# Metabolic Signatures of Resistance to Mycobacterium Tuberculosis Infection: Insights from a Multi-Country Plasma Metabolomics Study

**DOI:** 10.64898/2026.09.29.26364337

**Authors:** Chang Liu, Jeffrey M. Collins, Matheus Fernandes Gyorfy, Mariana Araújo Pereira, Vidya Mave, Senbagavalli Prakash, Kamakshi Prudhula Devalraju, Neil A. Martinson, Fay Willis, Marina C. Figueiredo, Marcelo Cordeiro-Santos, Artur Trancoso Lopo de Queiroz, Venkata Sanjeev Kumar Neela, Rajesh Karyakarte, Timothy R. Sterling, Jerrold J. Ellner, James C.M. Brust, Amita Gupta, Bruno B. Andrade, Yan V. Sun, Neel R. Gandhi, the TB GWAS collaboration

**Author notes:** Contributed equally (Yan V. Sun, Neel R. Gandhi). These authors jointly supervised this work. Corresponding Author: Yan V. Sun, PhD, VA Atlanta Health Care System, 1670 Clairmont Road, Decatur, Georgia 30033, Department of Epidemiology, Emory University Rollins School of Public Health 1518 Clifton Rd. NE, Atlanta, Georgia 30322.

## Abstract

**Background:** Tuberculosis (TB) remains the leading infectious disease cause of mortality worldwide. A subset of individuals exposed to *Mycobacterium tuberculosis* (*Mtb*) remain TST/IGRA-negative despite sustained exposure, termed “TB resisters”. The biological mechanisms underlying this resistance remain poorly understood. We applied untargeted high-resolution metabolomics to determine whether systemic metabolic profiles differ between TB resisters and matched *Mtb*-infected close contacts.

**Methods:** We compared untargeted high-resolution plasma metabolomics using liquid chromatography mass spectrometry between 269 TB resisters and 269 matched *Mtb*-infected close contacts, enrolled across Brazil, India, and South Africa. TB resisters were defined as highly exposed close contacts who slept in the same room or spent at least 5 hours indoors per day with an infectious pulmonary TB index patient, but remained TST/IGRA-negative. *Mtb*-infected close contacts were those who were TST/IGRA positive. Metabolome-wide association studies (MWAS) were conducted using paired t-tests.

**Results:** There were 1,787 features with nominal differences between TB resisters and matched *Mtb*-infected contacts (p < 0.05). Pathway enrichment identified fructose and mannose metabolism and bile acid biosynthesis in the overall cohort, while lipid-related pathways were enriched in Brazil. Among metabolites with confirmed chemical identities, glyceric acid concentrations were higher in TB resisters by 0.53 µM, whereas hydroxyproline was lower by 0.90 µM, butyrobetaine was lower by 0.06 µM, and homogentisate was lower by 0.0036 µM at false discovery rate of 0.2.

**Conclusions:** Plasma metabolomic profiles differed between TB resisters and matched *Mtb*-infected close contacts. These findings indicate that systemic metabolic differences are associated with resistance to *Mtb* infection.

## Introduction

Tuberculosis (TB) remains the leading infectious disease cause of death worldwide and continued transmission highlights the need for strategies that prevent *Mycobacterium tuberculosis* (*Mtb*) infection ^1^. A longstanding observation is that some close contacts of infectious TB patients remain uninfected, with persistently negative tuberculin skin test (TST) or interferon-γ release assays (IGRAs) despite sustained exposure. While it is unclear whether these “TB resisters” never become infected, clear bacilli early or become *Mtb* infected without an interferon gamma response ^2,3^, long-term follow-up data show that these individuals have a very low 10-year risk of developing TB disease or developing a positive TST or IGRA test ^4^. Identifying biologic mechanisms associated with this long-term protection may allow for the development of new prevention tools, including a new TB vaccine.

Metabolomic profiling captures the integrated biochemical consequences of immune activation and cellular energetics, providing a functional readout of immunometabolic pathways that may influence early *Mtb* clearance ^5,6^. Prior TB metabolomics studies have largely contrasted active TB with latent *Mtb* infection or more broadly defined “exposed/uninfected” comparators, and have repeatedly implicated disruptions in tryptophan-kynurenine catabolism, lipid remodeling and acylcarnitine metabolism, as well as amino acid metabolism ^7–9^. However, few studies have evaluated rigorously ascertained TB resisters, despite evidence that this phenotype reflects distinct host biology ^2,10,11^. To address this gap, we conducted a multi-country, matched resister-control study nested within close contact cohorts in India, Brazil, and South Africa. We applied untargeted high-resolution plasma metabolomics using the samples collected at enrollment to compare highly exposed contacts who remained persistently TST/IGRA-negative (“TB resisters”) with matched contacts with evidence of *Mtb* infection (TST/IGRA-positive). We hypothesized that TB resisters would exhibit distinct systemic metabolic profiles compared with *Mtb*-infected controls.

## Methods

### Study Population

Participants for this study were drawn from the parent TB genome-wide association study (GWAS) cohort, which enrolled 4,058 close contacts of pulmonary TB patients from nine prospective cohorts across 10 sites in Brazil, India, and South Africa, in collaboration with the RePORT consortia; 4,000 participants had sufficient data to determine resister/control status **Supplemental Table 1** ^12,13^. Detailed study methods of the parent TB study have been previously published ^13^. Briefly, close contacts of microbiologically confirmed (e.g., positive culture and/or Xpert) pulmonary TB index patients were eligible for enrollment if they: (i) resided in the same household for ≥1 month, or (ii) spent ≥4 hours per week with the index patient before the index patient’s diagnosis. Close contacts were excluded if they were HIV-coinfected or unable to provide informed consent/assent.

Characterization of TB resister and control phenotypes are detailed in **Supplemental Methods**. Briefly, we created eight phenotypic categories representing a hierarchy of resistance and infection, based on index patient infectiousness, exposure to the index patient, and IGRA and/or TST test results **Supplemental Figure 1**. We identified TB resisters among participants with high exposure (slept in the same room or ≥5 hours indoors) to a highly infectious index case (cavitary disease or 2+/3+ smear grade), who were persistently TST and/or IGRA negative were categorized as Resister A, B, or C categories, **Supplemental Figure 1**. Controls were selected from participants with a high likelihood of being *Mtb*-infected. These included contacts with positive TST and/or IGRA results or a history of TB disease, categorized as Infected A if both IGRA and TST were positive, and Infected B if either IGRA or TST was available and positive, **Supplemental Figure 1**.

In the current metabolomic study, we analyzed a subset of 538 participants from the parent TB GWAS study. Participants with a high likelihood of being resistant to *Mtb* infection (Resister A n = 35; Resister B n = 234; Resister C n = 0; total n=269) were matched 1:1 to participants with a high likelihood of being *Mtb*-infected (Infected A n = 77; Infected B n = 192; total n=269). For each Resister participant, a matched control was sought who shared the same index patient and who was similar in age. When matches with the same index patient were not possible, control participants with a similar age, same sex and within same country of enrollment were sought, **Supplemental Methods**.

All participants provided written informed consent/assent. Institutional review boards at Emory University and all participating sites approved the study, **Supplemental Methods**.

### Untargeted High-Resolution Metabolic Profiling

Baseline plasma samples were analyzed using previously established methodologies ^14–18^. The plasma specimens were collected, stored at –80°C, and thawed prior to analysis by liquid chromatography-mass spectrometry (LC-MS). To prepare the samples, aliquots were treated with acetonitrile containing an isotopic internal standard mix, then centrifuged at 4°C for 10 minutes to remove proteins. The resulting supernatant was kept in an Accela Open Autosampler at 4°C for subsequent analysis. Triplicate analyses were performed using liquid chromatography coupled to high-resolution Orbitrap mass spectrometry (Q Exactive, Thermo Fisher Scientific), acquired over an m/z range of 85-1100 with a 5-minute run time. Extracts were analyzed using HILIC with positive electrospray ionization and reverse-phase C18 chromatography with negative electrospray ionization. HILIC and reverse-phase C18 chromatography were used as complementary platforms to broaden metabolomic coverage, with HILIC enriching detection of polar metabolites and C18 improving detection of less polar and lipid-associated metabolites. HILIC separation was performed using a Waters XBridge BEH Amide XP HILIC column. C18 separation was performed using an endcapped Higgins Targa C18 column. The software apLCMS was utilized for metabolic feature extraction ^19^. A metabolic feature is defined by m/z (mass-to-charge ratio) and retention time. Only features showing a Pearson correlation coefficient greater than 0.7 across all triplicates were retained to ensure data quality. The intensities of metabolic features were summarized as median values of the non-zero readings across triplicates. Features with >50% missing values were excluded from downstream analyses. The remaining zero readings were considered values under detectable range and imputed as random values below the half of the minimum intensities across all samples. Intensities were then log2-transformed (log2(intensity+1)), mean-centered, and scaled by standard deviation.

Annotation of features was performed using xMSannotator ^20^ matching to the Human Metabolome Database ^21^. xMSannotator assigns confidence scores (0-3) by integrating multiple orthogonal pieces of evidence, including expected isotope/adduct relationships, retention-time clustering and intensity correlation patterns across samples, and network/pathway context, and then applying chemical plausibility filters ^20^. Annotations with a confidence score of 3 were classified as highly confident, and those with a score of 2 as moderately confident. Only annotations with a score of 2 or higher were used. In addition, a subset of metabolites were further confirmed based on established standards ^15,22^. Metabolite identification and confirmation were carried out using an in-house metabolite library, which was validated through comparisons of retention time and MS/MS data with authentic standards, as well as by matching MS/MS fragments with online spectral databases. The library was then used, allowing a tolerance of ± 5 ppm for m/z and ± 30 seconds for retention time, relative to standardized plasma, for accurate metabolite annotation. The intensities of confirmed metabolites were converted to concentrations in uM using a previously described protocol ^15^.

### Statistical Analysis

Subject characteristics were described as median (interquartile range) for continuous variables and frequency (percentage) for categorical variables. These characteristics are described for the overall cohort and stratified by study site, including Brazil, India, and South Africa.

For the untargeted metabolome-wide association study (MWAS), paired t-tests were conducted to compare the metabolite intensities between the paired TB resisters and matched controls for each metabolic feature. The paired t-test was chosen to account for the within-pair nature of the comparison. Statistical significance was assessed using a threshold of p < 0.05. To account for multiple testing, the Benjamini-Hochberg false discovery rate (FDR) ^23^ method was used with FDR corrected q value < 0.2 defined as statistical significance ^24,25^. For metabolites with the highest confidence annotations and concentrations, a separate analysis was performed on the concentrations using paired t-tests. Similarly, multiple testing correction was applied using the FDR approach. All analyses were performed in the overall cohort, and stratified by study site. Analyses were performed using R version 4.4.0 (R Foundation for Statistical Computing, Vienna, Austria).

Additionally, pathway enrichment analysis was performed following the untargeted MWAS using Mummichog (version 2.1.1) ^26^. Metabolic features that were different between TB resisters and non-resisters at a p value < 0.05 were chosen as target features and all features in the MWAS were utilized as the feature reference pool. Pathways with a FDR corrected q value < 0.2 were considered statistically significant.

## Results

The study included 269 matched pairs of TB resisters and *Mtb*-infected controls, including 155 pairs from Brazil, 103 from India, and 11 from South Africa, **Table 1**. By design, all resisters were QFT-negative and had high-intensity exposure to highly infectious index cases (cavities or AFB smear ≥2+/3+), with shared room/bed or ≥5 hours of indoor exposure. Across sites, males comprised 40% of resisters and 42% of controls; median age was 27-28 years overall (32-34 years in Brazil; youngest in South Africa at 13 years), and median BMI was 23.7 kg/m^2^ in resisters vs 23.1 kg/m^2^ in controls, highest in Brazil (24.3-24.6 kg/m^2^) and lowest in South Africa (17.2 kg/m^2^ in resisters; 14.5 kg/m^2^ in controls).

**Table 1.** Characteristics of the Study Cohort.

| Variable | Overall<br>269 Pairs |  | Brazil<br>155 Pairs |  | India<br>103 Pairs |  | South Africa<br>11 Pairs |  |
| --- | --- | --- | --- | --- | --- | --- | --- | --- |
|  | Resisters<br>N = 269 | Controls<br>N = 269 | Resisters<br>N = 155 | Controls<br>N = 155 | Resisters<br>N = 103 | Controls<br>N = 103 | Resisters<br>N = 11 | Controls<br>N = 11 |
| Age, years | 27<br>(13, 43) | 28<br>(13, 42) | 34<br>(15, 46) | 32<br>(15, 46) | 23<br>(12, 37) | 25<br>(12, 38) | 13<br>(12, 19) | 13<br>(12, 19) |
| **Sex (%)** |  |  |  |  |  |  |  |  |
| Female | 161 (60) | 155 (58) | 92 (59) | 94 (61) | 60 (58) | 53 (52) | 9 (82) | 8 (73) |
| Male | 108 (40) | 114 (42) | 63 (41) | 61 (39) | 43 (42) | 50 (49) | 2 (18) | 3 (27) |
| BMI, kg/m<sup>2</sup> | 23.7<br>(18.7, 28.2) | 23.1<br>(18.4, 27.9) | 24.6<br>(20.6, 29.3) | 24.3<br>(19.9, 28.7) | 21.7<br>(16.8, 26.5) | 20.3<br>(16.5, 25.2) | 17.2<br>(14.6, 19.5) | 14.5<br>(13.4, 19.8) |
| **QuantiFERON-TB Gold Test (%)** |  |  |  |  |  |  |  |  |
| Negative | 269 (100) | 0 (0) | 155 (100) | 0 (0.0) | 103 (100) | 0 (0) | 11 (100) | 0 (0) |
| Positive | 0 (0) | 269 (100) | 0 (0) | 155 (100) | 0 (0) | 103 (100) | 0 (0) | 11 (100) |
| **Resistance & Infection Hierarchy Category** |  |  |  |  |  |  |  |  |
| Resister A | 35 (13) | 0 (0) | 0 (0) | 0 (0) | 35 (34) | 0 (0) | 0 (0) | 0 (0) |
| Resister B | 234 (87) | 0 (0) | 155 (100) | 0 (0) | 68 (66) | 0 (0) | 11 (100) | 0 (0) |
| Resister C | 0 (0) | 0 (0) | 0 (0) | 0 (0) | 0 (0) | 0 (0) | 0 (0) | 0 (0) |
| Infected A | 0 (0) | 77 (29) | 0 (0) | 9 (6) | 0 (0) | 63 (61) | 0 (0) | 5 (45) |
| Infected B | 0 (0) | 192 (71) | 0 (0) | 146 (94) | 0 (0) | 40 (39) | 0 (0) | 6 (55) |
| **Index case infectiousness** |  |  |  |  |  |  |  |  |
| High: Cavities present or AFB smear 2+/3+ | 269 (100) | 201 (75) | 155 (100) | 114 (74) | 103 (100) | 85 (83) | 11 (100) | 2 (18) |
| Medium: AFB smear scanty/1+, OR<br>GeneXpert grade high or very high (no cavities) | 0 (0) | 44 (16) | 0 (0) | 31 (20) | 0 (0) | 13 (13) | 0 (0) | 0 (0) |
| Low: AFB smear negative OR<br>GeneXpert grade low or very low (no cavities) | 0 (0) | 16 (6) | 0 (0) | 10 (7) | 0 (0) | 5 (5) | 0 (0) | 1 (9) |
| Not available | 0 (0) | 8 (3) | 0 (0) | 0 (0.0) | 0 (0) | 0 (0.0) | 0 (0) | 8 (73) |
| **Exposure to index patient** |  |  |  |  |  |  |  |  |
| Slept in same bed | 53 (20) | 38 (14) | 35 (23) | 17 (11) | 18 (18) | 20 (19) | 0 (0.0) | 1 (9) |
| Slept in same room | 66 (25) | 61 (23) | 15 (10) | 16 (10) | 49 (48) | 43 (42) | 2 (18) | 2 (18) |
| Spent ≥5 hours indoors together | 150 (56) | 83 (31) | 105 (68) | 55 (36) | 36 (35) | 21 (20) | 9 (82) | 7 (64) |
| None of the above | 0 (0) | 18 (7) | 0 (0) | 8 (5) | 0 (0) | 10 (10) | 0 (0) | 0 (0) |
| Not available | 0 (0) | 69 (26) | 0 (0) | 59 (38) | 0 (0) | 9 (9) | 0 (0) | 1 (9) |

Median (interquartile range) shown for continuous variables. Count (percentage) shown for categorical variables.
AFB, acid-fast bacillus; BMI, body mass index; GeneXpert, Xpert MTB/RIF molecular test for Mycobacterium tuberculosis detection; TB, tuberculosis.

A total of 12,177 and 10,996 metabolic features, respectively for HILIC positive and C18 negative modes, passed quality control and entered the untargeted MWAS comparing resisters with controls. The overall cohort showed 911 features in HILIC and 876 in C18 significant at p < 0.05, with only 2 HILIC features significant at FDR<0.2. In the study site stratified MWAS, no features passed the FDR threshold, except for one in the South Africa cohort from the HILIC positive mode, **Supplementary Table 2, Supplementary Figures 2-5**. Among the metabolic features with p < 0.05, 307 in the overall cohort, 270 in Brazil, 260 in India, and 191 in South Africa had annotations and are presented in **Supplementary Tables 3-6**, respectively.

The pathway enrichment analysis of the untargeted MWAS identified 2 and 13 pathways in the analysis of overall cohort and Brazil, respectively. Fructose and mannose metabolism and bile acid biosynthesis were enriched in the overall cohort. Key lipid-related pathways, such as fatty acid activation, linoleate metabolism, omega-6 and omega-3 fatty acid metabolism, di-unsaturated fatty acid beta-oxidation were enriched in the Brazil cohort, **Table 2**, **Figure 1**.

**Figure 1.**
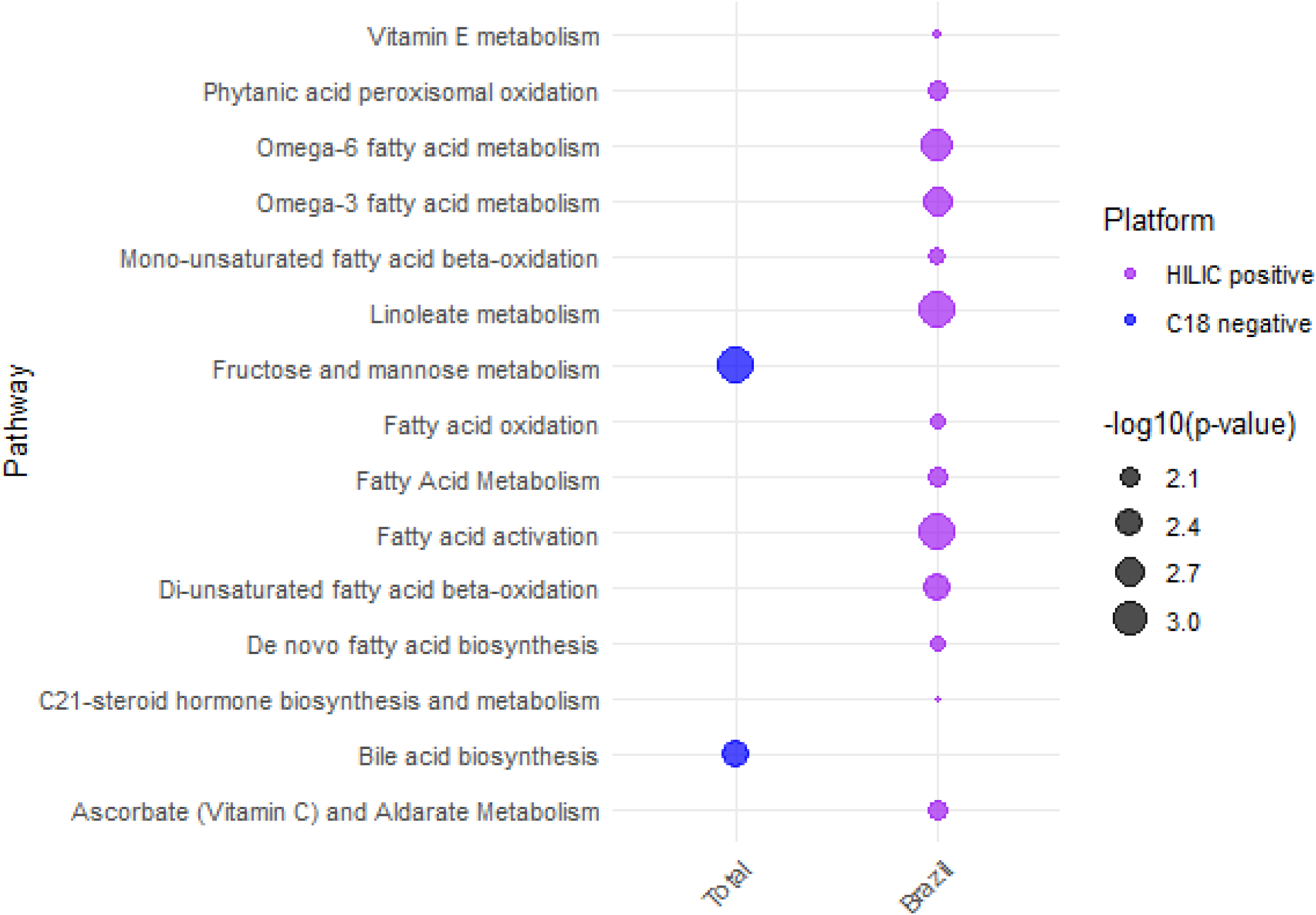
Pathway Enrichment Results of the Metabolome-wide Association Study Comparing TB Resisters and Controls. **Alt text:** Bubble plot showing enriched metabolic pathways among tuberculosis resisters versus controls. Metabolic features with p value < 0.05 were analyzed. Pathways with false discovery rate < 0.2 and overlapping size ≥ 3 are shown.

**Table 2.** Pathway Enrichment Results of the Untargeted Metabolome-wide Association Study Comparing TB Resisters and Controls. Metabolic Features with P Value < 0.05 Entered Pathway Analysis. Pathways with P Value < 0.05 and Overlapping Size ≥ 3 are Shown.

| Cohort | Platform | Pathway | Number of Significant Metabolites | Number of Metabolites in Pathway | P | FDR |
| --- | --- | --- | --- | --- | --- | --- |
| All Cohorts<br>(269 pairs) | C18 negative | Fructose and mannose metabolism | 8 | 12 | 0.0007 | 0.0800 |
|  | C18 negative | Bile acid biosynthesis | 15 | 45 | 0.0032 | 0.1900 |
| Brazil<br>(155 pairs) | HILIC positive | Fatty acid activation | 14 | 35 | 0.0006 | 0.0450 |
|  | HILIC positive | Linoleate metabolism | 14 | 36 | 0.0008 | 0.0450 |
|  | HILIC positive | Omega-6 fatty acid metabolism | 8 | 16 | 0.0017 | 0.0667 |
|  | HILIC positive | Omega-3 fatty acid metabolism | 8 | 17 | 0.0025 | 0.0740 |
|  | HILIC positive | Di-unsaturated fatty acid beta-oxidation | 5 | 8 | 0.0031 | 0.0740 |
|  | HILIC positive | Ascorbate (Vitamin C) and Aldarate Metabolism | 9 | 26 | 0.0073 | 0.1244 |
|  | HILIC positive | Fatty Acid Metabolism | 7 | 18 | 0.0085 | 0.1244 |
|  | HILIC positive | Phytanic acid peroxisomal oxidation | 6 | 14 | 0.0085 | 0.1244 |
|  | HILIC positive | Mono-unsaturated fatty acid beta-oxidation | 4 | 7 | 0.0094 | 0.1244 |
|  | HILIC positive | Fatty acid oxidation | 5 | 11 | 0.0115 | 0.1291 |
|  | HILIC positive | De novo fatty acid biosynthesis | 10 | 33 | 0.0119 | 0.1291 |
|  | HILIC positive | Vitamin E metabolism | 6 | 16 | 0.0151 | 0.1400 |
|  | HILIC positive | C21-steroid hormone biosynthesis and metabolism | 8 | 25 | 0.0153 | 0.1400 |

A total of 127 metabolites were confirmed, and their quantified concentrations were analyzed. In the overall cohort, glyceric acid concentrations were higher in TB resisters than in controls (4.04 vs. 3.51 µM; mean difference, 0.53 µM; 95% CI, 0.18-0.88; P = 0.0032; FDR = 0.1997), whereas hydroxyproline (6.44 vs. 7.34 µM; mean difference, –0.90 µM; 95% CI, –1.51 to –0.29; P = 0.0041; FDR = 0.1997) and butyrobetaine (0.76 vs. 0.82 µM; mean difference, – 0.06 µM; 95% CI, –0.10 to –0.02; P = 0.0047; FDR = 0.1997) were lower in TB resisters, **Table 3**. Notably, the direction of association for glyceric acid and butyrobetaine was generally consistent across Brazil and India, with no evidence of between-country heterogeneity (Pheterogeneity = 0.5667 and 0.5400, respectively). In contrast, hydroxyproline showed evidence of between-country heterogeneity (P heterogeneity = 0.0094), with a substantially larger inverse association in South Africa, although this estimate was based on only 11 pairs, **Table 3**. Country-specific analyses showed that homogentisate was lower in TB resisters than in controls in India (0.0189 vs. 0.0225 µM; mean difference, –0.0036 µM; 95% CI, –0.0057 to – 0.0014; P = 0.0015; FDR = 0.1853), the only country-specific association meeting an FDR <0.20 threshold, **Table 4**.

**Table 3.** Top Associations between Targeted Metabolites with Confirmed Annotations and TB Resistance in the Overall Cohort (269 Pairs). Quantified Concentrations in uM were Analyzed. Metabolites with P Value < 0.05 or False Discovery Rate < 0.2 in the Overall Cohort Analysis are Shown. Country-Specific Associations and Between-Country Heterogeneity for Targeted Metabolites Identified in the Overall Cohort are Shown.

| Platform | m/z | time | Name | Adduct | All Cohorts<br>(269 pairs) |  |  |  |  | Brazil<br>(155 pairs) |  |  |  |  | India<br>(103 pairs) |  |  |  |  | South Africa<br>(11 pairs) |  |  |  |  | Heterogeneity<br>Test P |
| --- | --- | --- | --- | --- | --- | --- | --- | --- | --- | --- | --- | --- | --- | --- | --- | --- | --- | --- | --- | --- | --- | --- | --- | --- | --- |
|  |  |  |  |  | Resister<br>Mean<br>(SD) | Control<br>Mean<br>(SD) | Difference<br>Resister<br>vs.<br>Control<br>(95% CI) | P | FDR | Resister<br>Mean<br>(SD) | Control<br>Mean<br>(SD) | Difference<br>Resister<br>vs.<br>Control<br>(95% CI) | P | FDR | Resister<br>Mean<br>(SD) | Control<br>Mean<br>(SD) | Difference<br>Resister<br>vs.<br>Control<br>(95% CI) | P | FDR | Resister<br>Mean<br>(SD) | Control<br>Mean<br>(SD) | Difference<br>Resister<br>vs.<br>Control<br>(95% CI) | P | FDR |  |
| C18 negative | 105.0193 | 20.2 | glyceric acid | M-H | 4.04<br>(2.51) | 3.51<br>(2.26) | 0.53<br>(0.18, 0.88) | 0.0032 | **0.1997** | 4.4 (2.5) | 3.73<br>(2.43) | 0.66<br>(0.15, 1.18) | 0.0119 | 0.5046 | 3.59<br>(2.46) | 3.19<br>(2.01) | 0.4<br>(-0.06, 0.86) | 0.0904 | 0.5669 | 3.31<br>(2.43) | 3.47<br>(1.85) | -0.17<br>(-1.92, 1.59) | 0.8358 | 0.9576 | 0.5667 |
| HILIC positive | 132.0656 | 70.7 | hydroxyproline | M+H | 6.44<br>(4.78) | 7.34<br>(5.75) | -0.9<br>(-1.51, -0.29) | 0.0041 | **0.1997** | 6.61<br>(4.21) | 7.32<br>(4.31) | -0.71<br>(-1.58, 0.17) | 0.1129 | 0.6034 | 5.77<br>(5.29) | 6.36<br>(6.49) | -0.59<br>(-1.37, 0.19) | 0.1349 | 0.6277 | 10.3<br>(5.64) | 16.8<br>(7.88) | -6.51<br>(-10.23, -2.78) | 0.0030 | 0.3812 | **0.0094** |
| HILIC positive | 146.1176 | 45.6 | butyrobetaine | M+H | 0.76<br>(0.41) | 0.82<br>(0.42) | -0.06<br>(-0.1, -0.02) | 0.0047 | **0.1997** | 0.77<br>(0.34) | 0.84<br>(0.4) | -0.07<br>(-0.12, -0.02) | 0.0056 | 0.5046 | 0.7 (0.5) | 0.76<br>(0.44) | -0.06<br>(-0.14, 0.01) | 0.0966 | 0.5669 | 1.06<br>(0.38) | 0.95<br>(0.42) | 0.11<br>(-0.21, 0.43) | 0.4585 | 0.8158 | 0.5400 |
| C18 negative | 129.0193 | 21 | citraconate | M-H | 7.85<br>(4.78) | 6.97 (4.1) | 0.88<br>(0.22, 1.53) | 0.0092 | 0.2936 | 9.18<br>(5.15) | 7.99<br>(4.52) | 1.19<br>(0.18, 2.2) | 0.0211 | 0.6034 | 5.85<br>(3.3) | 5.35<br>(2.93) | 0.5<br>(-0.26, 1.25) | 0.1965 | 0.6299 | 7.79<br>(4.83) | 7.76<br>(2.1) | 0.02<br>(-3.42, 3.46) | 0.9888 | 0.9888 | 0.5139 |
| C18 negative | 167.0342 | 21.3 | homogentisate | M-H | 0.0179<br>(0.0063) | 0.0191<br>(0.0079) | -0.0012<br>(-0.0022, -2e-04) | 0.0176 | 0.4296 | 0.0177<br>(0.005) | 0.0174<br>(0.0044) | 3e-04<br>(-6e-04, 0.0012) | 0.5156 | 0.7920 | 0.0189<br>(0.0077) | 0.0225<br>(0.0106) | -0.0036<br>(-0.0057, -0.0014) | 0.0015 | 0.1853 | 0.0116<br>(0.0034) | 0.0117<br>(0.0035) | -1e-04<br>(-0.0026, 0.0024) | 0.9493 | 0.9794 | **0.0052** |
| HILIC positive | 147.0764 | 77.1 | glutamine | M+H | 277.58<br>(104.41) | 290.43<br>(105.81) | -12.85<br>(-23.76, -1.94) | 0.0212 | 0.4296 | 286.62<br>(81.43) | 297.71<br>(92.68) | -11.09<br>(-22.6, 0.41) | 0.0586 | 0.6034 | 256.46<br>(130.74) | 267.08<br>(118.42) | -10.63<br>(-32.72, 11.47) | 0.3424 | 0.7518 | 347.94<br>(66.22) | 406.34<br>(59.54) | -58.4<br>(-117.57, 0.77) | 0.0525 | 0.7457 | 0.3018 |
| HILIC positive | 114.0662 | 36.1 | creatinine | M+H | 37.34<br>(14.14) | 38.71<br>(13.96) | -1.37<br>(-2.55, -0.18) | 0.0237 | 0.4296 | 40.26<br>(13.75) | 41.18<br>(14.19) | -0.92<br>(-2.43, 0.58) | 0.2280 | 0.6354 | 32.37<br>(13.47) | 34.94<br>(13.07) | -2.57<br>(-4.56, -0.59) | 0.0115 | 0.4117 | 42.89<br>(13.27) | 39.21<br>(11.94) | 3.68<br>(-3.54, 10.9) | 0.2825 | 0.7457 | 0.1629 |
| HILIC positive | 120.0032 | 65.8 | glycine | M+2Na-H | 105.15<br>(41.2) | 109.92<br>(42.22) | -4.76<br>(-9.27, -0.25) | 0.0385 | 0.5178 | 103.76<br>(36.06) | 107.09<br>(38.82) | -3.33<br>(-9.7, 3.04) | 0.3034 | 0.7136 | 104.39<br>(48.71) | 110.79<br>(47.23) | -6.4<br>(-12.97, 0.18) | 0.0563 | 0.4578 | 131.97<br>(20.83) | 141.66<br>(23.99) | -9.69<br>(-34.74, 15.37) | 0.4092 | 0.8031 | 0.7512 |
| HILIC positive | 269.2257 | 27.5 | retinol | M-H2O+H | 6.72<br>(2.97) | 6.39<br>(2.56) | 0.33<br>(0.01, 0.64) | 0.0406 | 0.5178 | 7.78<br>(2.91) | 7.33<br>(2.59) | 0.45<br>(0, 0.9) | 0.0511 | 0.6034 | 5.19<br>(2.44) | 5.14<br>(1.94) | 0.05<br>(-0.39, 0.5) | 0.8116 | 0.9539 | 6.01<br>(1.75) | 4.78<br>(0.75) | 1.22<br>(-0.23, 2.68) | 0.0905 | 0.7457 | 0.2101 |
| HILIC positive | 163.123 | 41.4 | nicotine | M+H | 0.02<br>(0.07) | 0.03 (0.1) | -0.01<br>(-0.03, 0) | 0.0423 | 0.5178 | 0.01<br>(0.03) | 0.02<br>(0.05) | -0.01<br>(-0.01, 0) | 0.2303 | 0.6354 | 0.03<br>(0.1) | 0.05<br>(0.14) | -0.02<br>(-0.05, 0.01) | 0.1588 | 0.6299 | 0 (0) | 0.06<br>(0.21) | -0.06<br>(-0.2, 0.08) | 0.3379 | 0.7599 | 0.4473 |

SD: Standard Deviation; CI: Confidence Interval; FDR: False Discovery Rate. Heterogeneity P values were calculated using Cochran’s Q test from a random-effects meta-analysis of the site-specific effect estimates from Brazil, India, and South Africa;

**Table 4.**
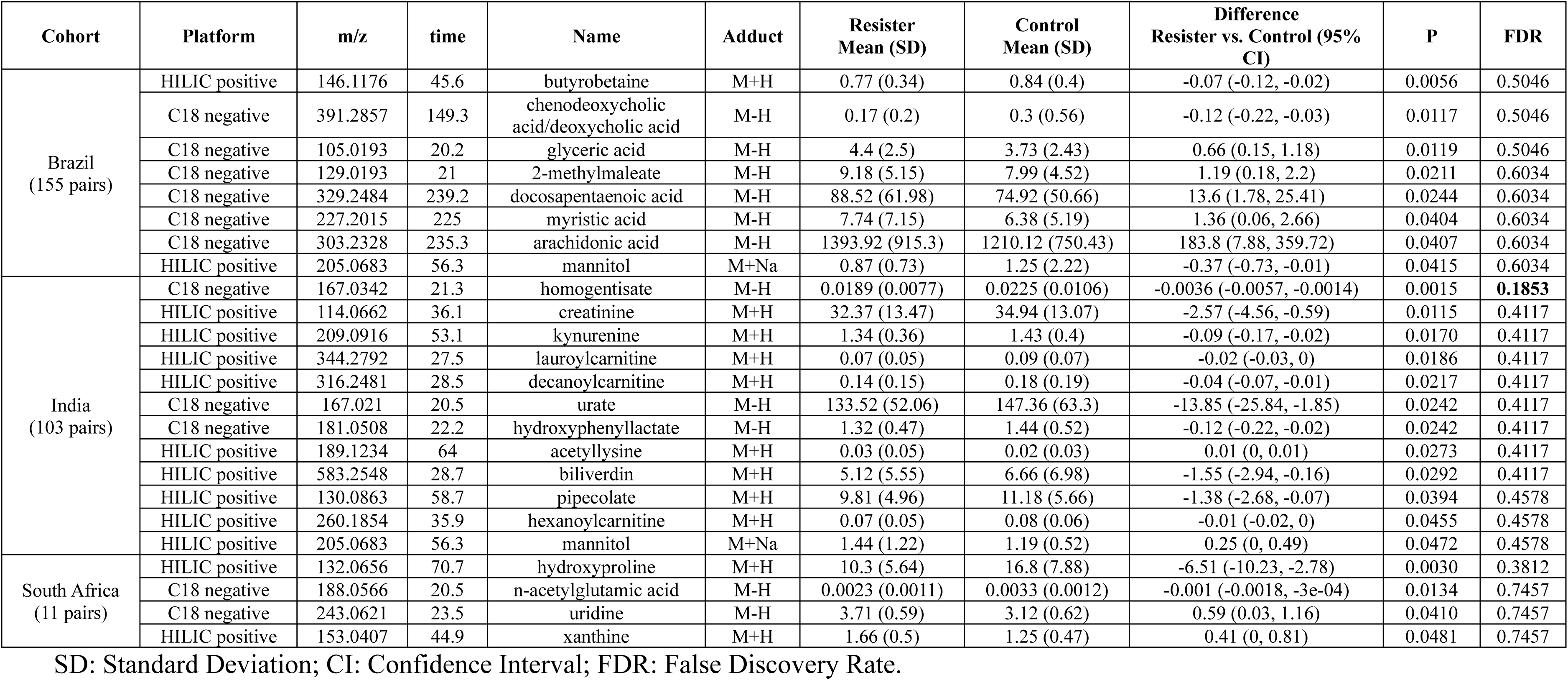
Top Associations between Targeted Metabolites with Confirmed Annotations and TB Resistance in the Country-Specific Cohorts. Quantified Concentrations in uM were Analyzed. Metabolites with P Value < 0.05 or False Discovery Rate < 0.2 in Each Country-Specific Analysis are Shown.

## Discussion

This study investigated the metabolic differences between TB resisters and matched *Mtb*-infected controls using untargeted high-resolution metabolic profiling of plasma samples and additionally investigated confirmed metabolites using a targeted design. Pathway enrichment suggested involvement of carbohydrate metabolism, bile acid biosynthesis, and lipid-related pathways, particularly in the Brazil cohort. In the targeted study of metabolite concentrations, glyceric acid, hydroxyproline, butyrobetaine and homogentisate were significantly different between TB resisters and *Mtb*-infected controls. Together, these findings suggest that resistance to sustained *Mtb* infection may be associated with differences in systemic metabolic states related to energy metabolism, amino acid turnover, lipid handling, and oxidative or inflammatory processes.

Fructose and mannose metabolism intersects with central carbon metabolism and glycolytic flux, processes that are dynamically remodeled during macrophage responses to *Mtb* infection. Prior plasma metabolomics studies of pulmonary TB have also implicated fructose and mannose metabolism, glycolysis/gluconeogenesis, pyruvate metabolism, and bile acid biosynthesis as altered pathways in TB patients ^27^. The mannose-related signal is also biologically plausible in the context of *Mtb*-host interactions, as mannose-capped mycobacterial cell-wall components can engage macrophage mannose receptors and influence *Mtb* uptake and phagosome-lysosome trafficking ^28,29^. Similarly, bile acids are increasingly recognized as immunometabolic signaling molecules that regulate innate immune and inflammatory responses through bile acid-activated receptors ^30,31^. Thus, the enrichment of fructose/mannose and bile acid pathways may reflect systemic metabolic programs linked to macrophage activation, microbial sensing, or inflammatory regulation after high-intensity *Mtb* exposure. However, because these are pathway-level findings from cross-sectional plasma metabolomics, they should be interpreted as hypothesis-generating rather than evidence of causal mechanisms.

Our findings also highlight the critical role of lipid metabolism in shaping host immune responses to *Mtb*. This reflects differences in systemic lipid handling after high-intensity exposure, which may shape the metabolic milieu available to immune cells and influence antimicrobial programs. Intracellular *Mtb* can preferentially utilize host-derived fatty acids and cholesterol and can acquire a survival advantage in lipid-rich macrophage environments ^32–34^. In this context, enrichment of fatty-acid pathways could indicate a host state with altered lipid mobilization/turnover or lipid-mediator production that is associated with resistance. Notably, linoleate and omega-3/omega-6 pathways are upstream of bioactive lipid mediators that can modulate inflammation and macrophage fate decisions in TB, offering a mechanistic framework for how lipid signatures might relate to resistance ^35^. This interpretation is also consistent with prior work showing distinct monocyte transcriptional programs in highly exposed individuals who resist TST/IGRA conversion ^10,36^. Because our measurements are cross-sectional in plasma, we cannot determine whether these lipid-associated differences precede exposure or reflect very early host responses to exposure to *Mtb*; nevertheless, they nominate lipid metabolism as a key candidate axis associated with the TB resister phenotype that warrants longitudinal and cell-specific follow-up.

We also examined individual metabolites with high-confidence identifications, and several specific metabolites emerged as significantly different between resisters and *Mtb*-infected controls. One of the most notable was glyceric acid, which was elevated in resisters. Glyceric acid is a sugar acid produced from the oxidation of glycerol or intermediate steps of glycolysis ^37^, which are pathways that generate and use cellular fuel ^38^. Although glyceric acid itself has not been widely reported in TB, its higher levels in resisters may reflect the possibility that resisters differ in how rapidly they engage energy metabolism that supports innate immune activation. Hydroxyproline was significantly lower in resisters compared to *Mtb*-infected controls. Hydroxyproline is a non-standard amino acid released largely from the breakdown of collagen. It serves as a classic marker of collagen turnover and tissue remodeling ^39^. The lower plasma hydroxyproline in resisters implies less collagen degradation and tissue matrix remodeling in these individuals. Active TB disease is known to elevate hydroxyproline levels. A recent study found significantly higher hydroxyproline in the serum and even bronchoalveolar fluid of TB patients, correlating with inflammatory markers ^40^. This finding reinforces the notion that resisters did not develop an infection that triggered tissue reorganization. Butyrobetaine was lower in TB resisters, pointing to differences in carnitine-linked lipid metabolism. Butyrobetaine is the immediate precursor of L-carnitine, which supports mitochondrial transport and oxidation of long-chain fatty acids ^41^. Because butyrobetaine can also arise from microbial metabolism of dietary carnitine and feed into the TMA/TMAO pathway, this finding suggests potential host-microbiome influences on energy metabolism that warrant follow-up with longitudinal sampling and related metabolites ^42^. Homogentisate is formed from tyrosine metabolism, and previous study identified a link between host tyrosine metabolism and susceptibility to TB through genetic regulation of the *FAH* gene, which encodes the enzyme responsible for the final step of tyrosine catabolism.^43^ These results highlight tyrosine metabolism as a potential candidate pathway for future studies on TB and underscore the importance of integrating host genetics and metabolomics to uncover novel mechanisms of TB susceptibility.

The cross-sectional design limits causal inference, and metabolite differences may reflect consequences rather than causes of infection status. The modest sample size, particularly in South Africa, and inter-site variability may introduce heterogeneity. Further, plasma metabolomics may not fully capture lung-specific metabolic processes critical to early *Mtb* clearance, highlighting the need for longitudinal and tissue-focused follow-up studies. The major strength of the study is the use of a rigorously characterized, multi-ancestry cohort of TB resisters and matched *Mtb*-infected controls, enabling robust assessment of resistance to *Mtb* infection. The application of untargeted high-resolution metabolomics provided a comprehensive and unbiased view of systemic metabolic states, with rigorous quality control and pathway-level validation enhancing reliability.

## Conclusion

TB resisters showed distinct plasma metabolic patterns involving energy metabolism, bile acid biosynthesis, lipid-related pathways, and selected metabolites, including glyceric acid, hydroxyproline, butyrobetaine, and homogentisate. These findings support systemic metabolism as a candidate axis associated with resistance to sustained *Mtb* infection. Longitudinal and cell-specific studies are needed to validate these findings and determine whether these metabolic differences represent biomarkers or mechanisms of protection.

## Author Contributions

CL, JMC, YVS, and NRG conceived and designed the study. CL performed the statistical analyses, interpreted the results, created the figures and tables, and drafted the manuscript. JMC, MFG, and FW contributed to the metabolomics methodology, data curation, and interpretation of the findings. MAP, VM, SP, KPD, NAM, MCF, MCS, ATLQ, VSKN, RK, TRS, JJE, JCMB, AG, and BBA contributed to participant recruitment, cohort management, phenotyping, sample or data acquisition, and interpretation of the findings. YVS and NRG supervised the study and obtained funding. CL and YVS accessed and verified the underlying data. All authors reviewed and edited the manuscript, approved the final version, and accept responsibility for the decision to submit for publication.

## Data Sharing Statement

Deidentified participant-level data are not publicly available because of participant consent, institutional review board requirements, and data-use restrictions across the participating cohorts. Requests for access may be submitted to the corresponding author and will be considered by the relevant study investigators and institutions, subject to applicable approvals and data-use agreements. Summary-level metabolomics results are provided in the Supplementary Materials.

## RePORT South Africa and HomeACF Acknowledgements

Participant recruitment and follow up was supported by the UK/South Africa Medical Research Council (MRC) Newton Fund (006Newton TB); and by the NIH through CRDF Global (DAA2-16-62117-1). The Free State, Limpopo and the North West Provincial Departments of Health facilitated and hosted these studies.

## RePORT India Acknowledgements

Participant recruitment and follow up was supported by Federal funds from the Government of India’s Department of Biotechnology (DBT) Project number BT/MB/COMMON PROTOCOL/01/2014-15 sanctioned to Bhagawan Mahavir Medical Research Centre (BMMRC), the Indian Council of Medical Research (ICMR), the United States NIH, NIAID, Office of AIDS Research (OAR), and distributed in part by CRDF Global.

RePORT India Participating Institutions, Principal investigators, co-principal investigators, DBT, ICMR, and NIAID staff include: Byramjee Jeejeebhoy Government Medical College (BJGMC): Aarti Kinikar, Sanjay Gaikwad, Rajesh Kayakarte; Byramjee Jeejeebhoy Government Medical College–Johns Hopkins India Clinical Research Site: Vidya Mave, Mandar Paradkar, Nikhil Gupte, Nishi Suryavanshi; National Institute for Research in Tuberculosis (NIRT): P. K. Bhavani, Elizabeth Hanna; Johns Hopkins University (JHU): Amita Gupta, Robert Bollinger, Jeff Tornheim; Bhagawan Mahavir Medical Research Centre (BMMRC): Vijaya Valluri; Saint Louis University: Ramakrishna Vankayalapati; Jawaharlal Institute of Postgraduate Medical Education and Research (JIPMER): Sonali Sarkar, Gautam Roy; Rutgers: Padmini Salgame, Jerrold Ellner; Centre for Cellular & Molecular Biology (CCMB): Vinay Nandicoori; DBT: Jyoti Logani; NIAID: Peter Kim, Fatima Jones; and CRDF Global: Tara Moore.

## RePORT-Brazil Acknowledgements

Data in this manuscript were collected as part of the RePORT-Brazil Consortium. This project has been funded in part with Federal funds from the Brazilian Ministry of Health (MoH), the National Council for Scientific and Technological Development (CNPq), the Fundação Oswaldo Cruz (Fiocruz), and the US NIH. It was also supported in part by NIH/NIAID grants: U01AI069923 (MPIs: Castillo, Cahn, Duda), R01AI120790 (MPIs: Sterling, Rolla), U01AI172064 (MPIs: Sterling, Andrade), U01AI174268 (MPIs: Ellner/Sterling/Andrade), R01AI147765 (MPIs: Sterling/Andrade/Hawn), AK, BBA, and MC-S are fellows from the Conselho Nacional de Desenvolvimento Científico e Tecnológico (CNPq), Brazil.

RePORT-Brazil Participating Institutions, Principal and Co-Investigators include: Fundação José Silveira (FJS), Instituto Brasileiro para a Investigação da Tuberculose (IBIT), and Fundação Oswaldo Cruz–Bahia (Fiocruz-BA): Bruno B. Andrade; Fundação de Medicina Tropical Dr. Heitor Vieira Dourado (FMT-HVD): Marcelo Cordeiro-Santos; Instituto Nacional de Infectologia Evandro Chagas (INI–Fiocruz): Valeria Rolla; Municipal Health Rinaldo De Lamare (Rocinha), Rio de Janeiro: Solange Cavalcante, Betina Durovni; Municipal Health Department of Duque de Caxias, and Universidade Federal do Rio de Janeiro (UFRJ): Afranio Kritski; Vanderbilt University Medical Center (VUMC): Timothy R. Sterling, Marina Cruvinel Figueiredo, and Megan Turner.

The contents of this publication are solely the responsibility of the authors and do not necessarily represent the official views of the US NIH; South Africa MRC; India DBT or ICMR; Brazil MoH, CNPq, Fiocruz, or CRDF Global. Any mention of trade names, commercial products, or organizations does not imply endorsement by any of the sponsoring agencies.

For the purpose of open access, the author has applied a CC BY public copyright license to any author accepted manuscript version arising from this submission.

