## Supplemental Material for "Metabolic Signatures of Resistance to Mycobacterium Tuberculosis Infection: Insights from a Multi-Country Plasma Metabolomics Study"

**Supplemental Methods**

Study Population

The parent TB GWAS study [1] was an observational study among close contacts of active, pulmonary TB patients enrolled from nine cohorts from 10 research centers across Brazil, India, and South Africa, in collaboration with the RePORT consortia [2]. Participants of all ages were eligible if exposed to microbiologically confirmed (e.g., positive culture, Xpert) index patients. The minimum exposure for eligibility was living in the same house for ≥1 month, or spending ≥4 hours per week in the 6 months before the index patient's diagnosis (i.e., non-household close contacts). Close contacts were excluded if they were living with HIV or could not provide informed consent.

### Measurements of *Mtb* infection, index patient infectiousness, and exposure in the TB GWAS study

From each participating cohort in the parent TB GWAS study, we harmonized data that assessed index patient infectiousness (e.g., chest x-ray, smear microscopy), contact exposure (e.g., hours spent indoors, sleeping arrangements), and results and timing of IGRA and/or TST tests. Because many cohorts began enrollment before the initiation of the TB GWAS study, the availability of data varied across cohorts.

IGRA results: We collated results of all interferon gamma release assays (IGRA) performed for each participant. Cohorts utilized the Quantiferon Gold-in-Tube (3$^{rd}$ generation, Qiagen) or Quantiferon Gold Plus (4$^{th}$ generation, Qiagen) assays. A threshold of 0.35 IU/ml was used to determine positivity. For persistently IGRA-negative participants, the number of days between the index patient's diagnosis and the contact's last IGRA test measured was determined; this measurement allowed us to assess if sufficient time had passed to allow for IGRA conversion as a result of this most recent TB exposure. Participants were categorized as: IGRA-positive; IGRA-negative (>90 days); IGRA-negative (<90 days).

TST results: We collated results of all tuberculin skin tests (TST) performed for each participant. A threshold of 5 mm of induration was used to determine positivity. For persistently TST-negative participants, the number of days between the index patient's diagnosis and the contact's last TST test measured was determined; this measurement allowed us to assess if sufficient time had passed to allow for TST conversion. Participants were categorized as: TST-positive; TST-negative (>90 days); TST-negative (<90 days). TST were routinely performed only in cohorts from India. Difficulty importing tuberculin and issues related to standardization TST resulted in only a small minority of participants from Brazil and South Africa having TST results available.

Index patient infectiousness: Chest x-ray and AFB smear microscopy data of the participants' index patients were collated to categorize infectiousness of the TB patients that participants were exposed to. Infectiousness was categorized as:

- *High*: Cavities present on chest x-ray, or AFB smear positive with a grade of 2+ or 3+.
- *Medium*: No cavities on chest x-ray and AFB smear positive with a grade scanty or 1+.
- *Low*: No cavities on chest x-ray and AFB smear negative.

Exposure to the index case: Participants exposure to the TB index patient was characterized based on whether they slept in the same bed or room as the index patient and the number of hours spent indoors with the patient.

- *High*: Participant slept in same bed or room or spent ≥5 hours indoors with the index patient before their TB diagnosis.

- *Low*: Participant did not sleep in the same room and spent <5 hours indoors with the index patient.

Characterization of Resister Phenotype in the TB GWAS study

In the parent TB GWAS study, we created phenotypic categories characterizing the likelihood of participants being resistant to *Mtb* infection ("resister") or being *Mtb*-infected. Initially, we assessed IGRA and/or TST results to create three categories: a) infected: all TST/IGRA tests performed were positive; b) discordant: at least one positive TST/IGRA and one negative TST/IGRA; c) uninfected: all TST/IGRA tests performed were negative. The timing of TST/IGRA results (>90 days or ≤90 days after index patient's TB diagnosis date) was assessed to account for potential TST/IGRA conversion after exposure. Among uninfected participants, we characterized the index patient's infectiousness and participants' exposure to the index case, **Supplemental Figure 1**.

Using these criteria, we created eight phenotypic categories representing a hierarchy of resistance and infection. Participants with negative IGRA and/or TST were categorized into five "resisters" categories, reflecting differing probabilities of being resistant to *Mtb* infection. Participants with high exposure (slept in the same room or ≥5 hours indoors) to a highly infectious index case (cavitary disease or 2+/3+ smear grade), were placed in the Resister A, B, and C categories, **Supplemental Figure 1**. Participants with medium, low or missing exposure or infectiousness data were in Uninfected A and B categories. Contacts with positive TST and/or IGRA or history of TB disease were categorized as *Mtb* infected (Infected A and B categories, **Supplemental Figure 1**). Participants with discordant IGRA and TST results were categorized as "Discordant." The results categories are listed below:

Uninfected categories: Participants whose results for all IGRA or TST tests performed were negative were classified into 5 categories: Resister A-C and Uninfected A/B. Participants who were exposed to an index patient with "high" infectiousness (e.g., cavities, smear 2+/3+) and had a "high" degree of exposure (e.g., slept in same room) were placed into categories Resister A-C.

- *Resister A*: Highly exposed participants who had both a negative IGRA and negative TST and at least one of these tests was performed ≥90 days after the index patient's diagnosis.
- *Resister B*: Highly exposed participants with either only IGRA result(s) or TST result(s), but not both; the latest of those results was ≥90 days after index case diagnosis.
- *Resister C*: Highly exposed participants with IGRAs or TSTs, whose latest of those tests is <90 days after index patient diagnosis.
- *Uninfected A*: Uninfected participants whose index patient's infectiousness was medium or low, or whose exposure to the index patient was low.
- *Uninfected B*: Uninfected participants who were missing either index patient's infectiousness data or exposure data.

Discordant category: Participants who had both IGRA and TST results available, with at least one result positive and one result negative.

Infected categories: Participants whose results for all IGRA and TST tests were positive were separated into two categories: Infected A & B.

- *Infected A* had both IGRA and TST results available.
- *Infected B* had either an IGRA result available or a TST result, but not both.

Matching process for participants

A subset of 269 participants with a high likelihood of resistance to *Mtb* infection (Resisters A-C, **Supplemental Figure 1**) from the parent TB GWAS study were selected for the metabolomic analysis in this study. Resisters were matched 1:1 to *Mtb*-infected close contacts. The strategy for matching was to match TB resisters to an Mtb-infected contact who was exposed to the same infectious TB index patient (within-index matches). Within-index matches were made on age as closely as possible; however, when a precise age match was not available, the shared-index nature of the pair was prioritized over age matching. For within-index matches, sex was not considered in order to prioritize matching on the shared experience of exposure to the same index case over demographic similarity.

Irrespective of index matching, the closest available match on age was sought in each pair. All children 12 years of age and younger in the resister sample were matched on exact age (by year) with their respective control due to greater age-dependent variability in the metabolome in younger children [3]. Adolescents 13-20 years of age were not matched on exact age but were matched to other adolescents within the same age range. Matches on age among adult participants were made using 10-year age categories, with the lowest category representing individuals aged 21-30 years and the highest representing those aged 70 years or older. When a suitable match within the same 10-year age category was not possible, a matched control was sought from the closest adjacent age category to the resister's.

When within-index matching was not possible, matches between resisters and controls with different index cases were made on age, sex, and study cohort/site where possible, with age prioritized over sex in the matching process. All matches were restricted to participants from study sites within the same country; however, when a suitable match was not available within the same cohort, matches were allowed between resisters and controls from different cohorts enrolled in the same country.

Ethical Considerations

This study was approved by the institutional review boards at the following institutions: Emory University, Johns Hopkins University, Vanderbilt University, Rutgers University, Commisão Nacional De Ética em Pesquisa (CONEP), University of Witswatersrand, Byramjee Jeejeebhoy Government Medical College, Jawaharlal Institute of Postgraduate Medical Education and Research, Bhagwan Mahavir Medical Research Centre, and National Institute for Research in Tuberculosis (Chennai).

**Supplemental Table 1.** Categories of Likelihood of Being Resistant to *Mtb* Infection and Having *Mtb* Infection by Country.

| **Hierarchy** | **QFT/TST (Final)** | **Contact Exposure Level** | **Index infectiousness** | **Total N=4000 N (%)** | **Brazil N=1557 N (%)** | **India N=1655 N (%)** | **South Africa N=788 N (%)** |
|---|---|---|---|---|---|---|---|
| Resister A | IGRA- & TST- (>90 days) | High | High | 35 (0.9%) | 0 ( 0%) | 35 (2.1%) | 0 ( 0%) |
| Resister B | IGRA- or TST- (>90 days) | High | High | 317 (7.9%) | 226 (15%) | 79 (4.8%) | 12 (1.5%) |
| Resister C | IGRA- or TST- (<90 days) | High | High | 120 (3%) | 5 (0.3%) | 109 (6.6%) | 6 (0.8%) |
| Uninfected A | IGRA- or TST- (any time) | Low | Medium/Low | 671 (16.7%) | 337 (22%) | 278 (17%) | 56 (7.1%) |
| Uninfected B | IGRA- or TST- (any time) | Unknown or Missing | Unknown or Missing | 504 (12.6%) | 222 (14%) | 36 (2.2%) | 246 (31%) |
| Discordant | IGRA+ & TST- or IGRA- & TST+ | Any/Unk | Any/Unk | 364 (9.1%) | 11 (0.7%) | 332 (20%) | 21 (2.6%) |
| Infected B | IGRA+ OR TST+ | Any/Unk | Any/Unk | 1432 (35.8%) | 720 (46%) | 384 (23%) | 328 (42%) |
| Infected A | IGRA+ & TST+ | Any/Unk | Any/Unk | 557 (13.9%) | 36 (2.3%) | 402 (24%) | 119 (15%) |

**Supplemental Table 2.** Results Summary of the Untargeted Metabolome-wide Association Study Comparing TB Resisters and Controls.

| Statistical Significance Threshold | Number of Metabolic Features | |
|---|---|---|
| | HILIC positive | C18 negative |
| **All Cohorts (269 pairs)** | | |
| $P < 0.05$ | 911 | 876 |
| FDR Corrected $q < 0.2$ | 2 | 0 |
| **Brazil (155 pairs)** | | |
| $P < 0.05$ | 732 | 882 |
| FDR Corrected $q < 0.2$ | 0 | 0 |
| **India (103 pairs)** | | |
| $P < 0.05$ | 750 | 612 |
| FDR Corrected $q < 0.2$ | 0 | 0 |
| **South Africa (11 pairs)** | | |
| $P < 0.05$ | 555 | 485 |
| FDR Corrected $q < 0.2$ | 1 | 0 |

FDR, false discover rate.

**Supplemental Table 3.** Results of the Untargeted Metabolome-wide Association Study Comparing TB Resisters and Controls in the Overall Cohort (269 Pairs). Metabolites with P Value < 0.05 and Level 1 or 2 Confirmation are Shown.

| Platform | m/z | time | Resister Mean (SD) | Control Mean (SD) | Difference Resister vs. Control (95% CI) | P | FDR | Level 2 Confirmation | | | Level 1 Confirmation | |
|---|---|---|---|---|---|---|---|---|---|---|---|---|
| | | | | | | | | Name | HMDB ID | Adduct | Name | Adduct |
| HILIC positive | 463.3411 | 27.2 | -0.16 (1.02) | 0.16 (0.95) | -0.32 (-0.47, -0.17) | 0.00002 | 0.1483 | Dolichosterone; Polyporusterone F | HMDB34336; HMDB38499 | M+H | - | - |
| HILIC positive | 579.4969 | 26.9 | -0.15 (1.14) | 0.15 (0.82) | -0.29 (-0.45, -0.14) | 0.0002 | 0.3164 | DG(15:018:2(9Z12Z)0:0); DG(18:2(9Z12Z)15:00:0); Glycerol triundecanoate; DG(14:00:018:2n6); DG(14:00:020:2n6) | HMDB07074; HMDB07242; HMDB31089; HMDB55967; HMDB55969 | M+H; M+H-H2O | - | - |
| HILIC positive | 559.4711 | 26.8 | -0.13 (1.14) | 0.13 (0.82) | -0.27 (-0.41, -0.13) | 0.0003 | 0.3164 | DG(15:018:3(6Z9Z12Z)0:0); DG(15:018:3(9Z12Z15Z)0:0); DG(18:3(6Z9Z12Z)15:00:0); DG(18:3(9Z12Z15Z)15:00:0); DG(15:00:018:3n6); DG(15:00:018:3n3); DG(14:1n50:018:2n6); DG(14:1n50:020:2n6) | HMDB07075; HMDB07076; HMDB07271; HMDB07300; HMDB55996; HMDB56003; HMDB56142; HMDB56144 | M+H-H2O | - | - |
| HILIC positive | 517.3881 | 26.8 | -0.14 (1.07) | 0.14 (0.91) | -0.29 (-0.44, -0.13) | 0.0005 | 0.3164 | 22-Acetylpriverogenin B | HMDB34645 | M+H | - | - |
| HILIC positive | 280.1754 | 250.7 | 0.15 (0.74) | -0.15 (1.19) | 0.31 (0.14, 0.48) | 0.0005 | 0.3164 | Etamiphylline | HMDB41889 | M+H | - | - |
| HILIC positive | 251.2005 | 26.9 | -0.13 (1.11) | 0.13 (0.86) | -0.25 (-0.39, -0.11) | 0.0007 | 0.3164 | Norambreinolide; 3-Methyl-alpha-ionyl acetate; 22-Bis(2-methylpropoxy)ethylbenzene | HMDB35293; HMDB37631; HMDB37712 | M+H | - | - |
| HILIC positive | 132.0565 | 74.5 | -0.12 (0.97) | 0.12 (1.02) | -0.24 (-0.38, -0.1) | 0.0007 | 0.3164 | Pyroglutamic acid; Pyrrolidonecarboxylic acid; Pyrroline hydroxycarboxylic acid; N-Acryloylglycine; 1-Pyrroline-4-hydroxy-2-carboxylate; 5-Oxoprolinate; dimethadione | HMDB00267; HMDB00805; HMDB01369; HMDB01843; HMDB02234; HMDB60262; HMDB61093 | M+H_[+2] | - | - |
| HILIC positive | 675.5427 | 35.1 | 0.13 (0.99) | -0.13 (0.99) | 0.27 (0.11, 0.43) | 0.0007 | 0.3164 | SM(d18:114:0) | HMDB12097 | M+H | - | - |
| HILIC positive | 592.4932 | 26.7 | -0.14 (1.17) | 0.14 (0.78) | -0.27 (-0.43, -0.11) | 0.0008 | 0.3164 | DG(15:018:4(6Z9Z12Z15Z)0:0); DG(18:4(6Z9Z12Z15Z)15:00:0); DG(15:00:018:4n3) | HMDB07077; HMDB07329; HMDB56004 | M+NH4 | - | - |
| HILIC positive | 151.024 | 90.6 | -0.13 (1.06) | 0.13 (0.92) | -0.27 (-0.42, -0.11) | 0.0008 | 0.3164 | Tartaric acid; D-Tartaric acid; 2-Methyl-2-(methyldithio)propanal; 23-Dihydroxybutanedioic acid | HMDB00956; HMDB29878; HMDB38889; HMDB59916 | M+H | - | - |
| HILIC positive | 501.3932 | 26.8 | -0.13 (1.08) | 0.13 (0.9) | -0.26 (-0.41, -0.11) | 0.0008 | 0.3164 | Norambreinolide; 3-Methyl-alpha-ionyl acetate; 22-Bis(2-methylpropoxy)ethylbenzene; Ganoderiol C | HMDB35293; HMDB37631; HMDB37712; HMDB37781 | 2M+H; M+H-H2O | - | - |
| HILIC positive | 491.3725 | 27.4 | -0.14 (1.15) | 0.14 (0.79) | -0.28 (-0.45, -0.12) | 0.0008 | 0.3164 | Barringtogenol C; Camelliagenin C; Ganoderiol H | HMDB34525; HMDB34532; HMDB37783 | M+H | - | - |
| C18 negative | 515.3742 | 230 | -0.14 (1.06) | 0.14 (0.92) | -0.28 (-0.44, -0.11) | 0.0010 | 0.4498 | 22-Acetylpriverogenin B | HMDB34645 | M-H | - | - |
| C18 negative | 325.311 | 294.4 | 0.14 (0.68) | -0.14 (1.23) | 0.27 (0.11, 0.43) | 0.0011 | 0.4498 | Homophytanic acid; Heneicosanoic acid; 2371115-Pentamethylhexadecanoic acid; 6-Hydroxy-8-heneicosanone; 8-Hydroxy-6-heneicosanone; 4-Hydroxy-6-heneicosanone; 6-Hydroxy-4-heneicosanone | HMDB02337; HMDB02345; HMDB31319; HMDB35668; HMDB35669; HMDB35670; HMDB35671 | M-H | - | - |
| HILIC positive | 241.1546 | 56.1 | -0.13 (1.17) | 0.13 (0.78) | -0.26 (-0.42, -0.11) | 0.0011 | 0.3622 | Pirbuterol | HMDB15407 | M+H | - | - |
| C18 negative | 477.3219 | 178.9 | -0.13 (1.12) | 0.13 (0.85) | -0.25 (-0.4, -0.1) | 0.0012 | 0.4652 | Dolicholide; Polyporusterone A | HMDB34086; HMDB38495 | M-H | - | - |
| HILIC positive | 193.1224 | 107.2 | 0.13 (0.55) | -0.13 (1.29) | 0.27 (0.1, 0.43) | 0.0014 | 0.3917 | 2-Methyl-1-phenyl-2-propanyl acetate; 4-Phenyl-2-butyl acetate; Ethyl 4-phenylbutanoate; Benzyl 3-methylbutanoate; 3-Methylbutyl benzoate; 2-Methylpropyl phenylacetate; 2-Phenylethyl butanoate; 2-Phenylethyl 2-methylpropanoate; 5-Isopropyl-2-methylphenol acetate; 3-Phenylpropyl propanoate; 111213-Trinor-1(10)-spirovetivene-27-dione; 4-Methyl-2-(1-phenylethyl)-13-dioxolane 9CI; 4-Methylphenyl 3-methylbutanoate; 1-Phenylethyl isobutyrate; 1-Phenylethyl butyrate; 1-Ethoxy-2-methoxy-4-(1-propenyl)benzene; 2-(2-Hydroxy-4-methylphenyl)-3-pentanone; 2-Ethoxy-1-methoxy-4-(1-propenyl)benzene; 111213-Trinor-135-bisabolatrien-10-oic acid; 2-Benzyl-45-dimethyl-13-dioxolane; Butyl phenylacetate | HMDB31571; HMDB31614; HMDB31618; HMDB32043; HMDB33380; HMDB35010; HMDB35014; HMDB35015; HMDB35447; HMDB36388; HMDB36734; HMDB37165; HMDB37709; HMDB37716; HMDB37717; HMDB37792; HMDB38180; HMDB39496; HMDB39643; HMDB39841; HMDB40427 | M+H | - | - |
| C18 negative | 491.3378 | 179.4 | -0.12 (1.07) | 0.12 (0.92) | -0.23 (-0.37, -0.09) | 0.0014 | 0.4652 | Homodolicholide | HMDB33907 | M-H | - | - |
| C18 negative | 412.3066 | 180.3 | -0.13 (0.99) | 0.13 (1) | -0.25 (-0.41, -0.1) | 0.0015 | 0.4652 | 3-Hydroxy-9-hexadecenoylcarnitine | HMDB13333 | M-H | - | - |
| HILIC positive | 279.2315 | 26.9 | -0.13 (1.11) | 0.13 (0.86) | -0.25 (-0.41, -0.1) | 0.0016 | 0.4010 | Alpha-Linolenic acid; Gamma-Linolenic acid; Calendic acid; Punicic acid; Linolenelaidic acid | HMDB01388; HMDB03073; HMDB30962; HMDB30963; HMDB30964 | M+H | - | - |
| C18 negative | 158.0096 | 294.2 | 0.14 (0.95) | -0.14 (1.04) | 0.27 (0.1, 0.44) | 0.0016 | 0.4652 | (E)-4-Isothiocyanato-1-(methylthio)-1-butene; (E)-Raphanusanin | HMDB31576; HMDB41083 | M-H | - | - |
| C18 negative | 165.0408 | 288.3 | 0.14 (0.63) | -0.14 (1.25) | 0.28 (0.11, 0.45) | 0.0016 | 0.4652 | Arabinonic acid; Ribonic acid; S-Propyl 1-propanesulfinothioate; 2345-Tetrahydroxypentanoic acid; L-Lyxonate; L-Xylonate | HMDB00539; HMDB00867; HMDB34394; HMDB59750; HMDB60255; HMDB60256 | M-H | - | - |

| | | | | | | | | | | | | |
|---|---|---|---|---|---|---|---|---|---|---|---|---|
| HILIC positive | 597.4484 | 25.7 | -0.11 (1.15) | 0.11 (0.82) | -0.22 (-0.36, -0.08) | 0.0017 | 0.4010 | DG(15:018:4(6Z9Z12Z15Z)0:0); DG(18:4(6Z9Z12Z15Z)15:00:0); DG(15:00:018:4n3) | HMDB07077; HMDB07329; HMDB56004 | M+Na | - | - |
| HILIC positive | 127.1117 | 295.5 | 0.13 (0.42) | -0.13 (1.34) | 0.26 (0.1, 0.43) | 0.0019 | 0.4010 | (E)-2-octenal; 2-Octenal; (3xi5Z)-15-Octadien-3-ol; 244-Trimethylcyclopentanone; 2-Octen-4-one; 1-Octen-3-one; 3-Methyl-3-hepten-2-one; 5-Methyl-5-hepten-2-one; 6-Methyl-3-hepten-2-one trans-; 4-Octen-3-one; 3-Octen-2-one; (SE)-Filbertone; 5-Octen-2-one; Sulcatone; 2-Ethylidenehexanal; 6-Octenal; (Z)-5-Octenal; 25-Octadien-1-ol; (3E5Z)-35-Octadien-1-ol; (2E4E)-24-Octadien-1-ol | HMDB13809; HMDB30961; HMDB30966; HMDB31197; HMDB31301; HMDB31309; HMDB31590; HMDB31591; HMDB32403; HMDB32451; HMDB33547; HMDB35242; HMDB35390; HMDB35915; HMDB37152; HMDB39769; HMDB39770; HMDB40146; HMDB40149; HMDB40151 | M+H | - | - |
| HILIC positive | 146.1176 | 45.6 | -0.12 (1.11) | 0.12 (0.87) | -0.23 (-0.38, -0.09) | 0.0019 | 0.4010 | 3-Dehydroxycarnitine | HMDB06831 | M+H | Acetylcholine | M+H |
| HILIC positive | 541.4613 | 26.4 | -0.12 (1.06) | 0.12 (0.93) | -0.24 (-0.39, -0.09) | 0.0019 | 0.4010 | Momordicilin | HMDB30896 | M+H | - | - |
| C18 negative | 138.0196 | 191.2 | -0.14 (1.25) | 0.14 (0.63) | -0.27 (-0.44, -0.1) | 0.0020 | 0.4652 | 4-Nitrophenol; 6-Hydroxynicotinic acid; 3-Hydroxypicolinic acid; 2-Hydroxynicotinic acid | HMDB01232; HMDB02658; HMDB13188; HMDB59710 | M-H | - | - |
| HILIC positive | 575.466 | 26.9 | -0.11 (1.14) | 0.11 (0.83) | -0.23 (-0.37, -0.08) | 0.0021 | 0.4232 | DG(15:018:4(6Z9Z12Z15Z)0:0); DG(18:4(6Z9Z12Z15Z)15:00:0); DG(15:00:018:4n3) | HMDB07077; HMDB07329; HMDB56004 | M+H | - | - |
| C18 negative | 203.1804 | 171.7 | 0.12 (0.87) | -0.12 (1.1) | 0.24 (0.09, 0.4) | 0.0023 | 0.4652 | delta-Amorphene; (-)-35-Cadinadiene; alpha-Santalene; Sesquisabinene; beta-Selinene; ()-1(10)4-Cadinadiene; Aristolene; 2710-Bisabolatriene; Sesquithujene; Daucene; Valencene; (2Z4E6E)-24610-Farnesatetraene; alpha-Selinene; (-)-beta-Elemene; beta-Farnesene; beta-Bisabolene; alpha-Guaiene; alpha-Cyperene; alpha-Farnesene; (3E6E)-alpha-Farnesene; (Z)-2610-Bisabolatriene; beta-Sesquiphellandrene; Zingiberene; (R)-2511-Bisabolatriene; beta-Santalene; alpha-Cubebene; 1-Epi-alpha-gurjunene; (-)-Aromadendrene; (1beta4alpha5beta6alpha7alpha)-9-Aromadendrene; (4alpha5beta6alpha7alpha10alpha)-1-Aromadendrene; alpha-Bulnesene; (4alpha10alpha)-1(5)6-Guaiadiene; alpha-Humulene; Germacrene B; beta-Bulnesene; alpha-Bergamotene; Cascarilladiene; beta-Neoclovene; beta-Patchoulene; delta-Patchoulene; beta-Caryophyllene; Cycloseychellene; gamma1-Cadinene; 1-Epi-bicyclosesquiphellandrene; 37(11)-Eudesmadiene; (-)-Tamariscene; ()-1(9)10-Pacifigorgiadiene; (-)-1(6)10-Pacifigorgiadiene; ()-alpha-Muurolene; gamma-Selinene; beta-Panasinsene; alpha-Panasinsene; Bicyclogermacrene; epsilon-Muurolene; beta-Bourbonene; beta-Guaiene; Bicycloelemene; gamma-Himachalene; omega-Cadinene; alpha-Cadinene; gamma-Muurolene; gamma-Cadinene; beta-Humulene; beta-Cadinene; Benzylalcohol alpha-isobutyl-246-trimethyl(1-mestyl-3-methyl-1-butanol); alpha-Cedrene; alpha-Alaskene | HMDB30644; HMDB30673; HMDB34939; HMDB34943; HMDB34997; HMDB35084; HMDB35146; HMDB35161; HMDB35198; HMDB35361; HMDB35688; HMDB35787; HMDB35810; HMDB35816; HMDB35913; HMDB35992; HMDB36003; HMDB36004; HMDB36065; HMDB36066; HMDB36154; HMDB36163; HMDB36164; HMDB36200; HMDB36364; HMDB36413; HMDB36417; HMDB36418; HMDB36419; HMDB36428; HMDB36444; HMDB36450; HMDB36467; HMDB36575; HMDB36644; HMDB36678; HMDB36682; HMDB36731; HMDB36736; HMDB36737; HMDB36792; HMDB36799; HMDB37029; HMDB37030; HMDB37060; HMDB37676; HMDB37677; HMDB37678; HMDB37784; HMDB38118; HMDB38124; HMDB38135; HMDB38137; HMDB38139; HMDB38155; HMDB38157; HMDB38166; HMDB38168; HMDB38203; HMDB38205; HMDB38206; HMDB38207; HMDB38214; HMDB41576; HMDB59689; HMDB59695; HMDB59818 | M-H | - | - |
| HILIC positive | 121.0509 | 272.4 | 0.13 (0.81) | -0.13 (1.14) | 0.26 (0.09, 0.43) | 0.0024 | 0.4568 | Purine | HMDB01366 | M+H | - | - |
| HILIC positive | 280.2351 | 27 | -0.12 (1.12) | 0.12 (0.84) | -0.24 (-0.4, -0.09) | 0.0024 | 0.4568 | Alpha-Linolenic acid; Gamma-Linolenic acid; Calendic acid; Punicic acid; Linolenelaidic acid | HMDB01388; HMDB03073; HMDB30962; HMDB30963; HMDB30964 | M+H_[+1] | - | - |
| C18 negative | 473.3633 | 250.6 | -0.11 (1.17) | 0.11 (0.78) | -0.21 (-0.35, -0.08) | 0.0024 | 0.4652 | Soyasapogenol A; Camelliagenin A; Priverogenin B; (3alphaOH20S24S)-319:2024-Diepoxydammarane-325-diol; Ganoderiol A; 2024-Epoxy-2526-dihydroxydammaran-3-one | HMDB34505; HMDB34528; HMDB34644; HMDB34683; HMDB35326; HMDB39692 | M-H | - | - |
| C18 negative | 415.3217 | 227.9 | -0.11 (1.09) | 0.11 (0.89) | -0.23 (-0.37, -0.08) | 0.0025 | 0.4652 | 2425-Dihydroxyvitamin D; 2526-dihydroxyvitamin D; Calcitriol; 7a12a-Dihydroxy-cholestene-3-one; 24R25-Dihydroxyvitamin D3; 23S25-dihydroxyvitamin D3; 3 beta-Hydroxy-5-cholestenoate; 7 alpha24-Dihydroxy-4-cholesten-3-one; 7 alpha26-Dihydroxy-4-cholesten-3-one; Paricalcitol; Sarsasapogenin; Neotigogenin; Smilagenin; Octadecyl cis-p- | HMDB00430; HMDB01420; HMDB01903; HMDB02197; HMDB06226; HMDB06720; HMDB12453; HMDB12457; HMDB12459; HMDB15046; HMDB30024; HMDB30047; | M-H | - | - |

| | | | | | | | | | | | | |
|---|---|---|---|---|---|---|---|---|---|---|---|---|
| | | | | | | | | coumarate; (24R)-2425-Dihydroxycalciol; 7alpha25-Dihydroxy-4-cholesten-3-one; Secalciferol | HMDB30048; HMDB35300; HMDB60133; HMDB60425; HMDB60508 | | | |
| C18 negative | 605.3332 | 139.1 | -0.12 (1) | 0.12 (0.98) | -0.25 (-0.41, -0.09) | 0.0028 | 0.4652 | (23S)-2325-dihdroxy-24-oxovitamine D3 23-(beta-glucuronide) | HMDB10361 | M-H | - | - |
| C18 negative | 209.0666 | 26 | -0.12 (1.07) | 0.12 (0.91) | -0.24 (-0.4, -0.08) | 0.0029 | 0.4778 | Sedoheptulose; L-Galacto-2-heptulose; D-Manno-2-heptulose; D-altro-D-manno-Heptose | HMDB03219; HMDB29901; HMDB29935; HMDB29952 | M-H | - | - |
| C18 negative | 861.5478 | 226.6 | -0.12 (1.04) | 0.12 (0.95) | -0.23 (-0.38, -0.08) | 0.0030 | 0.4779 | PI(16:020:2(11Z14Z)); PI(18:018:2(9Z12Z)); PI(18:1(11Z)18:1(11Z)); PI(18:1(11Z)18:1(9Z)); PI(18:1(9Z)18:1(11Z)); PI(18:1(9Z)18:1(9Z)); PI(18:2(9Z12Z)18:0); PI(20:2(11Z14Z)16:0) | HMDB09786; HMDB09809; HMDB09824; HMDB09825; HMDB09836; HMDB09837; HMDB09847; HMDB09875 | M-H | - | - |
| C18 negative | 239.1652 | 147.7 | 0.12 (0.96) | -0.12 (1.02) | 0.25 (0.09, 0.41) | 0.0030 | 0.4779 | L-Menthyl acetoacetate | HMDB32369 | M-H | - | - |
| HILIC positive | 597.5086 | 26.8 | -0.12 (1.1) | 0.12 (0.87) | -0.24 (-0.39, -0.08) | 0.0031 | 0.4853 | 3-Oxooctadecanoic acid; 9-Oxooctadecanoic acid; 10-Oxooctadecanoic acid; 11-Oxooctadecanoic acid; 5-Hexyltetrahydro-2-furanoctanoic acid; 5-Oxooctadecanoic acid; Ricinoleic acid; Glycerol triundecanoate | HMDB10736; HMDB30979; HMDB30980; HMDB30981; HMDB31127; HMDB34074; HMDB34297; HMDB31089 | 2M+H; M+H | - | - |
| HILIC positive | 599.4646 | 25.6 | -0.12 (1.06) | 0.12 (0.92) | -0.23 (-0.39, -0.08) | 0.0033 | 0.4853 | DG(15:018:3(6Z9Z12Z)0:0); DG(15:018:3(9Z12Z15Z)0:0); DG(18:3(6Z9Z12Z)15:00:0); DG(18:3(9Z12Z15Z)15:00:0); DG(15:00:018:3n6); DG(15:00:018:3n3); DG(14:1n50:018:2n6); DG(14:1n50:020:2n6) | HMDB07075; HMDB07076; HMDB07271; HMDB07300; HMDB55996; HMDB56003; HMDB56142; HMDB56144 | M+Na | - | - |
| HILIC positive | 405.3721 | 26.8 | -0.12 (1.01) | 0.12 (0.98) | -0.24 (-0.39, -0.08) | 0.0033 | 0.4853 | 3a7a-Dihydroxy-5b-cholestane; 5-Heneicosyl-13-benzenediol; Lepidiumterpenoid | HMDB06893; HMDB31035; HMDB36844 | M+H | - | - |
| HILIC positive | 184.0372 | 96.4 | -0.11 (1.05) | 0.11 (0.93) | -0.22 (-0.37, -0.07) | 0.0034 | 0.4853 | Dipropyl trisulfide; Ethyl 1-(ethylthio)ethyl disulfide; Ethyl 1-(methylthio)propyl disulfide; 11-Thiobis-1-propanethiol | HMDB32825; HMDB33055; HMDB39654; HMDB40235 | M+H_[+1] | - | - |
| HILIC positive | 450.3211 | 29.6 | -0.12 (1.08) | 0.12 (0.9) | -0.24 (-0.4, -0.08) | 0.0034 | 0.4853 | Deoxycholic acid glycine conjugate; Chenodeoxycholic acid glycine conjugate; Glycoursodeoxycholic acid; Chenodeoxyglycocholic acid | HMDB00631; HMDB00637; HMDB00708; HMDB06898 | M+H | - | - |
| HILIC positive | 315.2528 | 25.9 | -0.12 (1.06) | 0.12 (0.92) | -0.25 (-0.41, -0.08) | 0.0037 | 0.4853 | Octadecanedioic acid; 910-DHOME; 1213-DHOME; (9xi10xi12xi)-910-Dihydroxy-12-octadecenoic acid; Dibutyl decanedioate | HMDB00782; HMDB04704; HMDB04705; HMDB31679; HMDB41220 | M+H | - | - |
| HILIC positive | 188.1758 | 65.2 | -0.12 (1.09) | 0.12 (0.89) | -0.25 (-0.41, -0.08) | 0.0037 | 0.4853 | N1-Acetylspermidine; N8-Acetylspermidine; N(8)-Acetylspermidinium | HMDB01276; HMDB02189; HMDB60288 | M+H | - | - |
| C18 negative | 199.0378 | 22.8 | -0.12 (1.05) | 0.12 (0.93) | -0.23 (-0.39, -0.08) | 0.0038 | 0.4928 | Potassium 2-(1-ethoxy) ethoxypropanoate | HMDB37192 | M-H | - | - |
| C18 negative | 463.343 | 215.7 | -0.12 (1.14) | 0.12 (0.83) | -0.24 (-0.41, -0.08) | 0.0040 | 0.4928 | Trihydroxycoprostanoic acid; 2-Deoxybrassinolide | HMDB02163; HMDB41135 | M-H | - | - |
| C18 negative | 165.0556 | 160.3 | -0.12 (1.05) | 0.12 (0.94) | -0.25 (-0.42, -0.08) | 0.0041 | 0.4928 | 3-(3-Hydroxyphenyl)propanoic acid; D-Phenyllactic acid; L-3-Phenyllactic acid; Phenyllactic acid; 4-Methoxyphenylacetic acid; Desaminotyrosine; 3-Phenoxypropionic acid; Homovanillin; Ethyl vanillin; Ethyl salicylate; 34-Dihydroxyphenylacetone; Guaicyl acetate; 34-Dimethoxybenzaldehyde; Ethylparaben; Methyl 2-methoxybenzoate; Methyl 4-methoxybenzoate; 3-(2-Hydroxyphenyl)propanoic acid; Ipomeanine; 4-Methoxybenzyl formate; 3-Hydroxy-1-(4-hydroxyphenyl)-1-propanone; Ethyl 2-furanacrylate; 4-Hydroxyphenyl-2-propionic acid; 3-Methoxyphenylacetic acid | HMDB00375; HMDB00563; HMDB00748; HMDB00779; HMDB02072; HMDB02199; HMDB02229; HMDB05175; HMDB29665; HMDB29817; HMDB31132; HMDB32030; HMDB32138; HMDB32573; HMDB32605; HMDB32639; HMDB33752; HMDB34243; HMDB34993; HMDB40645; HMDB41611; HMDB41683; HMDB59969 | M-H | - | - |
| HILIC positive | 108.0808 | 31.4 | -0.11 (1.04) | 0.11 (0.94) | -0.22 (-0.38, -0.07) | 0.0042 | 0.5110 | 3-Ethylpyridine; 24-Dimethylpyridine; 26-Dimethylpyridine; Benzylamine; O-Toluidine | HMDB29734; HMDB32244; HMDB32972; HMDB33871; HMDB41965 | M+H | - | - |
| HILIC positive | 314.1037 | 282.4 | -0.1 (1.04) | 0.1 (0.95) | -0.2 (-0.34, -0.06) | 0.0042 | 0.5110 | Avenanthramide 1f; Avenanthramide E | HMDB29283; HMDB38944 | M+H | - | - |
| C18 negative | 862.5536 | 227.6 | -0.11 (1.03) | 0.11 (0.96) | -0.22 (-0.37, -0.07) | 0.0043 | 0.4928 | PI(16:020:2(11Z14Z)); PI(18:018:2(9Z12Z)); PI(18:1(11Z)18:1(11Z)); PI(18:1(11Z)18:1(9Z)); PI(18:1(9Z)18:1(11Z)); PI(18:1(9Z)18:1(9Z)); PI(18:2(9Z12Z)18:0); PI(20:2(11Z14Z)16:0) | HMDB09786; HMDB09809; HMDB09824; HMDB09825; HMDB09836; HMDB09837; HMDB09847; HMDB09875 | M-H_[-1] | - | - |
| C18 negative | 570.2769 | 155.3 | 0.11 (0.83) | -0.11 (1.13) | 0.23 (0.07, 0.38) | 0.0044 | 0.4933 | Ganoderic acid F; Kinetensin 4-7 | HMDB35988; HMDB12986 | M-H_[-1]; M-H | - | - |
| HILIC positive | 241.1065 | 214.1 | 0.12 (0.61) | -0.12 (1.27) | 0.25 (0.08, 0.42) | 0.0044 | 0.5233 | 3-(345-Trimethoxyphenyl)propanoic acid; Ethyl 345-trimethoxybenzoate; Isopropyl 3-(34-dihydroxyphenyl)-2-hydroxypropanoate; 3-Carboxy-4-methyl-5-propyl-2-furanpropionic acid | HMDB30254; HMDB38627; HMDB41756; HMDB61112 | M+H | - | - |
| C18 negative | 313.1655 | 292.7 | -0.12 (1.04) | 0.12 (0.94) | -0.24 (-0.4, -0.07) | 0.0049 | 0.4940 | (-)-trans-Carveol glucoside; ()-trans-Carveol glucoside; Perilloside A | HMDB29848; HMDB29849; HMDB38706 | M-H | - | - |
| HILIC positive | 250.0642 | 48.6 | -0.09 (1.06) | 0.09 (0.92) | -0.17 (-0.29, -0.05) | 0.0051 | 0.5312 | Sulfapyridine | HMDB15028 | M+H | - | - |
| C18 negative | 210.07 | 25.6 | -0.11 (1.15) | 0.11 (0.81) | -0.23 (-0.39, -0.07) | 0.0051 | 0.4940 | 1-Hydroxylorcaserin; 5-Hydroxylorcaserin; 7-Hydroxylorcaserin; N-Hydroxylorcaserin; 4-(4-chlorophenyl)-4-hydroxypiperidine | HMDB60704; HMDB60778; HMDB60800; HMDB60844; HMDB60902 | M-H | - | - |

| | | | | | | | | | | | | |
|---|---|---|---|---|---|---|---|---|---|---|---|---|
| HILIC positive | 601.4808 | 25.4 | -0.11 (1.09) | 0.11 (0.89) | -0.22 (-0.37, -0.07) | 0.0053 | 0.5312 | DG(15:020:5(5Z8Z11Z14Z17Z)0:0); DG(20:5(5Z8Z11Z14Z17Z)15:00:0); DG(15:00:020:5n3) | HMDB07085; HMDB07561; HMDB56006 | M+H | - | - |
| HILIC positive | 577.4812 | 27 | -0.11 (1.14) | 0.11 (0.82) | -0.22 (-0.37, -0.06) | 0.0055 | 0.5312 | DG(15:018:3(6Z9Z12Z)0:0); DG(15:018:3(9Z12Z15Z)0:0); DG(18:3(6Z9Z12Z)15:00:0); DG(18:3(9Z12Z15Z)15:00:0); DG(15:00:018:3n6); DG(15:00:018:3n3); DG(14:1n50:018:2n6); DG(14:1n50:020:2n6) | HMDB07075; HMDB07076; HMDB07271; HMDB07300; HMDB55996; HMDB56003; HMDB56142; HMDB56144 | M+H | - | - |
| C18 negative | 113.989 | 34.7 | -0.11 (1.06) | 0.11 (0.92) | -0.22 (-0.37, -0.06) | 0.0055 | 0.4989 | Trifluoroacetic acid | HMDB14118 | M-H_[-1] | - | - |
| HILIC positive | 87.0934 | 46.4 | -0.12 (1.01) | 0.12 (0.97) | -0.24 (-0.41, -0.07) | 0.0057 | 0.5325 | Piperidine | HMDB34301 | M+H_[+1] | - | - |
| HILIC positive | 854.5693 | 29.3 | 0.11 (0.71) | -0.11 (1.22) | 0.22 (0.06, 0.38) | 0.0061 | 0.5416 | PC(20:4(5Z8Z11Z14Z)22:6(4Z7Z10Z13Z16Z19Z)); PC(20:4(8Z11Z14Z17Z)22:6(4Z7Z10Z13Z16Z19Z)); PC(20:5(5Z8Z11Z14Z17Z)22:5(4Z7Z10Z13Z16Z)); PC(20:5(5Z8Z11Z14Z17Z)22:5(7Z10Z13Z16Z19Z)); PC(22:5(4Z7Z10Z13Z16Z)20:5(5Z8Z11Z14Z17Z)); PC(22:5(7Z10Z13Z16Z19Z)20:5(5Z8Z11Z14Z17Z)); PC(22:6(4Z7Z10Z13Z16Z19Z)20:4(5Z8Z11Z14Z)); PC(22:6(4Z7Z10Z13Z16Z19Z)20:4(8Z11Z14Z17Z)) | HMDB08452; HMDB08485; HMDB08516; HMDB08517; HMDB08675; HMDB08708; HMDB08739; HMDB08740 | M+H | - | - |
| C18 negative | 478.3253 | 178.6 | -0.1 (1.12) | 0.1 (0.86) | -0.21 (-0.35, -0.06) | 0.0062 | 0.5103 | Dolicholide; Polyporusterone A | HMDB34086; HMDB38495 | M-H_[-1] | - | - |
| C18 negative | 183.1026 | 25.1 | -0.12 (1.06) | 0.12 (0.93) | -0.24 (-0.41, -0.07) | 0.0064 | 0.5108 | (1R)-Nepetalic acid; (S)-Oleuropeic acid; (4S8R)-89-Dihydroxy-p-menth-1(6)-en-2-one; 1-Acetylcyclohexyl acetate; (E)-10-Oxo-8-decenoic acid | HMDB36117; HMDB36998; HMDB39052; HMDB39710; HMDB40883 | M-H | - | - |
| C18 negative | 860.5343 | 220.4 | -0.11 (1.06) | 0.11 (0.93) | -0.21 (-0.37, -0.06) | 0.0068 | 0.5108 | PI(16:020:3(5Z8Z11Z)); PI(16:020:3(8Z11Z14Z)); PI(18:018:3(6Z9Z12Z)); PI(18:018:3(9Z12Z15Z)); PI(18:1(11Z)18:2(9Z12Z)); PI(18:1(9Z)18:2(9Z12Z)); PI(18:2(9Z12Z)18:1(11Z)); PI(18:2(9Z12Z)18:1(9Z)); PI(18:3(6Z9Z12Z)18:0); PI(18:3(9Z12Z15Z)18:0); PI(20:3(5Z8Z11Z)16:0); PI(20:3(8Z11Z14Z)16:0) | HMDB09787; HMDB09788; HMDB09810; HMDB09811; HMDB09826; HMDB09838; HMDB09848; HMDB09849; HMDB09858; HMDB09862; HMDB09879; HMDB09886 | M-H_[-1] | - | - |
| C18 negative | 105.0193 | 20.2 | 0.11 (1.03) | -0.11 (0.96) | 0.22 (0.06, 0.37) | 0.0069 | 0.5108 | Glyceric acid; L-Glyceric acid; D-23-Dihydroxypropanoic acid | HMDB00139; HMDB06372; HMDB31818 | M-H | Glyceric acid | M-H |
| C18 negative | 243.1235 | 295.2 | 0.11 (0.74) | -0.11 (1.2) | 0.23 (0.06, 0.39) | 0.0071 | 0.5108 | Polyethylene oxidized | HMDB32472 | M-H | - | - |
| C18 negative | 174.056 | 25 | -0.12 (1.14) | 0.12 (0.83) | -0.24 (-0.42, -0.06) | 0.0077 | 0.5123 | Indoleacetic acid; 5-Hydroxyindoleacetaldehyde; Hydroxymethyl indol-3-yl ketone; 1-Methoxy-1H-indole-3-carboxaldehyde | HMDB00197; HMDB04073; HMDB38628; HMDB40972 | M-H | Indoleacet ate | M-H |
| C18 negative | 158.057 | 289 | 0.11 (0.99) | -0.11 (1) | 0.22 (0.06, 0.38) | 0.0083 | 0.5123 | L-Hexahydro-3-imino-124-oxadiazepine-3-carboxylic acid | HMDB30402 | M-H | - | - |
| C18 negative | 198.158 | 155.8 | 0.1 (0.78) | -0.1 (1.18) | 0.2 (0.05, 0.34) | 0.0085 | 0.5123 | 5-Dodecenoic acid; trans-Dodec-2-enoic acid; Methyl 10-undecenoate; Allyl nonanoate; xi-Dihydro-5-octyl-2(3H)-furanone; cis-4-Decenyl acetate; 26-Dimethyl-5-heptenal propyleneglycol acetal; 11-Dodecenoic acid; cis-3-Hexenyl hexanoate; ()-Citronellyl acetate; Citronellyl acetate; 55-Dibutyl-45-dihydro-2(3H)furanone; Rhodinyl acetate; Methyl 9-undecenoate; Ethyl 2-decenoate; 1-Ethenylhexyl butanoate; xi-5-Dodecanolide; 3-Heptyldihydro-5-methyl-2(3H)-furanone; 2-Octenyl butyrate; Hexyl 2E-hexenoate; 7-Hexyl-2-oxepanone; 2-Hexenyl hexanoate; Ethyl 4-decenoate; Hexyl 2-methyl-3-pentenoate; Hexyl 2-methyl-4-pentenoate; 11-Dimethoxy-37-dimethyl-26-octadiene; ()-Menthyl acetate; (37-Dimethyl-6-octenyl)oxyacetaldehyde | HMDB00529; HMDB10729; HMDB29585; HMDB29763; HMDB31683; HMDB32214; HMDB32235; HMDB32248; HMDB33378; HMDB34160; HMDB34844; HMDB36182; HMDB37186; HMDB37305; HMDB37329; HMDB37498; HMDB37742; HMDB37813; HMDB38081; HMDB38269; HMDB38895; HMDB38924; HMDB39220; HMDB40158; HMDB40163; HMDB40361; HMDB41264; HMDB41449 | M-H_[-1] | - | - |
| HILIC positive | 182.049 | 124 | -0.1 (0.88) | 0.1 (1.1) | -0.19 (-0.34, -0.05) | 0.0086 | 0.5886 | Acamprosate | HMDB14797 | M+H | - | - |
| C18 negative | 771.5178 | 224.5 | -0.11 (1.1) | 0.11 (0.88) | -0.21 (-0.37, -0.05) | 0.0088 | 0.5123 | PG(16:020:3(5Z8Z11Z)); PG(16:020:3(8Z11Z14Z)); PG(18:018:3(6Z9Z12Z)); PG(18:018:3(9Z12Z15Z)); PG(18:1(11Z)18:2(9Z12Z)); PG(18:1(9Z)18:2(9Z12Z)); PG(18:2(9Z12Z)18:1(11Z)); PG(18:2(9Z12Z)18:1(9Z)); PG(18:3(6Z9Z12Z)18:0); PG(18:3(9Z12Z15Z)18:0) | HMDB10578; HMDB10579; HMDB10606; HMDB10607; HMDB10620; HMDB10635; HMDB10648; HMDB10649; HMDB10662; HMDB10677 | M-H | - | - |
| HILIC positive | 414.3 | 29.5 | -0.11 (1.05) | 0.11 (0.93) | -0.22 (-0.38, -0.05) | 0.0093 | 0.5915 | Deoxycholic acid glycine conjugate; Chenodeoxycholic acid glycine conjugate; Glycoursodeoxycholic acid; Chenodeoxyglycocholic acid | HMDB00631; HMDB00637; HMDB00708; HMDB06898 | M+H-2H2O | - | - |
| C18 negative | 859.5313 | 220.8 | -0.11 (1.07) | 0.11 (0.92) | -0.21 (-0.37, -0.05) | 0.0093 | 0.5164 | PI(16:020:3(5Z8Z11Z)); PI(16:020:3(8Z11Z14Z)); PI(18:018:3(6Z9Z12Z)); PI(18:018:3(9Z12Z15Z)); PI(18:1(11Z)18:2(9Z12Z)); PI(18:1(9Z)18:2(9Z12Z)); PI(18:2(9Z12Z)18:1(11Z)); PI(18:2(9Z12Z)18:1(9Z)); PI(18:3(6Z9Z12Z)18:0); PI(18:3(9Z12Z15Z)18:0); PI(20:3(5Z8Z11Z)16:0); PI(20:3(8Z11Z14Z)16:0) | HMDB09787; HMDB09788; HMDB09810; HMDB09811; HMDB09826; HMDB09838; HMDB09848; HMDB09849; HMDB09858; HMDB09862; HMDB09879; HMDB09886 | M-H | - | - |
| C18 negative | 163.0612 | 25.8 | -0.11 (1.09) | 0.11 (0.89) | -0.21 (-0.37, -0.05) | 0.0093 | 0.5164 | L-Fucose; Rhamnose; 15-Anhydrosorbitol; Beta-D-Fucose; 3-Deoxyfructose; L-Rhamnulose; 2-Deoxygalactopyranose; D-Fucose; 2-O-Methyl-D-xylose; Alpha-L-fucose; beta-L-fucose; L-Fuculose | HMDB00174; HMDB00849; HMDB02712; HMDB03081; HMDB05876; HMDB10207; HMDB12327; HMDB29196; | M-H | - | - |

| | | | | | | | | | | | | |
|---|---|---|---|---|---|---|---|---|---|---|---|---|
| | | | | | | | | | HMDB33821; HMDB59624; HMDB59625; HMDB60267 | | | |
| HILIC positive | 780.0018 | 38.7 | -0.1 (1.15) | 0.1 (0.81) | -0.2 (-0.35, -0.05) | 0.0096 | 0.5915 | CI Food Black 1 | HMDB33389 | M+H | - | - |
| C18 negative | 505.3536 | 165.5 | -0.1 (1.1) | 0.1 (0.88) | -0.2 (-0.36, -0.05) | 0.0097 | 0.5164 | Theasapogenol A | HMDB34519 | M-H | - | - |
| HILIC positive | 548.372 | 36.2 | -0.11 (1.23) | 0.11 (0.68) | -0.22 (-0.38, -0.05) | 0.0099 | 0.5915 | LysoPC(20:2(11Z14Z)) | HMDB10392 | M+H | - | - |
| HILIC positive | 451.3243 | 29.8 | -0.11 (1.04) | 0.11 (0.95) | -0.22 (-0.38, -0.05) | 0.0100 | 0.5915 | Deoxycholic acid glycine conjugate; Chenodeoxycholic acid glycine conjugate; Glycoursodeoxycholic acid; Chenodeoxyglycocholic acid | HMDB00631; HMDB00637; HMDB00708; HMDB06898 | M+H_[+1] | - | - |
| HILIC positive | 358.2011 | 292.2 | 0.11 (0.79) | -0.11 (1.16) | 0.23 (0.06, 0.4) | 0.0101 | 0.5915 | Nalbuphine; (S)-Laudanosine; 5-hydroxypropafenone | HMDB14982; HMDB30213; HMDB60988 | M+H | - | - |
| HILIC positive | 222.1124 | 179.3 | -0.11 (1.15) | 0.11 (0.81) | -0.22 (-0.39, -0.05) | 0.0102 | 0.5915 | Metaxalone; Carbofuran; Hydrocotarnine | HMDB14798; HMDB31770; HMDB33701 | M+H | - | - |
| C18 negative | 453.1563 | 111.8 | -0.08 (1.07) | 0.08 (0.92) | -0.16 (-0.29, -0.04) | 0.0103 | 0.5164 | KB 2 | HMDB33666 | M-H | - | - |
| C18 negative | 189.067 | 294.3 | 0.1 (0.83) | -0.1 (1.14) | 0.21 (0.05, 0.37) | 0.0103 | 0.5164 | 6-Hydroxy-1H-indole-3-acetamide | HMDB31173 | M-H | - | - |
| C18 negative | 255.1244 | 24.8 | -0.11 (1.15) | 0.11 (0.81) | -0.21 (-0.38, -0.05) | 0.0106 | 0.5178 | 1-Methoxy-1-(245-trimethoxyphenyl)-2-propanol; 2-4-(3-Hydroxypropyl)-2-methoxyphenoxy-13-propanediol | HMDB31772; HMDB40352 | M-H | - | - |
| HILIC positive | 153.0658 | 37.7 | 0.1 (1.1) | -0.1 (0.88) | 0.21 (0.05, 0.37) | 0.0107 | 0.5915 | N1-Methyl-2-pyridone-5-carboxamide; N1-Methyl-4-pyridone-3-carboxamide | HMDB04193; HMDB04194 | M+H | - | - |
| C18 negative | 353.2333 | 152 | 0.11 (0.96) | -0.11 (1.03) | 0.22 (0.05, 0.39) | 0.0111 | 0.5178 | Prostaglandin F2a; Prostaglandin E1; Prostaglandin F2b; Troxilin B3; Trioxilin A3; 8-isoprostaglandin PGF2b; 111215-THETA; 1314-Dihydro-15-keto PGF2a; 8-Isoprostaglandin E1; 111415-THETA; 8-Isoprostaglandin F2a; Prostaglandin D1; 8-iso-1314-dihydro-15-keto-PGF2a; 111215-TriHETRE; 11b-PGF2a; 11-Epi-PGF2a; 1314-Dihydro- lipoxin A4; Prostaglandin H1; Sterebin G; 15-keto-PGF1alpha; (5Z9E12S14Z)-81112-Trihydroxyicosa-5914-trienoate | HMDB01139; HMDB01442; HMDB01483; HMDB01965; HMDB01977; HMDB02115; HMDB04684; HMDB04685; HMDB04686; HMDB04694; HMDB05083; HMDB05102; HMDB06562; HMDB10170; HMDB10199; HMDB11137; HMDB12563; HMDB13041; HMDB35380; HMDB60045; HMDB60107 | M-H | - | - |
| C18 negative | 443.317 | 221.6 | -0.1 (1.15) | 0.1 (0.82) | -0.21 (-0.37, -0.05) | 0.0112 | 0.5178 | 1-a24R25-Trihydroxyvitamin D2; 59-Epidioxy-3-hydroxyergost-7-en-6-one; (3beta5alpha6alpha7beta14alpha22E24R)-56-Epoxyergosta-822-diene-3714-triol; 56:89-Diepoxyergost-22-ene-37beta-diol; (3beta5alpha6beta9alpha22E24R)-59-Epidioxyergosta-722-diene-36-diol | HMDB06227; HMDB32667; HMDB32691; HMDB33647; HMDB38977 | M-H | - | - |
| HILIC positive | 121.1011 | 28.6 | 0.1 (1) | -0.1 (0.99) | 0.21 (0.05, 0.37) | 0.0114 | 0.5991 | 124-Trimethylbenzene; Isopropylbenzene; 124-Tris(methylene)cyclohexane; Mesitylene; o-Ethyltoluene; m-Ethyltoluene; Propylbenzene; Hemimellitene | HMDB13733; HMDB34029; HMDB40458; HMDB41924; HMDB59819; HMDB59848; HMDB59877; HMDB59901 | M+H | - | - |
| C18 negative | 213.1131 | 292.7 | 0.1 (0.97) | -0.1 (1.02) | 0.2 (0.04, 0.35) | 0.0116 | 0.5247 | 5-Hexyltetrahydro-2-oxo-3-furancarboxylic acid; alpha-Carboxy-delta-decalactone; 2-Carboxy-57-dimethyl-4-octanolide | HMDB30984; HMDB30985; HMDB30986 | M-H | - | - |
| C18 negative | 437.3275 | 214.4 | -0.1 (1.02) | 0.1 (0.97) | -0.19 (-0.34, -0.04) | 0.0117 | 0.5247 | 27-Nor-5b-cholestane-3a7a12a2425-pentol | HMDB02126 | M-H | - | - |
| HILIC positive | 324.326 | 27.7 | -0.1 (1.05) | 0.1 (0.94) | -0.19 (-0.34, -0.04) | 0.0117 | 0.5992 | N-(14-Methylhexadecanoyl)pyrrolidine | HMDB34373 | M+H | - | - |
| C18 negative | 315.2537 | 208.5 | 0.11 (0.84) | -0.11 (1.13) | 0.21 (0.05, 0.38) | 0.0118 | 0.5250 | MG(0:015:00:0); MG(15:00:00:0); ()-1516-Dihydroxyoctadecanoic acid; (9S10S)-910-dihydroxyoctadecanoate | HMDB11532; HMDB11563; HMDB31008; HMDB59633 | M-H | - | - |
| HILIC positive | 311.221 | 229.6 | 0.11 (0.71) | -0.11 (1.21) | 0.22 (0.05, 0.4) | 0.0119 | 0.5992 | 12(13)Ep-9-KODE; (9Z11R12S13S15Z)-1213-Epoxy-11-hydroxy-915-octadecadienoic acid; Sterebin A; Auxin b | HMDB13623; HMDB33505; HMDB35337; HMDB38484 | M+H | - | - |
| HILIC positive | 86.0964 | 43.5 | -0.1 (1.03) | 0.1 (0.96) | -0.2 (-0.35, -0.04) | 0.0126 | 0.5992 | Piperidine | HMDB34301 | M+H | - | - |
| HILIC positive | 277.2162 | 27.5 | -0.1 (1.07) | 0.1 (0.91) | -0.19 (-0.35, -0.04) | 0.0126 | 0.5992 | 19-Norandrosterone; 19-Nor-5-androstenediol; 19-Noretiocholanolone; Stearidonic acid; Phenethyl decanoate; 481215-Octadecatetraenoic acid; (EE)-1113-Octadecadien-9-ynoic acid | HMDB02697; HMDB04590; HMDB05886; HMDB06547; HMDB32464; HMDB32672; HMDB34382 | M+H | - | - |
| C18 negative | 856.7194 | 46.9 | 0.1 (0.79) | -0.1 (1.17) | 0.2 (0.04, 0.37) | 0.0129 | 0.5331 | PC(24:0P-18:0); PC(P-18:024:0); PC(o-18:1(9Z)24:0); PC(o-22:020:1(11Z)) | HMDB08785; HMDB11263; HMDB13434; HMDB13447 | M-H | - | - |
| C18 negative | 347.1501 | 139.8 | -0.09 (1.01) | 0.09 (0.98) | -0.19 (-0.34, -0.04) | 0.0132 | 0.5387 | Gibberellin A60; Gibberellin A58; Gibberellin A63; Gibberellin A54; Gibberellin A67; Gibberellin A77; 8-Angeloylegelolide; Gibberellin A81; Triphasiol; Gibberellin A90; 3-Epigibberellin A1 | HMDB34988; HMDB35043; HMDB35044; HMDB35048; HMDB35049; HMDB37004; HMDB37773; HMDB38080; HMDB39062; HMDB39622; HMDB40944 | M-H | - | - |
| HILIC positive | 290.1596 | 113.8 | 0.11 (0.86) | -0.11 (1.12) | 0.21 (0.04, 0.39) | 0.0135 | 0.5992 | 3-Methylglutarylcarnitine | HMDB00552 | M+H | - | - |
| C18 negative | 405.1917 | 107.5 | -0.11 (1.22) | 0.11 (0.71) | -0.21 (-0.38, -0.04) | 0.0136 | 0.5387 | (1R3R4R5S6S8x)-1-Acetoxy-8-angeloyloxy-34-epoxy-5-hydroxy-7(14)10-bisaboladien-2-one; 1-(4-Hydroxy-35- | HMDB39971; HMDB41091 | M-H | - | - |

| | | | | | | | | | | | | |
|---|---|---|---|---|---|---|---|---|---|---|---|---|
| | | | | | | | | dimethoxyphenyl)-7-(4-hydroxy-3-methoxyphenyl)-35-heptanediol | | | | |
| C18 negative | 129.0193 | 21 | 0.1 (0.98) | -0.1 (1.01) | 0.2 (0.04, 0.35) | 0.0136 | 0.5387 | Citramalic acid; 3-Hydroxyglutaric acid; D-2-Hydroxyglutaric acid; L-2-Hydroxyglutaric acid; Ribonolactone; D-Xylono-15-lactone; 2-Hydroxyglutarate; Glutaconic acid; Citraconic acid; Mesaconic acid; Itaconic acid; Gamma-delta-Dioxovaleric acid; Methyl hydrogen fumarate; 2-Hydroxyglutaric acid lactone; 25-Dioxopentanoate; 2-Pentendioate | HMDB00426; HMDB00428; HMDB00606; HMDB00694; HMDB01900; HMDB11676; HMDB59655; HMDB00620; HMDB00634; HMDB00749; HMDB02092; HMDB13233; HMDB33809; HMDB59743; HMDB60365; HMDB61185 | M-H2O-H; M-H | 2-methylmaleate | M-H |
| HILIC positive | 221.1536 | 297.2 | 0.11 (1.01) | -0.11 (0.98) | 0.21 (0.04, 0.38) | 0.0138 | 0.5992 | 26-Di-tert-butylbenzoquinone; 3-Phenylpropyl isovalerate; 13-Nor-6-eremophilene-811-dione; Hexyl phenylacetate; 2-Phenylethyl hexanoate; (Z)-6-Tetradecene-13-diyne-58-diol; 13-Tetradecene-13-diyne-67-diol; 34-Dihydro-22578-pentamethyl-2H-1-benzopyran-6-ol; 2-Methyl-1-phenyl-2-propanyl butyrate | HMDB13817; HMDB36389; HMDB37606; HMDB37714; HMDB37718; HMDB38996; HMDB39539; HMDB40217; HMDB40226 | M+H | - | - |
| C18 negative | 490.3621 | 219.8 | -0.1 (1.02) | 0.1 (0.97) | -0.2 (-0.35, -0.04) | 0.0139 | 0.5387 | Barringtogenol C; Camelliagenin C; Ganoderiol H | HMDB34525; HMDB34532; HMDB37783 | M-H_[-1] | - | - |
| HILIC positive | 154.983 | 121 | -0.09 (1.07) | 0.09 (0.91) | -0.17 (-0.31, -0.03) | 0.0139 | 0.5992 | (E)-S-1-Propenyl thiosulfate | HMDB41079 | M+H | - | - |
| HILIC positive | 273.0048 | 84.9 | 0.09 (0.99) | -0.09 (1) | 0.18 (0.04, 0.32) | 0.0141 | 0.5992 | Zoledronate | HMDB14543 | M+H | - | - |
| HILIC positive | 411.2506 | 26.3 | 0.1 (0.76) | -0.1 (1.18) | 0.2 (0.04, 0.36) | 0.0143 | 0.5992 | LPA(0:016:0); LPA(16:00:0) | HMDB07849; HMDB07853 | M+H | - | - |
| HILIC positive | 288.1665 | 86.1 | -0.09 (1.07) | 0.09 (0.92) | -0.18 (-0.33, -0.04) | 0.0147 | 0.5992 | Arginyl-Hydroxyproline; Hydroxyprolyl-Arginine | HMDB28710; HMDB28857 | M+H | - | - |
| HILIC positive | 322.186 | 236 | 0.1 (0.92) | -0.1 (1.06) | 0.21 (0.04, 0.37) | 0.0148 | 0.5992 | Arginyl-Phenylalanine; Phenylalanyl-Arginine | HMDB28716; HMDB28989 | M+H | - | - |
| HILIC positive | 87.0998 | 46.3 | -0.1 (1.02) | 0.1 (0.97) | -0.19 (-0.35, -0.04) | 0.0148 | 0.5992 | Piperidine | HMDB34301 | M+H_[+1] | - | - |
| HILIC positive | 209.0129 | 175.8 | -0.09 (1.03) | 0.09 (0.96) | -0.17 (-0.31, -0.03) | 0.0148 | 0.5992 | (Z)-3-(Methylsulfinyl)-1-propenyl 2-propenyl disulfide | HMDB32653 | M+H | - | - |
| C18 negative | 453.3223 | 153.2 | -0.1 (1.05) | 0.1 (0.94) | -0.2 (-0.36, -0.04) | 0.0154 | 0.5427 | 27-Norcholestanehexol | HMDB02157 | M-H | - | - |
| HILIC positive | 263.0874 | 297.1 | 0.07 (0.98) | -0.07 (1.01) | 0.13 (0.03, 0.24) | 0.0156 | 0.5992 | L-beta-aspartyl-L-glutamic acid ; Na-L-Glutamyl-L-aspartic acid; Ng-L-Glutamyl-L-aspartic acid | HMDB11164; HMDB30413; HMDB30419 | M+H | - | - |
| HILIC positive | 135.0441 | 294.7 | -0.1 (1.34) | 0.1 (0.44) | -0.21 (-0.38, -0.04) | 0.0156 | 0.5992 | Phthalide | HMDB32469 | M+H | - | - |
| HILIC positive | 224.1281 | 184.7 | 0.11 (0.76) | -0.11 (1.19) | 0.22 (0.04, 0.4) | 0.0159 | 0.5992 | Cerulenin | HMDB15168 | M+H | - | - |
| HILIC positive | 287.2366 | 27.1 | -0.1 (1.13) | 0.1 (0.84) | -0.19 (-0.35, -0.04) | 0.0160 | 0.5992 | Vitamin A; 11-cis-Retinol; 9-cis-Retinol; 13-cis Retinol; (x)-p-Menth-1-en-4-yl 5-isopropyl-2-methylphenyl ether; Abietinal; ent-16-Kauren-19-al; ent-19-Trachylobanal; 813-Abietadien-18-al | HMDB00305; HMDB06216; HMDB06217; HMDB06221; HMDB30403; HMDB34735; HMDB36728; HMDB36841; HMDB38709 | M+H | - | - |
| HILIC positive | 760.5136 | 53.7 | -0.09 (1.12) | 0.09 (0.85) | -0.18 (-0.33, -0.03) | 0.0162 | 0.5992 | PS(16:018:2(9Z12Z)); PS(16:1(9Z)18:1(9Z)); PS(18:1(9Z)16:1(9Z)); PS(18:2(9Z12Z)16:0) | HMDB12358; HMDB12368; HMDB12388; HMDB12398 | M+H | - | - |
| HILIC positive | 519.4039 | 27.2 | -0.09 (1.08) | 0.09 (0.9) | -0.18 (-0.33, -0.03) | 0.0162 | 0.5992 | Ganoderiol C | HMDB37781 | M+H | - | - |
| C18 negative | 241.2172 | 235.1 | 0.1 (0.92) | -0.1 (1.06) | 0.2 (0.04, 0.36) | 0.0164 | 0.5488 | Pentadecanoic acid; Methyl tetradecanoate; Dodecyl propionate; Hexyl nonanoate; Heptyl octanoate; Octyl heptanoate; Pentyl decanoate; 3-Methylbutyl decanoate; 7(14)-Farnesene-912-diol; 2610-Trimethyldodecanoic acid; Isopropyllaurate; Ethyl tridecanoate | HMDB00826; HMDB30469; HMDB32250; HMDB32328; HMDB36218; HMDB36220; HMDB37306; HMDB38907; HMDB40998; HMDB41588; HMDB59820; HMDB59833 | M-H | - | - |
| HILIC positive | 88.0757 | 94.8 | -0.1 (1.31) | 0.1 (0.51) | -0.21 (-0.37, -0.04) | 0.0164 | 0.5992 | 4-Aminobutyraldehyde; N-Ethylacetamide; Morpholine; Butyramide; 4-Ammoniobutanal | HMDB01080; HMDB31215; HMDB31581; HMDB33870; HMDB60247 | M+H | - | - |
| HILIC positive | 263.0661 | 92.7 | 0.09 (0.98) | -0.09 (1.01) | 0.18 (0.03, 0.32) | 0.0169 | 0.5992 | Cinoxacin; 5-Fluorouridine | HMDB14965; HMDB60396 | M+H | - | - |
| C18 negative | 427.3065 | 252.4 | 0.09 (0.96) | -0.09 (1.03) | 0.18 (0.03, 0.33) | 0.0169 | 0.5492 | Sorbitan oleate; Glyceryl lactooleate | HMDB29886; HMDB32298 | M-H | - | - |
| HILIC positive | 137.1325 | 102.9 | 0.1 (0.74) | -0.1 (1.2) | 0.2 (0.04, 0.36) | 0.0170 | 0.5992 | D-Limonene; ()-Limonene; Gamma-terpinene; ()-a-Pinene; cis-Ocimene; trans-Ocimen; Polylimonene; 5-Isosylveterpinolene; ()-alpha-Carene; (-)-4(10)-Thujene; (S)-alpha-Phellandrene; (-)-alpha-Carene; alpha-Carene; (-)-77-Dimethyl-2-methylenebicyclo221heptane; (-)-alpha-Pinene; alpha-Phellandrene; (R)-alpha-Phellandrene; ()-4(10)-Thujene; beta-Phellandrene; (Z)-37-Dimethyl-137-octatriene; (-)-3-Thujene; (-)-beta-Pinene; beta-Pinene; Terpinolene; alpha-Terpinene; beta-Terpinene; Sylveterpinolene; 177-Trimethyltricyclo221026heptane; p-Mentha-24-diene; 7-Methyl-3-methylene-16-octadiene; Artemisiatriene; (-)-beta-Phellandrene; ()-beta-Phellandrene; ()-4-Carene; Camphene; 2-Carene | HMDB03375; HMDB04321; HMDB05806; HMDB06525; HMDB30088; HMDB30089; HMDB32473; HMDB33200; HMDB34697; HMDB34928; HMDB34970; HMDB35087; HMDB35619; HMDB35625; HMDB35658; HMDB35850; HMDB35851; HMDB36076; HMDB36081; HMDB36104; HMDB36116; HMDB36559; HMDB36560; HMDB36994; HMDB36995; HMDB36997; HMDB37046; HMDB38121; HMDB38150; HMDB38169; HMDB41574; HMDB41633; | M+H | - | - |

| | | | | | | | | | | | | |
|---|---|---|---|---|---|---|---|---|---|---|---|---|
| | | | | | | | | | HMDB41634; HMDB59726; HMDB59839; HMDB59884 | | | |
| HILIC positive | 299.258 | 27.2 | -0.09 (0.99) | 0.09 (1) | -0.19 (-0.34, -0.03) | 0.0170 | 0.5992 | 3-Oxooctadecanoic acid; MG(0:015:00:0); MG(15:00:00:0); 9-Oxooctadecanoic acid; 10-Oxooctadecanoic acid; 11-Oxooctadecanoic acid; ()-1516-Dihydroxyoctadecanoic acid; 5-Hexyltetrahydro-2-furanoctanoic acid; 5-Oxooctadecanoic acid; Ricinoleic acid; (9S10S)-910-dihydroxyoctadecanoate; Glycerol triundecanoate | HMDB10736; HMDB11532; HMDB11563; HMDB30979; HMDB30980; HMDB30981; HMDB31008; HMDB31127; HMDB34074; HMDB34297; HMDB59633; HMDB31089 | M+H; M+H-H2O; M+2H | - | - |
| C18 negative | 166.9938 | 31.2 | -0.09 (1.15) | 0.09 (0.82) | -0.18 (-0.34, -0.03) | 0.0170 | 0.5493 | Desflurane; Hexafluoroisopropanol | HMDB15320; HMDB60648 | M-H | - | - |
| HILIC positive | 403.3576 | 26.7 | -0.1 (1.03) | 0.1 (0.96) | -0.2 (-0.36, -0.04) | 0.0172 | 0.6003 | 24-Hydroxycholesterol; 7a-Hydroxycholesterol; 27-Hydroxycholesterol; Delta-Tocopherol; 22b-Hydroxycholesterol; 7b-Hydroxycholesterol; 25-Hydroxycholesterol; 20alpha-Hydroxycholesterol; 7a-Hydroxy-5b-cholestan-3-one; (24R)-Cholest-5-ene-3-beta24-diol ; 4b-Hydroxycholesterol; (3beta5alpha6alpha)-Cholest-8-ene-36-diol; eta-Tocopherol; 3-Hydroxy-1-phenyl-1-heneicosanone; 2-Methyl-6-phytylhydroquinone; 5-(12-Heneicosenyl)-13-benzenediol; 5-(14-Heneicosenyl)-13-benzenediol; 5-(16-Heneicosenyl)-13-benzenediol; Cholest-5-ene-3-beta7-alpha-diol | HMDB01419; HMDB01496; HMDB02103; HMDB02902; HMDB04035; HMDB06119; HMDB06247; HMDB06283; HMDB06892; HMDB11643; HMDB13643; HMDB29320; HMDB34254; HMDB35681; HMDB38959; HMDB39868; HMDB39869; HMDB39870; HMDB59592 | M+H | - | - |
| C18 negative | 611.3796 | 137 | -0.1 (1.05) | 0.1 (0.94) | -0.19 (-0.35, -0.03) | 0.0173 | 0.5532 | Cholestane-371225-tetrol-3-glucuronide | HMDB10355 | M-H | - | - |
| C18 negative | 205.0174 | 22.8 | -0.1 (1.21) | 0.1 (0.73) | -0.2 (-0.37, -0.04) | 0.0177 | 0.5554 | Daucic acid | HMDB31665 | M-H_[+2] | - | - |
| C18 negative | 375.1812 | 141.4 | -0.1 (1.07) | 0.1 (0.92) | -0.2 (-0.36, -0.03) | 0.0177 | 0.5554 | 18-Oxocortisol; 111214-Trihydroxy-7-methoxy-81113-abietatrien-206-olide; 12alpha-Hydroxy-1318-dehydroparain; 16alpha;-hydroxyprednisolone | HMDB00332; HMDB31932; HMDB39557; HMDB61016 | M-H | - | - |
| C18 negative | 408.2107 | 106.5 | -0.1 (1.19) | 0.1 (0.76) | -0.21 (-0.38, -0.04) | 0.0179 | 0.5563 | Cascarillin; Anhydrocinnzeylanine | HMDB36836; HMDB36864 | M-H_[-1] | - | - |
| C18 negative | 157.0619 | 293.3 | 0.1 (0.88) | -0.1 (1.1) | 0.19 (0.03, 0.35) | 0.0179 | 0.5563 | 1-(Hydroxymethyl)-55-dimethyl-24-imidazolidinedione | HMDB31670 | M-H | - | - |
| C18 negative | 114.9899 | 35.1 | -0.1 (1.05) | 0.1 (0.93) | -0.19 (-0.35, -0.03) | 0.0180 | 0.5592 | Trifluoroacetic acid | HMDB14118 | M-H_[+2] | - | - |
| C18 negative | 433.3326 | 235.9 | -0.09 (1.06) | 0.09 (0.92) | -0.19 (-0.34, -0.03) | 0.0182 | 0.5595 | 3a7a-Dihydroxycoprostanic acid; 3a7a12a-Trihydroxy-5b-cholestan-26-al | HMDB00359; HMDB03533 | M-H | - | - |
| C18 negative | 367.2253 | 275.1 | 0.09 (0.9) | -0.09 (1.09) | 0.17 (0.03, 0.32) | 0.0184 | 0.5599 | Perindopril | HMDB14928 | M-H | - | - |
| C18 negative | 605.2395 | 248.5 | -0.1 (1.02) | 0.1 (0.97) | -0.19 (-0.35, -0.03) | 0.0184 | 0.5599 | Phaeophorbide b | HMDB31149 | M-H | - | - |
| C18 negative | 433.3323 | 293.1 | -0.1 (1.21) | 0.1 (0.72) | -0.2 (-0.37, -0.03) | 0.0185 | 0.5599 | 3a7a-Dihydroxycoprostanic acid; 3a7a12a-Trihydroxy-5b-cholestan-26-al | HMDB00359; HMDB03533 | M-H | - | - |
| HILIC positive | 422.9844 | 67.2 | 0.1 (0.83) | -0.1 (1.14) | 0.21 (0.03, 0.38) | 0.0186 | 0.6117 | Chlorophenol red | HMDB36699 | M+H | - | - |
| HILIC positive | 129.0658 | 66.5 | -0.07 (1.04) | 0.07 (0.95) | -0.14 (-0.26, -0.02) | 0.0187 | 0.6117 | Dihydrothymine; Squamolone; L-Cyclo(alanylglycyl) | HMDB00079; HMDB29874; HMDB31547 | M+H | - | - |
| HILIC positive | 552.4025 | 36.1 | -0.09 (1.17) | 0.09 (0.78) | -0.19 (-0.35, -0.03) | 0.0188 | 0.6117 | LysoPC(20:0); Aliskiren | HMDB10390; HMDB15387 | M+H | - | - |
| C18 negative | 413.3425 | 298.1 | -0.09 (1.04) | 0.09 (0.95) | -0.18 (-0.33, -0.03) | 0.0190 | 0.5637 | (22Alpha)-hydroxy-campest-4-en-3-one; 4alpha-Formyl-5alpha-cholesta-8-en-3beta-ol; 4Alpha-hydroxymethyl-5alpha-cholesta-824-dien-3beta-ol; Ergostane-36-dione; Stenocereol; 1-Phenyl-13-docosanedione; (3beta5alpha7alpha)-14-Methylcholesta-9(11)24-diene-37-diol | HMDB12113; HMDB12169; HMDB12172; HMDB32004; HMDB34329; HMDB35586; HMDB41111 | M-H | - | - |
| HILIC positive | 131.0469 | 74.9 | -0.09 (1.01) | 0.09 (0.99) | -0.18 (-0.32, -0.03) | 0.0190 | 0.6139 | Pipecolic acid; Pyroglutamic acid; L-Pipecolic acid; Pyrrolidonecarboxylic acid; Pyrroline hydroxycarboxylic acid; N-Acryloylglycine; 1-Pyrroline-4-hydroxy-2-carboxylate; N4-Acetylaminobutanal; D-Pipecolic acid; Vigabatrin; Flucytosine; L-trans-4-Methyl-2-pyrrolidinecarboxylic acid; 2-Pyrrolidineacetic acid; 3-Acetamidobutanal; 5-Oxoprolinate; dimethadione | HMDB00070; HMDB00267; HMDB00716; HMDB00805; HMDB01369; HMDB01843; HMDB02234; HMDB04226; HMDB05960; HMDB15212; HMDB15231; HMDB29435; HMDB29444; HMDB59649; HMDB60262; HMDB61093 | M+H_[+1] | - | - |
| C18 negative | 203.0568 | 32.7 | 0.08 (0.93) | -0.08 (1.06) | 0.16 (0.03, 0.28) | 0.0193 | 0.5637 | 356-Trihydroxy-5-(hydroxymethyl)-2-methoxy-2-cyclohexen-1-one | HMDB41031 | M-H | - | - |
| C18 negative | 517.3894 | 238.5 | -0.09 (1.06) | 0.09 (0.93) | -0.19 (-0.34, -0.03) | 0.0193 | 0.5637 | Ganoderiol C | HMDB37781 | M-H | - | - |
| C18 negative | 766.5388 | 290.7 | -0.1 (1.15) | 0.1 (0.82) | -0.19 (-0.36, -0.03) | 0.0196 | 0.5637 | PC(15:020:4(5Z8Z11Z14Z)); PC(15:020:4(8Z11Z14Z17Z)); PC(20:4(5Z8Z11Z14Z)15:0); PC(20:4(8Z11Z14Z17Z)15:0); PE(16:022:4(7Z10Z13Z16Z)); PE(16:1(9Z)20:3(8Z11Z14Z)); PE(18:020:4(5Z8Z11Z14Z)); PE(18:020:4(8Z11Z14Z17Z)); PE(18:1(11Z)20:3(5Z8Z11Z)); PE(18:1(11Z)20:3(8Z11Z14Z)); PE(18:1(9Z)20:3(5Z8Z11Z)); PE(18:1(9Z)20:3(8Z11Z14Z)); PE(18:2(9Z12Z)20:2(11Z14Z)); PE(18:3(6Z9Z12Z)20:1(11Z)); PE(18:3(9Z12Z15Z)20:1(11Z)); PE(18:4(6Z9Z12Z15Z)20:0); PE(20:018:4(6Z9Z12Z15Z)); PE(20:1(11Z)18:3(6Z9Z12Z)); PE(20:1(11Z)18:3(9Z12Z15Z)); PE(20:2(11Z14Z)18:2(9Z12Z)); PE(20:3(5Z8Z11Z)18:1(11Z)); | HMDB07949; HMDB07950; HMDB08428; HMDB08461; HMDB08943; HMDB08969; HMDB09003; HMDB09004; HMDB09034; HMDB09035; HMDB09067; HMDB09068; HMDB09099; HMDB09131; HMDB09164; HMDB09196; HMDB09228; HMDB09259; HMDB09260; HMDB09291; HMDB09322; HMDB09323; HMDB09355; HMDB09356; | M-H | - | - |

| | | | | | | | | | | | |
|---|---|---|---|---|---|---|---|---|---|---|---|
| | | | | | | | | PE(20:3(5Z8Z11Z)18:1(9Z)); PE(20:3(8Z11Z14Z)18:1(11Z)); PE(20:3(8Z11Z14Z)18:1(9Z)); PE(20:4(5Z8Z11Z14Z)18:0); PE(20:4(8Z11Z14Z17Z)18:0); PE(22:4(7Z10Z13Z16Z)16:0) | HMDB09387; HMDB09420; HMDB09583 | | | |
| HILIC positive | 331.2637 | 27.2 | 0.09 (0.97) | -0.09 (1.03) | 0.18 (0.03, 0.33) | 0.0199 | 0.6191 | Docosapentaenoic acid (22n-6); Docosapentaenoic acid; Ethyl abietate; ent-16-Kauren-19-ol acetate; 1-Phenyl-13-hexadecanedione; 2-Methyl-5-(811-pentadecadienyl)-13-benzenediol; 48121519-Docosapentaenoic acid; Ethyl icosapentate; Docosa-47101316-pentaenoic acid | HMDB01976; HMDB06528; HMDB32257; HMDB35024; HMDB35581; HMDB38909; HMDB39133; HMDB39530; HMDB60113 | M+H | - | - |
| HILIC positive | 314.0748 | 96.1 | -0.08 (1.04) | 0.08 (0.96) | -0.17 (-0.31, -0.03) | 0.0200 | 0.6191 | Phosphoribosylformylglycineamidine; 5-phosphoribosyl-a-N-formylglycineamidine | HMDB00999; HMDB02305 | M+H | - | - |
| C18 negative | 230.0125 | 23.6 | -0.09 (1.02) | 0.09 (0.97) | -0.18 (-0.33, -0.03) | 0.0201 | 0.5637 | Paracetamol sulfate; Benzeneacetamide-4-O-sulphate | HMDB59911; HMDB59994 | M-H | - | - |
| HILIC positive | 508.3762 | 36.8 | -0.1 (1.18) | 0.1 (0.76) | -0.2 (-0.37, -0.03) | 0.0202 | 0.6191 | LysoPC(P-18:0) | HMDB13122 | M+H | - | - |
| HILIC positive | 114.0662 | 36.1 | -0.06 (1.06) | 0.06 (0.94) | -0.12 (-0.21, -0.02) | 0.0206 | 0.6191 | Creatinine | HMDB00562 | M+H | Creatinine | M+H |
| C18 negative | 305.0344 | 45.4 | -0.1 (1.2) | 0.1 (0.74) | -0.2 (-0.37, -0.03) | 0.0207 | 0.5659 | 4-Hydroxy-5-(dihydroxyphenyl)-valeric acid-O-sulphate; 4-Hydroxy-5-(dihydroxyphenyl)-valeric acid-O-sulphate III | HMDB59978; HMDB59979 | M-H | - | - |
| C18 negative | 242.1843 | 139.1 | 0.09 (0.89) | -0.09 (1.09) | 0.19 (0.03, 0.34) | 0.0208 | 0.5659 | 3-Oxotetradecanoic acid; 6-Ketomyristic acid; L-Menthyl (RS)-3-hydroxybutyrate | HMDB10730; HMDB30982; HMDB32370 | M-H_[-1] | - | - |
| HILIC positive | 173.0298 | 64.1 | -0.07 (1.07) | 0.07 (0.92) | -0.15 (-0.27, -0.02) | 0.0209 | 0.6191 | Dihydrothymine; Squamolone; L-Cyclo(alanylglycyl) | HMDB00079; HMDB29874; HMDB31547 | M+2Na-H | - | - |
| C18 negative | 257.1032 | 293.8 | 0.1 (0.89) | -0.1 (1.1) | 0.19 (0.03, 0.36) | 0.0209 | 0.5659 | 11-(Tetrahydro-6a-hydroxy-23a5-trimethylfuro23-d-13-dioxole-25-diyl)bis-ethanone | HMDB32527 | M-H | - | - |
| HILIC positive | 267.1706 | 29.9 | -0.08 (1) | 0.08 (0.99) | -0.16 (-0.3, -0.02) | 0.0210 | 0.6191 | Atenolol; Practolol | HMDB01924; HMDB15411 | M+H | - | - |
| HILIC positive | 181.1588 | 110.9 | 0.1 (0.75) | -0.1 (1.19) | 0.2 (0.03, 0.37) | 0.0212 | 0.6191 | Homodihydrojasmone; 2-trans-6-cis-Dodecadienal; cis-Quinceoxepane; Tricycloekasantalol; (2E4E)-24-Dodecadienal | HMDB31181; HMDB32531; HMDB38108; HMDB39339; HMDB39590 | M+H | - | - |
| HILIC positive | 257.2474 | 294.9 | 0.1 (0.78) | -0.1 (1.17) | 0.19 (0.03, 0.36) | 0.0215 | 0.6191 | Palmitic acid; Trimethyltridecanoic acid; Isopalmitic acid; Butyl dodecanoate; Dodecyl butyrate; Hexyl decanoate; Octyl octanoate; Ethyl tetradecanoate; Dodecyl 2-methylpropanoate; Hexadecanoate (n-C16:0) | HMDB00220; HMDB02396; HMDB31068; HMDB32065; HMDB32249; HMDB32324; HMDB33166; HMDB34153; HMDB36210; HMDB60083 | M+H | - | - |
| HILIC positive | 714.5445 | 208.3 | 0.09 (0.91) | -0.09 (1.07) | 0.18 (0.03, 0.34) | 0.0219 | 0.6191 | PC(14:1(9Z)P-18:1(11Z)); PC(14:1(9Z)P-18:1(9Z)); PC(P-18:1(11Z)14:1(9Z)); PC(P-18:1(9Z)14:1(9Z)) | HMDB07930; HMDB07931; HMDB11270; HMDB11303 | M+H | - | - |
| HILIC positive | 193.0858 | 285.3 | 0.1 (0.82) | -0.1 (1.14) | 0.21 (0.03, 0.39) | 0.0223 | 0.6191 | (R)-Shinanolone; Methyl trans-p-methoxycinnamate; Allyl phenoxyacetate; Ethyl phenylglycidate; Dehydrozingerone; Isomyristicin; Myristicin; (Z)-4-Methoxy-6-(1-propenyl)-13-benzodioxole; Isoeugenol formate; Eugenyl formate; Ethyl 3-oxo-3-phenylpropanoate; Benzyl acetoacetate; 1-Hydroxychavicol acetate; 4-(34-Methylenedioxyphenyl)-2-butanone | HMDB30580; HMDB30752; HMDB31610; HMDB31623; HMDB32591; HMDB35275; HMDB35873; HMDB35894; HMDB37277; HMDB37877; HMDB40268; HMDB40446; HMDB41007; HMDB41481 | M+H | - | - |
| HILIC positive | 473.3618 | 26.9 | -0.09 (1.03) | 0.09 (0.96) | -0.19 (-0.35, -0.03) | 0.0223 | 0.6191 | Maslinic acid; Lucidumol A; Queretaroic acid; Priverogenin A; (3beta23xi)-323-Dihydroxycycloart-24-en-26-oic acid; Pomolic acid; 20beta-Hydroxyursolic acid; Azukisapogenol; delta-Maslinic acid; Sebiferenic acid; Momordicin I; Ganoderic acid U; Ganodermanontriol; Epoxyganoderiol A; (3b22S24E)-322-Dihydroxycycloart-24-en-26-oic acid; 27-Hydroxyisomangiferolic acid; Albigenic acid; Rubitic acid; Ganoderiol E; 3alpha-Corosolic acid | HMDB02392; HMDB33233; HMDB34523; HMDB34530; HMDB34567; HMDB35106; HMDB35107; HMDB35258; HMDB35260; HMDB35748; HMDB35758; HMDB35962; HMDB35977; HMDB36001; HMDB36063; HMDB36064; HMDB36638; HMDB36654; HMDB37779; HMDB41043 | M+H | - | - |
| C18 negative | 335.0345 | 20.4 | -0.04 (1.05) | 0.04 (0.95) | -0.08 (-0.15, -0.01) | 0.0226 | 0.5770 | Malaoxon | HMDB60627 | M+Na-2H | - | - |
| C18 negative | 321.0411 | 19 | -0.05 (1.03) | 0.05 (0.97) | -0.09 (-0.18, -0.01) | 0.0229 | 0.5770 | 77-Dihydroxy-68-bicoumarin | HMDB33559 | M-H | - | - |
| HILIC positive | 130.0499 | 76.6 | -0.09 (1) | 0.09 (0.99) | -0.17 (-0.32, -0.02) | 0.0229 | 0.6191 | Pyroglutamic acid; Pyrrolidonecarboxylic acid; Pyrroline hydroxycarboxylic acid; N-Acryloylglycine; 1-Pyrroline-4-hydroxy-2-carboxylate; 5-Oxoprolinate; dimethadione | HMDB00267; HMDB00805; HMDB01369; HMDB01843; HMDB02234; HMDB60262; HMDB61093 | M+H | - | - |
| HILIC positive | 131.0533 | 75.4 | -0.09 (1) | 0.09 (0.99) | -0.17 (-0.32, -0.02) | 0.0230 | 0.6191 | Pipecolic acid; Pyroglutamic acid; L-Pipecolic acid; Pyrrolidonecarboxylic acid; Pyrroline hydroxycarboxylic acid; N-Acryloylglycine; 1-Pyrroline-4-hydroxy-2-carboxylate; N4-Acetylaminobutanal; D-Pipecolic acid; Vigabatrin; L-trans-4-Methyl-2-pyrrolidinecarboxylic acid; 2-Pyrrolidineacetic acid; 3-Acetamidobutanal; 5-Oxoprolinate; dimethadione | HMDB00070; HMDB00267; HMDB00716; HMDB00805; HMDB01369; HMDB01843; HMDB02234; HMDB04226; HMDB05960; HMDB15212; HMDB29435; HMDB29444; HMDB59649; HMDB60262; HMDB61093 | M+H_[+1] | - | - |
| C18 negative | 471.3485 | 237 | -0.09 (1.06) | 0.09 (0.93) | -0.19 (-0.35, -0.03) | 0.0230 | 0.5770 | Maslinic acid; Lucidumol A; Queretaroic acid; Priverogenin A; (3beta23xi)-323-Dihydroxycycloart-24-en-26-oic acid; Pomolic acid; 20beta-Hydroxyursolic acid; Azukisapogenol; | HMDB02392; HMDB33233; HMDB34523; HMDB34530; HMDB34567; HMDB35106; | M-H | - | - |

| | | | | | | | | | | | | |
|---|---|---|---|---|---|---|---|---|---|---|---|---|
| | | | | | | | | delta-Maslinic acid; Sebiferenic acid; Momordicin I; Ganoderic acid U; Ganodermanontriol; Epoxyganoderiol A; (3b22S24E)-322-Dihydroxycycloart-24-en-26-oic acid; 27-Hydroxyisomangiferolic acid; Albigenic acid; Rubitic acid; Ganoderiol E; 3alpha-Corosolic acid | HMDB35107; HMDB35258; HMDB35260; HMDB35748; HMDB35758; HMDB35962; HMDB35977; HMDB36001; HMDB36063; HMDB36064; HMDB36638; HMDB36654; HMDB37779; HMDB41043 | | | |
| HILIC positive | 742.4534 | 54.2 | -0.06 (1) | 0.06 (1) | -0.12 (-0.23, -0.02) | 0.0230 | 0.6191 | Lycoperoside D | HMDB34484 | M+H_[+2] | - | - |
| HILIC positive | 157.0681 | 83.9 | -0.07 (1.1) | 0.07 (0.89) | -0.14 (-0.26, -0.02) | 0.0231 | 0.6191 | 2-(Isopropylthio)methylfuran | HMDB36181 | M+H | - | - |
| HILIC positive | 477.3923 | 26.7 | -0.09 (1.03) | 0.09 (0.96) | -0.18 (-0.33, -0.02) | 0.0232 | 0.6191 | Panaxatriol | HMDB38244 | M+H | - | - |
| C18 negative | 833.5188 | 219.7 | -0.09 (1.06) | 0.09 (0.93) | -0.18 (-0.33, -0.02) | 0.0235 | 0.5783 | PI(16:018:2(9Z12Z)); PI(16:1(9Z)18:1(11Z)); PI(16:1(9Z)18:1(9Z)); PI(16:2(9Z12Z)18:0); PI(18:016:2(9Z12Z)); PI(18:1(11Z)16:1(9Z)); PI(18:1(9Z)16:1(9Z)); PI(18:2(9Z12Z)16:0) | HMDB09784; HMDB09800; HMDB09801; HMDB09803; HMDB09807; HMDB09823; HMDB09835; HMDB09846 | M-H | - | - |
| C18 negative | 475.3427 | 228.1 | -0.09 (1.09) | 0.09 (0.9) | -0.18 (-0.33, -0.02) | 0.0237 | 0.5789 | Homodolichosterone; (2alpha3alpha5alpha22R23R)-232223-Tetrahydroxy-25-methylergost-24(28)en-6-one | HMDB34079; HMDB39443 | M-H | - | - |
| HILIC positive | 958.3128 | 54.4 | -0.08 (1.15) | 0.08 (0.81) | -0.16 (-0.3, -0.02) | 0.0238 | 0.6191 | Pentaglutamyl folate | HMDB06487 | M+H | - | - |
| HILIC positive | 435.3458 | 26.7 | -0.09 (1.1) | 0.09 (0.88) | -0.19 (-0.35, -0.03) | 0.0240 | 0.6191 | 3a7a-Dihydroxycoprostanic acid; 3a7a12a-Trihydroxy-5b-cholestan-26-al | HMDB00359; HMDB03533 | M+H | - | - |
| C18 negative | 264.1087 | 294.5 | -0.1 (1.11) | 0.1 (0.87) | -0.2 (-0.37, -0.03) | 0.0246 | 0.5797 | D-1-(3-Carboxypropyl)amino-1-deoxyfructose | HMDB38663 | M-H | - | - |
| HILIC positive | 191.0662 | 86.2 | -0.08 (1.07) | 0.08 (0.92) | -0.17 (-0.31, -0.02) | 0.0247 | 0.6191 | L-beta-aspartyl-L-glycine ; Carglumic acid; Aspartyl-Glycine; Glycyl-Aspartate | HMDB11165; HMDB15673; HMDB28753; HMDB28837 | M+H | - | - |
| C18 negative | 315.05 | 19.3 | -0.04 (1.05) | 0.04 (0.95) | -0.07 (-0.14, -0.01) | 0.0252 | 0.5797 | Isorhamnetin; 3457-Tetrahydroxy-6-methoxyflavone; 2-(34-Dihydroxyphenyl)-56-dihydroxy-7-methoxy-4H-1-benzopyran-4-one; Pollenitin; 1358-Tetrahydroxy-6-methoxy-2-methylanthraquinone; Malaoxon | HMDB02655; HMDB30547; HMDB30804; HMDB38572; HMDB39210; HMDB60627 | M-H; M-H_[+2] | - | - |
| HILIC positive | 295.2267 | 27.9 | -0.09 (1.1) | 0.09 (0.88) | -0.17 (-0.32, -0.02) | 0.0252 | 0.6191 | 13-OxoODE; 9-OxoODE; A-12(13)-EpODE; 13-HOTE; 15(16)-EpODE; 9(10)-EpODE; 9-HOTE; 17-Hydroxylinolenic acid; 10-Oxo-11-octadecen-13-olide; Squamostanal A; 1516-Epoxy-912-octadecadienoic acid; (2E4Z8E)-Colneleic acid; 1213-Epoxy-915-octadecadienoic acid; 2-Hydroxylinolenic acid; (9S10E12Z15Z)-9-Hydroxy-101215-octadecatrienoic acid; (9Z12Z14E)-16-Hydroxy-91214-octadecatrienoic acid; Sterebin D; 34-Dimethyl-5-pentyl-2-furanheptanoic acid | HMDB04668; HMDB04669; HMDB10200; HMDB10203; HMDB10206; HMDB10220; HMDB10224; HMDB11108; HMDB29786; HMDB29969; HMDB30950; HMDB30995; HMDB31088; HMDB31103; HMDB31934; HMDB34586; HMDB36832; HMDB39603 | M+H | - | - |
| HILIC positive | 133.1011 | 26.7 | -0.1 (1.1) | 0.1 (0.88) | -0.19 (-0.37, -0.02) | 0.0254 | 0.6191 | p-Mentha-1358-tetraene | HMDB29641 | M+H | - | - |
| C18 negative | 165.0921 | 30.1 | -0.08 (1.07) | 0.08 (0.92) | -0.16 (-0.3, -0.02) | 0.0257 | 0.5797 | Perillic acid; Wine lactone; 6-Pentyl-2H-pyran-2-one; 2-tert-Butyl-14-benzenediol; 2-Methoxy-4-propylphenol; Pentyl 2-furyl ketone; 11-Dimethoxy-2-phenylethane; alpha-Teresantalic acid; Epinepetalactone; 247-Decatrienoic acid; Perilla ketone; Marmelolactone A; Isomintlactone; 14-Epidioxy-p-mentha-28-diene; 12:34-Diepoxy-p-menth-8-ene; 5-Hydroxy-p-mentha-68-dien-2-one; 2-Hydroxy-p-mentha-18-dien-6-one; Rotundifolone; (1xi4xi6xi)-Carvone oxide; 4-Ethyl-12-dimethoxybenzene; Karahana lactone; 3-Hydroxy-p-mentha-18-dien-7-al | HMDB04586; HMDB29691; HMDB31085; HMDB32062; HMDB32135; HMDB32462; HMDB32615; HMDB34922; HMDB35063; HMDB35235; HMDB35328; HMDB36042; HMDB36084; HMDB36878; HMDB36879; HMDB37011; HMDB37012; HMDB37044; HMDB39382; HMDB40176; HMDB41000; HMDB41584 | M-H | - | - |
| HILIC positive | 716.522 | 28.3 | -0.09 (1.08) | 0.09 (0.9) | -0.18 (-0.33, -0.02) | 0.0259 | 0.6191 | PE(14:020:2(11Z14Z)); PE(14:1(9Z)20:1(11Z)); PE(16:018:2(9Z12Z)); PE(16:1(9Z)18:1(11Z)); PE(16:1(9Z)18:1(9Z)); PE(18:1(11Z)16:1(9Z)); PE(18:1(9Z)16:1(9Z)); PE(18:2(9Z12Z)16:0); PE(20:1(11Z)14:1(9Z)); PE(20:2(11Z14Z)14:0) | HMDB08835; HMDB08867; HMDB08928; HMDB08959; HMDB08960; HMDB09023; HMDB09056; HMDB09088; HMDB09251; HMDB09283 | M+H | - | - |
| HILIC positive | 313.2372 | 28.1 | -0.09 (1.03) | 0.09 (0.96) | -0.19 (-0.35, -0.02) | 0.0263 | 0.6191 | 13-L-Hydroperoxylinoleic acid; 8(R)-Hydroperoxylinoleic acid; 9(S)-HPODE; 1213-DiHODE; 1516-DiHODE; 910-DiHODE; ()-(E)-13-Hydroxy-10-oxo-11-octadecenoic acid | HMDB03871; HMDB04706; HMDB06940; HMDB10201; HMDB10208; HMDB10221; HMDB40900 | M+H | - | - |
| HILIC positive | 148.0736 | 75 | -0.08 (1.01) | 0.08 (0.99) | -0.17 (-0.32, -0.02) | 0.0264 | 0.6191 | L-Lysine; 2-Methylglutaric acid; Adipic acid; L-Glutamine; Methylglutaric acid; Monomethyl glutaric acid; Ureidoisobutyric acid; 22-Dimethylsuccinic acid; Solerol; D-Lysine; D-Glutamine; Alanylglycine; (S)-2-Aceto-2-hydroxybutanoic acid; (3S)-36-Diaminohexanoate; (3S5S)-35-Diaminohexanoate; Alanyl-Glycine; Dimethyl succinate | HMDB00182; HMDB00422; HMDB00448; HMDB00641; HMDB00752; HMDB00858; HMDB02031; HMDB02074; HMDB02173; HMDB03405; HMDB03423; HMDB06899; HMDB06900; HMDB12114; HMDB12115; HMDB28687; HMDB33837 | M+H_[+1] | - | - |

| | | | | | | | | | | | | |
|---|---|---|---|---|---|---|---|---|---|---|---|---|
| C18 negative | 291.0072 | 35.7 | -0.08 (1.05) | 0.08 (0.95) | -0.15 (-0.29, -0.02) | 0.0264 | 0.5797 | Carboxyphosphamide; Carboxycyclophosphamide; Carboxyifosfamide | HMDB60449; HMDB60687; HMDB60692 | M-H | - | - |
| HILIC positive | 413.3624 | 223.7 | -0.09 (1.13) | 0.09 (0.84) | -0.19 (-0.35, -0.02) | 0.0267 | 0.6191 | MG(0:022:1(13Z)0:0); MG(22:1(13Z)0:00:0) | HMDB11552; HMDB11582 | M+H | - | - |
| C18 negative | 567.3176 | 128.6 | -0.09 (1.04) | 0.09 (0.95) | -0.17 (-0.32, -0.02) | 0.0269 | 0.5797 | (3a5b7a)-23-Carboxy-7-hydroxy-24-norcholan-3-yl-b-D-Glucopyranosiduronic acid; Deoxycholic acid 3-glucuronide | HMDB02430; HMDB02596 | M-H | - | - |
| C18 negative | 199.1703 | 189.8 | 0.09 (0.94) | -0.09 (1.05) | 0.17 (0.02, 0.33) | 0.0270 | 0.5797 | Dodecanoic acid; Ethyl decanoate; 2-Heptyl butyrate; Nonanal propyleneglycol acetal; Hexyl hexanoate; Isopropyl nonanoate; Octyl butanoate; Octyl 2-methylpropanoate; (R)-Dihydrocitronellol acetate; Pentyl heptanoate; Decyl acetate; Isobutyl octanoate | HMDB00638; HMDB30998; HMDB32310; HMDB32440; HMDB33619; HMDB34128; HMDB34136; HMDB34137; HMDB35410; HMDB36225; HMDB40166; HMDB59868 | M-H | Lauric acid | M-H |
| HILIC positive | 148.0427 | 46.7 | -0.08 (1) | 0.08 (1) | -0.17 (-0.32, -0.02) | 0.0271 | 0.6191 | Thiomorpholine 3-carboxylate | HMDB59611 | M+H | - | - |
| C18 negative | 111.02 | 220.5 | 0.09 (0.73) | -0.09 (1.21) | 0.18 (0.02, 0.35) | 0.0272 | 0.5797 | Uracil; 4-Carboxypyrazole | HMDB00300; HMDB60760 | M-H | - | - |
| C18 negative | 429.3223 | 283.6 | 0.1 (1.03) | -0.1 (0.96) | 0.19 (0.02, 0.36) | 0.0274 | 0.5797 | Sorbitan stearate | HMDB29888 | M-H | - | - |
| HILIC positive | 101.0961 | 108.1 | 0.09 (0.81) | -0.09 (1.15) | 0.19 (0.02, 0.36) | 0.0275 | 0.6206 | 3-Hexanone; 4-Methylpentanal; Methyl isobutyl ketone; 2-Oxohexane; Ethyl isopropyl ketone; Hexanal; (E)-3-Hexen-1-ol; 2-Hexen-1-ol; 2-Ethylbutanal; (E)-4-Hexen-1-ol; 1-Hexen-3-ol; 2-Methylpentanal | HMDB00753; HMDB01318; HMDB02939; HMDB05842; HMDB05846; HMDB05994; HMDB30003; HMDB30952; HMDB31220; HMDB31502; HMDB31503; HMDB31578 | M+H | - | - |
| C18 negative | 351.1812 | 77.8 | 0.1 (0.84) | -0.1 (1.13) | 0.19 (0.02, 0.36) | 0.0275 | 0.5797 | 4-Gingerdiol 35-diacetate | HMDB39132 | M-H | - | - |
| C18 negative | 151.0401 | 23.8 | 0.1 (0.89) | -0.1 (1.09) | 0.19 (0.02, 0.36) | 0.0280 | 0.5797 | p-Hydroxyphenylacetic acid; 3-Hydroxyphenylacetic acid; Ortho-Hydroxyphenylacetic acid; Mandelic acid; p-Anisic acid; 3-Cresotinic acid; 34-Dihydroxyphenylacetaldehyde; 4-Hydroxy-3-methylbenzoic acid; Vanillin; 24-Dihydroxyacetophenone; 23-Dihydroxyacetophenone; 26-Dihydroxyacetophenone; 34-Dihydroxyacetophenone; 35-Dihydroxyacetophenone; Phenoxyacetic acid; Methyl furfuracrylate; Methylparaben; 2-(Hydroxymethyl)benzoic acid; 2-Methoxybenzoic acid; 3-Methoxybenzoic acid; 25-Dihydroxyacetophenone; Ethyl 2-furanyl diketone; 1-(5-Methyl-2-furanyl)-12-propanedione; Methyl 2-hydroxybenzoate; 2-Propenyl 2-furancarboxylate; 1-(2-Furanyl)-13-butanedione; 4-Hydroxyphenyl acetate | HMDB00020; HMDB00440; HMDB00669; HMDB00703; HMDB01101; HMDB02390; HMDB03791; HMDB04815; HMDB12308; HMDB29657; HMDB29658; HMDB29659; HMDB29660; HMDB29661; HMDB29662; HMDB31609; HMDB32399; HMDB32572; HMDB32594; HMDB32604; HMDB32606; HMDB32629; HMDB32919; HMDB33003; HMDB34172; HMDB36907; HMDB41283; HMDB60390 | M-H | Hydroxyphenylacetate | M-H |
| C18 negative | 487.3431 | 211.4 | -0.1 (1.1) | 0.1 (0.89) | -0.19 (-0.36, -0.02) | 0.0281 | 0.5797 | Pitheduloside I; Arjunolic acid; Camelliagenin B; Madasiatic acid; Esculentic acid (Diplazium); Hovenolactone; Centellasapogenol A; Glyyunnansapogenin B; Euscaphic acid; (3beta6alpha19alpha)-3619-Trihydroxy-12-ursen-28-oic acid; Ganoderiol D; Ananasic acid; 16beta-Hydroxystellatogenin; 21beta-Hydroxyhederagenin | HMDB34036; HMDB34502; HMDB34531; HMDB35118; HMDB35782; HMDB35957; HMDB36311; HMDB36639; HMDB36651; HMDB36961; HMDB37782; HMDB38116; HMDB40391; HMDB41039 | M-H | - | - |
| C18 negative | 197.1546 | 156 | 0.09 (0.87) | -0.09 (1.11) | 0.17 (0.02, 0.33) | 0.0282 | 0.5797 | 5-Dodecenoic acid; trans-Dodec-2-enoic acid; Methyl 10-undecenoate; Allyl nonanoate; xi-Dihydro-5-octyl-2(3H)-furanone; cis-4-Decenyl acetate; 26-Dimethyl-5-heptenal propyleneglycol acetal; 11-Dodecenoic acid; cis-3-Hexenyl hexanoate; ()-Citronellyl acetate; Citronellyl acetate; 55-Dibutyl-45-dihydro-2(3H)furanone; Rhodinyl acetate; Methyl 9-undecenoate; Ethyl 2-decenoate; 1-Ethenylhexyl butanoate; xi-5-Dodecanolide; 3-Heptyldihydro-5-methyl-2(3H)-furanone; 2-Octenyl butyrate; Hexyl 2E-hexenoate; 7-Hexyl-2-oxepanone; 2-Hexenyl hexanoate; Ethyl 4-decenoate; Hexyl 2-methyl-3-pentenoate; Hexyl 2-methyl-4-pentenoate; 11-Dimethoxy-37-dimethyl-26-octadiene; ()-Menthyl acetate; (37-Dimethyl-6-octenyl)oxyacetaldehyde | HMDB00529; HMDB10729; HMDB29585; HMDB29763; HMDB31683; HMDB32214; HMDB32235; HMDB32248; HMDB33378; HMDB34160; HMDB34844; HMDB36182; HMDB37186; HMDB37305; HMDB37329; HMDB37498; HMDB37742; HMDB37813; HMDB38081; HMDB38269; HMDB38895; HMDB38924; HMDB39220; HMDB40158; HMDB40163; HMDB40361; HMDB41264; HMDB41449 | M-H | - | - |
| HILIC positive | 203.1431 | 284.9 | 0.09 (0.72) | -0.09 (1.21) | 0.18 (0.02, 0.34) | 0.0282 | 0.6230 | ()-Anisoxide; Feniculin; 2-Pentyl-3-phenyl-2-propenal; Dictagymnin; Panaquinquecol 5 | HMDB30572; HMDB30838; HMDB31313; HMDB39274; HMDB39356 | M+H | - | - |
| C18 negative | 394.2961 | 278.1 | 0.09 (0.97) | -0.09 (1.02) | 0.17 (0.02, 0.33) | 0.0287 | 0.5797 | 912-Hexadecadienoylcarnitine | HMDB13334 | M-H | - | - |
| C18 negative | 834.5221 | 219.7 | -0.09 (1.05) | 0.09 (0.94) | -0.17 (-0.32, -0.02) | 0.0288 | 0.5797 | PI(16:018:2(9Z12Z)); PI(16:1(9Z)18:1(11Z)); PI(16:1(9Z)18:1(9Z)); PI(16:2(9Z12Z)18:0); PI(18:016:2(9Z12Z)); PI(18:1(11Z)16:1(9Z)); PI(18:1(9Z)16:1(9Z)); PI(18:2(9Z12Z)16:0) | HMDB09784; HMDB09800; HMDB09801; HMDB09803; HMDB09807; HMDB09823; HMDB09835; HMDB09846 | M-H_[-1] | - | - |

| | | | | | | | | | | | | |
|---|---|---|---|---|---|---|---|---|---|---|---|---|
| HILIC positive | 361.2008 | 30.2 | -0.1 (1.07) | 0.1 (0.92) | -0.19 (-0.37, -0.02) | 0.0288 | 0.6254 | Aldosterone; Cortisone; 19-Oic-deoxycorticosterone; Prednisolone; Tricyclodehydroisohumulone; 7-Methylrosmanol | HMDB00037; HMDB02802; HMDB12613; HMDB14998; HMDB30165; HMDB35813 | M+H | - | - |
| HILIC positive | 310.201 | 32.6 | -0.08 (1.2) | 0.08 (0.74) | -0.16 (-0.31, -0.02) | 0.0292 | 0.6262 | Nadolol; Metipranolol | HMDB15334; HMDB15345 | M+H | - | - |
| HILIC positive | 147.0764 | 77.1 | -0.08 (1) | 0.08 (0.99) | -0.17 (-0.32, -0.02) | 0.0294 | 0.6262 | Pyroglutamic acid; L-Glutamine; Pyrrolidonecarboxylic acid; Pyrroline hydroxycarboxylic acid; N-Acryloylglycine; Ureidoisobutyric acid; 1-Pyrroline-4-hydroxy-2-carboxylate; D-Glutamine; Alanylglycine; Alanyl-Glycine; 5-Oxoprolinate; dimethadione | HMDB00267; HMDB00641; HMDB00805; HMDB01369; HMDB01843; HMDB02031; HMDB02234; HMDB03423; HMDB06899; HMDB28687; HMDB60262; HMDB61093 | M+NH4; M+H | Glutamine | M+H |
| C18 negative | 112.0404 | 289.8 | 0.09 (0.95) | -0.09 (1.04) | 0.17 (0.02, 0.33) | 0.0295 | 0.5832 | 1-Pyrroline-5-carboxylic acid; 1-Pyrroline-2-carboxylic acid | HMDB01301; HMDB06875 | M-H | - | - |
| C18 negative | 112.0517 | 26.1 | -0.06 (1.21) | 0.06 (0.73) | -0.13 (-0.25, -0.01) | 0.0296 | 0.5832 | Creatinine | HMDB00562 | M-H | - | - |
| C18 negative | 184.0729 | 293.2 | 0.09 (0.73) | -0.09 (1.21) | 0.18 (0.02, 0.35) | 0.0296 | 0.5832 | (S)-N-(45-Dihydro-1-methyl-4-oxo-1H-imidazol-2-yl)alanine | HMDB34912 | M-H | - | - |
| HILIC positive | 152.0319 | 76.9 | -0.08 (0.97) | 0.08 (1.03) | -0.16 (-0.31, -0.02) | 0.0296 | 0.6262 | Pyroglutamic acid; Pyrrolidonecarboxylic acid; Pyrroline hydroxycarboxylic acid; N-Acryloylglycine; 1-Pyrroline-4-hydroxy-2-carboxylate; 5-Oxoprolinate; dimethadione | HMDB00267; HMDB00805; HMDB01369; HMDB01843; HMDB02234; HMDB60262; HMDB61093 | M+Na | - | - |
| HILIC positive | 445.3674 | 27.3 | -0.09 (1.12) | 0.09 (0.86) | -0.19 (-0.36, -0.02) | 0.0297 | 0.6262 | 3-beta-Hydroxy-4-beta-methyl-5-alpha-cholest-7-ene-4-alpha-carboxylate; 4alpha-Carboxy-4beta-methyl-5alpha-cholesta-8-en-3beta-ol; (3beta5alpha6beta22E24R)-23-Methylergosta-722-diene-356-triol; Schleicherastatin 5 | HMDB11662; HMDB12165; HMDB33633; HMDB35804 | M+H | - | - |
| HILIC positive | 130.0408 | 75 | -0.08 (0.97) | 0.08 (1.02) | -0.16 (-0.31, -0.02) | 0.0298 | 0.6270 | Flucytosine | HMDB15231 | M+H | - | - |
| C18 negative | 277.0114 | 217.8 | -0.09 (1) | 0.09 (1) | -0.18 (-0.34, -0.02) | 0.0299 | 0.5832 | Fenthion | HMDB33209 | M-H | - | - |
| C18 negative | 204.9954 | 21.9 | -0.09 (1.07) | 0.09 (0.92) | -0.18 (-0.35, -0.02) | 0.0302 | 0.5832 | O-methoxycatechol-O-sulphate | HMDB60013 | M-H_[+2] | - | - |
| C18 negative | 135.0451 | 289.5 | 0.09 (0.99) | -0.09 (1) | 0.17 (0.02, 0.33) | 0.0303 | 0.5832 | Phenylacetic acid; 2-Methylbenzoic acid; 4-Hydroxyphenylacetaldehyde; 4-Methylbenzoic acid; 4-Methoxybenzaldehyde; 3-Methoxybenzaldehyde; 3-(5-Methyl-2-furyl)prop-2-enal; 2-Hydroxyacetophenone; 2-Hydroxy-4-methylbenzaldehyde; 4-(2-Furanyl)-3-buten-2-one; 2-Methoxybenzaldehyde; Methyl benzoate; alpha-Methyl-2-furanacrolein; 3-(3-Furanyl)-2-methyl-2-propenal; Phenyl acetate; Benzyl formate | HMDB00209; HMDB02340; HMDB03767; HMDB29635; HMDB29686; HMDB31459; HMDB32400; HMDB32568; HMDB32603; HMDB33128; HMDB33766; HMDB33968; HMDB38176; HMDB40638; HMDB40733; HMDB41485 | M-H | - | - |
| C18 negative | 180.0302 | 295.2 | 0.09 (0.94) | -0.09 (1.05) | 0.19 (0.02, 0.36) | 0.0305 | 0.5832 | 2-Methyl-3-hydroxy-5-formylpyridine-4-carboxylate; 24-Dihydroxy-2H-14-benzoxazin-3(4H)-one; (R)-27-Dihydroxy-2H-14-benzoxazin-3(4H)-one | HMDB06954; HMDB33733; HMDB38054 | M-H | - | - |
| C18 negative | 291.0831 | 20 | -0.09 (1.18) | 0.09 (0.77) | -0.18 (-0.34, -0.02) | 0.0308 | 0.5832 | Edetic Acid | HMDB15109 | M-H | - | - |
| HILIC positive | 276.0431 | 63.8 | -0.09 (1.06) | 0.09 (0.93) | -0.18 (-0.35, -0.02) | 0.0309 | 0.6365 | N-(2-formyl-3-chlorophenyl)anthranilic acid | HMDB60006 | M+H | - | - |
| C18 negative | 847.7781 | 41.2 | -0.08 (1.12) | 0.08 (0.86) | -0.16 (-0.31, -0.02) | 0.0312 | 0.5854 | Glycerol triheptadecanoate; TG(14:015:022:0); TG(14:022:015:0); TG(15:014:022:0); TG(15:016:020:0); TG(15:018:018:0); TG(15:020:016:0); TG(16:015:020:0); TG(18:015:018:0) | HMDB31106; HMDB42096; HMDB42212; HMDB42964; HMDB43021; HMDB43049; HMDB43077; HMDB43859; HMDB44695 | M-H | - | - |
| HILIC positive | 527.1418 | 74.1 | -0.09 (1.2) | 0.09 (0.74) | -0.18 (-0.35, -0.02) | 0.0312 | 0.6365 | Melleolide D | HMDB37042 | M+2Na-H | - | - |
| C18 negative | 112.9857 | 35.1 | -0.08 (1.07) | 0.08 (0.92) | -0.17 (-0.32, -0.02) | 0.0312 | 0.5854 | Trifluoroacetic acid | HMDB14118 | M-H | - | - |
| C18 negative | 250.0931 | 50 | -0.08 (1.09) | 0.08 (0.89) | -0.17 (-0.32, -0.01) | 0.0317 | 0.5865 | Muramic acid; N-(1-Deoxy-1-fructosyl)alanine | HMDB03254; HMDB38662 | M-H | - | - |
| HILIC positive | 152.0512 | 74.7 | -0.08 (1.13) | 0.08 (0.84) | -0.17 (-0.32, -0.01) | 0.0317 | 0.6365 | Mevalonic acid; (R) 23-Dihydroxy-3-methylvalerate; Glycerol 1-propanoate; (R)-mevalonate; (2-Methoxyethoxy)propanoic acid | HMDB00227; HMDB12140; HMDB31639; HMDB59629; HMDB59705 | M+H_[+3] | - | - |
| HILIC positive | 148.0827 | 74.9 | -0.09 (1.14) | 0.09 (0.83) | -0.17 (-0.33, -0.02) | 0.0317 | 0.6365 | L-Lysine; L-Glutamine; Ureidoisobutyric acid; D-Lysine; D-Glutamine; Alanylglycine; (3S)-36-Diaminohexanoate; (3S5S)-35-Diaminohexanoate; Alanyl-Glycine | HMDB00182; HMDB00641; HMDB02031; HMDB03405; HMDB03423; HMDB06899; HMDB12114; HMDB12115; HMDB28687 | M+H_[+1] | - | - |
| C18 negative | 306.0853 | 154.4 | 0.09 (1.02) | -0.09 (0.97) | 0.18 (0.02, 0.34) | 0.0320 | 0.5865 | (S)-Succinyldihydrolipoamide | HMDB01177 | M-H | - | - |
| HILIC positive | 219.142 | 40.7 | -0.09 (1.17) | 0.09 (0.79) | -0.17 (-0.33, -0.01) | 0.0322 | 0.6413 | Propionylcarnitine; (S)-3-Mercaptohexyl pentanoate; (S)-3-Methylthiohexyl butyrate | HMDB00824; HMDB38945; HMDB38946 | M+H_[+1]; M+H | - | - |
| HILIC positive | 119.0604 | 286.2 | -0.09 (0.96) | 0.09 (1.03) | -0.18 (-0.35, -0.02) | 0.0323 | 0.6414 | 7-Azaindolizine; 5H-Cyclopentapyrazine | HMDB33576; HMDB40040 | M+H | - | - |
| HILIC positive | 132.0542 | 75 | -0.08 (0.98) | 0.08 (1.02) | -0.16 (-0.31, -0.01) | 0.0324 | 0.6414 | Pipecolic acid; Pyroglutamic acid; L-Pipecolic acid; Pyrrolidonecarboxylic acid; Pyrroline hydroxycarboxylic acid; N-Acryloylglycine; 1-Pyrroline-4-hydroxy-2-carboxylate; N4-Acetylaminobutanal; D-Pipecolic acid; Vigabatrin; L-trans-4-Methyl-2-pyrrolidinecarboxylic acid; 2-Pyrrolidineacetic acid; 3-Acetamidobutanal; 5-Oxoprolinate; dimethadione | HMDB00070; HMDB00267; HMDB00716; HMDB00805; HMDB01369; HMDB01843; HMDB02234; HMDB04226; HMDB05960; HMDB15212; HMDB29435; HMDB29444; HMDB59649; HMDB60262; HMDB61093 | M+H_[+2] | - | - |

| | | | | | | | | | | | | |
|---|---|---|---|---|---|---|---|---|---|---|---|---|
| C18 negative | 200.041 | 22.8 | -0.09 (1) | 0.09 (1) | -0.17 (-0.33, -0.01) | 0.0328 | 0.5897 | Potassium 2-(1-ethoxy) ethoxypropanoate | HMDB37192 | M-H_[-1] | - | - |
| C18 negative | 253.0928 | 24.9 | -0.09 (0.99) | 0.09 (1.01) | -0.18 (-0.34, -0.01) | 0.0329 | 0.5897 | Galactosylglycerol; (2R)-1-O-beta-D-Galactopyranosylglycerol | HMDB06790; HMDB38664 | M-H | - | - |
| HILIC positive | 116.107 | 97.5 | 0.09 (0.74) | -0.09 (1.2) | 0.19 (0.02, 0.36) | 0.0332 | 0.6443 | N-(3-Methylbutyl)acetamide; N-(2-Methylpropyl)acetamide | HMDB31651; HMDB34203 | M+H | - | - |
| C18 negative | 389.2698 | 243.5 | -0.09 (1.05) | 0.09 (0.94) | -0.17 (-0.34, -0.01) | 0.0337 | 0.5934 | 12-Ketodeoxycholic acid; 7-Hydroxy-3-oxocholanoic acid; Nutriacholic acid; 7a-Hydroxy-3-oxo-5b-cholanoic acid; 9-Carboxy-alpha-chromanol; D8-Merulinic acid A | HMDB00328; HMDB00460; HMDB00467; HMDB00503; HMDB12866; HMDB41454 | M-H | - | - |
| HILIC positive | 181.0854 | 24.2 | -0.08 (1.09) | 0.08 (0.9) | -0.17 (-0.32, -0.01) | 0.0337 | 0.6443 | 3-Methoxybenzenepropanoic acid; Coniferyl alcohol; 4-Ethoxy-3-methoxybenzaldehyde; 2-Phenyl-13-dioxolane-4-methanol; 1-(24-Dihydroxyphenyl)-1-butanone; Benzaldehyde glyceryl acetal; Propylparaben; Ethyl p-anisate; 4-Methoxybenzyl acetate; 4-Hydroxy-266-trimethyl-3-oxo-14-cyclohexadiene-1-carboxaldehyde; Propyl 2-furanacrylate; 3-(3-Hydroxyphenyl)-2-methylpropionic acid | HMDB11751; HMDB12915; HMDB29685; HMDB31240; HMDB32128; HMDB32174; HMDB32574; HMDB32640; HMDB33761; HMDB37602; HMDB37733; HMDB60734 | M+H | - | - |
| HILIC positive | 198.1277 | 27.5 | -0.08 (1.04) | 0.08 (0.96) | -0.16 (-0.3, -0.01) | 0.0340 | 0.6443 | 2-(3-Phenylpropyl)pyridine | HMDB36173 | M+H | - | - |
| HILIC positive | 137.1325 | 28 | -0.09 (0.95) | 0.09 (1.04) | -0.17 (-0.33, -0.01) | 0.0343 | 0.6447 | D-Limonene; ()-Limonene; Gamma-terpinene; ()-a-Pinene; cis-Ocimene; trans-Ocimen; Polylimonene; 5-Isosylveterpinolene; ()-alpha-Carene; (-)-4(10)-Thujene; (S)-alpha-Phellandrene; (-)-alpha-Carene; alpha-Carene; (-)-77-Dimethyl-2-methylenebicyclo221heptane; (-)-alpha-Pinene; alpha-Phellandrene; (R)-alpha-Phellandrene; ()-4(10)-Thujene; beta-Phellandrene; (Z)-37-Dimethyl-137-octatriene; (-)-3-Thujene; (-)-beta-Pinene; beta-Pinene; Terpinolene; alpha-Terpinene; beta-Terpinene; Sylveterpinolene; 177-Trimethyltricyclo221026heptane; p-Mentha-24-diene; 7-Methyl-3-methylene-16-octadiene; Artemisiatriene; (-)-beta-Phellandrene; ()-beta-Phellandrene; ()-4-Carene; Camphene; 2-Carene | HMDB03375; HMDB04321; HMDB05806; HMDB06525; HMDB30088; HMDB30089; HMDB32473; HMDB33200; HMDB34697; HMDB34928; HMDB34970; HMDB35087; HMDB35619; HMDB35625; HMDB35658; HMDB35850; HMDB35851; HMDB36076; HMDB36081; HMDB36104; HMDB36116; HMDB36559; HMDB36560; HMDB36994; HMDB36995; HMDB36997; HMDB37046; HMDB38121; HMDB38150; HMDB38169; HMDB41574; HMDB41633; HMDB41634; HMDB59726; HMDB59839; HMDB59884 | M+H | - | - |
| HILIC positive | 169.0089 | 122.5 | -0.08 (0.94) | 0.08 (1.05) | -0.15 (-0.29, -0.01) | 0.0345 | 0.6447 | Desflurane; Hexafluoroisopropanol | HMDB15320; HMDB60648 | M+H | - | - |
| HILIC positive | 148.0798 | 75.8 | -0.08 (1) | 0.08 (1) | -0.16 (-0.31, -0.01) | 0.0346 | 0.6447 | L-Lysine; L-Glutamine; Ureidoisobutyric acid; D-Lysine; D-Glutamine; Alanylglycine; (3S)-36-Diaminohexanoate; (3S5S)-35-Diaminohexanoate; Alanyl-Glycine | HMDB00182; HMDB00641; HMDB02031; HMDB03405; HMDB03423; HMDB06899; HMDB12114; HMDB12115; HMDB28687 | M+H_[+1] | - | - |
| HILIC positive | 143.0704 | 178.3 | 0.09 (0.75) | -0.09 (1.19) | 0.18 (0.01, 0.35) | 0.0349 | 0.6447 | 5-Ethyl-3-hydroxy-4-methyl-2(5H)-furanone; Homofureanol; Mesifurane; Erinapyrone A; Erinapyrone B | HMDB33551; HMDB33736; HMDB34230; HMDB41426; HMDB41427 | M+H | - | - |
| C18 negative | 229.1231 | 105.8 | 0.06 (1) | -0.06 (0.99) | 0.12 (0.01, 0.23) | 0.0349 | 0.6002 | 812-Epoxy-4(15)711-eudesmatrien-1-one; (2E8Z)-Decadiene-46-diyn-1-yl 3-methylbutanoate; Curzerenone; Furanodienone; Cyclohexyl cinnamate | HMDB33235; HMDB35425; HMDB36155; HMDB36768; HMDB37706 | M-H | - | - |
| C18 negative | 171.0776 | 294.3 | -0.09 (1.17) | 0.09 (0.78) | -0.17 (-0.34, -0.01) | 0.0352 | 0.6025 | Glycylproline; L-prolyl-L-glycine | HMDB00721; HMDB11178 | M-H | - | - |
| C18 negative | 573.4522 | 270.2 | -0.08 (1.07) | 0.08 (0.92) | -0.16 (-0.31, -0.01) | 0.0353 | 0.6025 | DG(15:018:4(6Z9Z12Z15Z)0:0); DG(18:4(6Z9Z12Z15Z)15:00:0); DG(15:00:018:4n3) | HMDB07077; HMDB07329; HMDB56004 | M-H | - | - |
| C18 negative | 429.301 | 182.1 | -0.09 (1.15) | 0.09 (0.81) | -0.17 (-0.33, -0.01) | 0.0356 | 0.6037 | MG(0:024:6(6Z9Z12Z15Z18Z21Z)0:0); MG(24:6(6Z9Z12Z15Z18Z21Z)0:00:0); 7 alpha-Hydroxy-3-oxo-4-cholestenoate; Australigenin; Barogenin; Schidigeragenin C; 25-Hydroxyvitamin D3-2623-lactol; 24-Oxo-1alpha25-dihydroxyvitamin D3 | HMDB11560; HMDB11590; HMDB12458; HMDB30066; HMDB34403; HMDB36249; HMDB60127; HMDB60128 | M-H | - | - |
| C18 negative | 174.0409 | 292.7 | 0.09 (0.95) | -0.09 (1.04) | 0.18 (0.01, 0.35) | 0.0356 | 0.6037 | N-Acetyl-L-aspartic acid; N-Formyl-L-glutamic acid; Berteroin; D-N-(Carboxyacetyl)alanine; 2-Amino-3-oxoadipate | HMDB00812; HMDB03470; HMDB31577; HMDB39163; HMDB60273 | M-H | - | - |
| C18 negative | 329.066 | 18.9 | -0.03 (1.05) | 0.03 (0.95) | -0.07 (-0.13, 0) | 0.0357 | 0.6037 | 37-Dimethylquercetin; Aflatoxin G2; Aflatoxin M2; Demethoxysudachitin; Prudomestin; 34-Dihydroxyphenacyl caffeate; 135-Trihydroxy-67-dimethoxy-2-methylanthraquinone; Trifolian; Aflatoxin B2a; Pilosin; Caryatin; 34-Di-O-methylquercetin; Cicerin; 57-Dihydroxy-84-dimethoxyisoflavone | HMDB29263; HMDB30475; HMDB30480; HMDB30696; HMDB30811; HMDB31706; HMDB33268; HMDB34009; HMDB35598; HMDB37096; HMDB37351; HMDB37354; HMDB38659; HMDB41690 | M-H | - | - |
| C18 negative | 420.2539 | 242.5 | 0.07 (0.94) | -0.07 (1.06) | 0.14 (0.01, 0.26) | 0.0363 | 0.6067 | PC-M6 | HMDB38568 | M-H | - | - |
| HILIC positive | 828.5506 | 37.5 | -0.09 (1.16) | 0.09 (0.8) | -0.17 (-0.34, -0.01) | 0.0364 | 0.6519 | PC(18:3(6Z9Z12Z)22:6(4Z7Z10Z13Z16Z19Z)); PC(18:3(9Z12Z15Z)22:6(4Z7Z10Z13Z16Z19Z)); PC(18:4(6Z9Z12Z15Z)22:5(4Z7Z10Z13Z16Z)); | HMDB08189; HMDB08222; HMDB08253; HMDB08254; HMDB08445; HMDB08478; | M+H | - | - |

| | | | | | | | | | | | | |
|---|---|---|---|---|---|---|---|---|---|---|---|---|
| | | | | | | | | PC(18:4(6Z9Z12Z15Z)22:5(7Z10Z13Z16Z19Z)); PC(20:4(5Z8Z11Z14Z)20:5(5Z8Z11Z14Z17Z)); PC(20:4(8Z11Z14Z17Z)20:5(5Z8Z11Z14Z17Z)); PC(20:5(5Z8Z11Z14Z17Z)20:4(5Z8Z11Z14Z)); PC(20:5(5Z8Z11Z14Z17Z)20:4(8Z11Z14Z17Z)); PC(22:5(4Z7Z10Z13Z16Z)18:4(6Z9Z12Z15Z)); PC(22:5(7Z10Z13Z16Z19Z)18:4(6Z9Z12Z15Z)); PC(22:6(4Z7Z10Z13Z16Z19Z)18:3(6Z9Z12Z)); PC(22:6(4Z7Z10Z13Z16Z19Z)18:3(9Z12Z15Z)) | HMDB08509; HMDB08510; HMDB08667; HMDB08700; HMDB08731; HMDB08732 | | | |
| HILIC positive | 229.1546 | 51.3 | -0.07 (1.1) | 0.07 (0.89) | -0.14 (-0.26, -0.01) | 0.0365 | 0.6519 | L-isoleucyl-L-proline ; L-leucyl-L-proline ; Isoleucyl-Proline; Leucyl-Proline | HMDB11174; HMDB11175; HMDB28915; HMDB28937 | M+H | - | - |
| C18 negative | 363.2549 | 166.1 | -0.09 (1.03) | 0.09 (0.96) | -0.17 (-0.33, -0.01) | 0.0366 | 0.6087 | 6-O-Acetylaustroinulin; 7-O-Acetylaustroinulin | HMDB36803; HMDB36804 | M-H | - | - |
| C18 negative | 445.3319 | 235.9 | -0.07 (1.13) | 0.07 (0.84) | -0.15 (-0.29, -0.01) | 0.0368 | 0.6090 | 13-Carboxy-gamma-tocopherol; (3beta5alpha6beta7alpha22E24R)-Ergosta-822-diene-3567-tetrol; (3beta5alpha6alpha9alpha22E24R)-Ergosta-722-diene-3569-tetrol; 359-Trihydroxyergost-7-en-6-one; Secasterone; 3-Dehydroteasterone | HMDB12557; HMDB32107; HMDB32123; HMDB32875; HMDB40999; HMDB41527 | M-H | - | - |
| HILIC positive | 114.055 | 78.7 | -0.08 (1.01) | 0.08 (0.99) | -0.16 (-0.31, -0.01) | 0.0373 | 0.6529 | 1-Pyrroline-5-carboxylic acid; 1-Pyrroline-2-carboxylic acid | HMDB01301; HMDB06875 | M+H | - | - |
| C18 negative | 331.1912 | 142 | -0.09 (1.07) | 0.09 (0.91) | -0.18 (-0.35, -0.01) | 0.0379 | 0.6164 | Carnosic acid; 11b-Hydroxyprogesterone; 7-Carboxy-gamma-tocotrienol; Hulupone; Adhulupone; (1(10)E4E6a9b)-9-(2-Methylbutanoyloxy)-1(10)411(13)-germacratrien-126-olide; (1(10)E4E6a9b)-9-(3-Methylbutanoyloxy)-1(10)411(13)-germacratrien-126-olide; Petasitin; 6-Angeloylfuranofukinol; 4-Deoxycohumulone; ent-7alpha12beta-Dihydroxy-16-kauren-196beta-olide; ent-15-Kaurene-1719-dioic acid; 718-Dihydroxykaurenolide; (9Z11S16S)-1-Acetoxy-917-octadecadiene-1214-diyne-1116-diol | HMDB02358; HMDB04031; HMDB12851; HMDB30103; HMDB30136; HMDB31374; HMDB31375; HMDB35637; HMDB36146; HMDB36623; HMDB36700; HMDB36726; HMDB36761; HMDB38966 | M-H | - | - |
| C18 negative | 475.3062 | 182.4 | -0.09 (1.18) | 0.09 (0.77) | -0.17 (-0.33, -0.01) | 0.0381 | 0.6164 | Polyporusterone B; Polyporusterone C | HMDB38496; HMDB38497 | M-H | - | - |
| HILIC positive | 149.0558 | 94.5 | -0.07 (1.1) | 0.07 (0.88) | -0.14 (-0.28, -0.01) | 0.0381 | 0.6551 | Hydroxylated lecithin | HMDB32332 | M+H | - | - |
| HILIC positive | 256.9967 | 69.4 | 0.07 (0.94) | -0.07 (1.06) | 0.15 (0.01, 0.29) | 0.0383 | 0.6551 | Ascorbic acid-2-sulfate | HMDB60649 | M+H | - | - |
| C18 negative | 316.9477 | 40.3 | -0.07 (1.06) | 0.07 (0.94) | -0.14 (-0.27, -0.01) | 0.0384 | 0.6164 | Mitotane | HMDB14786 | M-H | - | - |
| C18 negative | 476.3096 | 183 | -0.09 (1.16) | 0.09 (0.8) | -0.17 (-0.33, -0.01) | 0.0385 | 0.6164 | Polyporusterone B; Polyporusterone C; Gentamicin | HMDB38496; HMDB38497; HMDB14936 | M-H_[-1]; M-H | - | - |
| HILIC positive | 489.3566 | 27.5 | -0.08 (1.04) | 0.08 (0.96) | -0.17 (-0.33, -0.01) | 0.0390 | 0.6551 | Pitheduloside I; Arjunolic acid; Camelliagenin B; Madasiatic acid; Esculentic acid (Diplazium); Hovenolactone; Centellasapogenol A; Glyyunnansapogenin B; Euscaphic acid; (3beta6alpha19alpha)-3619-Trihydroxy-12-ursen-28-oic acid; Ganoderiol D; Ananasic acid; 16beta-Hydroxystellatogenin; 21beta-Hydroxyhederagenin | HMDB34036; HMDB34502; HMDB34531; HMDB35118; HMDB35782; HMDB35957; HMDB36311; HMDB36639; HMDB36651; HMDB36961; HMDB37782; HMDB38116; HMDB40391; HMDB41039 | M+H | - | - |
| C18 negative | 125.0245 | 23.3 | -0.09 (1.09) | 0.09 (0.9) | -0.18 (-0.34, -0.01) | 0.0391 | 0.6184 | 123-Trihydroxybenzene; 135-Trihydroxybenzene; Methyl 2-furoate; Maltol; 4-Hydroxy-2H-pyran-3-carboxaldehyde; (Z)-Tamarindienal; 5-Hydroxymethyl-2-furancarboxaldehyde; 5-Methylfuran-2-carboxylic acid | HMDB13674; HMDB13675; HMDB29750; HMDB30776; HMDB31256; HMDB32651; HMDB34355; HMDB59735 | M-H | - | - |
| C18 negative | 424.1976 | 106.5 | -0.09 (1.13) | 0.09 (0.85) | -0.18 (-0.35, -0.01) | 0.0403 | 0.6197 | O-Desmethyl-tramado glucuronide; NO-Didesmethylvenlafaxine glucuronide | HMDB60856; HMDB61167 | M-H | - | - |
| C18 negative | 383.1532 | 24.5 | -0.07 (1.1) | 0.07 (0.88) | -0.15 (-0.28, -0.01) | 0.0404 | 0.6197 | 3b16a-Dihydroxyandrostenone sulfate | HMDB00386 | M-H | - | - |
| C18 negative | 266.1841 | 215 | 0.08 (0.86) | -0.08 (1.12) | 0.16 (0.01, 0.32) | 0.0406 | 0.6197 | 4-Hydroxy-3-methoxy-210-bisaboladien-9-one; Tetranor 12-HETE | HMDB33917; HMDB60055 | M-H_[-1] | - | - |
| C18 negative | 583.4199 | 221.6 | -0.08 (1.05) | 0.08 (0.94) | -0.15 (-0.3, -0.01) | 0.0407 | 0.6197 | Antibiotic X 14889A | HMDB33336 | M-H | - | - |
| C18 negative | 265.1807 | 214.7 | 0.08 (0.92) | -0.08 (1.07) | 0.15 (0.01, 0.29) | 0.0409 | 0.6197 | 4-Hydroxy-3-methoxy-210-bisaboladien-9-one; Tetranor 12-HETE | HMDB33917; HMDB60055 | M-H | - | - |
| HILIC positive | 293.1454 | 75.1 | -0.08 (1.02) | 0.08 (0.98) | -0.16 (-0.31, -0.01) | 0.0411 | 0.6557 | L-Glutamine; Ureidoisobutyric acid; D-Glutamine; Alanylglycine; Alanyl-Glycine | HMDB00641; HMDB02031; HMDB03423; HMDB06899; HMDB28687 | 2M+H | - | - |
| C18 negative | 209.0815 | 21.8 | -0.08 (1.01) | 0.08 (0.98) | -0.16 (-0.31, -0.01) | 0.0411 | 0.6197 | Sinapyl alcohol; 5-(35)-Dihydroxyphenyl-gamma-valerolactone; 34-Dihydroxyphenylvaleric acid; 3-Methyl-1-(246-trihydroxyphenyl)-1-butanone; 2-Hydroxy-46-dimethoxy-3-methylacetophenone; 2-Methoxy-4-(4-methyl-13-dioxolan-2-yl)phenol; 2-Methoxy-3-(4-methoxyphenyl)propanoic acid; Bancroftinone; 3-(4-Hydroxy-3-methoxyphenyl)-2-methylpropionic acid | HMDB13070; HMDB29187; HMDB29233; HMDB33798; HMDB34047; HMDB36199; HMDB39428; HMDB41406; HMDB60737 | M-H | - | - |
| C18 negative | 464.2775 | 190.2 | 0.08 (0.88) | -0.08 (1.1) | 0.16 (0.01, 0.31) | 0.0414 | 0.6197 | LysoPC(14:1(9Z)) | HMDB10380 | M-H | - | - |
| C18 negative | 613.1796 | 146.1 | -0.08 (0.98) | 0.08 (1.02) | -0.15 (-0.3, -0.01) | 0.0416 | 0.6197 | Catechin 34-diglucoside; Catechin 35-diglucoside; Catechin 37-diglucoside | HMDB37950; HMDB37951; HMDB37952 | M-H | - | - |

| | | | | | | | | | | | | |
|---|---|---|---|---|---|---|---|---|---|---|---|---|
| C18 negative | 297.2432 | 211.2 | -0.08 (1.12) | 0.08 (0.85) | -0.15 (-0.3, -0.01) | 0.0417 | 0.6197 | 3-Oxooctadecanoic acid; 9-Oxooctadecanoic acid; 10-Oxooctadecanoic acid; 11-Oxooctadecanoic acid; 5-Hexyltetrahydro-2-furanoctanoic acid; 5-Oxooctadecanoic acid; Ricinoleic acid | HMDB10736; HMDB30979; HMDB30980; HMDB30981; HMDB31127; HMDB34074; HMDB34297 | M-H | - | - |
| C18 negative | 338.3145 | 300.7 | -0.08 (1.05) | 0.08 (0.94) | -0.16 (-0.31, -0.01) | 0.0417 | 0.6197 | Erucic acid; Cetoleic acid; Catelaidic acid; Phytyl acetate; 79-Docosanedione; 68-Docosanedione; 57-Docosanedione; 46-Docosanedione | HMDB02068; HMDB02884; HMDB31013; HMDB32470; HMDB35566; HMDB35567; HMDB35568; HMDB35569 | M-H_[-1] | - | - |
| HILIC positive | 101.0709 | 75.7 | -0.08 (1) | 0.08 (1) | -0.16 (-0.31, -0.01) | 0.0422 | 0.6557 | N-Nitroso-pyrrolidine; Gyromitrin | HMDB31642; HMDB33952 | M+H | - | - |
| HILIC positive | 585.4296 | 27.1 | 0.08 (0.94) | -0.08 (1.05) | 0.16 (0.01, 0.32) | 0.0422 | 0.6557 | Cryptochrome; Cryptoxanthin diepoxide; Cryptoxanthin 56:58-diepoxide; (3S3R4xi)-betabeta-Carotene-334-triol; Cucurbitaxanthin A; Antheraxanthin A; Capsanthin; Flavoxanthin; Triphasiaxanthin; Prenigroxanthin; Mutatoxanthin; Lutein 56-epoxide | HMDB29857; HMDB29896; HMDB30537; HMDB30586; HMDB35319; HMDB35831; HMDB36590; HMDB36868; HMDB36914; HMDB37121; HMDB37564; HMDB41590 | M+H | - | - |
| C18 negative | 283.1552 | 294.5 | 0.09 (0.81) | -0.09 (1.15) | 0.18 (0.01, 0.35) | 0.0422 | 0.6197 | Alpha-dihydroartemisinin; Dihydroartemisinin (DHA) | HMDB60593; HMDB61088 | M-H | - | - |
| HILIC positive | 277.1796 | 27.9 | -0.09 (1.16) | 0.09 (0.8) | -0.17 (-0.34, -0.01) | 0.0425 | 0.6557 | Cyclandelate; 8-Shogaol; Panaquinquecol 2; Ginsenoyne C; Ginsenoyne K; Panaquinquecol 7 | HMDB15586; HMDB31463; HMDB38938; HMDB38994; HMDB40375; HMDB41201 | M+H | - | - |
| C18 negative | 433.2808 | 174.1 | -0.08 (1.06) | 0.08 (0.93) | -0.17 (-0.33, -0.01) | 0.0425 | 0.6197 | Polysorbate 60 | HMDB37183 | M-H | - | - |
| C18 negative | 105.0378 | 26.1 | -0.07 (1.03) | 0.07 (0.97) | -0.15 (-0.29, -0.01) | 0.0425 | 0.6197 | 3-(Methylthio)-1-propanol; 3-Mercapto-2-butanol; 4-Mercapto-2-butanol | HMDB31716; HMDB40185; HMDB41475 | M-H | - | - |
| HILIC positive | 180.102 | 247.4 | -0.09 (1.28) | 0.09 (0.59) | -0.18 (-0.34, -0.01) | 0.0426 | 0.6559 | 2(N)-Methyl-norsalsolinol; (R)-Salsolinol; alpha-Methylphenylalanine; N-methylphenylalanine; Maltoxazine; 35-Dimethylphenyl methylcarbamate; Ethyl N-methylanthranilate; 123456-Hexahydro-5-(1-hydroxyethylidene)-7H-cyclopentabpyridin-7-one; 23-Dihydro-5-(3-hydroxypropanoyl)-1H-pyrrolizine; 34-Methylenedioxyamphetamine; Salsolinol | HMDB01189; HMDB05199; HMDB29223; HMDB29224; HMDB30372; HMDB31811; HMDB33763; HMDB39655; HMDB40021; HMDB41931; HMDB42012 | M+H | - | - |
| C18 negative | 349.2746 | 259.2 | 0.08 (0.87) | -0.08 (1.11) | 0.17 (0.01, 0.33) | 0.0426 | 0.6197 | 34-Dimethyl-5-pentyl-2-furanundecanoic acid | HMDB31126 | M-H | - | - |
| C18 negative | 435.3479 | 282.2 | -0.08 (1.1) | 0.08 (0.88) | -0.16 (-0.32, -0.01) | 0.0431 | 0.6197 | 5a-Cholestane-3a7a12a25-tetrol; 5b-Cholestane-3a7a12a25-tetrol; Cholestane-371225-tetrol; 27-Deoxy-5b-cyprinol; 5b-Cholestane-3a7a12a23-Tetrol; 27-Deoxy-5alpha-cyprinol | HMDB00520; HMDB00524; HMDB00655; HMDB01231; HMDB01968; HMDB29851 | M-H | - | - |
| C18 negative | 171.1391 | 295.2 | 0.09 (0.77) | -0.09 (1.18) | 0.18 (0.01, 0.35) | 0.0431 | 0.6197 | Capric acid; 11-Diethoxy-2-hexene; Isopentyl isopentanoate; Methyl nonanoate; 7-Hydroxy-37-dimethyloctanal; Pentyl 3-methylbutanoate; Isooctyl acetate; Acetaldehyde 13-octanediol acetal; Ethyl 2-ethylhexanoate; Heptanal propyleneglycol acetal; Hexanal butane-23-diol acetal; Hexyl butyrate; Propyl heptanoate; 4-Methylnonanoic acid; trans-p-Menthane-18-diol; (1R2S3S4R)-p-Menthane-23-diol; 4-Ethyloctanoic acid; p-Menthane-38-diol; 2-Methylpropyl hexanoate; (1S3R4R)-p-Menthane-13-diol; Hexyl 2-methylpropanoate; 2-Methylbutyl 3-methylbutanoate; Octyl acetate; (Z)-1-(1-Ethoxyethoxy)-3-hexene; 2-Methylbutyl 2-methylbutanoate; 3-Methylbutyl 2-methylbutanoate; ()-3-Octyl acetate; Ethyl octanoate; Butyl hexanoate; 3-Methylbutyl pentanoate; 26-Dimethyl-7-octene-26-diol | HMDB00511; HMDB29760; HMDB30029; HMDB31264; HMDB31739; HMDB32045; HMDB32070; HMDB32155; HMDB32264; HMDB32302; HMDB32315; HMDB33620; HMDB34457; HMDB34849; HMDB35595; HMDB35723; HMDB36136; HMDB36145; HMDB36228; HMDB37387; HMDB37849; HMDB38038; HMDB38602; HMDB38783; HMDB39218; HMDB40157; HMDB40167; HMDB40195; HMDB40211; HMDB40529; HMDB41216 | M-H | - | - |
| C18 negative | 395.1897 | 118.5 | -0.08 (1.1) | 0.08 (0.88) | -0.15 (-0.3, 0) | 0.0432 | 0.6197 | Pregnenolone sulfate; 3beta-Hydroxypregn-5-en-20-one sulfate | HMDB00774; HMDB60382 | M-H | - | - |
| HILIC positive | 820.6212 | 30.1 | -0.09 (1.22) | 0.09 (0.7) | -0.18 (-0.35, -0.01) | 0.0434 | 0.6587 | PC(22:4(7Z10Z13Z16Z)P-18:1(11Z)); PC(22:4(7Z10Z13Z16Z)P-18:1(9Z)); PC(22:5(4Z7Z10Z13Z16Z)P-18:0); PC(22:5(7Z10Z13Z16Z19Z)P-18:0); PC(P-18:022:5(4Z7Z10Z13Z16Z)); PC(P-18:022:5(7Z10Z13Z16Z19Z)); PC(P-18:1(11Z)22:4(7Z10Z13Z16Z)); PC(P-18:1(9Z)22:4(7Z10Z13Z16Z)); PC(o-18:022:6(4Z7Z10Z13Z16Z19Z)) | HMDB08654; HMDB08655; HMDB08686; HMDB08719; HMDB11260; HMDB11261; HMDB11292; HMDB11325; HMDB13422 | M+H | - | - |
| C18 negative | 575.3591 | 152.4 | -0.08 (1.18) | 0.08 (0.78) | -0.17 (-0.33, 0) | 0.0440 | 0.6197 | Collettiside I | HMDB29310 | M-H | - | - |
| C18 negative | 204.0664 | 23.5 | -0.08 (1.04) | 0.08 (0.95) | -0.16 (-0.31, 0) | 0.0441 | 0.6197 | Indolelactic acid; 5-Methoxyindoleacetate; Cinnamoylglycine; Edulitine; Methyl 1-methoxy-1H-indole-3-carboxylate; 3-Indolehydracrylic acid | HMDB00671; HMDB04096; HMDB11621; HMDB30179; HMDB32755; HMDB59765 | M-H | - | - |
| HILIC positive | 482.3236 | 37.2 | 0.08 (0.93) | -0.08 (1.06) | 0.16 (0, 0.32) | 0.0441 | 0.6587 | LysoPC(15:0); LysoPE(0:018:0); LysoPE(18:00:0) | HMDB10381; HMDB11129; HMDB11130 | M+H | - | - |
| HILIC positive | 203.0342 | 60.6 | -0.08 (1.09) | 0.08 (0.9) | -0.16 (-0.33, 0) | 0.0441 | 0.6587 | Bergaptol; Xanthotoxol | HMDB13679; HMDB29457 | M+H | - | - |

| | | | | | | | | | | | |
|---|---|---|---|---|---|---|---|---|---|---|---|
| HILIC positive | 154.0479 | 71.7 | -0.08 (1.05) | 0.08 (0.94) | -0.17 (-0.33, 0) | 0.0441 | 0.6587 | 4-Hydroxyproline; N-Acetyl-L-alanine; Propionylglycine; 5-Aminolevulinic acid; L-Glutamic gamma-semialdehyde; 3-Hydroxy-L-proline; 4-Hydroxy-L-proline; 5-Amino-2-oxopentanoic acid; 4-Hydroxy-2-pyrrolidinecarboxylic acid; Trans-3-hydroxy-L-proline; cis-4-Hydroxy-D-proline | HMDB00725; HMDB00766; HMDB00783; HMDB01149; HMDB02104; HMDB02113; HMDB06055; HMDB06272; HMDB36576; HMDB59659; HMDB60460 | M+Na | - | - |
| C18 negative | 378.2404 | 254 | 0.08 (0.91) | -0.08 (1.08) | 0.16 (0, 0.32) | 0.0443 | 0.6197 | Sphingosine 1-phosphate; N-palmitoyl-phosphoethanolamine | HMDB00277; HMDB59630 | M-H | - | - |
| HILIC positive | 286.2376 | 244.4 | 0.08 (0.9) | -0.08 (1.09) | 0.16 (0, 0.32) | 0.0445 | 0.6587 | Myristoylglycine | HMDB13250 | M+H | - | - |
| C18 negative | 225.1652 | 203.5 | 0.08 (0.98) | -0.08 (1.01) | 0.16 (0, 0.32) | 0.0446 | 0.6197 | (8E15E)-1815-Heptadecatriene-1113-diyne | HMDB31041 | M-H | - | - |
| C18 negative | 149.0607 | 25.1 | -0.08 (1.04) | 0.08 (0.95) | -0.17 (-0.33, 0) | 0.0447 | 0.6197 | Hydrocinnamic acid; 4-Ethylbenzoic acid; 3-Methylphenylacetic acid; 34-Dimethylbenzoic acid; 4-Coumaryl alcohol; 2-Phenylpropionate; 2-Methoxy-4-vinylphenol; Benzyl acetate; o-Tolyl acetate; p-Tolyl acetate; 2-Methoxyacetophenone; 4-Methoxyacetophenone; 2-Hydroxy-5-methylacetophenone; Methyl phenylacetate; Ethyl benzoate; 4-Ethoxybenzaldehyde; 1-Phenylethyl formate; 2-Ethyl-4-(2-furanyl)-2-propenal; 2-Phenylethyl formate; 2-(2-Furanyl)-3-methyl-2-butenal; 4-Hydroxy-3-methylacetophenone | HMDB00764; HMDB02097; HMDB02222; HMDB02237; HMDB03654; HMDB11743; HMDB13744; HMDB31310; HMDB32073; HMDB32075; HMDB32569; HMDB32570; HMDB32592; HMDB32617; HMDB33967; HMDB33970; HMDB37715; HMDB38355; HMDB40428; HMDB40925; HMDB59824 | M-H | - | - |
| HILIC positive | 296.2298 | 27.4 | -0.08 (1.01) | 0.08 (0.98) | -0.16 (-0.32, 0) | 0.0447 | 0.6587 | 13-OxoODE; 9-OxoODE; A-12(13)-EpODE; 13-HOTE; 15(16)-EpODE; 9(10)-EpODE; 9-HOTE; 17-Hydroxylinolenic acid; 10-Oxo-11-octadecen-13-olide; Squamostanal A; 1516-Epoxy-912-octadecadienoic acid; (2E4Z8E)-Colneleic acid; 1213-Epoxy-915-octadecadienoic acid; 2-Hydroxylinolenic acid; (9S10E12Z15Z)-9-Hydroxy-101215-octadecatrienoic acid; (9Z12Z14E)-16-Hydroxy-91214-octadecatrienoic acid; Sterebin D; 34-Dimethyl-5-pentyl-2-furanheptanoic acid | HMDB04668; HMDB04669; HMDB10200; HMDB10203; HMDB10206; HMDB10220; HMDB10224; HMDB11108; HMDB29786; HMDB29969; HMDB30950; HMDB30995; HMDB31088; HMDB31103; HMDB31934; HMDB34586; HMDB36832; HMDB39603 | M+H_[+1] | - | - |
| C18 negative | 431.317 | 218.6 | -0.09 (1.04) | 0.09 (0.95) | -0.17 (-0.34, 0) | 0.0451 | 0.6197 | 24-Hydroxycalcitriol; 3 beta7 alpha-Dihydroxy-5-cholestenoate; Neochlorogenin; Rockogenin; 23S2526-Trihydroxyvitamin D3 | HMDB06228; HMDB12454; HMDB30702; HMDB33769; HMDB60134 | M-H | - | - |
| HILIC positive | 395.1176 | 100.8 | 0.08 (0.87) | -0.08 (1.11) | 0.15 (0, 0.3) | 0.0453 | 0.6587 | Garcimangosone D | HMDB38066 | M+H_[+2] | - | - |
| HILIC positive | 150.0761 | 77.6 | -0.08 (0.98) | 0.08 (1.01) | -0.15 (-0.3, 0) | 0.0454 | 0.6587 | L-Glutamine; Ureidoisobutyric acid; D-Glutamine; Alanylglycine; Alanyl-Glycine | HMDB00641; HMDB02031; HMDB03423; HMDB06899; HMDB28687 | M+H_[+3] | - | - |
| C18 negative | 191.0197 | 20.5 | 0.08 (0.79) | -0.08 (1.17) | 0.16 (0, 0.32) | 0.0454 | 0.6197 | Citric acid; Isocitric acid; D-threo-Isocitric acid; Diketogulonic acid; 23-Diketo-L-gulonate; (1R2R)-Isocitric acid; D-Glucaro-14-lactone | HMDB00094; HMDB00193; HMDB01874; HMDB05971; HMDB06511; HMDB33717; HMDB41862 | M-H | Citric acid | M-H |
| C18 negative | 431.2438 | 69.2 | 0.08 (0.87) | -0.08 (1.11) | 0.16 (0, 0.31) | 0.0456 | 0.6197 | Erinacine E; Colupdox a; Erinacine A; Erinacine B | HMDB29656; HMDB39597; HMDB40291; HMDB40390 | M-H | - | - |
| C18 negative | 961.7307 | 41.7 | -0.09 (1.06) | 0.09 (0.93) | -0.17 (-0.34, 0) | 0.0457 | 0.6197 | TG(18:2(9Z12Z)20:5(5Z8Z11Z14Z17Z)22:6(4Z7Z10Z13Z16Z19Z)); TG(18:2(9Z12Z)22:6(4Z7Z10Z13Z16Z19Z)20:5(5Z8Z11Z14Z17Z)); TG(20:2n620:5(5Z8Z11Z14Z17Z)22:6(4Z7Z10Z13Z16Z19Z)); TG(20:2n622:6(4Z7Z10Z13Z16Z19Z)20:5(5Z8Z11Z14Z17Z)); TG(20:5(5Z8Z11Z14Z17Z)20:2n622:6(4Z7Z10Z13Z16Z19Z)) | HMDB52799; HMDB52827; HMDB53613; HMDB53637; HMDB55755 | M-H | - | - |
| HILIC positive | 316.1754 | 236.9 | 0.09 (0.9) | -0.09 (1.09) | 0.18 (0, 0.35) | 0.0460 | 0.6587 | Alizapride | HMDB15494 | M+H | - | - |
| HILIC positive | 378.7914 | 26.1 | -0.09 (1.11) | 0.09 (0.87) | -0.17 (-0.34, 0) | 0.0461 | 0.6587 | ()-Naled | HMDB31806 | M+H | - | - |
| HILIC positive | 152.0706 | 33.8 | -0.08 (1.06) | 0.08 (0.93) | -0.16 (-0.32, 0) | 0.0466 | 0.6587 | Acetaminophen; 2-Phenylglycine; Dopamine quinone; Leukoaminochrome; Methyl 2-aminobenzoate; 2-(Methylamino)benzoic acid; 3alpha477alpha-Tetrahydro-1H-isoindole-13(2H)-dione; Ethyl nicotinate; 2-Amino-3-methylbenzoate | HMDB01859; HMDB02210; HMDB12219; HMDB12992; HMDB29703; HMDB32609; HMDB33903; HMDB59846; HMDB60680 | M+H | - | - |
| C18 negative | 209.0858 | 147.4 | 0.09 (0.98) | -0.09 (1.01) | 0.17 (0, 0.34) | 0.0467 | 0.6245 | 2-Ethyl-1-hexanol sulfate | HMDB33373 | M-H | - | - |
| HILIC positive | 243.1079 | 182.6 | -0.08 (1.02) | 0.08 (0.97) | -0.15 (-0.3, 0) | 0.0468 | 0.6587 | Histidinyl-Serine; Serinyl-Histidine; D-erythro-D-galacto-octitol; (SS)-Nt-Histidinylalanine; (SS)-Np-Histidinylalanine | HMDB28894; HMDB29041; HMDB29953; HMDB40248; HMDB40249 | M+H | - | - |
| HILIC positive | 223.1692 | 27.2 | -0.08 (1.04) | 0.08 (0.96) | -0.16 (-0.32, 0) | 0.0468 | 0.6587 | Rishitin; Kobusone; Isokobusone; (10betaH11xi)-11-Hydroxy-13-nor-6-eremophilen-8-one; Acetaldehyde butyl phenethyl acetal; 26-Di-tert-butyl-14-benzenediol | HMDB35593; HMDB36790; HMDB36791; HMDB37605; HMDB37820; HMDB40178 | M+H | - | - |
| HILIC positive | 142.9837 | 265.6 | 0.07 (0.79) | -0.07 (1.17) | 0.14 (0, 0.29) | 0.0471 | 0.6587 | Mesna | HMDB03745 | M+H | - | - |

| | | | | | | | | | | | | |
|---|---|---|---|---|---|---|---|---|---|---|---|---|
| HILIC positive | 283.061 | 124.6 | 0.08 (0.89) | -0.08 (1.1) | 0.15 (0, 0.3) | 0.0475 | 0.6587 | 3-O-Methylcoumestrol; 9-O-Methylcoumestrol; Cochliophilin A; Pseudobaptigenin | HMDB29584; HMDB33588; HMDB34445; HMDB36616 | M+H | - | - |
| C18 negative | 167.0342 | 21.3 | -0.08 (1.16) | 0.08 (0.81) | -0.17 (-0.33, 0) | 0.0481 | 0.6248 | Homogentisic acid; Vanillic acid; 3-Hydroxymandelic acid; p-Hydroxymandelic acid; 34-Dihydroxybenzeneacetic acid; 5-Methoxysalicylic acid; 34-Dihydroxymandelaldehyde; 246-Trihydroxyacetophenone; 26-Dimethoxy-14-benzoquinone; 3-Acetyl-4-hydroxy-6-methyl-2H-pyran-2-one; Thermophillin; 3-Methoxysalicylic acid; Isovanillic acid | HMDB00130; HMDB00484; HMDB00750; HMDB00822; HMDB01336; HMDB01868; HMDB06242; HMDB29644; HMDB29673; HMDB33129; HMDB34335; HMDB59713; HMDB60003 | M-H | Homogent isate | M-H |
| C18 negative | 144.0455 | 247.5 | 0.08 (0.95) | -0.08 (1.04) | 0.17 (0, 0.34) | 0.0481 | 0.6248 | 1H-Indole-3-carboxaldehyde | HMDB29737 | M-H | - | - |
| C18 negative | 163.1128 | 196 | -0.09 (1.01) | 0.09 (0.98) | -0.17 (-0.34, 0) | 0.0482 | 0.6248 | 4-(1133-Tetramethylbutyl)-phenol; 1-(1-Methoxy-1-methylethyl)-4-methylbenzene; Benzyl butyl ether; trans-Jasmone; 5-Phenyl-1-pentanol; 1-Phenyl-2-pentanol; 2-Methyl-4-phenyl-2-butanol; 2-Butyl-4-methylphenol; (Z)-3-Methyl-2-(2-pentenyl)-2-cyclopenten-1-one; Thymol methyl ether; Norbicycloekasantalal; Santalone; Norecasantalal; 2-Methyl-3-(2-pentenyl)-2-cyclopenten-1-one | HMDB13825; HMDB29653; HMDB31311; HMDB31454; HMDB31624; HMDB31625; HMDB31866; HMDB32060; HMDB35601; HMDB35989; HMDB36719; HMDB36814; HMDB36816; HMDB37294 | M-H | - | - |
| C18 negative | 624.3416 | 38 | -0.07 (1.01) | 0.07 (0.99) | -0.13 (-0.26, 0) | 0.0484 | 0.6248 | Glycochenodeoxycholic acid 3-glucuronide | HMDB02579 | M-H | - | - |
| C18 negative | 315.1449 | 294.2 | 0.08 (0.99) | -0.08 (1.01) | 0.16 (0, 0.31) | 0.0489 | 0.6248 | 10-Hydroxy-8-nor-2-fenchanone glucoside; Glucosyl (E)-26-Dimethyl-25-heptadienoate | HMDB33643; HMDB35136 | M-H | - | - |
| C18 negative | 306.9555 | 27.3 | -0.07 (1.19) | 0.07 (0.76) | -0.15 (-0.3, 0) | 0.0491 | 0.6248 | Alphachloralose | HMDB31727 | M-H | - | - |
| C18 negative | 446.3356 | 235.7 | -0.08 (1.09) | 0.08 (0.89) | -0.16 (-0.31, 0) | 0.0491 | 0.6248 | 13-Carboxy-gamma-tocopherol; (3beta5alpha6beta7alpha22E24R)-Ergosta-822-diene-3567-tetrol; (3beta5alpha6alpha9alpha22E24R)-Ergosta-722-diene-3569-tetrol; 359-Trihydroxyergost-7-en-6-one; Secasterone; 3-Dehydroteasterone | HMDB12557; HMDB32107; HMDB32123; HMDB32875; HMDB40999; HMDB41527 | M-H_[-1] | - | - |
| C18 negative | 337.0199 | 18.9 | -0.03 (1.01) | 0.03 (0.99) | -0.06 (-0.12, 0) | 0.0493 | 0.6248 | Famotidine | HMDB01919 | M-H_[-1] | - | - |
| HILIC positive | 260.0531 | 51.3 | -0.07 (0.98) | 0.07 (1.01) | -0.14 (-0.28, 0) | 0.0493 | 0.6587 | Glucosamine-1P; Glucosamine 6-phosphate; Aminofructose 6-phosphate | HMDB01109; HMDB01254; HMDB60436 | M+H | - | - |
| HILIC positive | 211.1433 | 117.1 | 0.08 (0.84) | -0.08 (1.13) | 0.16 (0, 0.33) | 0.0494 | 0.6587 | LL-Cyclo(leucylprolyl) | HMDB34276 | M+H | - | - |
| C18 negative | 157.1234 | 73.2 | -0.08 (1.05) | 0.08 (0.94) | -0.16 (-0.31, 0) | 0.0494 | 0.6248 | Oenanthic ether; Pelargonic acid; xi-2-Ethylheptanoic acid; Methyl caprylate; Heptyl acetate; 4-Methyloctanoic acid; Butyl 3-methylbutanoate; 3-Heptyl acetate; 5-Methylhexyl acetate; 3-Octyl formate; Pentyl butanoate; Propyl hexanoate; Hexyl propionate; 3-Methylbutyl 2-methylpropanoate; 1-Hydroxy-3-nonanone; 3-(Hydroxymethyl)-2-octanone; 4-Methyl-2-pentyl-13-dioxolane; 2-Methylpropyl 3-methylbutanoate; Octyl formate; Butyl 2-methylbutanoate; 1-Methylbutyl butanoate; Isoamyl butyrate; Butyl pentanoate; Isopropyl hexanoate; Ethyl 5-methylhexanoate | HMDB00798; HMDB00847; HMDB30331; HMDB31291; HMDB31480; HMDB31557; HMDB32042; HMDB32309; HMDB32406; HMDB32454; HMDB34162; HMDB34165; HMDB34606; HMDB36234; HMDB37143; HMDB37164; HMDB37288; HMDB38040; HMDB38603; HMDB39216; HMDB39219; HMDB40221; HMDB40295; HMDB40430; HMDB59822 | M-H | - | - |
| C18 negative | 509.184 | 183 | 0.08 (0.84) | -0.08 (1.13) | 0.16 (0, 0.32) | 0.0498 | 0.6248 | Physalin B; Physalin C; 2527-Dihydro-47-didehydro-7-deoxyphysalin A; Isophysalin B | HMDB30128; HMDB34094; HMDB39696; HMDB39697 | M-H | - | - |
| HILIC positive | 193.0351 | 90.3 | -0.07 (1.1) | 0.07 (0.89) | -0.14 (-0.28, 0) | 0.0499 | 0.6587 | Citric acid; Isocitric acid; D-threo-Isocitric acid; Diketogulonic acid; 23-Diketo-L-gulonate; (1R2R)-Isocitric acid; D-Glucaro-14-lactone | HMDB00094; HMDB00193; HMDB01874; HMDB05971; HMDB06511; HMDB33717; HMDB41862 | M+H | - | - |
| HILIC positive | 103.548 | 93.4 | -0.08 (0.97) | 0.08 (1.03) | -0.16 (-0.32, 0) | 0.0500 | 0.6587 | Cycloserine; 3-Amino-2-oxazolidone | HMDB14405; HMDB60739 | M+H_[+1] | - | - |

SD: Standard Deviation; CI: Confidence Interval; FDR: False Discovery Rate.

**Supplemental Table 4.** Results of the Untargeted Metabolome-wide Association Study Comparing TB Resisters and Controls in the Brazilian Cohort (155 Pairs). Metabolites with P Value < 0.05 and Level 1 or 2 Confirmation are Shown.

| Platform | m/z | time | Resister Mean (SD) | Control Mean (SD) | Difference Resister vs. Control (95% CI) | P | FDR | Level 2 Confirmation | | | Level 1 Confirmation | |
|---|---|---|---|---|---|---|---|---|---|---|---|---|
| | | | | | | | | Name | HMDB ID | Adduct | Name | Adduct |
| C18 negative | 239.1652 | 147.7 | 0.21 (0.95) | -0.27 (1.24) | 0.48 (0.24, 0.72) | 0.0001 | 0.3700 | L-Menthyl acetoacetate | HMDB32369 | M-H | - | - |
| C18 negative | 183.1026 | 25.1 | -0.16 (1.1) | 0.29 (0.89) | -0.44 (-0.68, -0.2) | 0.0003 | 0.4015 | (1R)-Nepetalic acid; (S)-Oleuropeic acid; (4S8R)-89-Dihydroxy-p-menth-1(6)-en-2-one; 1-Acetylcyclohexyl acetate; (E)-10-Oxo-8-decenoic acid | HMDB36117; HMDB36998; HMDB39052; HMDB39710; HMDB40883 | M-H | - | - |
| HILIC positive | 809.5859 | 31.8 | 0.23 (0.96) | -0.1 (0.96) | 0.33 (0.15, 0.52) | 0.0005 | 0.7057 | PC(16:022:5(4Z7Z10Z13Z16Z)); PC(16:022:5(7Z10Z13Z16Z19Z)); PC(16:1(9Z)22:4(7Z10Z13Z16Z)); PC(18:020:5(5Z8Z11Z14Z17Z)); PC(18:1(11Z)20:4(5Z8Z11Z14Z)); PC(18:1(11Z)20:4(8Z11Z14Z17Z)); PC(18:1(9Z)20:4(5Z8Z11Z14Z)); PC(18:1(9Z)20:4(8Z11Z14Z17Z)); PC(18:2(9Z12Z)20:3(5Z8Z11Z)); PC(18:2(9Z12Z)20:3(8Z11Z14Z)); PC(18:3(6Z9Z12Z)20:2(11Z14Z)); PC(18:3(9Z12Z15Z)20:2(11Z14Z)); PC(18:4(6Z9Z12Z15Z)20:1(11Z)); PC(20:1(11Z)18:4(6Z9Z12Z15Z)); PC(20:2(11Z14Z)18:3(6Z9Z12Z)); PC(20:2(11Z14Z)18:3(9Z12Z15Z)); PC(20:3(5Z8Z11Z)18:2(9Z12Z)); PC(20:3(8Z11Z14Z)18:2(9Z12Z)); PC(20:4(5Z8Z11Z14Z)18:1(11Z)); PC(20:4(5Z8Z11Z14Z)18:1(9Z)); PC(20:4(8Z11Z14Z17Z)18:1(11Z)); PC(20:4(8Z11Z14Z17Z)18:1(9Z)); PC(20:5(5Z8Z11Z14Z17Z)18:0); PC(22:4(7Z10Z13Z16Z)16:1(9Z)); PC(22:5(4Z7Z10Z13Z16Z)16:0); PC(22:5(7Z10Z13Z16Z19Z)16:0) | HMDB07989; HMDB07990; HMDB08021; HMDB08050; HMDB08081; HMDB08082; HMDB08114; HMDB08115; HMDB08139; HMDB08146; HMDB08177; HMDB08210; HMDB08242; HMDB08306; HMDB08337; HMDB08338; HMDB08369; HMDB08402; HMDB08432; HMDB08433; HMDB08465; HMDB08466; HMDB08497; HMDB08627; HMDB08659; HMDB08692 | M+H_[+1] | - | - |
| HILIC positive | 808.5832 | 31.4 | 0.23 (0.95) | -0.09 (0.95) | 0.33 (0.14, 0.51) | 0.0005 | 0.7057 | PC(16:022:5(4Z7Z10Z13Z16Z)); PC(16:022:5(7Z10Z13Z16Z19Z)); PC(16:1(9Z)22:4(7Z10Z13Z16Z)); PC(18:020:5(5Z8Z11Z14Z17Z)); PC(18:1(11Z)20:4(5Z8Z11Z14Z)); PC(18:1(11Z)20:4(8Z11Z14Z17Z)); PC(18:1(9Z)20:4(5Z8Z11Z14Z)); PC(18:1(9Z)20:4(8Z11Z14Z17Z)); PC(18:2(9Z12Z)20:3(5Z8Z11Z)); PC(18:2(9Z12Z)20:3(8Z11Z14Z)); PC(18:3(6Z9Z12Z)20:2(11Z14Z)); PC(18:3(9Z12Z15Z)20:2(11Z14Z)); PC(18:4(6Z9Z12Z15Z)20:1(11Z)); PC(20:1(11Z)18:4(6Z9Z12Z15Z)); PC(20:2(11Z14Z)18:3(6Z9Z12Z)); PC(20:2(11Z14Z)18:3(9Z12Z15Z)); PC(20:3(5Z8Z11Z)18:2(9Z12Z)); PC(20:3(8Z11Z14Z)18:2(9Z12Z)); PC(20:4(5Z8Z11Z14Z)18:1(11Z)); PC(20:4(5Z8Z11Z14Z)18:1(9Z)); PC(20:4(8Z11Z14Z17Z)18:1(11Z)); PC(20:4(8Z11Z14Z17Z)18:1(9Z)); PC(20:5(5Z8Z11Z14Z17Z)18:0); PC(22:4(7Z10Z13Z16Z)16:1(9Z)); PC(22:5(4Z7Z10Z13Z16Z)16:0); PC(22:5(7Z10Z13Z16Z19Z)16:0) | HMDB07989; HMDB07990; HMDB08021; HMDB08050; HMDB08081; HMDB08082; HMDB08114; HMDB08115; HMDB08139; HMDB08146; HMDB08177; HMDB08210; HMDB08242; HMDB08306; HMDB08337; HMDB08338; HMDB08369; HMDB08402; HMDB08432; HMDB08433; HMDB08465; HMDB08466; HMDB08497; HMDB08627; HMDB08659; HMDB08692 | M+H | - | - |

| | | | | | | | | | | | | |
|---|---|---|---|---|---|---|---|---|---|---|---|---|
| HILIC positive | 463.3411 | 27.2 | -0.52 (0.99) | -0.14 (1.02) | -0.38 (-0.59, -0.16) | 0.0007 | 0.7057 | Dolichosterone; Polyporusterone F | HMDB34336; HMDB38499 | M+H | - | - |
| C18 negative | 158.0096 | 294.2 | 0.2 (0.9) | -0.18 (1.1) | 0.39 (0.16, 0.61) | 0.0008 | 0.4015 | (E)-4-Isothiocyanato-1-(methylthio)-1-butene; (E)-Raphanusanin | HMDB31576; HMDB41083 | M-H | - | - |
| C18 negative | 325.311 | 294.4 | 0.11 (0.6) | -0.3 (1.37) | 0.4 (0.17, 0.63) | 0.0008 | 0.4015 | Homophytanic acid; Heneicosanoic acid; 2371115-Pentamethylhexadecanoic acid; 6-Hydroxy-8-heneicosanone; 8-Hydroxy-6-heneicosanone; 4-Hydroxy-6-heneicosanone; 6-Hydroxy-4-heneicosanone | HMDB02337; HMDB02345; HMDB31319; HMDB35668; HMDB35669; HMDB35670; HMDB35671 | M-H | - | - |
| C18 negative | 412.3066 | 180.3 | -0.23 (0.99) | 0.14 (1) | -0.37 (-0.58, -0.15) | 0.0010 | 0.4015 | 3-Hydroxy-9-hexadecenoylcarnitine | HMDB13333 | M-H | - | - |
| HILIC positive | 279.2315 | 26.9 | -0.29 (1.18) | 0.07 (0.87) | -0.37 (-0.58, -0.15) | 0.0010 | 0.7057 | Alpha-Linolenic acid; Gamma-Linolenic acid; Calendic acid; Punicic acid; Linolenelaidic acid | HMDB01388; HMDB03073; HMDB30962; HMDB30963; HMDB30964 | M+H | - | - |
| HILIC positive | 597.5086 | 26.8 | -0.25 (1.07) | 0.07 (0.76) | -0.32 (-0.51, -0.13) | 0.0013 | 0.7057 | 3-Oxooctadecanoic acid; 9-Oxooctadecanoic acid; 10-Oxooctadecanoic acid; 11-Oxooctadecanoic acid; 5-Hexyltetrahydro-2-furanoctanoic acid; 5-Oxooctadecanoic acid; Ricinoleic acid; Glycerol triundecanoate | HMDB10736; HMDB30979; HMDB30980; HMDB30981; HMDB31127; HMDB34074; HMDB34297; HMDB31089 | 2M+H; M+H | - | - |
| C18 negative | 203.1804 | 171.7 | 0.23 (0.73) | -0.07 (1) | 0.3 (0.12, 0.48) | 0.0014 | 0.4342 | delta-Amorphene; (-)-35-Cadinadiene; alpha-Santalene; Sesquisabinene; beta-Selinene; ()-1(10)4-Cadinadiene; Aristolene; 2710-Bisabolatriene; Sesquithujene; Daucene; Valencene; (2Z4E6E)-24610-Farnesatetraene; alpha-Selinene; (-)-beta-Elemene; beta-Farnesene; beta-Bisabolene; alpha-Guaiene; alpha-Cyperene; alpha-Farnesene; (3E6E)-alpha-Farnesene; (Z)-2610-Bisabolatriene; beta-Sesquiphellandrene; Zingiberene; (R)-2511-Bisabolatriene; beta-Santalene; alpha-Cubebene; 1-Epi-alpha-gurjunene; (-)-Aromadendrene; (1beta4alpha5beta6alpha7alpha)-9-Aromadendrene; (4alpha5beta6alpha7alpha10alpha)-1-Aromadendrene; alpha-Bulnesene; (4alpha10alpha)-1(5)6-Guaiadiene; alpha-Humulene; Germacrene B; beta-Bulnesene; alpha-Bergamotene; Cascarilladiene; beta-Neoclovene; beta-Patchoulene; delta-Patchoulene; beta-Caryophyllene; Cycloseychellene; gamma1-Cadinene; 1-Epi-bicyclosesquiphellandrene; 37(11)-Eudesmadiene; (-)-Tamariscene; ()-1(9)10-Pacifigorgiadiene; (-)-1(6)10-Pacifigorgiadiene; ()-alpha-Muurolene; gamma-Selinene; beta-Panasinsene; alpha-Panasinsene; Bicyclogermacrene; epsilon-Muurolene; beta-Bourbonene; beta-Guaiene; Bicycloelemene; gamma-Himachalene; omega-Cadinene; alpha-Cadinene; gamma-Muurolene; gamma-Cadinene; beta-Humulene; beta-Cadinene; Benzylalcohol alpha-isobutyl-246-trimethyl(1-mestyl-3-methyl-1-butanol); alpha-Cedrene; alpha-Alaskene | HMDB30644; HMDB30673; HMDB34939; HMDB34943; HMDB34997; HMDB35084; HMDB35146; HMDB35161; HMDB35198; HMDB35361; HMDB35688; HMDB35787; HMDB35810; HMDB35816; HMDB35913; HMDB35992; HMDB36003; HMDB36004; HMDB36065; HMDB36066; HMDB36154; HMDB36163; HMDB36164; HMDB36200; HMDB36364; HMDB36413; HMDB36417; HMDB36418; HMDB36419; HMDB36428; HMDB36444; HMDB36450; HMDB36467; HMDB36575; HMDB36644; HMDB36678; HMDB36682; HMDB36731; HMDB36736; HMDB36737; HMDB36792; HMDB36799; HMDB37029; HMDB37030; HMDB37060; HMDB37676; HMDB37677; HMDB37678; HMDB37784; HMDB38118; HMDB38124; HMDB38135; HMDB38137; HMDB38139; HMDB38155; HMDB38157; HMDB38166; HMDB38168; HMDB38203; HMDB38205; HMDB38206; HMDB38207; HMDB38214; HMDB41576; HMDB59689; HMDB59695; HMDB59818 | M-H | - | - |
| HILIC positive | 579.4969 | 26.9 | -0.29 (1.18) | 0.05 (0.91) | -0.34 (-0.55, -0.13) | 0.0014 | 0.7057 | DG(15:018:2(9Z12Z)0:0); DG(18:2(9Z12Z)15:00:0); Glycerol triundecanoate; DG(14:00:018:2n6); DG(14:00:020:2n6) | HMDB07074; HMDB07242; HMDB31089; HMDB55967; HMDB55969 | M+H; M+H-H2O | - | - |
| HILIC positive | 224.1281 | 184.7 | 0.21 (0.56) | -0.12 (1.16) | 0.33 (0.13, 0.54) | 0.0015 | 0.7057 | Cerulenin | HMDB15168 | M+H | - | - |
| C18 negative | 257.1032 | 293.8 | 0.2 (0.89) | -0.13 (0.93) | 0.33 (0.13, 0.53) | 0.0016 | 0.4654 | 11-(Tetrahydro-6a-hydroxy-23a5-trimethylfuro23-d-13-dioxole-25-diyl)bis-ethanone | HMDB32527 | M-H | - | - |
| HILIC positive | 559.4711 | 26.8 | -0.25 (1.02) | 0.03 (0.84) | -0.28 (-0.45, -0.11) | 0.0017 | 0.7057 | DG(15:018:3(6Z9Z12Z)0:0); DG(15:018:3(9Z12Z15Z)0:0); DG(18:3(6Z9Z12Z)15:00:0); DG(18:3(9Z12Z15Z)15:00:0); DG(15:00:018:3n6); DG(15:00:018:3n3); DG(14:1n50:018:2n6); DG(14:1n50:020:2n6) | HMDB07075; HMDB07076; HMDB07271; HMDB07300; HMDB55996; HMDB56003; HMDB56142; HMDB56144 | M+H-H2O | - | - |
| C18 negative | 605.3332 | 139.1 | -0.3 (0.97) | 0.06 (1) | -0.37 (-0.6, -0.14) | 0.0018 | 0.5034 | (23S)-2325-dihdroxy-24-oxovitamine D3 23-(beta-glucuronide) | HMDB10361 | M-H | - | - |
| C18 negative | 198.158 | 155.8 | 0.12 (0.88) | -0.23 (1.36) | 0.35 (0.13, 0.57) | 0.0020 | 0.5100 | 5-Dodecenoic acid; trans-Dodec-2-enoic acid; Methyl 10-undecenoate; Allyl nonanoate; xi-Dihydro-5-octyl-2(3H)-furanone; cis-4-Decenyl acetate; 26-Dimethyl-5-heptenal propyleneglycol acetal; 11-Dodecenoic acid; cis-3-Hexenyl hexanoate; ()-Citronellyl acetate; Citronellyl acetate; 55- | HMDB00529; HMDB10729; HMDB29585; HMDB29763; HMDB31683; HMDB32214; HMDB32235; HMDB32248; HMDB33378; HMDB34160; | M-H_[-1] | - | - |

| | | | | | | | | | | | | |
|---|---|---|---|---|---|---|---|---|---|---|---|---|
| | | | | | | | | Dibutyl-45-dihydro-2(3H)furanone; Rhodinyl acetate; Methyl 9-undecenoate; Ethyl 2-decenoate; 1-Ethenylhexyl butanoate; xi-5-Dodecanolide; 3-Heptyldihydro-5-methyl-2(3H)-furanone; 2-Octenyl butyrate; Hexyl 2E-hexenoate; 7-Hexyl-2-oxepanone; 2-Hexenyl hexanoate; Ethyl 4-decenoate; Hexyl 2-methyl-3-pentenoate; Hexyl 2-methyl-4-pentenoate; 11-Dimethoxy-37-dimethyl-26-octadiene; ()-Menthyl acetate; (37-Dimethyl-6-octenyl)oxyacetaldehyde | HMDB34844; HMDB36182; HMDB37186; HMDB37305; HMDB37329; HMDB37498; HMDB37742; HMDB37813; HMDB38081; HMDB38269; HMDB38895; HMDB38924; HMDB39220; HMDB40158; HMDB40163; HMDB40361; HMDB41264; HMDB41449 | | | |
| HILIC positive | 592.4932 | 26.7 | -0.22 (1.18) | 0.12 (0.76) | -0.34 (-0.56, -0.13) | 0.0020 | 0.7057 | DG(15:018:4(6Z9Z12Z15Z)0:0); DG(18:4(6Z9Z12Z15Z)15:00:0); DG(15:00:018:4n3) | HMDB07077; HMDB07329; HMDB56004 | M+NH4 | - | - |
| HILIC positive | 675.5427 | 35.1 | 0.2 (0.99) | -0.14 (1.02) | 0.34 (0.12, 0.55) | 0.0022 | 0.7079 | SM(d18:114:0) | HMDB12097 | M+H | - | - |
| HILIC positive | 181.1588 | 110.9 | 0.15 (0.59) | -0.19 (1.24) | 0.34 (0.12, 0.56) | 0.0023 | 0.7246 | Homodihydrojasmone; 2-trans-6-cis-Dodecadienal; cis-Quinceoxepane; Tricycloekasantalol; (2E4E)-24-Dodecadienal | HMDB31181; HMDB32531; HMDB38108; HMDB39339; HMDB39590 | M+H | - | - |
| HILIC positive | 241.1546 | 56.1 | -0.1 (0.81) | 0.12 (0.79) | -0.22 (-0.36, -0.08) | 0.0027 | 0.7296 | Pirbuterol | HMDB15407 | M+H | - | - |
| HILIC positive | 714.5445 | 208.3 | 0.17 (0.81) | -0.16 (1.11) | 0.33 (0.12, 0.55) | 0.0029 | 0.7296 | PC(14:1(9Z)P-18:1(11Z)); PC(14:1(9Z)P-18:1(9Z)); PC(P-18:1(11Z)14:1(9Z)); PC(P-18:1(9Z)14:1(9Z)) | HMDB07930; HMDB07931; HMDB11270; HMDB11303 | M+H | - | - |
| HILIC positive | 193.1224 | 107.2 | 0.12 (0.44) | -0.16 (1.14) | 0.28 (0.1, 0.46) | 0.0029 | 0.7296 | 2-Methyl-1-phenyl-2-propanyl acetate; 4-Phenyl-2-butyl acetate; Ethyl 4-phenylbutanoate; Benzyl 3-methylbutanoate; 3-Methylbutyl benzoate; 2-Methylpropyl phenylacetate; 2-Phenylethyl butanoate; 2-Phenylethyl 2-methylpropanoate; 5-Isopropyl-2-methylphenol acetate; 3-Phenylpropyl propanoate; 111213-Trinor-1(10)-spirovetivene-27-dione; 4-Methyl-2-(1-phenylethyl)-13-dioxolane 9CI; 4-Methylphenyl 3-methylbutanoate; 1-Phenylethyl isobutyrate; 1-Phenylethyl butyrate; 1-Ethoxy-2-methoxy-4-(1-propenyl)benzene; 2-(2-Hydroxy-4-methylphenyl)-3-pentanone; 2-Ethoxy-1-methoxy-4-(1-propenyl)benzene; 111213-Trinor-135-bisabolatrien-10-oic acid; 2-Benzyl-45-dimethyl-13-dioxolane; Butyl phenylacetate | HMDB31571; HMDB31614; HMDB31618; HMDB32043; HMDB33380; HMDB35010; HMDB35014; HMDB35015; HMDB35447; HMDB36388; HMDB36734; HMDB37165; HMDB37709; HMDB37716; HMDB37717; HMDB37792; HMDB38180; HMDB39496; HMDB39643; HMDB39841; HMDB40427 | M+H | - | - |
| C18 negative | 155.0713 | 217.2 | -0.16 (1.05) | 0.15 (0.88) | -0.31 (-0.51, -0.11) | 0.0032 | 0.5285 | 8-Hydroxy-56-octadienoic acid; 25-Dimethyl-4-ethoxy-3(2H)-furanone | HMDB31101; HMDB32232 | M-H | - | - |
| HILIC positive | 411.2506 | 26.3 | 0.16 (0.9) | -0.23 (1.45) | 0.39 (0.13, 0.64) | 0.0033 | 0.7296 | LPA(0:016:0); LPA(16:00:0) | HMDB07849; HMDB07853 | M+H | - | - |
| HILIC positive | 403.3576 | 26.7 | -0.11 (0.99) | 0.19 (0.86) | -0.3 (-0.49, -0.1) | 0.0034 | 0.7296 | 24-Hydroxycholesterol; 7a-Hydroxycholesterol; 27-Hydroxycholesterol; Delta-Tocopherol; 22b-Hydroxycholesterol; 7b-Hydroxycholesterol; 25-Hydroxycholesterol; 20alpha-Hydroxycholesterol; 7a-Hydroxy-5b-cholestan-3-one; (24R)-Cholest-5-ene-3-beta24-diol ; 4b-Hydroxycholesterol; (3beta5alpha6alpha)-Cholest-8-ene-36-diol; eta-Tocopherol; 3-Hydroxy-1-phenyl-1-heneicosanone; 2-Methyl-6-phytylhydroquinone; 5-(12-Heneicosenyl)-13-benzenediol; 5-(14-Heneicosenyl)-13-benzenediol; 5-(16-Heneicosenyl)-13-benzenediol; Cholest-5-ene-3-beta7-alpha-diol | HMDB01419; HMDB01496; HMDB02103; HMDB02902; HMDB04035; HMDB06119; HMDB06247; HMDB06283; HMDB06892; HMDB11643; HMDB13643; HMDB29320; HMDB34254; HMDB35681; HMDB38959; HMDB39868; HMDB39869; HMDB39870; HMDB59592 | M+H | - | - |
| HILIC positive | 288.1665 | 86.1 | -0.04 (0.98) | 0.23 (0.74) | -0.26 (-0.44, -0.09) | 0.0035 | 0.7296 | Arginyl-Hydroxyproline; Hydroxyprolyl-Arginine | HMDB28710; HMDB28857 | M+H | - | - |
| C18 negative | 463.343 | 215.7 | -0.26 (1.25) | 0.09 (0.77) | -0.34 (-0.57, -0.11) | 0.0036 | 0.5310 | Trihydroxycoprostanoic acid; 2-Deoxybrassinolide | HMDB02163; HMDB41135 | M-H | - | - |
| C18 negative | 385.2594 | 211.8 | -0.17 (1) | 0.16 (0.94) | -0.33 (-0.55, -0.11) | 0.0036 | 0.5310 | Glycerol trihexanoate; Mangalkanyl glucoside; Cryptomeridiol 11-rhamnoside | HMDB31125; HMDB36015; HMDB38018 | M-H | - | - |
| C18 negative | 435.3479 | 282.2 | -0.26 (1.22) | 0.1 (0.91) | -0.35 (-0.59, -0.12) | 0.0037 | 0.5310 | 5a-Cholestane-3a7a12a25-tetrol; 5b-Cholestane-3a7a12a25-tetrol; Cholestane-371225-tetrol; 27-Deoxy-5b-cyprinol; 5b-Cholestane-3a7a12a23-Tetrol; 27-Deoxy-5alpha-cyprinol | HMDB00520; HMDB00524; HMDB00655; HMDB01231; HMDB01968; HMDB29851 | M-H | - | - |
| HILIC positive | 277.2162 | 27.5 | -0.28 (1.2) | 0.06 (0.96) | -0.34 (-0.57, -0.11) | 0.0041 | 0.7373 | 19-Norandrosterone; 19-Nor-5-androstenediol; 19-Noretiocholanolone; Stearidonic acid; Phenethyl decanoate; 481215-Octadecatetraenoic acid; (EE)-1113-Octadecadien-9-ynoic acid | HMDB02697; HMDB04590; HMDB05886; HMDB06547; HMDB32464; HMDB32672; HMDB34382 | M+H | - | - |
| C18 negative | 347.1501 | 139.8 | -0.03 (0.96) | 0.26 (0.88) | -0.29 (-0.49, -0.09) | 0.0044 | 0.5664 | Gibberellin A60; Gibberellin A58; Gibberellin A63; Gibberellin A54; Gibberellin A67; Gibberellin A77; 8-Angeloylegelolide; Gibberellin A81; Triphasiol; Gibberellin A90; 3-Epigibberellin A1 | HMDB34988; HMDB35043; HMDB35044; HMDB35048; HMDB35049; HMDB37004; HMDB37773; HMDB38080; HMDB39062; HMDB39622; HMDB40944 | M-H | - | - |
| C18 negative | 490.3621 | 219.8 | -0.42 (1.09) | -0.09 (1.05) | -0.34 (-0.57, -0.11) | 0.0045 | 0.5664 | Barringtogenol C; Camelliagenin C; Ganoderiol H | HMDB34525; HMDB34532; HMDB37783 | M-H_[-1] | - | - |
| HILIC positive | 491.3725 | 27.4 | -0.35 (1.22) | 0.02 (0.93) | -0.36 (-0.61, -0.11) | 0.0045 | 0.7477 | Barringtogenol C; Camelliagenin C; Ganoderiol H | HMDB34525; HMDB34532; HMDB37783 | M+H | - | - |

| | | | | | | | | | | | | |
|---|---|---|---|---|---|---|---|---|---|---|---|---|
| HILIC positive | 854.5693 | 29.3 | 0.2 (0.53) | -0.07 (1.13) | 0.27 (0.08, 0.46) | 0.0050 | 0.7477 | PC(20:4(5Z8Z11Z14Z)22:6(4Z7Z10Z13Z16Z19Z)); PC(20:4(8Z11Z14Z17Z)22:6(4Z7Z10Z13Z16Z19Z)); PC(20:5(5Z8Z11Z14Z17Z)22:5(4Z7Z10Z13Z16Z)); PC(20:5(5Z8Z11Z14Z17Z)22:5(7Z10Z13Z16Z19Z)); PC(22:5(4Z7Z10Z13Z16Z)20:5(5Z8Z11Z14Z17Z)); PC(22:5(7Z10Z13Z16Z19Z)20:5(5Z8Z11Z14Z17Z)); PC(22:6(4Z7Z10Z13Z16Z19Z)20:4(5Z8Z11Z14Z)); PC(22:6(4Z7Z10Z13Z16Z19Z)20:4(8Z11Z14Z17Z)) | HMDB08452; HMDB08485; HMDB08516; HMDB08517; HMDB08675; HMDB08708; HMDB08739; HMDB08740 | M+H | - | - |
| C18 negative | 473.3633 | 250.6 | -0.22 (1.18) | 0.05 (0.76) | -0.28 (-0.47, -0.08) | 0.0051 | 0.5787 | Soyasapogenol A; Camelliagenin A; Priverogenin B; (3alphaOH20S24S)-319:2024-Diepoxydammarane-325-diol; Ganoderiol A; 2024-Epoxy-2526-dihydroxydammaran-3-one | HMDB34505; HMDB34528; HMDB34644; HMDB34683; HMDB35326; HMDB39692 | M-H | - | - |
| HILIC positive | 263.0874 | 297.1 | 0.63 (0.54) | 0.45 (0.74) | 0.18 (0.06, 0.31) | 0.0052 | 0.7477 | L-beta-aspartyl-L-glutamic acid ; Na-L-Glutamyl-L-aspartic acid; Ng-L-Glutamyl-L-aspartic acid | HMDB11164; HMDB30413; HMDB30419 | M+H | - | - |
| HILIC positive | 405.3721 | 26.8 | -0.52 (0.9) | -0.22 (0.99) | -0.31 (-0.52, -0.09) | 0.0053 | 0.7477 | 3a7a-Dihydroxy-5b-cholestane; 5-Heneicosyl-13-benzenediol; Lepidiumterpenoid | HMDB06893; HMDB31035; HMDB36844 | M+H | - | - |
| C18 negative | 283.1552 | 294.5 | 0.15 (0.79) | -0.2 (1.33) | 0.35 (0.11, 0.6) | 0.0054 | 0.5787 | Alpha-dihydroartemisinin; Dihydroartemisinin (DHA) | HMDB60593; HMDB61088 | M-H | - | - |
| C18 negative | 165.0408 | 288.3 | 0.16 (0.62) | -0.15 (1.19) | 0.31 (0.09, 0.52) | 0.0054 | 0.5787 | Arabinonic acid; Ribonic acid; S-Propyl 1-propanesulfinothioate; 2345-Tetrahydroxypentanoic acid; L-Lyxonate; L-Xylonate | HMDB00539; HMDB00867; HMDB34394; HMDB59750; HMDB60255; HMDB60256 | M-H | - | - |
| HILIC positive | 241.1065 | 214.1 | 0.17 (0.64) | -0.2 (1.48) | 0.37 (0.11, 0.63) | 0.0054 | 0.7477 | 3-(345-Trimethoxyphenyl)propanoic acid; Ethyl 345-trimethoxybenzoate; Isopropyl 3-(34-dihydroxyphenyl)-2-hydroxypropanoate; 3-Carboxy-4-methyl-5-propyl-2-furanpropionic acid | HMDB30254; HMDB38627; HMDB41756; HMDB61112 | M+H | - | - |
| C18 negative | 331.1912 | 142 | -0.27 (1.24) | 0.09 (0.99) | -0.36 (-0.61, -0.11) | 0.0054 | 0.5787 | Carnosic acid; 11b-Hydroxyprogesterone; 7-Carboxy-gamma-tocotrienol; Hulupone; Adhulupone; (1(10)E4E6a9b)-9-(2-Methylbutanoyloxy)-1(10)411(13)-germacratrien-126-olide; (1(10)E4E6a9b)-9-(3-Methylbutanoyloxy)-1(10)411(13)-germacratrien-126-olide; Petasitin; 6-Angeloylfuranofukinol; 4-Deoxycohumulone; ent-7alpha12beta-Dihydroxy-16-kauren-196beta-olide; ent-15-Kaurene-1719-dioic acid; 718-Dihydroxykaurenolide; (9Z11S16S)-1-Acetoxy-917-octadecadiene-1214-diyne-1116-diol | HMDB02358; HMDB04031; HMDB12851; HMDB30103; HMDB30136; HMDB31374; HMDB31375; HMDB35637; HMDB36146; HMDB36623; HMDB36700; HMDB36726; HMDB36761; HMDB38966 | M-H | - | - |
| C18 negative | 291.6261 | 187.9 | 0.18 (0.88) | -0.12 (1.05) | 0.29 (0.09, 0.5) | 0.0059 | 0.5787 | 1112-Dimethoxydihydrokawain; Coriandrone A; Coriandrone B | HMDB30767; HMDB37774; HMDB37775 | M-H_[-1] | - | - |
| C18 negative | 197.1546 | 156 | 0.13 (0.96) | -0.2 (1.25) | 0.33 (0.09, 0.56) | 0.0061 | 0.5787 | 5-Dodecenoic acid; trans-Dodec-2-enoic acid; Methyl 10-undecenoate; Allyl nonanoate; xi-Dihydro-5-octyl-2(3H)-furanone; cis-4-Decenyl acetate; 26-Dimethyl-5-heptenal propyleneglycol acetal; 11-Dodecenoic acid; cis-3-Hexenyl hexanoate; ()-Citronellyl acetate; Citronellyl acetate; 55-Dibutyl-45-dihydro-2(3H)furanone; Rhodinyl acetate; Methyl 9-undecenoate; Ethyl 2-decenoate; 1-Ethenylhexyl butanoate; xi-5-Dodecanolide; 3-Heptyldihydro-5-methyl-2(3H)-furanone; 2-Octenyl butyrate; Hexyl 2E-hexenoate; 7-Hexyl-2-oxepanone; 2-Hexenyl hexanoate; Ethyl 4-decenoate; Hexyl 2-methyl-3-pentenoate; Hexyl 2-methyl-4-pentenoate; 11-Dimethoxy-37-dimethyl-26-octadiene; ()-Menthyl acetate; (37-Dimethyl-6-octenyl)oxyacetaldehyde | HMDB00529; HMDB10729; HMDB29585; HMDB29763; HMDB31683; HMDB32214; HMDB32235; HMDB32248; HMDB33378; HMDB34160; HMDB34844; HMDB36182; HMDB37186; HMDB37305; HMDB37329; HMDB37498; HMDB37742; HMDB37813; HMDB38081; HMDB38269; HMDB38895; HMDB38924; HMDB39220; HMDB40158; HMDB40163; HMDB40361; HMDB41264; HMDB41449 | M-H | - | - |
| HILIC positive | 541.4613 | 26.4 | -0.31 (1.07) | -0.02 (0.98) | -0.29 (-0.49, -0.08) | 0.0062 | 0.7477 | Momordicilin | HMDB30896 | M+H | - | - |
| HILIC positive | 108.0808 | 31.4 | -0.34 (1.3) | 0.02 (1.21) | -0.36 (-0.62, -0.1) | 0.0063 | 0.7477 | 3-Ethylpyridine; 24-Dimethylpyridine; 26-Dimethylpyridine; Benzylamine; O-Toluidine | HMDB29734; HMDB32244; HMDB32972; HMDB33871; HMDB41965 | M+H | - | - |
| HILIC positive | 575.466 | 26.9 | -0.14 (1.1) | 0.1 (0.82) | -0.24 (-0.42, -0.07) | 0.0067 | 0.7477 | DG(15:018:4(6Z9Z12Z15Z)0:0); DG(18:4(6Z9Z12Z15Z)15:00:0); DG(15:00:018:4n3) | HMDB07077; HMDB07329; HMDB56004 | M+H | - | - |
| HILIC positive | 315.2528 | 25.9 | -0.01 (0.56) | 0.17 (0.62) | -0.19 (-0.32, -0.05) | 0.0068 | 0.7477 | Octadecanedioic acid; 910-DHOME; 1213-DHOME; (9xi10xi12xi)-910-Dihydroxy-12-octadecenoic acid; Dibutyl decanedioate | HMDB00782; HMDB04704; HMDB04705; HMDB31679; HMDB41220 | M+H | - | - |
| HILIC positive | 974.2879 | 54.3 | 0.02 (1.05) | 0.25 (0.6) | -0.23 (-0.39, -0.06) | 0.0068 | 0.7477 | 5-cis-8-cis-Tetradecadienoyl-CoA; (2S6R10R)-Trimethyl-hendecanoyl-CoA | HMDB02288; HMDB12471 | M+H | - | - |
| HILIC positive | 260.0531 | 51.3 | -0.06 (0.98) | 0.18 (1.02) | -0.24 (-0.42, -0.07) | 0.0069 | 0.7477 | Glucosamine-1P; Glucosamine 6-phosphate; Aminofructose 6-phosphate | HMDB01109; HMDB01254; HMDB60436 | M+H | - | - |
| HILIC positive | 451.3243 | 29.8 | -0.38 (1.05) | -0.05 (0.99) | -0.33 (-0.56, -0.09) | 0.0070 | 0.7477 | Deoxycholic acid glycine conjugate; Chenodeoxycholic acid glycine conjugate; Glycoursodeoxycholic acid; Chenodeoxyglycocholic acid | HMDB00631; HMDB00637; HMDB00708; HMDB06898 | M+H_[+1] | - | - |

| | | | | | | | | | | | | |
|---|---|---|---|---|---|---|---|---|---|---|---|---|
| HILIC positive | 450.3211 | 29.6 | -0.17 (1.05) | 0.12 (0.85) | -0.3 (-0.51, -0.08) | 0.0071 | 0.7477 | Deoxycholic acid glycine conjugate; Chenodeoxycholic acid glycine conjugate; Glycoursodeoxycholic acid; Chenodeoxyglycocholic acid | HMDB00631; HMDB00637; HMDB00708; HMDB06898 | M+H | - | - |
| HILIC positive | 251.2005 | 26.9 | -0.26 (0.97) | 0 (0.92) | -0.26 (-0.45, -0.07) | 0.0071 | 0.7477 | Norambreinolide; 3-Methyl-alpha-ionyl acetate; 22-Bis(2-methylpropoxy)ethylbenzene | HMDB35293; HMDB37631; HMDB37712 | M+H | - | - |
| C18 negative | 165.0556 | 160.3 | -0.15 (0.99) | 0.15 (0.84) | -0.29 (-0.51, -0.08) | 0.0072 | 0.5925 | 3-(3-Hydroxyphenyl)propanoic acid; D-Phenyllactic acid; L-3-Phenyllactic acid; Phenyllactic acid; 4-Methoxyphenylacetic acid; Desaminotyrosine; 3-Phenoxypropionic acid; Homovanillin; Ethyl vanillin; Ethyl salicylate; 34-Dihydroxyphenylacetone; Guaicyl acetate; 34-Dimethoxybenzaldehyde; Ethylparaben; Methyl 2-methoxybenzoate; Methyl 4-methoxybenzoate; 3-(2-Hydroxyphenyl)propanoic acid; Ipomeanine; 4-Methoxybenzyl formate; 3-Hydroxy-1-(4-hydroxyphenyl)-1-propanone; Ethyl 2-furanacrylate; 4-Hydroxyphenyl-2-propionic acid; 3-Methoxyphenylacetic acid | HMDB00375; HMDB00563; HMDB00748; HMDB00779; HMDB02072; HMDB02199; HMDB02229; HMDB05175; HMDB29665; HMDB29817; HMDB31132; HMDB32030; HMDB32138; HMDB32573; HMDB32605; HMDB32639; HMDB33752; HMDB34243; HMDB34993; HMDB40645; HMDB41611; HMDB41683; HMDB59969 | M-H | - | - |
| C18 negative | 515.3742 | 230 | -0.28 (1.07) | 0.04 (0.98) | -0.32 (-0.56, -0.09) | 0.0073 | 0.5937 | 22-Acetylpriverogenin B | HMDB34645 | M-H | - | - |
| HILIC positive | 313.2372 | 28.1 | -0.13 (1.06) | 0.17 (0.95) | -0.3 (-0.52, -0.08) | 0.0073 | 0.7477 | 13-L-Hydroperoxylinoleic acid; 8(R)-Hydroperoxylinoleic acid; 9(S)-HPODE; 1213-DiHODE; 1516-DiHODE; 910-DiHODE; ()-(E)-13-Hydroxy-10-oxo-11-octadecenoic acid | HMDB03871; HMDB04706; HMDB06940; HMDB10201; HMDB10208; HMDB10221; HMDB40900 | M+H | - | - |
| HILIC positive | 414.3 | 29.5 | -0.34 (1.12) | -0.02 (1.01) | -0.33 (-0.56, -0.09) | 0.0076 | 0.7502 | Deoxycholic acid glycine conjugate; Chenodeoxycholic acid glycine conjugate; Glycoursodeoxycholic acid; Chenodeoxyglycocholic acid | HMDB00631; HMDB00637; HMDB00708; HMDB06898 | M+H-2H2O | - | - |
| HILIC positive | 286.1434 | 28.7 | 0.13 (0.93) | -0.12 (0.96) | 0.25 (0.07, 0.43) | 0.0080 | 0.7502 | Morphine; Hydromorphone; Piperine; Erysopine; Secodemethylclausenamide; (-)-Morphine; Norcodeine; Dihydroisomorphine; norhydrocodone | HMDB14440; HMDB14472; HMDB29377; HMDB30256; HMDB32961; HMDB40987; HMDB60657; HMDB60929; HMDB61070 | M+H | - | - |
| HILIC positive | 146.1176 | 45.6 | -0.13 (1.02) | 0.12 (0.92) | -0.24 (-0.42, -0.06) | 0.0081 | 0.7502 | 3-Dehydroxycarnitine | HMDB06831 | M+H | Acetylcholine | M+H |
| C18 negative | 163.0612 | 25.8 | -0.04 (0.73) | 0.18 (0.73) | -0.22 (-0.38, -0.06) | 0.0084 | 0.5963 | L-Fucose; Rhamnose; 15-Anhydrosorbitol; Beta-D-Fucose; 3-Deoxyfructose; L-Rhamnulose; 2-Deoxygalactopyranose; D-Fucose; 2-O-Methyl-D-xylose; Alpha-L-fucose; beta-L-fucose; L-Fuculose | HMDB00174; HMDB00849; HMDB02712; HMDB03081; HMDB05876; HMDB10207; HMDB12327; HMDB29196; HMDB33821; HMDB59624; HMDB59625; HMDB60267 | M-H | - | - |
| C18 negative | 225.1652 | 203.5 | 0.19 (0.95) | -0.09 (1.01) | 0.28 (0.07, 0.49) | 0.0087 | 0.5963 | (8E15E)-1815-Heptadecatriene-1113-diyne | HMDB31041 | M-H | - | - |
| C18 negative | 491.193 | 169 | -0.11 (0.84) | 0.12 (0.57) | -0.22 (-0.38, -0.06) | 0.0087 | 0.5963 | Isolariciresinol 9-O-alpha-L-arabinofuranoside | HMDB32906 | M-H | - | - |
| C18 negative | 584.2597 | 186.5 | 0.15 (0.96) | -0.18 (1.29) | 0.33 (0.08, 0.58) | 0.0088 | 0.5963 | Bilirubin; 4E15Z-Bilirubin IXa | HMDB00054; HMDB00488 | M-H_[-1] | - | - |
| HILIC positive | 456.4044 | 25.8 | -0.17 (1.14) | 0.13 (0.86) | -0.3 (-0.52, -0.08) | 0.0088 | 0.7502 | Arachidyl carnitine | HMDB06460 | M+H | - | - |
| C18 negative | 423.275 | 90.1 | -0.29 (1.21) | 0.04 (1.09) | -0.33 (-0.57, -0.08) | 0.0089 | 0.5963 | 1b-Hydroxycholic acid; 3b4b7a12a-Tetrahydroxy-5b-cholanoic acid; 2b3a7a12a-Tetrahydroxy-5b-cholanoic acid; 3a7a12a19-Tetrahydroxy-5b-cholanoic acid; 3a6b7b12a-Tetrahydroxy-5b-cholanoic acid; 3a6b7b12b-Tetrahydroxy-5b-cholanoic acid; 3a6b7a12a-Tetrahydroxy-5b-cholanoic acid; 13712-Tetrahydroxycholan-24-oic acid; 3a4b7a12a-Tetrahydroxy-5b-cholanoic acid; 1b3a7b-Trihydroxy-5b-cholanoic acid; 3a7b21-Trihydroxy-5b-cholanoic acid | HMDB00307; HMDB00311; HMDB00316; HMDB00340; HMDB00367; HMDB00377; HMDB00399; HMDB00433; HMDB00437; HMDB13158; HMDB13192 | M-H | - | - |
| C18 negative | 157.0254 | 288.5 | 0.1 (0.72) | -0.11 (0.83) | 0.22 (0.05, 0.38) | 0.0091 | 0.5963 | 45-Dihydroorotic acid; L-Dihydroorotic acid | HMDB00528; HMDB03349 | M-H | - | - |
| HILIC positive | 958.3128 | 54.4 | -0.08 (0.66) | 0.09 (0.73) | -0.16 (-0.29, -0.04) | 0.0091 | 0.7502 | Pentaglutamyl folate | HMDB06487 | M+H | - | - |
| HILIC positive | 190.138 | 74.7 | 0.07 (0.86) | -0.17 (1.09) | 0.24 (0.06, 0.42) | 0.0095 | 0.7502 | Homo-L-arginine; L-Targinine | HMDB00670; HMDB29416 | M+H_[+1] | - | - |
| HILIC positive | 235.1321 | 26.9 | -0.15 (0.84) | 0.13 (0.99) | -0.27 (-0.47, -0.07) | 0.0095 | 0.7502 | (10S11S)-Pterosin C; Dehydrooreadone; (S)-Pterosin P; Pterosin N | HMDB30763; HMDB36046; HMDB36607; HMDB38218 | M+H | - | - |
| C18 negative | 291.2329 | 234.6 | 0.16 (0.97) | -0.1 (0.97) | 0.26 (0.06, 0.46) | 0.0096 | 0.5963 | 3b17b-Dihydroxyetiocholane; 3a17a-Dihydroxy-5b-androstane; 3b17a-Dihydroxy-5a-androstane; 5a-Androstane-3a17a-diol; 5a-Androstane-3b17b-diol; Androstanediol; Etiocholanediol; Dihydroandrosterone; Androstan-3alpha17beta-diol | HMDB00369; HMDB00383; HMDB00412; HMDB00458; HMDB00493; HMDB00495; HMDB00551; HMDB00554; HMDB60437 | M-H | - | - |
| C18 negative | 567.3176 | 128.6 | -0.28 (1.16) | 0.03 (1.05) | -0.3 (-0.53, -0.07) | 0.0100 | 0.6021 | (3a5b7a)-23-Carboxy-7-hydroxy-24-norcholan-3-yl-b-D-Glucopyranosiduronic acid; Deoxycholic acid 3-glucuronide | HMDB02430; HMDB02596 | M-H | - | - |
| C18 negative | 171.0414 | 19.6 | -0.29 (1.08) | -0.07 (0.99) | -0.22 (-0.39, -0.05) | 0.0101 | 0.6021 | Hydantoin-5-propionic acid; L-alpha-Amino-5-oxo-2(5H)-isoxazolepropanoic acid | HMDB01212; HMDB34322 | M-H | - | - |
| C18 negative | 477.3219 | 178.9 | -0.16 (1.08) | 0.1 (0.93) | -0.27 (-0.47, -0.06) | 0.0101 | 0.6021 | Dolicholide; Polyporusterone A | HMDB34086; HMDB38495 | M-H | - | - |

| | | | | | | | | | | | | |
|---|---|---|---|---|---|---|---|---|---|---|---|---|
| HILIC positive | 295.2267 | 27.9 | -0.17 (1.15) | 0.13 (0.91) | -0.3 (-0.53, -0.07) | 0.0103 | 0.7502 | 13-OxoODE; 9-OxoODE; A-12(13)-EpODE; 13-HOTE; 15(16)-EpODE; 9(10)-EpODE; 9-HOTE; 17-Hydroxylinolenic acid; 10-Oxo-11-octadecen-13-olide; Squamostanal A; 1516-Epoxy-912-octadecadienoic acid; (2E4Z8E)-Colneleic acid; 1213-Epoxy-915-octadecadienoic acid; 2-Hydroxylinolenic acid; (9S10E12Z15Z)-9-Hydroxy-101215-octadecatrienoic acid; (9Z12Z14E)-16-Hydroxy-91214-octadecatrienoic acid; Sterebin D; 34-Dimethyl-5-pentyl-2-furanheptanoic acid | HMDB04668; HMDB04669; HMDB10200; HMDB10203; HMDB10206; HMDB10220; HMDB10224; HMDB11108; HMDB29786; HMDB29969; HMDB30950; HMDB30995; HMDB31088; HMDB31103; HMDB31934; HMDB34586; HMDB36832; HMDB39603 | M+H | - | - |
| HILIC positive | 578.4854 | 27 | -0.08 (0.67) | 0.11 (0.67) | -0.19 (-0.33, -0.04) | 0.0104 | 0.7502 | DG(15:018:3(6Z9Z12Z)0:0); DG(15:018:3(9Z12Z15Z)0:0); DG(18:3(6Z9Z12Z)15:00:0); DG(18:3(9Z12Z15Z)15:00:0); DG(15:00:018:3n6); DG(15:00:018:3n3); DG(14:1n50:018:2n6); DG(14:1n50:020:2n6) | HMDB07075; HMDB07076; HMDB07271; HMDB07300; HMDB55996; HMDB56003; HMDB56142; HMDB56144 | M+H_[+1] | - | - |
| HILIC positive | 189.1346 | 75.3 | 0.07 (0.91) | -0.16 (1.03) | 0.22 (0.05, 0.39) | 0.0104 | 0.7502 | Homo-L-arginine; L-Targinine | HMDB00670; HMDB29416 | M+H | - | - |
| HILIC positive | 243.2319 | 266.1 | 0.15 (0.92) | -0.16 (1.15) | 0.31 (0.07, 0.54) | 0.0104 | 0.7502 | Pentadecanoic acid; Methyl tetradecanoate; Dodecyl propionate; Hexyl nonanoate; Heptyl octanoate; Octyl heptanoate; Pentyl decanoate; 3-Methylbutyl decanoate; 7(14)-Farnesene-912-diol; 2610-Trimethyldodecanoic acid; Isopropyllaurate; Ethyl tridecanoate | HMDB00826; HMDB30469; HMDB32250; HMDB32328; HMDB36218; HMDB36220; HMDB37306; HMDB38907; HMDB40998; HMDB41588; HMDB59820; HMDB59833 | M+H | - | - |
| HILIC positive | 324.326 | 27.7 | -0.44 (1.18) | -0.12 (1.08) | -0.32 (-0.56, -0.07) | 0.0109 | 0.7502 | N-(14-Methylhexadecanoyl)pyrrolidine | HMDB34373 | M+H | - | - |
| C18 negative | 461.3276 | 284.7 | -0.04 (0.87) | 0.17 (0.78) | -0.21 (-0.37, -0.05) | 0.0112 | 0.6059 | Dolichosterone; Polyporusterone F | HMDB34336; HMDB38499 | M-H | - | - |
| C18 negative | 103.0036 | 239.7 | 0.13 (0.6) | -0.06 (0.71) | 0.18 (0.04, 0.32) | 0.0114 | 0.6059 | Malonic acid; Hydroxypyruvic acid; Tartronate semialdehyde | HMDB00691; HMDB01352; HMDB06938 | M-H | - | - |
| C18 negative | 135.0451 | 289.5 | 0.18 (0.94) | -0.09 (1.04) | 0.27 (0.06, 0.49) | 0.0117 | 0.6059 | Phenylacetic acid; 2-Methylbenzoic acid; 4-Hydroxyphenylacetaldehyde; 4-Methylbenzoic acid; 4-Methoxybenzaldehyde; 3-Methoxybenzaldehyde; 3-(5-Methyl-2-furyl)prop-2-enal; 2-Hydroxyacetophenone; 2-Hydroxy-4-methylbenzaldehyde; 4-(2-Furanyl)-3-buten-2-one; 2-Methoxybenzaldehyde; Methyl benzoate; alpha-Methyl-2-furanacrolein; 3-(3-Furanyl)-2-methyl-2-propenal; Phenyl acetate; Benzyl formate | HMDB00209; HMDB02340; HMDB03767; HMDB29635; HMDB29686; HMDB31459; HMDB32400; HMDB32568; HMDB32603; HMDB33128; HMDB33766; HMDB33968; HMDB38176; HMDB40638; HMDB40733; HMDB41485 | M-H | - | - |
| HILIC positive | 193.0858 | 285.3 | 0.17 (0.81) | -0.16 (1.3) | 0.33 (0.07, 0.59) | 0.0119 | 0.7502 | (R)-Shinanolone; Methyl trans-p-methoxycinnamate; Allyl phenoxyacetate; Ethyl phenylglycidate; Dehydrozingerone; Isomyristicin; Myristicin; (Z)-4-Methoxy-6-(1-propenyl)-13-benzodioxole; Isoeugenol formate; Eugenyl formate; Ethyl 3-oxo-3-phenylpropanoate; Benzyl acetoacetate; 1-Hydroxychavicol acetate; 4-(34-Methylenedioxyphenyl)-2-butanone | HMDB30580; HMDB30752; HMDB31610; HMDB31623; HMDB32591; HMDB35275; HMDB35873; HMDB35894; HMDB37277; HMDB37877; HMDB40268; HMDB40446; HMDB41007; HMDB41481 | M+H | - | - |
| C18 negative | 171.0662 | 290.3 | 0.13 (0.92) | -0.15 (0.89) | 0.28 (0.06, 0.5) | 0.0119 | 0.6059 | 2-Octenedioic acid; cis-4-Octenedioic acid; trans-3-Octenedioic acid; 3-Isopropenylpentanedioic acid; Ethyl 24-dioxohexanoate; Diethyl fumarate | HMDB00341; HMDB04982; HMDB13312; HMDB32352; HMDB34851; HMDB38601 | M-H | - | - |
| C18 negative | 329.2331 | 72 | -0.13 (1.03) | 0.15 (1.01) | -0.28 (-0.5, -0.06) | 0.0121 | 0.6059 | 91213-TriHOME; 91013-TriHOME; 5812-Trihydroxy-9-octadecenoic acid; (9S10E12S13S)-91213-Trihydroxy-10-octadecenoic acid | HMDB04708; HMDB04710; HMDB30936; HMDB38555 | M-H | - | - |
| HILIC positive | 125.071 | 93.2 | -0.13 (1.25) | 0.16 (0.56) | -0.29 (-0.51, -0.06) | 0.0122 | 0.7502 | Methylimidazole acetaldehyde; 2-Methoxy-3-methylpyrazine; 2-Methoxy-5-methylpyrazine; 2-Methoxy-6-methylpyrazine | HMDB04181; HMDB31852; HMDB40141; HMDB40143 | M+H | - | - |
| HILIC positive | 203.0342 | 60.6 | -0.22 (1.03) | 0.05 (0.91) | -0.26 (-0.47, -0.06) | 0.0122 | 0.7502 | Bergaptol; Xanthotoxol | HMDB13679; HMDB29457 | M+H | - | - |
| C18 negative | 261.1342 | 294.7 | 0.15 (0.91) | -0.1 (0.73) | 0.25 (0.06, 0.45) | 0.0123 | 0.6059 | Phaseolic acid | HMDB31897 | M-H | - | - |
| HILIC positive | 153.0658 | 37.7 | 0.16 (1.21) | -0.12 (0.93) | 0.29 (0.06, 0.51) | 0.0123 | 0.7502 | N1-Methyl-2-pyridone-5-carboxamide; N1-Methyl-4-pyridone-3-carboxamide | HMDB04193; HMDB04194 | M+H | - | - |
| C18 negative | 85.0295 | 293.5 | 0.17 (0.97) | -0.1 (1.06) | 0.27 (0.06, 0.48) | 0.0126 | 0.6059 | Gamma-Butyrolactone; Oxolan-3-one; Diacetyl; But-2-enoic acid; Ethenyl acetate; Methyl acrylate; Isocrotonic acid | HMDB00549; HMDB02523; HMDB03407; HMDB10720; HMDB31209; HMDB33977; HMDB34439 | M-H | - | - |
| HILIC positive | 585.4296 | 27.1 | -0.1 (0.97) | -0.39 (1.11) | 0.29 (0.06, 0.51) | 0.0127 | 0.7502 | Cryptochrome; Cryptoxanthin diepoxide; Cryptoxanthin 56:58-diepoxide; (3S3R4xi)-betabeta-Carotene-334-triol; Cucurbitaxanthin A; Antheraxanthin A; Capsanthin; Flavoxanthin; Triphasiaxanthin; Prenigroxanthin; Mutatoxanthin; Lutein 56-epoxide | HMDB29857; HMDB29896; HMDB30537; HMDB30586; HMDB35319; HMDB35831; HMDB36590; HMDB36868; HMDB36914; HMDB37121; HMDB37564; HMDB41590 | M+H | - | - |
| C18 negative | 514.3057 | 182.7 | 0.11 (0.94) | -0.17 (1.13) | 0.28 (0.06, 0.5) | 0.0129 | 0.6059 | Flavidulol C | HMDB39150 | M-H_[-1] | - | - |
| C18 negative | 105.0193 | 20.2 | 0.14 (1.09) | -0.15 (1.05) | 0.29 (0.06, 0.53) | 0.0131 | 0.6059 | Glyceric acid; L-Glyceric acid; D-23-Dihydroxypropanoic acid | HMDB00139; HMDB06372; HMDB31818 | M-H | Glyceric acid | M-H |
| C18 negative | 453.1563 | 111.8 | -0.29 (1.16) | -0.06 (1.04) | -0.23 (-0.42, -0.05) | 0.0132 | 0.6059 | KB 2 | HMDB33666 | M-H | - | - |

| | | | | | | | | | | | | |
|---|---|---|---|---|---|---|---|---|---|---|---|---|
| C18 negative | 429.2627 | 78.9 | -0.34 (0.93) | -0.05 (1.08) | -0.29 (-0.52, -0.06) | 0.0133 | 0.6059 | 2-(2-Methylbutanoyl)-9-(3-methyl-2E-pentenoyl)-2b9a-dihydroxy-4Z10(14)-oplopadien-3-one | HMDB40994 | M-H | - | - |
| C18 negative | 175.0611 | 296 | 0.15 (0.97) | -0.1 (0.99) | 0.26 (0.05, 0.46) | 0.0133 | 0.6059 | 2-Isopropylmalic acid; 23-Dimethyl-3-hydroxyglutaric acid; 3-Isopropylmalate; 3-Hydroxy-2-methylglutarate; ()-Glycerol 12-diacetate | HMDB00402; HMDB02025; HMDB12156; HMDB29169; HMDB31712 | M-H | - | - |
| C18 negative | 446.3356 | 235.7 | -0.21 (1.14) | 0.07 (0.8) | -0.28 (-0.5, -0.06) | 0.0134 | 0.6059 | 13-Carboxy-gamma-tocopherol; (3beta5alpha6beta7alpha22E24R)-Ergosta-822-diene-3567-tetrol; (3beta5alpha6alpha9alpha22E24R)-Ergosta-722-diene-3569-tetrol; 359-Trihydroxyergost-7-en-6-one; Secasterone; 3-Dehydroteasterone | HMDB12557; HMDB32107; HMDB32123; HMDB32875; HMDB40999; HMDB41527 | M-H_[-1] | - | - |
| C18 negative | 129.0193 | 21 | 0.18 (0.93) | -0.07 (1) | 0.25 (0.05, 0.46) | 0.0140 | 0.6173 | Citramalic acid; 3-Hydroxyglutaric acid; D-2-Hydroxyglutaric acid; L-2-Hydroxyglutaric acid; Ribonolactone; D-Xylono-15-lactone; 2-Hydroxyglutarate; Glutaconic acid; Citraconic acid; Mesaconic acid; Itaconic acid; Gamma-delta-Dioxovaleric acid; Methyl hydrogen fumarate; 2-Hydroxyglutaric acid lactone; 25-Dioxopentanoate; 2-Pentendioate | HMDB00426; HMDB00428; HMDB00606; HMDB00694; HMDB01900; HMDB11676; HMDB59655; HMDB00620; HMDB00634; HMDB00749; HMDB02092; HMDB13233; HMDB33809; HMDB59743; HMDB60365; HMDB61185 | M-H2O-H; M-H | 2-methylmaleate | M-H |
| C18 negative | 144.0455 | 247.5 | 0.14 (0.74) | -0.05 (0.71) | 0.19 (0.04, 0.34) | 0.0144 | 0.6179 | 1H-Indole-3-carboxaldehyde | HMDB29737 | M-H | - | - |
| HILIC positive | 280.2351 | 27 | -0.26 (1.17) | 0.03 (0.97) | -0.29 (-0.52, -0.06) | 0.0144 | 0.7563 | Alpha-Linolenic acid; Gamma-Linolenic acid; Calendic acid; Punicic acid; Linolenelaidic acid | HMDB01388; HMDB03073; HMDB30962; HMDB30963; HMDB30964 | M+H_[+1] | - | - |
| C18 negative | 229.1231 | 105.8 | 0.64 (0.74) | 0.44 (0.86) | 0.2 (0.04, 0.35) | 0.0145 | 0.6179 | 812-Epoxy-4(15)711-eudesmatrien-1-one; (2E8Z)-Decadiene-46-diyn-1-yl 3-methylbutanoate; Curzerenone; Furanodienone; Cyclohexyl cinnamate | HMDB33235; HMDB35425; HMDB36155; HMDB36768; HMDB37706 | M-H | - | - |
| C18 negative | 241.2172 | 235.1 | 0.16 (0.94) | -0.09 (0.94) | 0.25 (0.05, 0.45) | 0.0146 | 0.6179 | Pentadecanoic acid; Methyl tetradecanoate; Dodecyl propionate; Hexyl nonanoate; Heptyl octanoate; Octyl heptanoate; Pentyl decanoate; 3-Methylbutyl decanoate; 7(14)-Farnesene-912-diol; 2610-Trimethyldodecanoic acid; Isopropyllaurate; Ethyl tridecanoate | HMDB00826; HMDB30469; HMDB32250; HMDB32328; HMDB36218; HMDB36220; HMDB37306; HMDB38907; HMDB40998; HMDB41588; HMDB59820; HMDB59833 | M-H | - | - |
| HILIC positive | 267.1706 | 29.9 | -0.28 (1.05) | -0.01 (1.07) | -0.27 (-0.49, -0.05) | 0.0153 | 0.7726 | Atenolol; Practolol | HMDB01924; HMDB15411 | M+H | - | - |
| HILIC positive | 149.045 | 297.3 | 0.14 (0.23) | -0.18 (1.61) | 0.32 (0.06, 0.59) | 0.0154 | 0.7726 | Citramalic acid; 3-Hydroxyglutaric acid; D-2-Hydroxyglutaric acid; L-2-Hydroxyglutaric acid; Ribonolactone; D-Xylono-15-lactone; 2-Propenyl propyl disulfide; Methyl 3-methyl-1-butenyl disulfide; 1-Propenyl propyl disulfide; 2-Hydroxyglutarate | HMDB00426; HMDB00428; HMDB00606; HMDB00694; HMDB01900; HMDB11676; HMDB33912; HMDB38890; HMDB41392; HMDB59655 | M+H | - | - |
| HILIC positive | 280.1754 | 250.7 | 0.19 (0.75) | -0.07 (1.06) | 0.27 (0.05, 0.48) | 0.0155 | 0.7748 | Etamiphylline | HMDB41889 | M+H | - | - |
| C18 negative | 171.1391 | 295.2 | 0.13 (0.7) | -0.17 (1.39) | 0.31 (0.06, 0.56) | 0.0168 | 0.6181 | Capric acid; 11-Diethoxy-2-hexene; Isopentyl isopentanoate; Methyl nonanoate; 7-Hydroxy-37-dimethyloctanal; Pentyl 3-methylbutanoate; Isooctyl acetate; Acetaldehyde 13-octanediol acetal; Ethyl 2-ethylhexanoate; Heptanal propyleneglycol acetal; Hexanal butane-23-diol acetal; Hexyl butyrate; Propyl heptanoate; 4-Methylnonanoic acid; trans-p-Menthane-18-diol; (1R2S3S4R)-p-Menthane-23-diol; 4-Ethyloctanoic acid; p-Menthane-38-diol; 2-Methylpropyl hexanoate; (1S3R4R)-p-Menthane-13-diol; Hexyl 2-methylpropanoate; 2-Methylbutyl 3-methylbutanoate; Octyl acetate; (Z)-1-(1-Ethoxyethoxy)-3-hexene; 2-Methylbutyl 2-methylbutanoate; 3-Methylbutyl 2-methylbutanoate; ()-3-Octyl acetate; Ethyl octanoate; Butyl hexanoate; 3-Methylbutyl pentanoate; 26-Dimethyl-7-octene-26-diol | HMDB00511; HMDB29760; HMDB30029; HMDB31264; HMDB31739; HMDB32045; HMDB32070; HMDB32155; HMDB32264; HMDB32302; HMDB32315; HMDB33620; HMDB34457; HMDB34849; HMDB35595; HMDB35723; HMDB36136; HMDB36145; HMDB36228; HMDB37387; HMDB37849; HMDB38038; HMDB38602; HMDB38783; HMDB39218; HMDB40157; HMDB40167; HMDB40195; HMDB40211; HMDB40529; HMDB41216 | M-H | - | - |
| HILIC positive | 296.2298 | 27.4 | -0.36 (0.99) | -0.07 (1.09) | -0.29 (-0.53, -0.05) | 0.0168 | 0.7938 | 13-OxoODE; 9-OxoODE; A-12(13)-EpODE; 13-HOTE; 15(16)-EpODE; 9(10)-EpODE; 9-HOTE; 17-Hydroxylinolenic acid; 10-Oxo-11-octadecen-13-olide; Squamostanal A; 1516-Epoxy-912-octadecadienoic acid; (2E4Z8E)-Colneleic acid; 1213-Epoxy-915-octadecadienoic acid; 2-Hydroxylinolenic acid; (9S10E12Z15Z)-9-Hydroxy-101215-octadecatrienoic acid; (9Z12Z14E)-16-Hydroxy-91214-octadecatrienoic acid; Sterebin D; 34-Dimethyl-5-pentyl-2-furanheptanoic acid | HMDB04668; HMDB04669; HMDB10200; HMDB10203; HMDB10206; HMDB10220; HMDB10224; HMDB11108; HMDB29786; HMDB29969; HMDB30950; HMDB30995; HMDB31088; HMDB31103; HMDB31934; HMDB34586; HMDB36832; HMDB39603 | M+H_[+1] | - | - |
| HILIC positive | 282.279 | 289.6 | -0.15 (0.95) | 0.12 (1.15) | -0.27 (-0.49, -0.05) | 0.0175 | 0.7957 | Oleamide | HMDB02117 | M+H | - | - |
| C18 negative | 651.4491 | 295.7 | -0.02 (1.01) | -0.28 (1.08) | 0.27 (0.05, 0.48) | 0.0176 | 0.6181 | Ginsenoside Rh5 | HMDB38016 | M-H | - | - |

| | | | | | | | | | | | | |
|---|---|---|---|---|---|---|---|---|---|---|---|---|
| HILIC positive | 225.185 | 115.6 | 0.07 (0.87) | -0.19 (1.05) | 0.26 (0.05, 0.47) | 0.0179 | 0.7957 | 58-Tetradecadienoic acid; Linalyl isobutyrate; Linalyl butyrate; Ethyl (2E6Z)-dodecadienoate; alpha-Terpineol butanoate; Isobornyl isobutyrate; Terpinyl isobutyrate; Allyl undecylenate; 2-Propenyl cyclohexanepentanoate; Bornyl butyrate; Neryl butyrate; Geranyl 2-methylpropanoate; Goshuyic acid | HMDB00560; HMDB30426; HMDB30427; HMDB31015; HMDB32053; HMDB32342; HMDB32524; HMDB36424; HMDB37814; HMDB38246; HMDB38259; HMDB38260; HMDB39129 | M+H | - | - |
| HILIC positive | 274.0595 | 68.6 | 0.09 (0.82) | -0.14 (1.17) | 0.22 (0.04, 0.41) | 0.0180 | 0.7957 | Ergothioneine | HMDB03045 | M+2Na-H | - | - |
| C18 negative | 963.7763 | 36.3 | -0.26 (1.04) | 0.01 (1.03) | -0.28 (-0.5, -0.05) | 0.0181 | 0.6181 | TG(22:6(4Z7Z10Z13Z16Z19Z)22:6(4Z7Z10Z13Z16Z19Z)o-18:0); TG(22:6(4Z7Z10Z13Z16Z19Z)o-18:022:6(4Z7Z10Z13Z16Z19Z)) | HMDB55950; HMDB55951 | M-H | - | - |
| C18 negative | 672.4943 | 22.4 | 0.64 (0.17) | 0.68 (0.15) | -0.04 (-0.08, -0.01) | 0.0181 | 0.6181 | PE(14:0P-18:1(11Z)); PE(14:0P-18:1(9Z)); PE(14:1(9Z)P-18:0); PE(16:1(9Z)P-16:0); PE(P-16:016:1(9Z)); PE(P-18:014:1(9Z)); PE(P-18:1(11Z)14:0); PE(P-18:1(9Z)14:0) | HMDB08852; HMDB08853; HMDB08884; HMDB08982; HMDB11339; HMDB11369; HMDB11401; HMDB11434 | M-H | - | - |
| C18 negative | 158.1186 | 171.9 | -0.16 (1) | 0.11 (0.91) | -0.27 (-0.49, -0.05) | 0.0181 | 0.6181 | DL-2-Aminooctanoic acid; Pregabalin | HMDB00991; HMDB14375 | M-H | - | - |
| HILIC positive | 87.0934 | 46.4 | -0.13 (0.88) | 0.13 (0.98) | -0.26 (-0.47, -0.04) | 0.0182 | 0.7957 | Piperidine | HMDB34301 | M+H_[+1] | - | - |
| C18 negative | 367.2253 | 275.1 | 0.31 (0.56) | 0.13 (0.75) | 0.18 (0.03, 0.33) | 0.0182 | 0.6181 | Perindopril | HMDB14928 | M-H | - | - |
| HILIC positive | 548.372 | 36.2 | -0.24 (1.47) | 0.08 (0.77) | -0.32 (-0.58, -0.05) | 0.0184 | 0.7957 | LysoPC(20:2(11Z14Z)) | HMDB10392 | M+H | - | - |
| HILIC positive | 290.1596 | 113.8 | 0.14 (0.82) | -0.13 (1.19) | 0.28 (0.05, 0.51) | 0.0185 | 0.7957 | 3-Methylglutarylcarnitine | HMDB00552 | M+H | - | - |
| HILIC positive | 939.8383 | 64.1 | 0.23 (0.39) | 0.12 (0.63) | 0.11 (0.02, 0.2) | 0.0194 | 0.7957 | TG(18:020:020:4(5Z8Z11Z14Z))iso6; TG(18:2(9Z12Z)20:1(11Z)20:1(11Z))iso3; TG(14:022:022:4(7Z10Z13Z16Z)); TG(14:024:020:4(5Z8Z11Z14Z)); TG(14:024:020:4(8Z11Z14Z17Z)); TG(14:020:3(5Z8Z11Z)24:1(15Z)); TG(14:024:1(15Z)20:3(5Z8Z11Z)); TG(14:024:1(15Z)20:3n6); TG(14:020:3n624:1(15Z)); TG(14:020:4(5Z8Z11Z14Z)24:0); TG(14:022:2(13Z16Z)22:2(13Z16Z)); TG(14:022:4(7Z10Z13Z16Z)22:0); TG(14:020:4(8Z11Z14Z17Z)24:0); TG(16:020:022:4(7Z10Z13Z16Z)); TG(16:022:020:4(5Z8Z11Z14Z)); TG(16:022:020:4(8Z11Z14Z17Z)); TG(16:024:018:4(6Z9Z12Z15Z)); TG(16:020:3(5Z8Z11Z)22:1(13Z)); TG(16:022:1(13Z)20:3(5Z8Z11Z)); TG(16:022:1(13Z)20:3n6); TG(16:024:1(15Z)18:3(6Z9Z12Z)); TG(16:024:1(15Z)18:3(9Z12Z15Z)); TG(16:018:3(6Z9Z12Z)24:1(15Z)); TG(16:020:3n622:1(13Z)); TG(16:020:4(5Z8Z11Z14Z)22:0); TG(16:022:4(7Z10Z13Z16Z)20:0); TG(16:018:3(9Z12Z15Z)24:1(15Z)); TG(16:018:4(6Z9Z12Z15Z)24:0); TG(16:020:4(8Z11Z14Z17Z)22:0); TG(18:018:022:4(7Z10Z13Z16Z)); TG(18:020:020:4(8Z11Z14Z17Z)); TG(18:022:018:4(6Z9Z12Z15Z)); TG(18:020:1(11Z)20:3(5Z8Z11Z)); TG(18:020:1(11Z)20:3n6); TG(18:020:3(5Z8Z11Z)20:1(11Z)); TG(18:022:1(13Z)18:3(6Z9Z12Z)); TG(18:022:1(13Z)18:3(9Z12Z15Z)); TG(18:018:3(6Z9Z12Z)22:1(13Z)); TG(18:020:3n620:1(11Z)); TG(18:020:4(5Z8Z11Z14Z)20:0); TG(18:022:4(7Z10Z13Z16Z)18:0); TG(18:018:3(9Z12Z15Z)22:1(13Z)); TG(18:018:4(6Z9Z12Z15Z)22:0); TG(18:020:4(8Z11Z14Z17Z)20:0); TG(20:016:022:4(7Z10Z13Z16Z)); TG(20:018:020:4(5Z8Z11Z14Z)); TG(20:018:020:4(8Z11Z14Z17Z)); TG(20:020:018:4(6Z9Z12Z15Z)); TG(20:018:1(11Z)20:3(5Z8Z11Z)); TG(20:018:1(11Z)20:3n6); TG(20:018:1(9Z)20:3(5Z8Z11Z)); TG(20:018:1(9Z)20:3n6); TG(20:020:1(11Z)18:3(6Z9Z12Z)); TG(20:020:1(11Z)18:3(9Z12Z15Z)); TG(20:020:3(5Z8Z11Z)18:1(11Z)); | HMDB05401; HMDB05470; HMDB42232; HMDB42260; HMDB42266; HMDB42435; HMDB42493; HMDB42499; HMDB42615; HMDB42637; HMDB42681; HMDB42696; HMDB42817; HMDB43960; HMDB43986; HMDB43992; HMDB44019; HMDB44176; HMDB44203; HMDB44209; HMDB44235; HMDB44242; HMDB44289; HMDB44344; HMDB44364; HMDB44419; HMDB44485; HMDB44505; HMDB44532; HMDB44767; HMDB44798; HMDB44824; HMDB44974; HMDB44980; HMDB45000; HMDB45032; HMDB45039; HMDB45110; HMDB45162; HMDB45182; HMDB45235; HMDB45299; HMDB45318; HMDB45344; HMDB45546; HMDB45570; HMDB45576; HMDB45601; HMDB45719; HMDB45725; HMDB45745; HMDB45751; HMDB45775; HMDB45782; HMDB45794; HMDB45795; HMDB45878; HMDB45880; HMDB45900; HMDB45930; HMDB45932; HMDB45950; HMDB45951; HMDB46082; HMDB46101; HMDB46272; HMDB46320; HMDB46326; HMDB46350; HMDB46463; HMDB46469; HMDB46492; HMDB46499; HMDB46517; HMDB46524; HMDB46559; HMDB46660; HMDB46661; HMDB46709; HMDB46835; HMDB46836; HMDB47018; HMDB47024; HMDB47071; HMDB47179; HMDB47185; HMDB47207; HMDB47214; HMDB47294; HMDB47391; HMDB47438; HMDB47559; HMDB47867; HMDB47873; | M+H | - | - |

| | | | | | | | | | | | | |
|---|---|---|---|---|---|---|---|---|---|---|---|---|
| | | | | | | | | TG(20:020:3(5Z8Z11Z)18:1(9Z)); TG(20:018:2(9Z12Z)18:2(9Z12Z)); TG(20:018:2(9Z12Z)20:2n6); TG(20:018:3(6Z9Z12Z)20:1(11Z)); TG(20:020:2n618:2(9Z12Z)); TG(20:020:2n620:2n6); TG(20:020:3n618:1(11Z)); TG(20:020:3n618:1(9Z)); TG(20:018:3(9Z12Z15Z)20:1(11Z)); TG(20:018:4(6Z9Z12Z15Z)20:0); TG(22:014:022:4(7Z10Z13Z16Z)); TG(22:016:020:4(5Z8Z11Z14Z)); TG(22:016:020:4(8Z11Z14Z17Z)); TG(22:018:018:4(6Z9Z12Z15Z)); TG(22:016:1(9Z)20:3(5Z8Z11Z)); TG(22:016:1(9Z)20:3n6); TG(22:018:1(11Z)18:3(6Z9Z12Z)); TG(22:018:1(11Z)18:3(9Z12Z15Z)); TG(22:018:1(9Z)18:3(6Z9Z12Z)); TG(22:018:1(9Z)18:3(9Z12Z15Z)); TG(22:020:3(5Z8Z11Z)16:1(9Z)); TG(22:018:3(6Z9Z12Z)18:1(11Z)); TG(22:018:3(6Z9Z12Z)18:1(9Z)); TG(22:020:3n616:1(9Z)); TG(22:018:3(9Z12Z15Z)18:1(11Z)); TG(22:018:3(9Z12Z15Z)18:1(9Z)); TG(24:014:020:4(5Z8Z11Z14Z)); TG(24:014:020:4(8Z11Z14Z17Z)); TG(24:016:018:4(6Z9Z12Z15Z)); TG(24:014:1(9Z)20:3(5Z8Z11Z)); TG(24:014:1(9Z)20:3n6); TG(24:016:1(9Z)18:3(6Z9Z12Z)); TG(24:016:1(9Z)18:3(9Z12Z15Z)); TG(24:020:3(5Z8Z11Z)14:1(9Z)); TG(24:018:3(6Z9Z12Z)16:1(9Z)); TG(24:020:3n614:1(9Z)); TG(24:018:3(9Z12Z15Z)16:1(9Z)); TG(14:1(9Z)24:020:3(5Z8Z11Z)); TG(14:1(9Z)24:020:3n6); TG(14:1(9Z)22:1(13Z)22:2(13Z16Z)); TG(14:1(9Z)22:2(13Z16Z)22:1(13Z)); TG(16:1(9Z)22:020:3(5Z8Z11Z)); TG(16:1(9Z)22:020:3n6); TG(16:1(9Z)24:018:3(6Z9Z12Z)); TG(16:1(9Z)24:018:3(9Z12Z15Z)); TG(16:1(9Z)20:1(11Z)22:2(13Z16Z)); TG(16:1(9Z)22:2(13Z16Z)20:1(11Z)); TG(18:1(11Z)20:020:3(5Z8Z11Z)); TG(18:1(11Z)20:020:3n6); TG(18:1(11Z)22:018:3(6Z9Z12Z)); TG(18:1(11Z)22:018:3(9Z12Z15Z)); TG(18:1(11Z)18:1(11Z)22:2(13Z16Z)); TG(18:1(11Z)18:1(9Z)22:2(13Z16Z)); TG(18:1(11Z)22:2(13Z16Z)18:1(11Z)); TG(18:1(11Z)22:2(13Z16Z)18:1(9Z)); TG(18:1(9Z)20:020:3(5Z8Z11Z)); TG(18:1(9Z)20:020:3n6); TG(18:1(9Z)22:018:3(6Z9Z12Z)); TG(18:1(9Z)22:018:3(9Z12Z15Z)); TG(18:1(9Z)18:1(11Z)22:2(13Z16Z)); TG(18:1(9Z)18:1(9Z)22:2(13Z16Z)); TG(18:1(9Z)22:2(13Z16Z)18:1(9Z)); TG(20:1(11Z)18:020:3(5Z8Z11Z)); TG(20:1(11Z)18:020:3n6); TG(20:1(11Z)20:018:3(6Z9Z12Z)); TG(20:1(11Z)20:018:3(9Z12Z15Z)); TG(20:1(11Z)16:1(9Z)22:2(13Z16Z)); TG(20:3(5Z8Z11Z)14:024:1(15Z)); TG(20:3(5Z8Z11Z)16:022:1(13Z)); TG(22:1(13Z)16:020:3n6); TG(22:1(13Z)18:018:3(6Z9Z12Z)); TG(22:1(13Z)18:018:3(9Z12Z15Z)); TG(22:1(13Z)14:1(9Z)22:2(13Z16Z)); TG(24:1(15Z)14:020:3n6); TG(24:1(15Z)16:018:3(6Z9Z12Z)); TG(24:1(15Z)16:018:3(9Z12Z15Z)); TG(18:2(9Z12Z)20:018:2(9Z12Z)); TG(18:2(9Z12Z)20:020:2n6); TG(20:2n620:020:2n6); TG(22:2(13Z16Z)14:022:2(13Z16Z)) | HMDB48036; HMDB48190; HMDB48527; HMDB48533; HMDB48553; HMDB48560; HMDB48667; HMDB48856; HMDB49159; HMDB49165; HMDB49184; HMDB49191; HMDB49272; HMDB49293; HMDB49492; HMDB49493; HMDB49783; HMDB49789; HMDB49807; HMDB49814; HMDB49891; HMDB49911; HMDB50101; HMDB50358; HMDB50364; HMDB50381; HMDB50388; HMDB50461; HMDB50871; HMDB50906; HMDB51447; HMDB51462; HMDB51469; HMDB51534; HMDB51921; HMDB51951; HMDB51958; HMDB52456; HMDB52458; HMDB53316; HMDB54341 | | | |
| C18 negative | 350.2097 | 148.3 | 0.17 (0.9) | -0.07 (0.97) | 0.25 (0.04, 0.45) | 0.0195 | 0.6181 | Sphingosine 1-phosphate (d16:1-P) | HMDB60061 | M-H | - | - |

| | | | | | | | | | | | | |
|---|---|---|---|---|---|---|---|---|---|---|---|---|
| C18 negative | 420.2539 | 242.5 | 0.26 (0.76) | 0.07 (1.02) | 0.19 (0.03, 0.34) | 0.0196 | 0.6181 | PC-M6 | HMDB38568 | M-H | - | - |
| HILIC positive | 517.3881 | 26.8 | -0.26 (1.02) | -0.01 (0.96) | -0.25 (-0.47, -0.04) | 0.0199 | 0.7957 | 22-Acetylpriverogenin B | HMDB34645 | M+H | - | - |
| HILIC positive | 207.0507 | 90.3 | 0.31 (0.96) | 0.09 (1.03) | 0.22 (0.04, 0.41) | 0.0202 | 0.7957 | 2-Methylcitric acid; (R)-lipoic acid; Homocitric acid; Methylisocitric acid; (S)-lipoic acid; 3-(Carboxymethyl)-3-hydroxypentanedioic acid | HMDB00379; HMDB01451; HMDB03518; HMDB06471; HMDB14312; HMDB59932 | M+H | - | - |
| HILIC positive | 143.1431 | 176.6 | -0.14 (1.1) | 0.11 (0.75) | -0.25 (-0.47, -0.04) | 0.0202 | 0.7957 | 2-Methyl-4-heptanone; Dihydroisophorol; (Z)-3-Nonen-1-ol; 2-Nonanone; 3-Nonanone; Amyl propyl ketone; 26-Dimethyl-4-heptanone; 2-Methyloctanal; 355-Trimethylhexanal; 1-Nonen-3-ol; ()-26-Dimethyl-6-hepten-1-ol; (Z)-6-Nonen-1-ol; 2-Nonen-1-ol; (Z)-2-Nonen-1-ol; (E)-3-Nonen-1-ol; Nonanal; (E)-6-Nonenol | HMDB13821; HMDB29557; HMDB30959; HMDB31266; HMDB31267; HMDB31268; HMDB31417; HMDB31684; HMDB31730; HMDB34152; HMDB37304; HMDB40581; HMDB41498; HMDB41499; HMDB41601; HMDB59835; HMDB59887 | M+H | - | - |
| C18 negative | 443.317 | 221.6 | -0.16 (1.14) | 0.1 (0.76) | -0.26 (-0.48, -0.04) | 0.0207 | 0.6181 | 1-a24R25-Trihydroxyvitamin D2; 59-Epidioxy-3-hydroxyergost-7-en-6-one; (3beta5alpha6alpha7beta14alpha22E24R)-56-Epoxyergosta-822-diene-3714-triol; 56:89-Diepoxyergost-22-ene-37beta-diol; (3beta5alpha6beta9alpha22E24R)-59-Epidioxyergosta-722-diene-36-diol | HMDB06227; HMDB32667; HMDB32691; HMDB33647; HMDB38977 | M-H | - | - |
| HILIC positive | 577.4812 | 27 | -0.14 (0.83) | 0.05 (0.85) | -0.18 (-0.34, -0.03) | 0.0212 | 0.7957 | DG(15:018:3(6Z9Z12Z)0:0); DG(15:018:3(9Z12Z15Z)0:0); DG(18:3(6Z9Z12Z)15:00:0); DG(18:3(9Z12Z15Z)15:00:0); DG(15:00:018:3n6); DG(15:00:018:3n3); DG(14:1n50:018:2n6); DG(14:1n50:020:2n6) | HMDB07075; HMDB07076; HMDB07271; HMDB07300; HMDB55996; HMDB56003; HMDB56142; HMDB56144 | M+H | - | - |
| HILIC positive | 489.3566 | 27.5 | -0.3 (1.14) | -0.02 (1.02) | -0.28 (-0.52, -0.04) | 0.0212 | 0.7957 | Pitheduloside I; Arjunolic acid; Camelliagenin B; Madasiatic acid; Esculentic acid (Diplazium); Hovenolactone; Centellasapogenol A; Glyyunnansapogenin B; Euscaphic acid; (3beta6alpha19alpha)-3619-Trihydroxy-12-ursen-28-oic acid; Ganoderiol D; Ananasic acid; 16beta-Hydroxystellatogenin; 21beta-Hydroxyhederagenin | HMDB34036; HMDB34502; HMDB34531; HMDB35118; HMDB35782; HMDB35957; HMDB36311; HMDB36639; HMDB36651; HMDB36961; HMDB37782; HMDB38116; HMDB40391; HMDB41039 | M+H | - | - |
| C18 negative | 296.2673 | 265.6 | 0.2 (0.71) | -0.02 (0.99) | 0.22 (0.03, 0.4) | 0.0213 | 0.6181 | Nonadeca-10(Z)-enoic acid; 46-Nonadecanedione | HMDB13622; HMDB35575 | M-H_[-1] | - | - |
| HILIC positive | 477.3923 | 26.7 | -0.4 (1.1) | -0.14 (1.06) | -0.27 (-0.49, -0.04) | 0.0217 | 0.7957 | Panaxatriol | HMDB38244 | M+H | - | - |
| C18 negative | 651.3366 | 48.5 | 0.6 (0.45) | 0.69 (0.29) | -0.1 (-0.18, -0.01) | 0.0221 | 0.6181 | Uzarigenin 3-xylosyl-(1-2)-rhamnoside | HMDB36296 | M-H | - | - |
| HILIC positive | 431.3896 | 26.9 | 0.1 (0.99) | -0.14 (0.99) | 0.23 (0.03, 0.43) | 0.0221 | 0.7957 | Alpha-Tocopherol; 4alpha-Hydroxymethyl-4beta-methyl-5alpha-cholesta-8-en-3beta-ol; Ikshusterol; 4-beta-Hydroxymethyl-4-alpha-methyl-5-alpha-cholest-7-en-3-beta-ol | HMDB01893; HMDB12171; HMDB30022; HMDB59642 | M+H | - | - |
| C18 negative | 545.4574 | 234.1 | 0.11 (1.01) | -0.14 (0.94) | 0.25 (0.04, 0.46) | 0.0225 | 0.6181 | Diepomuricanin A | HMDB40921 | M-H | - | - |
| C18 negative | 197.0815 | 295.1 | 0.08 (0.76) | -0.16 (1.09) | 0.25 (0.03, 0.46) | 0.0230 | 0.6181 | Guaifenesin; 5-(3E-Pentenyl)tetrahydro-2-oxo-3-furancarboxylic acid; 3-(4-Hydroxy-3-methoxyphenyl)-12-propanediol; (2E4E)-27-Dimethyl-24-octadienedioic acid; 34-Methylene suberic acid; 23-Methylene suberic acid | HMDB04998; HMDB30991; HMDB33093; HMDB34099; HMDB59768; HMDB59779 | M-H | - | - |
| C18 negative | 517.3894 | 238.5 | -0.13 (0.95) | 0.09 (0.86) | -0.22 (-0.41, -0.03) | 0.0231 | 0.6181 | Ganoderiol C | HMDB37781 | M-H | - | - |
| HILIC positive | 445.3674 | 27.3 | -0.26 (1.35) | 0.05 (0.99) | -0.31 (-0.58, -0.04) | 0.0233 | 0.7957 | 3-beta-Hydroxy-4-beta-methyl-5-alpha-cholest-7-ene-4-alpha-carboxylate; 4alpha-Carboxy-4beta-methyl-5alpha-cholesta-8-en-3beta-ol; (3beta5alpha6beta22E24R)-23-Methylergosta-722-diene-356-triol; Schleicherastatin 5 | HMDB11662; HMDB12165; HMDB33633; HMDB35804 | M+H | - | - |
| HILIC positive | 133.068 | 153.2 | -0.07 (1.16) | 0.16 (0.54) | -0.23 (-0.43, -0.03) | 0.0235 | 0.7957 | 4-Mercapto-4-methyl-2-pentanone; 25-Dimethyl-3-mercaptotetrahydrofuran; 4-(Methylthio)-2-pentanone; 3-Mercapto-2-methylpentanal; S-Methyl 3-methylthiobutyrate | HMDB31519; HMDB32237; HMDB32430; HMDB35176; HMDB39843 | M+H | - | - |
| HILIC positive | 119.0604 | 286.2 | -0.11 (1) | 0.12 (0.79) | -0.23 (-0.43, -0.03) | 0.0236 | 0.7957 | 7-Azaindolizine; 5H-Cyclopentapyrazine | HMDB33576; HMDB40040 | M+H | - | - |
| C18 negative | 479.3376 | 217.4 | -0.11 (1.11) | 0.14 (0.86) | -0.25 (-0.46, -0.03) | 0.0238 | 0.6181 | Brassinolide; 24-Epibrassinolide | HMDB34081; HMDB41130 | M-H | - | - |
| C18 negative | 265.1807 | 214.7 | 0.09 (0.84) | -0.14 (1.18) | 0.23 (0.03, 0.43) | 0.0239 | 0.6181 | 4-Hydroxy-3-methoxy-210-bisaboladien-9-one; Tetranor 12-HETE | HMDB33917; HMDB60055 | M-H | - | - |
| HILIC positive | 422.9844 | 67.2 | 0.13 (0.8) | -0.14 (1.26) | 0.27 (0.04, 0.51) | 0.0239 | 0.7957 | Chlorophenol red | HMDB36699 | M+H | - | - |
| C18 negative | 242.1843 | 139.1 | 0.1 (0.99) | -0.17 (1.25) | 0.27 (0.04, 0.5) | 0.0241 | 0.6181 | 3-Oxotetradecanoic acid; 6-Ketomyristic acid; L-Menthyl (RS)-3-hydroxybutyrate | HMDB10730; HMDB30982; HMDB32370 | M-H_[-1] | - | - |
| C18 negative | 150.0561 | 27.3 | -0.47 (0.96) | -0.25 (1.05) | -0.22 (-0.4, -0.03) | 0.0242 | 0.6181 | Acetaminophen; 2-Phenylglycine; Dopamine quinone; Leukoaminochrome; Methyl 2-aminobenzoate; 2-(Methylamino)benzoic acid; 3alpha477alpha-Tetrahydro-1H-isoindole-13(2H)-dione; Ethyl nicotinate; 2-Amino-3-methylbenzoate | HMDB01859; HMDB02210; HMDB12219; HMDB12992; HMDB29703; HMDB32609; HMDB33903; HMDB59846; HMDB60680 | M-H | - | - |
| HILIC positive | 201.0387 | 94.7 | 0.09 (0.5) | 0.23 (0.49) | -0.14 (-0.26, -0.02) | 0.0246 | 0.7957 | 4-Fumarylacetoacetic acid; Maleylacetoacetic acid | HMDB01268; HMDB02052 | M+H | - | - |
| HILIC positive | 497.1305 | 78 | -0.48 (1.01) | -0.26 (1.02) | -0.22 (-0.41, -0.03) | 0.0250 | 0.7957 | 4-Methyl-(-)-epigallocatechin 3-glucuronide; 4-Methyl-(-)-epigallocatechin 7-glucuronide | HMDB41675; HMDB41676 | M+H | - | - |

| | | | | | | | | | | | |
|---|---|---|---|---|---|---|---|---|---|---|---|
| C18 negative | 505.3536 | 165.5 | -0.19 (1.25) | 0.06 (0.93) | -0.25 (-0.47, -0.03) | 0.0250 | 0.6181 | Theasapogenol A | HMDB34519 | M-H | - | - |
| HILIC positive | 597.4484 | 25.7 | -0.12 (1.07) | 0.06 (0.86) | -0.18 (-0.34, -0.02) | 0.0252 | 0.7957 | DG(15:018:4(6Z9Z12Z15Z)0:0); DG(18:4(6Z9Z12Z15Z)15:00:0); DG(15:00:018:4n3) | HMDB07077; HMDB07329; HMDB56004 | M+Na | - | - |
| C18 negative | 766.5388 | 290.7 | -0.11 (1.09) | 0.13 (0.73) | -0.23 (-0.44, -0.03) | 0.0252 | 0.6181 | PC(15:020:4(5Z8Z11Z14Z)); PC(15:020:4(8Z11Z14Z17Z)); PC(20:4(5Z8Z11Z14Z)15:0); PC(20:4(8Z11Z14Z17Z)15:0); PE(16:022:4(7Z10Z13Z16Z)); PE(16:1(9Z)20:3(8Z11Z14Z)); PE(18:020:4(5Z8Z11Z14Z)); PE(18:020:4(8Z11Z14Z17Z)); PE(18:1(11Z)20:3(5Z8Z11Z)); PE(18:1(11Z)20:3(8Z11Z14Z)); PE(18:1(9Z)20:3(5Z8Z11Z)); PE(18:1(9Z)20:3(8Z11Z14Z)); PE(18:2(9Z12Z)20:2(11Z14Z)); PE(18:3(6Z9Z12Z)20:1(11Z)); PE(18:3(9Z12Z15Z)20:1(11Z)); PE(18:4(6Z9Z12Z15Z)20:0); PE(20:018:4(6Z9Z12Z15Z)); PE(20:1(11Z)18:3(6Z9Z12Z)); PE(20:1(11Z)18:3(9Z12Z15Z)); PE(20:2(11Z14Z)18:2(9Z12Z)); PE(20:3(5Z8Z11Z)18:1(11Z)); PE(20:3(5Z8Z11Z)18:1(9Z)); PE(20:3(8Z11Z14Z)18:1(11Z)); PE(20:3(8Z11Z14Z)18:1(9Z)); PE(20:4(5Z8Z11Z14Z)18:0); PE(20:4(8Z11Z14Z17Z)18:0); PE(22:4(7Z10Z13Z16Z)16:0) | HMDB07949; HMDB07950; HMDB08428; HMDB08461; HMDB08943; HMDB08969; HMDB09003; HMDB09004; HMDB09034; HMDB09035; HMDB09067; HMDB09068; HMDB09099; HMDB09131; HMDB09164; HMDB09196; HMDB09228; HMDB09259; HMDB09260; HMDB09291; HMDB09322; HMDB09323; HMDB09355; HMDB09356; HMDB09387; HMDB09420; HMDB09583 | M-H | - | - |
| HILIC positive | 217.1587 | 295 | 0.11 (0.56) | -0.11 (1.09) | 0.22 (0.03, 0.41) | 0.0253 | 0.7957 | 2-Hexyl-3-phenyl-2-propenal; (R)-ar-Turmerone; 5beta-137(11)-Eudesmatrien-8-one; 13511-Bisabolatetraen-10-one; 12-Dehydro-alpha-cyperone; Furanodiene; Isogermafurene; 110-Oxidocalamenene | HMDB31736; HMDB35612; HMDB35965; HMDB36108; HMDB36589; HMDB36769; HMDB38147; HMDB59863 | M+H | - | - |
| C18 negative | 433.2358 | 166.7 | 0.29 (0.53) | 0.4 (0.38) | -0.11 (-0.2, -0.01) | 0.0255 | 0.6181 | LPA(0:018:2(9Z12Z)); LPA(18:2(9Z12Z)0:0) | HMDB07852; HMDB07856 | M-H | - | - |
| HILIC positive | 250.0642 | 48.6 | -0.12 (0.94) | 0.08 (0.74) | -0.2 (-0.38, -0.03) | 0.0256 | 0.7957 | Sulfapyridine | HMDB15028 | M+H | - | - |
| C18 negative | 111.02 | 220.5 | 0.02 (0.92) | -0.3 (1.54) | 0.32 (0.04, 0.59) | 0.0261 | 0.6181 | Uracil; 4-Carboxypyrazole | HMDB00300; HMDB60760 | M-H | - | - |
| HILIC positive | 819.5132 | 39.5 | 0.67 (0.44) | 0.56 (0.57) | 0.11 (0.01, 0.21) | 0.0262 | 0.7957 | PG(18:2(9Z12Z)22:6(4Z7Z10Z13Z16Z19Z)); PG(18:3(6Z9Z12Z)22:5(4Z7Z10Z13Z16Z)); PG(18:3(6Z9Z12Z)22:5(7Z10Z13Z16Z19Z)); PG(18:3(9Z12Z15Z)22:5(4Z7Z10Z13Z16Z)); PG(18:3(9Z12Z15Z)22:5(7Z10Z13Z16Z19Z)); Quinquenoside F1 | HMDB10659; HMDB10672; HMDB10673; HMDB10687; HMDB10688; HMDB39398 | M+H | - | - |
| HILIC positive | 375.0654 | 74.4 | 0.33 (0.85) | 0.08 (1.26) | 0.25 (0.03, 0.47) | 0.0263 | 0.7957 | 3-Glucosyl-246-trihydroxyacetophenone; 7-Hydroxymethyl-12-methylbenzaanthracene sulfate | HMDB40621; HMDB60420 | M+2Na-H; M+Na | - | - |
| C18 negative | 337.205 | 233.5 | 0.2 (0.81) | -0.01 (0.79) | 0.2 (0.02, 0.38) | 0.0264 | 0.6181 | Lisuride | HMDB14727 | M-H | - | - |
| HILIC positive | 599.4646 | 25.6 | -0.12 (1) | 0.1 (0.93) | -0.22 (-0.41, -0.03) | 0.0266 | 0.7957 | DG(15:018:3(6Z9Z12Z)0:0); DG(15:018:3(9Z12Z15Z)0:0); DG(18:3(6Z9Z12Z)15:00:0); DG(18:3(9Z12Z15Z)15:00:0); DG(15:00:018:3n6); DG(15:00:018:3n3); DG(14:1n50:018:2n6); DG(14:1n50:020:2n6) | HMDB07075; HMDB07076; HMDB07271; HMDB07300; HMDB55996; HMDB56003; HMDB56142; HMDB56144 | M+Na | - | - |
| HILIC positive | 576.4699 | 27 | -0.07 (0.97) | 0.12 (0.84) | -0.19 (-0.36, -0.02) | 0.0271 | 0.7957 | DG(15:018:4(6Z9Z12Z15Z)0:0); DG(18:4(6Z9Z12Z15Z)15:00:0); DG(15:00:018:4n3) | HMDB07077; HMDB07329; HMDB56004 | M+H_[+1] | - | - |
| C18 negative | 297.2432 | 211.2 | 0.01 (0.97) | 0.19 (0.62) | -0.18 (-0.33, -0.02) | 0.0271 | 0.6181 | 3-Oxooctadecanoic acid; 9-Oxooctadecanoic acid; 10-Oxooctadecanoic acid; 11-Oxooctadecanoic acid; 5-Hexyltetrahydro-2-furanoctanoic acid; 5-Oxooctadecanoic acid; Ricinoleic acid | HMDB10736; HMDB30979; HMDB30980; HMDB30981; HMDB31127; HMDB34074; HMDB34297 | M-H | - | - |
| C18 negative | 163.1128 | 196 | -0.2 (0.88) | 0.05 (1.09) | -0.26 (-0.49, -0.03) | 0.0279 | 0.6181 | 4-(1133-Tetramethylbutyl)-phenol; 1-(1-Methoxy-1-methylethyl)-4-methylbenzene; Benzyl butyl ether; trans-Jasmone; 5-Phenyl-1-pentanol; 1-Phenyl-2-pentanol; 2-Methyl-4-phenyl-2-butanol; 2-Butyl-4-methylphenol; (Z)-3-Methyl-2-(2-pentenyl)-2-cyclopenten-1-one; Thymol methyl ether; Norbicycloekasantalal; Santalone; Norecasantalal; 2-Methyl-3-(2-pentenyl)-2-cyclopenten-1-one | HMDB13825; HMDB29653; HMDB31311; HMDB31454; HMDB31624; HMDB31625; HMDB31866; HMDB32060; HMDB35601; HMDB35989; HMDB36719; HMDB36814; HMDB36816; HMDB37294 | M-H | - | - |
| HILIC positive | 103.039 | 290.7 | 0.12 (0.56) | -0.1 (1.07) | 0.22 (0.02, 0.41) | 0.0280 | 0.7967 | 2-Ketobutyric acid; Acetoacetic acid; 2-Methyl-3-oxopropanoic acid; Succinic acid semialdehyde; (S)-Methylmalonic acid semialdehyde; 4-Hydroxycrotonic acid; Acetic anhydride | HMDB00005; HMDB00060; HMDB01172; HMDB01259; HMDB02217; HMDB03381; HMDB31646 | M+H | - | - |
| HILIC positive | 127.1117 | 295.5 | 0.11 (0.46) | -0.18 (1.62) | 0.29 (0.03, 0.55) | 0.0281 | 0.7967 | (E)-2-octenal; 2-Octenal; (3xi5Z)-15-Octadien-3-ol; 244-Trimethylcyclopentanone; 2-Octen-4-one; 1-Octen-3-one; 3-Methyl-3-hepten-2-one; 5-Methyl-5-hepten-2-one; 6-Methyl-3-hepten-2-one trans-; 4-Octen-3-one; 3-Octen-2-one; (SE)-Filbertone; 5-Octen-2-one; Sulcatone; 2-Ethylidenehexanal; 6-Octenal; (Z)-5-Octenal; 25-Octadien-1-ol; (3E5Z)-35-Octadien-1-ol; (2E4E)-24-Octadien-1-ol | HMDB13809; HMDB30961; HMDB30966; HMDB31197; HMDB31301; HMDB31309; HMDB31590; HMDB31591; HMDB32403; HMDB32451; HMDB33547; HMDB35242; HMDB35390; HMDB35915; HMDB37152; HMDB39769; HMDB39770; HMDB40146; HMDB40149; HMDB40151 | M+H | - | - |
| C18 negative | 596.4976 | 279.5 | -0.03 (0.84) | 0.15 (0.61) | -0.17 (-0.33, -0.02) | 0.0282 | 0.6181 | Glycerol triundecanoate | HMDB31089 | M-H_[-1] | - | - |

| | | | | | | | | | | | | |
|---|---|---|---|---|---|---|---|---|---|---|---|---|
| C18 negative | 228.2048 | 224.2 | 0.1 (0.97) | -0.13 (1.05) | 0.23 (0.02, 0.44) | 0.0286 | 0.6181 | Myristic acid; 2610-Trimethylundecanoic acid; 12-Methyltridecanoic acid; Hexanal octane-13-diol acetal; Undecanal propyleneglycol acetal; Ethyl dodecanoate; Heptyl heptanoate; Hexyl octanoate; Decyl butanoate; 3-Methylbutyl nonanoate; Nonyl isovalerate; Dodecyl acetate; Isobutyl decanoate | HMDB00806; HMDB02221; HMDB31072; HMDB32318; HMDB32548; HMDB33788; HMDB34461; HMDB36216; HMDB37312; HMDB37495; HMDB38034; HMDB38969; HMDB59865 | M-H_[-1] | - | - |
| HILIC positive | 133.1011 | 26.7 | -0.24 (1.28) | 0.06 (0.99) | -0.3 (-0.57, -0.03) | 0.0288 | 0.7968 | p-Mentha-1358-tetraene | HMDB29641 | M+H | - | - |
| HILIC positive | 501.3932 | 26.8 | -0.23 (1.06) | -0.01 (0.97) | -0.23 (-0.43, -0.02) | 0.0290 | 0.7968 | Norambreinolide; 3-Methyl-alpha-ionyl acetate; 22-Bis(2-methylpropoxy)ethylbenzene; Ganoderiol C | HMDB35293; HMDB37631; HMDB37712; HMDB37781 | 2M+H; M+H-H2O | - | - |
| C18 negative | 329.2484 | 239.2 | 0.14 (0.93) | -0.07 (0.91) | 0.21 (0.02, 0.4) | 0.0291 | 0.6181 | Docosapentaenoic acid (22n-6); Docosapentaenoic acid; Ethyl abietate; ent-16-Kauren-19-ol acetate; 1-Phenyl-13-hexadecanedione; 2-Methyl-5-(811-pentadecadienyl)-13-benzenediol; 48121519-Docosapentaenoic acid; Ethyl icosapentate; Docosa-47101316-pentanoic acid | HMDB01976; HMDB06528; HMDB32257; HMDB35024; HMDB35581; HMDB38909; HMDB39133; HMDB39530; HMDB60113 | M-H | C22-5-fa | M-H |
| C18 negative | 401.3428 | 299.1 | -0.69 (0.79) | -0.47 (0.93) | -0.21 (-0.41, -0.02) | 0.0291 | 0.6181 | 24-Hydroxycholesterol; 7a-Hydroxycholesterol; 27-Hydroxycholesterol; Delta-Tocopherol; 22b-Hydroxycholesterol; 7b-Hydroxycholesterol; 25-Hydroxycholesterol; 20alpha-Hydroxycholesterol; 7a-Hydroxy-5b-cholestan-3-one; (24R)-Cholest-5-ene-3-beta24-diol ; 4b-Hydroxycholesterol; (3beta5alpha6alpha)-Cholest-8-ene-36-diol; eta-Tocopherol; 3-Hydroxy-1-phenyl-1-heneicosanone; 2-Methyl-6-phytylhydroquinone; 5-(12-Heneicosenyl)-13-benzenediol; 5-(14-Heneicosenyl)-13-benzenediol; 5-(16-Heneicosenyl)-13-benzenediol; Cholest-5-ene-3-beta7-alpha-diol | HMDB01419; HMDB01496; HMDB02103; HMDB02902; HMDB04035; HMDB06119; HMDB06247; HMDB06283; HMDB06892; HMDB11643; HMDB13643; HMDB29320; HMDB34254; HMDB35681; HMDB38959; HMDB39868; HMDB39869; HMDB39870; HMDB59592 | M-H | - | - |
| C18 negative | 605.2395 | 248.5 | -0.18 (1.1) | 0.08 (1.07) | -0.25 (-0.48, -0.03) | 0.0291 | 0.6181 | Phaeophorbide b | HMDB31149 | M-H | - | - |
| C18 negative | 191.0197 | 20.5 | 0.15 (0.9) | -0.14 (1.45) | 0.28 (0.03, 0.54) | 0.0293 | 0.6181 | Citric acid; Isocitric acid; D-threo-Isocitric acid; Diketogulonic acid; 23-Diketo-L-gulonate; (1R2R)-Isocitric acid; D-Glucaro-14-lactone | HMDB00094; HMDB00193; HMDB01874; HMDB05971; HMDB06511; HMDB33717; HMDB41862 | M-H | Citric acid | M-H |
| C18 negative | 244.1998 | 151.1 | 0.12 (1.03) | -0.12 (1) | 0.24 (0.02, 0.46) | 0.0293 | 0.6181 | 2-Hydroxymyristic acid; (R)-3-Hydroxy-tetradecanoic acid; 3-(L-Menthoxy)-2-methylpropane-12-diol; Ethyl 3-hydroxydodecanoate | HMDB02261; HMDB10731; HMDB32417; HMDB59850 | M-H_[-1] | - | - |
| HILIC positive | 828.5506 | 37.5 | -0.16 (1.34) | 0.11 (0.84) | -0.27 (-0.52, -0.03) | 0.0294 | 0.7973 | PC(18:3(6Z9Z12Z)22:6(4Z7Z10Z13Z16Z19Z)); PC(18:3(9Z12Z15Z)22:6(4Z7Z10Z13Z16Z19Z)); PC(18:4(6Z9Z12Z15Z)22:5(4Z7Z10Z13Z16Z)); PC(18:4(6Z9Z12Z15Z)22:5(7Z10Z13Z16Z19Z)); PC(20:4(5Z8Z11Z14Z)20:5(5Z8Z11Z14Z17Z)); PC(20:4(8Z11Z14Z17Z)20:5(5Z8Z11Z14Z17Z)); PC(20:5(5Z8Z11Z14Z17Z)20:4(5Z8Z11Z14Z)); PC(20:5(5Z8Z11Z14Z17Z)20:4(8Z11Z14Z17Z)); PC(22:5(4Z7Z10Z13Z16Z)18:4(6Z9Z12Z15Z)); PC(22:5(7Z10Z13Z16Z19Z)18:4(6Z9Z12Z15Z)); PC(22:6(4Z7Z10Z13Z16Z19Z)18:3(6Z9Z12Z)); PC(22:6(4Z7Z10Z13Z16Z19Z)18:3(9Z12Z15Z)) | HMDB08189; HMDB08222; HMDB08253; HMDB08254; HMDB08445; HMDB08478; HMDB08509; HMDB08510; HMDB08667; HMDB08700; HMDB08731; HMDB08732 | M+H | - | - |
| C18 negative | 330.2517 | 239 | 0.14 (0.93) | -0.07 (0.9) | 0.21 (0.02, 0.39) | 0.0295 | 0.6181 | Docosapentaenoic acid (22n-6); Docosapentaenoic acid; Ethyl abietate; ent-16-Kauren-19-ol acetate; 1-Phenyl-13-hexadecanedione; 2-Methyl-5-(811-pentadecadienyl)-13-benzenediol; 48121519-Docosapentaenoic acid; Ethyl icosapentate; Docosa-47101316-pentanoic acid | HMDB01976; HMDB06528; HMDB32257; HMDB35024; HMDB35581; HMDB38909; HMDB39133; HMDB39530; HMDB60113 | M-H_[-1] | - | - |
| C18 negative | 157.1234 | 73.2 | -0.29 (1.16) | -0.03 (1.07) | -0.27 (-0.51, -0.03) | 0.0295 | 0.6181 | Oenanthic ether; Pelargonic acid; xi-2-Ethylheptanoic acid; Methyl caprylate; Heptyl acetate; 4-Methyloctanoic acid; Butyl 3-methylbutanoate; 3-Heptyl acetate; 5-Methylhexyl acetate; 3-Octyl formate; Pentyl butanoate; Propyl hexanoate; Hexyl propionate; 3-Methylbutyl 2-methylpropanoate; 1-Hydroxy-3-nonanone; 3-(Hydroxymethyl)-2-octanone; 4-Methyl-2-pentyl-13-dioxolane; 2-Methylpropyl 3-methylbutanoate; Octyl formate; Butyl 2-methylbutanoate; 1-Methylbutyl butanoate; Isoamyl butyrate; Butyl pentanoate; Isopropyl hexanoate; Ethyl 5-methylhexanoate | HMDB00798; HMDB00847; HMDB30331; HMDB31291; HMDB31480; HMDB31557; HMDB32042; HMDB32309; HMDB32406; HMDB32454; HMDB34162; HMDB34165; HMDB34606; HMDB36234; HMDB37143; HMDB37164; HMDB37288; HMDB38040; HMDB38603; HMDB39216; HMDB39219; HMDB40221; HMDB40295; HMDB40430; HMDB59822 | M-H | - | - |
| HILIC positive | 211.1329 | 112 | 0.08 (0.83) | -0.15 (1) | 0.24 (0.02, 0.45) | 0.0299 | 0.7973 | Jasmonic acid; Sedanonic acid; Dihydro-3-(2-octenyl)-25-furandione; (R)-8-Acetoxycarvotanacetone; 1-Benzyloxy-1-(2- | HMDB32797; HMDB32871; HMDB33102; HMDB37104; | M+H | - | - |

| | | | | | | | | | | | | |
|---|---|---|---|---|---|---|---|---|---|---|---|---|
| | | | | | | | | methoxyethoxy)ethane; 4-(Butoxymethyl)-2-methoxyphenol; Isoamyl 2-furonpropionate; Dihydro-3-(1-octenyl)-25-furandione; 3-Ethenyl-25-dimethyl-4-oxohex-5-en-2-yl acetate | HMDB37135; HMDB37642; HMDB37735; HMDB37816; HMDB41573 | | | |
| HILIC positive | 299.258 | 27.2 | -0.22 (0.92) | 0.01 (0.97) | -0.22 (-0.42, -0.02) | 0.0299 | 0.7973 | 3-Oxooctadecanoic acid; MG(0:015:00:0); MG(15:00:00:0); 9-Oxooctadecanoic acid; 10-Oxooctadecanoic acid; 11-Oxooctadecanoic acid; ()-1516-Dihydroxyoctadecanoic acid; 5-Hexyltetrahydro-2-furanoctanoic acid; 5-Oxooctadecanoic acid; Ricinoleic acid; (9S10S)-910-dihydroxyoctadecanoate; Glycerol triundecanoate | HMDB10736; HMDB11532; HMDB11563; HMDB30979; HMDB30980; HMDB30981; HMDB31008; HMDB31127; HMDB34074; HMDB34297; HMDB59633; HMDB31089 | M+H; M+H-H2O; M+2H | - | - |
| HILIC positive | 331.2482 | 27.9 | -0.01 (0.83) | 0.19 (0.68) | -0.2 (-0.37, -0.02) | 0.0302 | 0.7977 | 91213-TriHOME; 91013-TriHOME; 5812-Trihydroxy-9-octadecenoic acid; (9S10E12S13S)-91213-Trihydroxy-10-octadecenoic acid | HMDB04708; HMDB04710; HMDB30936; HMDB38555 | M+H | - | - |
| C18 negative | 100.0404 | 293.2 | 0.1 (0.92) | -0.11 (0.9) | 0.21 (0.02, 0.4) | 0.0303 | 0.6181 | L-3-Aminodihydro-2(3H)-furanone; (S)-2-Azetidinecarboxylic acid; 1-Aminocyclopropanecarboxylic acid | HMDB29387; HMDB29615; HMDB36458 | M-H | - | - |
| C18 negative | 413.3425 | 298.1 | -0.18 (1.1) | 0.05 (1.01) | -0.22 (-0.43, -0.02) | 0.0306 | 0.6181 | (22Alpha)-hydroxy-campest-4-en-3-one; 4alpha-Formyl-5alpha-cholesta-8-en-3beta-ol; 4Alpha-hydroxymethyl-5alpha-cholesta-824-dien-3beta-ol; Ergostane-36-dione; Stenocereol; 1-Phenyl-13-docosanedione; (3beta5alpha7alpha)-14-Methylcholesta-9(11)24-diene-37-diol | HMDB12113; HMDB12169; HMDB12172; HMDB32004; HMDB34329; HMDB35586; HMDB41111 | M-H | - | - |
| C18 negative | 229.1808 | 140.4 | -0.11 (1.04) | 0.14 (0.92) | -0.25 (-0.48, -0.02) | 0.0306 | 0.6181 | 3-5-Methyl-2-(1-methylethyl)cyclohexyloxy-12-propanediol | HMDB36133 | M-H | - | - |
| HILIC positive | 273.0048 | 84.9 | 0.15 (0.99) | -0.07 (1.02) | 0.22 (0.02, 0.41) | 0.0308 | 0.7993 | Zoledronate | HMDB14543 | M+H | - | - |
| HILIC positive | 241.1798 | 240.9 | 0.13 (0.54) | -0.11 (1.24) | 0.23 (0.02, 0.45) | 0.0316 | 0.7993 | L-Menthyl acetoacetate | HMDB32369 | M+H | - | - |
| C18 negative | 388.2575 | 82.1 | -0.1 (1.07) | 0.12 (0.89) | -0.23 (-0.43, -0.02) | 0.0316 | 0.6181 | Cavipetin C | HMDB30366 | M-H_[-1] | - | - |
| C18 negative | 217.0828 | 20.3 | 0.18 (0.66) | -0.09 (1.45) | 0.27 (0.02, 0.52) | 0.0317 | 0.6181 | Glutamylalanine; 5-L-Glutamyl-L-alanine; Hydroxyprolyl-Serine; Serinyl-Hydroxyproline; N-gamma-L-Glutamyl-D-alanine | HMDB03764; HMDB06248; HMDB28872; HMDB29040; HMDB36301 | M-H | - | - |
| C18 negative | 207.0145 | 19.4 | 0.15 (0.87) | -0.1 (1.11) | 0.25 (0.02, 0.47) | 0.0318 | 0.6181 | Garcinia acid | HMDB31159 | M-H | - | - |
| C18 negative | 573.4522 | 270.2 | -0.14 (1.07) | 0.07 (0.92) | -0.21 (-0.4, -0.02) | 0.0318 | 0.6181 | DG(15:018:4(6Z9Z12Z15Z)0:0); DG(18:4(6Z9Z12Z15Z)15:00:0); DG(15:00:018:4n3) | HMDB07077; HMDB07329; HMDB56004 | M-H | - | - |
| C18 negative | 135.0663 | 158.7 | 0 (0.93) | 0.16 (0.69) | -0.17 (-0.32, -0.01) | 0.0319 | 0.6181 | 2-Methylerythritol ; 3-Deoxy-D-arabinitol; 2-Deoxy-D-ribitol; 1-Deoxy-D-ribitol | HMDB11659; HMDB31201; HMDB33919; HMDB41486 | M-H | - | - |
| HILIC positive | 223.1329 | 254.7 | -0.18 (1.33) | 0.07 (0.72) | -0.26 (-0.5, -0.02) | 0.0319 | 0.7993 | Annuionone B; Dehydrovomifoliol; Isoamyl p-anisate; 1-Hydroxyibuprofen; 2-Hydroxyibuprofen; 3-Hydroxyibuprofen | HMDB32121; HMDB36819; HMDB38894; HMDB60565; HMDB60920; HMDB60921 | M+H | - | - |
| HILIC positive | 188.0918 | 97.6 | 0.11 (0.63) | -0.07 (0.91) | 0.19 (0.02, 0.35) | 0.0320 | 0.7993 | 2-Keto-6-acetamidocaproate | HMDB12150 | M+H | - | - |
| HILIC positive | 136.0618 | 133.3 | 0.09 (0.72) | -0.11 (1.02) | 0.2 (0.02, 0.39) | 0.0321 | 0.7993 | Adenine | HMDB00034 | M+H | - | - |
| C18 negative | 125.0245 | 23.3 | -0.31 (1.31) | -0.01 (1.02) | -0.3 (-0.57, -0.03) | 0.0321 | 0.6181 | 123-Trihydroxybenzene; 135-Trihydroxybenzene; Methyl 2-furoate; Maltol; 4-Hydroxy-2H-pyran-3-carboxaldehyde; (Z)-Tamarindienal; 5-Hydroxymethyl-2-furancarboxaldehyde; 5-Methylfuran-2-carboxylic acid | HMDB13674; HMDB13675; HMDB29750; HMDB30776; HMDB31256; HMDB32651; HMDB34355; HMDB59735 | M-H | - | - |
| C18 negative | 227.2015 | 225 | 0.09 (1.04) | -0.15 (1.09) | 0.24 (0.02, 0.47) | 0.0325 | 0.6181 | Myristic acid; 2610-Trimethylundecanoic acid; 12-Methyltridecanoic acid; Hexanal octane-13-diol acetal; Undecanal propyleneglycol acetal; Ethyl dodecanoate; Heptyl heptanoate; Hexyl octanoate; Decyl butanoate; 3-Methylbutyl nonanoate; Nonyl isovalerate; Dodecyl acetate; Isobutyl decanoate | HMDB00806; HMDB02221; HMDB31072; HMDB32318; HMDB32548; HMDB33788; HMDB34461; HMDB36216; HMDB37312; HMDB37495; HMDB38034; HMDB38969; HMDB59865 | M-H | Myristic acid | M-H |
| C18 negative | 199.1703 | 189.8 | -0.02 (1.1) | -0.28 (1.21) | 0.26 (0.02, 0.5) | 0.0329 | 0.6181 | Dodecanoic acid; Ethyl decanoate; 2-Heptyl butyrate; Nonanal propyleneglycol acetal; Hexyl hexanoate; Isopropyl nonanoate; Octyl butanoate; Octyl 2-methylpropanoate; (R)-Dihydrocitronellol acetate; Pentyl heptanoate; Decyl acetate; Isobutyl octanoate | HMDB00638; HMDB30998; HMDB32310; HMDB32440; HMDB33619; HMDB34128; HMDB34136; HMDB34137; HMDB35410; HMDB36225; HMDB40166; HMDB59868 | M-H | Lauric acid | M-H |
| C18 negative | 819.7434 | 40.8 | 0.3 (0.5) | 0.14 (0.73) | 0.15 (0.01, 0.29) | 0.0333 | 0.6181 | TG(14:015:020:0); TG(14:020:015:0); TG(15:014:020:0); TG(15:016:018:0); TG(15:018:016:0); TG(16:015:018:0) | HMDB42095; HMDB42182; HMDB42963; HMDB43020; HMDB43048; HMDB43858 | M-H | - | - |
| C18 negative | 138.0196 | 191.2 | -0.11 (1.17) | 0.13 (0.63) | -0.24 (-0.46, -0.02) | 0.0338 | 0.6181 | 4-Nitrophenol; 6-Hydroxynicotinic acid; 3-Hydroxypicolinic acid; 2-Hydroxynicotinic acid | HMDB01232; HMDB02658; HMDB13188; HMDB59710 | M-H | - | - |
| HILIC positive | 213.0747 | 79.2 | 0.22 (0.91) | -0.01 (1.01) | 0.23 (0.02, 0.44) | 0.0341 | 0.8026 | Vanillactic acid; Danielone; Propyl gallate; Eudesmic acid; Valtrate; 23-Dihydroxy-1-(4-hydroxy-3-methoxyphenyl)-1-propanone; 34-Dihydroxyphenyllactic acid methyl ester; 3-(34-Dihydroxyphenyl)-2-methoxypropionic acid; 3-Hydroxy-4-methoxyphenyllactic acid; beta-(2-Methoxyphenoxy)-lactic acid | HMDB00913; HMDB31704; HMDB33835; HMDB33839; HMDB34493; HMDB41286; HMDB41661; HMDB41663; HMDB41665; HMDB60557 | M+H | - | - |
| C18 negative | 306.2427 | 233.9 | 0.06 (0.87) | -0.2 (1.29) | 0.26 (0.02, 0.49) | 0.0343 | 0.6181 | Arachidonic acid; Cis-8111417-Eicosatetraenoic acid; Mesterolone; Drostanolone; Sideridiol; ent-17-Hydroxy- | HMDB01043; HMDB02177; HMDB06036; HMDB14996; | M-H_[+3] | - | - |

| | | | | | | | | | | | | |
|---|---|---|---|---|---|---|---|---|---|---|---|---|
| | | | | | | | | 16beta-kauran-19-al; Yucalexin P21; Copalic acid; Junicedral; 713-Eperudien-15-oic acid; Oryzalexin S; Oryzalexin E | HMDB36702; HMDB36721; HMDB36754; HMDB36829; HMDB36830; HMDB37824; HMDB39687; HMDB39702 | | | |
| HILIC positive | 432.392 | 27.1 | 0.09 (1.02) | -0.13 (0.97) | 0.22 (0.02, 0.42) | 0.0351 | 0.8026 | Alpha-Tocopherol; 4alpha-Hydroxymethyl-4beta-methyl-5alpha-cholesta-8-en-3beta-ol; Ikshusterol; 4-beta-Hydroxymethyl-4-alpha-methyl-5-alpha-cholest-7-en-3-beta-ol | HMDB01893; HMDB12171; HMDB30022; HMDB59642 | M+H_[+1] | - | - |
| C18 negative | 285.2263 | 152.8 | 0.14 (1.01) | -0.09 (0.96) | 0.23 (0.02, 0.44) | 0.0352 | 0.6181 | Myristoylglycine | HMDB13250 | M-H_[-1] | - | - |
| HILIC positive | 395.1176 | 100.8 | 0.25 (0.52) | 0.11 (0.79) | 0.14 (0.01, 0.28) | 0.0359 | 0.8083 | Garcimangosone D | HMDB38066 | M+H_[+2] | - | - |
| C18 negative | 176.0928 | 295.7 | 0.11 (0.76) | -0.08 (0.83) | 0.19 (0.01, 0.37) | 0.0366 | 0.6181 | N-Methyl-1-deoxynojirimycin | HMDB35360 | M-H | - | - |
| C18 negative | 471.3485 | 237 | -0.11 (0.99) | 0.1 (0.75) | -0.21 (-0.41, -0.01) | 0.0366 | 0.6181 | Maslinic acid; Lucidumol A; Queretaroic acid; Priverogenin A; (3beta23xi)-323-Dihydroxycycloart-24-en-26-oic acid; Pomolic acid; 20beta-Hydroxyursolic acid; Azukisapogenol; delta-Maslinic acid; Sebiferenic acid; Momordicin I; Ganoderic acid U; Ganodermanontriol; Epoxyganoderiol A; (3b22S24E)-322-Dihydroxycycloart-24-en-26-oic acid; 27-Hydroxyisomangiferolic acid; Albigenic acid; Rubitic acid; Ganoderiol E; 3alpha-Corosolic acid | HMDB02392; HMDB33233; HMDB34523; HMDB34530; HMDB34567; HMDB35106; HMDB35107; HMDB35258; HMDB35260; HMDB35748; HMDB35758; HMDB35962; HMDB35977; HMDB36001; HMDB36063; HMDB36064; HMDB36638; HMDB36654; HMDB37779; HMDB41043 | M-H | - | - |
| HILIC positive | 172.0718 | 108.5 | 0.08 (0.86) | -0.14 (1.07) | 0.23 (0.01, 0.44) | 0.0370 | 0.8083 | Metronidazole | HMDB15052 | M+H | - | - |
| HILIC positive | 172.1332 | 41.4 | -0.07 (1.14) | -0.28 (1.08) | 0.21 (0.01, 0.4) | 0.0371 | 0.8083 | Gabapentin | HMDB05015 | M+H | - | - |
| C18 negative | 847.7781 | 41.2 | -0.15 (1.33) | 0.09 (0.91) | -0.24 (-0.47, -0.01) | 0.0373 | 0.6181 | Glycerol triheptadecanoate; TG(14:015:022:0); TG(14:022:015:0); TG(15:014:022:0); TG(15:016:020:0); TG(15:018:018:0); TG(15:020:016:0); TG(16:015:020:0); TG(18:015:018:0) | HMDB31106; HMDB42096; HMDB42212; HMDB42964; HMDB43021; HMDB43049; HMDB43077; HMDB43859; HMDB44695 | M-H | - | - |
| C18 negative | 279.1966 | 140.2 | -0.14 (1.12) | 0.1 (0.89) | -0.24 (-0.47, -0.01) | 0.0374 | 0.6181 | 12S-HHT; Dihydropanaxacol; Fauronyl acetate; Valeracetate | HMDB12535; HMDB32675; HMDB36422; HMDB41388 | M-H | - | - |
| C18 negative | 219.1753 | 224.6 | -0.06 (0.64) | 0.13 (1) | -0.19 (-0.37, -0.01) | 0.0375 | 0.6181 | Nootkatol; (2R6S7S10Z)-beta-Santala-3(15)10-dien-12-ol; Dihydronootkatone; 26-Di-tert-butyl-4-methylphenol; alpha-Valerenol; Isospathulenol; alpha-Santal-10-en-12-ol; Acolamone; alpha-Cyperol; beta-Costol; Epishyobunone; Preisocalamendiol; Epoxyguaiene; Acorenone; Isocyperol; Isoacolamone; (R)-27(14)9-Bisabolatrien-11-ol; Fukinone; (3S6E)-6-Caryophyllen-15-al; Apritone; alpha-Bergamotenol; Spathulenol; beta-Santalol; trans-beta-Santalol; beta-Betulenol; Caryophyllene alpha-oxide; 315-Epoxy-6-caryophyllene; 8alpha-3-Copaen-8-ol; Acoragermacrone; 4(15)-Copaen-11-ol; 11-Copaen-4-ol; Vetiverol; Eremofukinone; 15-Epoxy-4(14)-salvialene; Bisabolene oxide; Calacone; 7(14)-Isodaucen-10-one; Isoshyobunone; Humuladienone; Humulene epoxide I; Humulene epoxide II; Humulenol I; Humulenol II; Bisacurol; 4-Nonylphenol; (6Z9Z12Z)-6912-Pentadecatrien-2-one; Aromadendrene epoxide; Italicene ether; Cabreuva oxide D; 2-trans6-trans-Farnesal | HMDB13688; HMDB30232; HMDB32220; HMDB33826; HMDB34661; HMDB34718; HMDB34940; HMDB35020; HMDB35026; HMDB35097; HMDB35306; HMDB35391; HMDB35645; HMDB35704; HMDB35718; HMDB35720; HMDB35739; HMDB35793; HMDB36118; HMDB36192; HMDB36402; HMDB36420; HMDB36716; HMDB36717; HMDB36788; HMDB36789; HMDB36793; HMDB36798; HMDB37069; HMDB37392; HMDB37395; HMDB37811; HMDB38119; HMDB38123; HMDB38134; HMDB38146; HMDB38160; HMDB38193; HMDB38208; HMDB38209; HMDB38210; HMDB38211; HMDB38212; HMDB38512; HMDB38982; HMDB39532; HMDB39711; HMDB40763; HMDB59836; HMDB60356 | M-H | - | - |
| HILIC positive | 536.4305 | 26.5 | -0.35 (1.02) | -0.11 (0.98) | -0.24 (-0.47, -0.01) | 0.0378 | 0.8105 | Ganoderiol C | HMDB37781 | M+NH4 | - | - |
| C18 negative | 425.2009 | 102.3 | -0.17 (1.1) | 0.08 (0.92) | -0.25 (-0.48, -0.01) | 0.0379 | 0.6181 | Paliperidone | HMDB15396 | M-H | - | - |
| HILIC positive | 413.3624 | 223.7 | -0.11 (1.2) | 0.12 (0.76) | -0.23 (-0.46, -0.01) | 0.0380 | 0.8105 | MG(0:022:1(13Z)0:0); MG(22:1(13Z)0:00:0) | HMDB11552; HMDB11582 | M+H | - | - |
| C18 negative | 353.2333 | 152 | 0.11 (1) | -0.15 (1.15) | 0.26 (0.01, 0.51) | 0.0382 | 0.6181 | Prostaglandin F2a; Prostaglandin E1; Prostaglandin F2b; Troxilin B3; Trioxilin A3; 8-isoprostaglandin PGF2b; 111215-THETA; 1314-Dihydro-15-keto PGF2a; 8-Isoprostaglandin E1; 111415-THETA; 8-Isoprostaglandin F2a; Prostaglandin D1; 8-iso-1314-dihydro-15-keto-PGF2a; 111215-TriHETRE; 11b-PGF2a; 11-Epi-PGF2a; 1314-Dihydro- lipoxin A4; Prostaglandin H1; Sterebin G; 15-keto-PGF1alpha; (5Z9E12S14Z)-81112-Trihydroxyicosa-5914-trienoate | HMDB01139; HMDB01442; HMDB01483; HMDB01965; HMDB01977; HMDB02115; HMDB04684; HMDB04685; HMDB04686; HMDB04694; HMDB05083; HMDB05102; HMDB06562; HMDB10170; HMDB10199; HMDB11137; HMDB12563; HMDB13041; | M-H | - | - |

| | | | | | | | | | | | | |
|---|---|---|---|---|---|---|---|---|---|---|---|---|
| | | | | | | | | | HMDB35380; HMDB60045; HMDB60107 | | | |
| C18 negative | 273.0076 | 19.8 | -0.55 (0.91) | -0.31 (1.06) | -0.23 (-0.45, -0.01) | 0.0383 | 0.6181 | Ferulic acid 4-sulfate; Isoferulic acid 3-sulfate | HMDB29200; HMDB41748 | M-H | - | - |
| C18 negative | 491.3378 | 179.4 | -0.09 (0.96) | 0.08 (0.94) | -0.17 (-0.34, -0.01) | 0.0384 | 0.6181 | Homodolicholide | HMDB33907 | M-H | - | - |
| C18 negative | 433.3326 | 235.9 | -0.28 (1.14) | -0.05 (0.94) | -0.23 (-0.45, -0.01) | 0.0389 | 0.6181 | 3a7a-Dihydroxycoprostanic acid; 3a7a12a-Trihydroxy-5b-cholestan-26-al | HMDB00359; HMDB03533 | M-H | - | - |
| C18 negative | 277.1447 | 294.5 | 0.11 (0.95) | -0.1 (1.08) | 0.22 (0.01, 0.42) | 0.0395 | 0.6181 | Alpha-CEHC; Monoethylhexyl phthalic acid; Diisobutyl phthalate; Dibutyl phthalate | HMDB01518; HMDB13248; HMDB13835; HMDB33244 | M-H | - | - |
| C18 negative | 377.2098 | 237.1 | 0.55 (0.66) | 0.38 (0.76) | 0.17 (0.01, 0.34) | 0.0397 | 0.6181 | Neotame | HMDB34566 | M-H | - | - |
| HILIC positive | 110.027 | 73.5 | -0.13 (1.05) | 0.06 (0.84) | -0.19 (-0.37, -0.01) | 0.0397 | 0.8113 | Hypotaurine | HMDB00965 | M+H | Hypotauri ne | M+H |
| HILIC positive | 265.216 | 26.6 | -0.2 (1.11) | 0.06 (1.11) | -0.26 (-0.5, -0.01) | 0.0399 | 0.8113 | Cedryl acetate; Shyobunyl acetate; Guaiol acetate; alpha-Caryophyllene alcohol acetate; beta-Caryophyllene alcohol acetate; Nerolidyl acetate | HMDB35910; HMDB36153; HMDB37218; HMDB37224; HMDB37273; HMDB39630 | M+H | - | - |
| HILIC positive | 519.4039 | 27.2 | -0.21 (1.1) | 0 (0.89) | -0.21 (-0.42, -0.01) | 0.0399 | 0.8113 | Ganoderiol C | HMDB37781 | M+H | - | - |
| HILIC positive | 151.024 | 90.6 | -0.01 (0.91) | 0.19 (0.97) | -0.2 (-0.39, -0.01) | 0.0402 | 0.8113 | Tartaric acid; D-Tartaric acid; 2-Methyl-2-(methyldithio)propanal; 23-Dihydroxybutanedioic acid | HMDB00956; HMDB29878; HMDB38889; HMDB59916 | M+H | - | - |
| C18 negative | 158.057 | 289 | 0.1 (0.87) | -0.12 (0.99) | 0.22 (0.01, 0.43) | 0.0403 | 0.6181 | L-Hexahydro-3-imino-124-oxadiazepine-3-carboxylic acid | HMDB30402 | M-H | - | - |
| HILIC positive | 783.5714 | 30.2 | 0.11 (0.96) | -0.07 (0.93) | 0.18 (0.01, 0.36) | 0.0403 | 0.8113 | PC(14:022:4(7Z10Z13Z16Z)); PC(16:020:4(5Z8Z11Z14Z)); PC(16:020:4(8Z11Z14Z17Z)); PC(16:1(9Z)20:3(5Z8Z11Z)); PC(16:1(9Z)20:3(8Z11Z14Z)); PC(18:018:4(6Z9Z12Z15Z)); PC(18:1(11Z)18:3(6Z9Z12Z)); PC(18:1(11Z)18:3(9Z12Z15Z)); PC(18:1(9Z)18:3(6Z9Z12Z)); PC(18:1(9Z)18:3(9Z12Z15Z)); PC(18:2(9Z12Z)18:2(9Z12Z)); PC(18:3(6Z9Z12Z)18:1(11Z)); PC(18:3(6Z9Z12Z)18:1(9Z)); PC(18:3(9Z12Z15Z)18:1(11Z)); PC(18:3(9Z12Z15Z)18:1(9Z)); PC(18:4(6Z9Z12Z15Z)18:0); PC(20:3(5Z8Z11Z)16:1(9Z)); PC(20:3(8Z11Z14Z)16:1(9Z)); PC(20:4(5Z8Z11Z14Z)16:0); PC(20:4(8Z11Z14Z17Z)16:0); PC(22:4(7Z10Z13Z16Z)14:0) | HMDB07889; HMDB07982; HMDB07983; HMDB08013; HMDB08014; HMDB08042; HMDB08073; HMDB08074; HMDB08106; HMDB08107; HMDB08138; HMDB08169; HMDB08170; HMDB08202; HMDB08203; HMDB08234; HMDB08365; HMDB08398; HMDB08429; HMDB08462; HMDB08623 | M+H_[+1] | - | - |
| C18 negative | 653.5197 | 22.8 | -0.27 (1.11) | -0.02 (0.96) | -0.25 (-0.49, -0.01) | 0.0405 | 0.6181 | SM(d18:012:0) | HMDB12084 | M-H_[+4] | - | - |
| C18 negative | 352.2572 | 244.8 | 0.1 (0.87) | -0.13 (1.13) | 0.23 (0.01, 0.45) | 0.0411 | 0.6181 | MG(0:018:3(6Z9Z12Z)0:0); MG(0:018:3(9Z12Z15Z)0:0); MG(18:3(6Z9Z12Z)0:00:0); MG(18:3(9Z12Z15Z)0:00:0) | HMDB11539; HMDB11540; HMDB11569; HMDB11570 | M-H_[-1] | - | - |
| HILIC positive | 276.0431 | 63.8 | -0.05 (0.81) | 0.12 (0.84) | -0.17 (-0.34, -0.01) | 0.0412 | 0.8113 | N-(2-formyl-3-chlorophenyl)anthranilic acid | HMDB60006 | M+H | - | - |
| HILIC positive | 124.0757 | 288.2 | 0.11 (1.03) | -0.11 (0.98) | 0.22 (0.01, 0.43) | 0.0413 | 0.8113 | p-Anisidine; 2-Propionylpyrrole; 4-Hydroxybenzylamine; 1-Ethyl-1H-pyrrole-2-carboxaldehyde; 2-Acetyl-1-methylpyrrole | HMDB29300; HMDB32492; HMDB32578; HMDB36058; HMDB40360 | M+H | - | - |
| C18 negative | 261.0075 | 19.9 | -0.5 (1.09) | -0.26 (1.04) | -0.24 (-0.47, -0.01) | 0.0417 | 0.6181 | Homovanillic acid sulfate; 2-Hydroxy-4-methoxyacetophenone 5-sulfate; Dihydrocaffeic acid 3-sulfate; 3-hydroxy-3-(3-hydroxyphenyl)propanoic acid-O-sulphate; 3-(35-dihydroxyphenyl)-1-propanoic acid sulphate | HMDB11719; HMDB41648; HMDB41721; HMDB59967; HMDB61117 | M-H | - | - |
| C18 negative | 243.1235 | 295.2 | 0.09 (0.77) | -0.18 (1.46) | 0.27 (0.01, 0.52) | 0.0417 | 0.6181 | Polyethylene oxidized | HMDB32472 | M-H | - | - |
| HILIC positive | 331.2637 | 27.2 | 0.05 (1.05) | -0.19 (1.17) | 0.24 (0.01, 0.46) | 0.0424 | 0.8113 | Docosapentaenoic acid (22n-6); Docosapentaenoic acid; Ethyl abietate; ent-16-Kauren-19-ol acetate; 1-Phenyl-13-hexadecanedione; 2-Methyl-5-(811-pentadecadienyl)-13-benzenediol; 48121519-Docosapentaenoic acid; Ethyl icosapentate; Docosa-47101316-pentaenoic acid | HMDB01976; HMDB06528; HMDB32257; HMDB35024; HMDB35581; HMDB38909; HMDB39133; HMDB39530; HMDB60113 | M+H | - | - |
| C18 negative | 284.2229 | 152.6 | 0.13 (0.95) | -0.07 (0.88) | 0.2 (0.01, 0.4) | 0.0424 | 0.6181 | Myristoylglycine | HMDB13250 | M-H | - | - |
| C18 negative | 297.2797 | 286.8 | 0.12 (1) | -0.11 (1.07) | 0.22 (0.01, 0.44) | 0.0424 | 0.6181 | Nonadecanoic acid; Pristanic acid; Tuberculostearic acid; Methyl stearate; Isopropyl hexadecanoate; 4-Hydroxy-6-nonadecanone; 6-Hydroxy-4-nonadecanone; xi-17-Methyloctadecanoic acid; xi-16-Methyloctadecanoic acid | HMDB00772; HMDB00795; HMDB04085; HMDB34154; HMDB35474; HMDB35674; HMDB35675; HMDB37397; HMDB38070 | M-H | - | - |
| C18 negative | 316.9477 | 40.3 | -0.25 (1.03) | -0.04 (0.85) | -0.21 (-0.41, -0.01) | 0.0425 | 0.6181 | Mitotane | HMDB14786 | M-H | - | - |
| HILIC positive | 211.1433 | 117.1 | 0.1 (0.75) | -0.12 (1.19) | 0.22 (0.01, 0.43) | 0.0428 | 0.8113 | LL-Cyclo(leucylprolyl) | HMDB34276 | M+H | - | - |
| HILIC positive | 135.0441 | 294.7 | -0.1 (1.33) | 0.13 (0.41) | -0.23 (-0.45, -0.01) | 0.0429 | 0.8113 | Phthalide | HMDB32469 | M+H | - | - |
| C18 negative | 347.1862 | 147 | -0.16 (1.33) | 0.07 (0.9) | -0.23 (-0.46, -0.01) | 0.0430 | 0.6181 | Gibberellin A110; Gibberellin A112; 3b-Hydroxy-6b-angeloyloxy-7(11)-eremophilen-128b-olide; Cohumulone; Bakkenolide C; Gibberellin A53; 3b-Hydroxy-6b-tigloyloxy-7(11)-eremophilen-128b-olide; 1220-Dioxo-leukotriene B4 | HMDB32010; HMDB32011; HMDB32895; HMDB33981; HMDB34999; HMDB36895; HMDB40989; HMDB60094 | M-H | - | - |
| HILIC positive | 248.1129 | 227.1 | -0.08 (1.03) | 0.12 (0.81) | -0.19 (-0.38, -0.01) | 0.0430 | 0.8113 | Malonylcarnitine; Linamarin | HMDB02095; HMDB33699 | M+H | - | - |
| HILIC positive | 241.1061 | 70.2 | 0.14 (0.77) | -0.12 (1.46) | 0.27 (0.01, 0.53) | 0.0430 | 0.8113 | 3-(345-Trimethoxyphenyl)propanoic acid; Ethyl 345-trimethoxybenzoate; Isopropyl 3-(34-dihydroxyphenyl)-2-hydroxypropanoate; 3-Carboxy-4-methyl-5-propyl-2-furanpropionic acid | HMDB30254; HMDB38627; HMDB41756; HMDB61112 | M+H | - | - |

| | | | | | | | | | | | | |
|---|---|---|---|---|---|---|---|---|---|---|---|---|
| HILIC positive | 157.1587 | 296.2 | 0.09 (0.81) | -0.14 (1.29) | 0.23 (0.01, 0.46) | 0.0433 | 0.8113 | Menthol; Decanal; (E)-3-decen-1-ol; 2-Decanone; cis-4-Decenol; 3-Decanone; 37-Dimethyloctanal; (-)-4-Ethyloctanal; 2-DECENOL; Menthanol; D-Citronellol; L-Citronellol; p-Menthan-4-ol; ()-Neomenthol; (-)-Neoisomenthol; p-Menthan-3-ol; p-Menthan-1-ol; alpha-Citronellol; ()-Carvomenthol; Dihydrocitronellal; 1-Decen-3-ol; ()-Neoisomenthol; 9-Decenol | HMDB03352; HMDB11623; HMDB13810; HMDB31409; HMDB32206; HMDB32212; HMDB32241; HMDB32274; HMDB32532; HMDB34717; HMDB35093; HMDB35094; HMDB35726; HMDB35763; HMDB35764; HMDB35765; HMDB37020; HMDB37171; HMDB37223; HMDB37806; HMDB39854; HMDB41628; HMDB59861 | M+H | - | - |
| C18 negative | 513.3026 | 182.9 | 0.09 (0.97) | -0.13 (1.07) | 0.22 (0.01, 0.44) | 0.0434 | 0.6181 | Flavidulol C | HMDB39150 | M-H | - | - |
| C18 negative | 211.1703 | 180.5 | 0.15 (0.94) | -0.06 (0.98) | 0.21 (0.01, 0.41) | 0.0434 | 0.6181 | 10-Undecenyl acetate; Methyl (E)-2-dodecenoate; Coniferan; ()-(E)-3-Methyl-4-decen-1-yl acetate; Ethyl 10-undecenoate; Rhodinyl propionate; Citronellyl propionate; 1-Ethylhexyl tiglate; 261010-Tetramethyl-1-oxaspiro45decan-6-ol; 11-Diethoxy-26-nonadiene; 4-(3-Hydroxybutyl)-335-trimethylcyclohexanone; Oxacyclotetradecan-2-one | HMDB31017; HMDB31028; HMDB31741; HMDB32792; HMDB34286; HMDB37187; HMDB37226; HMDB37626; HMDB37907; HMDB38078; HMDB39805; HMDB40451 | M-H | - | - |
| C18 negative | 771.5178 | 224.5 | -0.28 (1.24) | -0.03 (1) | -0.25 (-0.5, -0.01) | 0.0436 | 0.6181 | PG(16:020:3(5Z8Z11Z)); PG(16:020:3(8Z11Z14Z)); PG(18:018:3(6Z9Z12Z)); PG(18:018:3(9Z12Z15Z)); PG(18:1(11Z)18:2(9Z12Z)); PG(18:1(9Z)18:2(9Z12Z)); PG(18:2(9Z12Z)18:1(11Z)); PG(18:2(9Z12Z)18:1(9Z)); PG(18:3(6Z9Z12Z)18:0); PG(18:3(9Z12Z15Z)18:0) | HMDB10578; HMDB10579; HMDB10606; HMDB10607; HMDB10620; HMDB10635; HMDB10648; HMDB10649; HMDB10662; HMDB10677 | M-H | - | - |
| HILIC positive | 257.2474 | 294.9 | 0.11 (0.81) | -0.11 (1.13) | 0.22 (0.01, 0.44) | 0.0440 | 0.8113 | Palmitic acid; Trimethyltridecanoic acid; Isopalmitic acid; Butyl dodecanoate; Dodecyl butyrate; Hexyl decanoate; Octyl octanoate; Ethyl tetradecanoate; Dodecyl 2-methylpropanoate; Hexadecanoate (n-C16:0) | HMDB00220; HMDB02396; HMDB31068; HMDB32065; HMDB32249; HMDB32324; HMDB33166; HMDB34153; HMDB36210; HMDB60083 | M+H | - | - |
| HILIC positive | 508.3762 | 36.8 | -0.24 (1.35) | 0.03 (0.77) | -0.26 (-0.52, -0.01) | 0.0441 | 0.8113 | LysoPC(P-18:0) | HMDB13122 | M+H | - | - |
| C18 negative | 145.0327 | 293.1 | 0.19 (0.94) | -0.03 (1.03) | 0.21 (0.01, 0.42) | 0.0443 | 0.6181 | Dihydro-2-methoxy-2-methyl-3(2H)-thiophenone | HMDB40237 | M-H | - | - |
| HILIC positive | 88.0757 | 94.8 | -0.12 (1.35) | 0.11 (0.44) | -0.22 (-0.44, -0.01) | 0.0443 | 0.8113 | 4-Aminobutyraldehyde; N-Ethylacetamide; Morpholine; Butyramide; 4-Ammoniobutanal | HMDB01080; HMDB31215; HMDB31581; HMDB33870; HMDB60247 | M+H | - | - |
| C18 negative | 319.2278 | 184.5 | 0.61 (0.09) | 0.63 (0.12) | -0.02 (-0.04, 0) | 0.0447 | 0.6181 | 56-Epoxy-81114-eicosatrienoic acid; 89-Epoxyeicosatrienoic acid; 14R15S-EpETrE; 15(S)-HETE; 1415-Epoxy-5811-eicosatrienoic acid; 1112-Epoxyeicosatrienoic acid; 8-HETE; 16(R)-HETE; 11(R)-HETE; 20-Hydroxyeicosatetraenoic acid; 12-HETE; 18-Hydroxyarachidonic acid; 9-HETE; 1112-EpETrE; 5-HETE; 19(S)-HETE; 10-HETE; 13-HETE; 17-HETE; Annoglabasin E; Ucriol; 8alpha13R-Epoxy-14-labden-19-oic acid; 2-Hydroxy-6-tridecylbenzoic acid; (ent-2alpha3beta15beta16beta)-1516-Epoxy-23-kauranediol; (ent-2alpha3beta15beta)-16-Kaurene-2315-triol; Crispane; 12 Hydroxy arachidonic acid; Arachidonate | HMDB02190; HMDB02232; HMDB02283; HMDB03876; HMDB04264; HMDB04673; HMDB04679; HMDB04680; HMDB04682; HMDB05998; HMDB06111; HMDB06245; HMDB10222; HMDB10409; HMDB11134; HMDB11136; HMDB12508; HMDB12567; HMDB12598; HMDB36262; HMDB36705; HMDB36810; HMDB38520; HMDB38688; HMDB38689; HMDB38694; HMDB60101; HMDB60102 | M-H | - | - |
| HILIC positive | 537.1653 | 26.7 | 0.08 (0.9) | -0.17 (1.38) | 0.25 (0.01, 0.5) | 0.0447 | 0.8113 | 14-beta-D-Glucan | HMDB06944 | M+H | - | - |
| C18 negative | 378.2729 | 251.2 | 0.11 (1) | -0.08 (0.93) | 0.2 (0, 0.39) | 0.0451 | 0.6181 | 2-Arachidonylglycerol; MG(0:020:4(8Z11Z14Z17Z)0:0); MG(20:4(5Z8Z11Z14Z)0:00:0); MG(20:4(8Z11Z14Z17Z)0:00:0); 1-Acetoxy-2-hydroxy-51215-heneicosatrien-4-one; 12-Gingerol; Persenone A | HMDB04666; HMDB11549; HMDB11578; HMDB11579; HMDB35273; HMDB36356; HMDB36568 | M-H_[-1] | - | - |
| C18 negative | 331.255 | 239.5 | 0.14 (0.88) | -0.05 (0.9) | 0.19 (0, 0.37) | 0.0453 | 0.6181 | Docosapentaenoic acid (22n-6); Docosapentaenoic acid; Ethyl abietate; ent-16-Kauren-19-ol acetate; 1-Phenyl-13-hexadecanedione; 2-Methyl-5-(811-pentadecadienyl)-13-benzenediol; 48121519-Docosapentaenoic acid; Ethyl icosapentate; Docosa-47101316-pentaenoic acid | HMDB01976; HMDB06528; HMDB32257; HMDB35024; HMDB35581; HMDB38909; HMDB39133; HMDB39530; HMDB60113 | M-H_[+2] | - | - |
| HILIC positive | 966.3437 | 54.2 | -0.09 (0.7) | 0.06 (0.74) | -0.16 (-0.31, 0) | 0.0453 | 0.8113 | Disialosyl galactosyl globoside | HMDB06588 | M+H | - | - |
| HILIC positive | 311.257 | 23.6 | 0.1 (0.81) | -0.08 (1.02) | 0.18 (0, 0.36) | 0.0460 | 0.8113 | (R)-2-Hydroxysterculic acid | HMDB31060 | M+H | - | - |
| HILIC positive | 731.5411 | 31.4 | 0.16 (0.97) | -0.06 (0.98) | 0.22 (0, 0.43) | 0.0463 | 0.8113 | PC(14:018:2(9Z12Z)); PC(14:1(9Z)18:1(11Z)); PC(14:1(9Z)18:1(9Z)); PC(16:1(9Z)16:1(9Z)); PC(18:1(11Z)14:1(9Z)); PC(18:1(9Z)14:1(9Z)); PC(18:2(9Z12Z)14:0); PE(15:020:2(11Z14Z)); PE(20:2(11Z14Z)15:0) | HMDB07874; HMDB07905; HMDB07906; HMDB08002; HMDB08065; HMDB08098; HMDB08130; HMDB08901; HMDB09285 | M+H_[+1] | - | - |

| | | | | | | | | | | | | |
|---|---|---|---|---|---|---|---|---|---|---|---|---|
| HILIC positive | 373.2736 | 29.8 | -0.37 (1.06) | -0.13 (1.05) | -0.24 (-0.48, 0) | 0.0469 | 0.8113 | Cervonoyl ethanolamide; Nabilone; (3R 6Z)-34-Dihydro-8-hydroxy-3-(6-pentadecenyl)-1H-2-benzopyran-1-one | HMDB13627; HMDB14629; HMDB41301 | M+H | - | - |
| C18 negative | 225.113 | 293.7 | 0.11 (0.75) | -0.08 (0.93) | 0.19 (0, 0.37) | 0.0469 | 0.6181 | Allixin; 34-Methylenesebacic acid | HMDB40705; HMDB59729 | M-H | - | - |
| C18 negative | 427.3065 | 252.4 | -0.21 (1.06) | -0.45 (1.06) | 0.24 (0, 0.47) | 0.0471 | 0.6181 | Sorbitan oleate; Glyceryl lactooleate | HMDB29886; HMDB32298 | M-H | - | - |
| HILIC positive | 782.5684 | 30.4 | 0.11 (0.96) | -0.07 (0.92) | 0.18 (0, 0.35) | 0.0472 | 0.8113 | PC(14:022:4(7Z10Z13Z16Z)); PC(16:020:4(5Z8Z11Z14Z)); PC(16:020:4(8Z11Z14Z17Z)); PC(16:1(9Z)20:3(5Z8Z11Z)); PC(16:1(9Z)20:3(8Z11Z14Z)); PC(18:018:4(6Z9Z12Z15Z)); PC(18:1(11Z)18:3(6Z9Z12Z)); PC(18:1(11Z)18:3(9Z12Z15Z)); PC(18:1(9Z)18:3(6Z9Z12Z)); PC(18:1(9Z)18:3(9Z12Z15Z)); PC(18:2(9Z12Z)18:2(9Z12Z)); PC(18:3(6Z9Z12Z)18:1(11Z)); PC(18:3(6Z9Z12Z)18:1(9Z)); PC(18:3(9Z12Z15Z)18:1(11Z)); PC(18:3(9Z12Z15Z)18:1(9Z)); PC(18:4(6Z9Z12Z15Z)18:0); PC(20:3(5Z8Z11Z)16:1(9Z)); PC(20:3(8Z11Z14Z)16:1(9Z)); PC(20:4(5Z8Z11Z14Z)16:0); PC(20:4(8Z11Z14Z17Z)16:0); PC(22:4(7Z10Z13Z16Z)14:0) | HMDB07889; HMDB07982; HMDB07983; HMDB08013; HMDB08014; HMDB08042; HMDB08073; HMDB08074; HMDB08106; HMDB08107; HMDB08138; HMDB08169; HMDB08170; HMDB08202; HMDB08203; HMDB08234; HMDB08365; HMDB08398; HMDB08429; HMDB08462; HMDB08623 | M+H | - | - |
| C18 negative | 138.031 | 294.5 | 0.13 (0.83) | -0.09 (1.11) | 0.22 (0, 0.44) | 0.0473 | 0.6181 | 5-Hydroxypyrazinamide | HMDB01978 | M-H | - | - |
| C18 negative | 338.27 | 174.2 | 0.14 (1.05) | -0.07 (0.93) | 0.22 (0, 0.43) | 0.0475 | 0.6181 | Oleoyl glycine | HMDB13631 | M-H | - | - |
| C18 negative | 411.348 | 294.3 | -0.11 (1.05) | 0.1 (0.75) | -0.21 (-0.41, 0) | 0.0477 | 0.6181 | MG(0:022:1(13Z)0:0); MG(22:1(13Z)0:00:0) | HMDB11552; HMDB11582 | M-H | - | - |
| HILIC positive | 1013.497 | 143.4 | -0.08 (1.04) | -0.31 (1.18) | 0.22 (0, 0.45) | 0.0477 | 0.8113 | Licoricesaponin D3 | HMDB38431 | M+H | - | - |
| HILIC positive | 171.138 | 296.7 | 0.12 (0.67) | -0.13 (1.44) | 0.24 (0, 0.48) | 0.0483 | 0.8113 | cis-4-Decenoic acid; trans-Dec-2-enoic acid; 8-Methylnonenoate; 5Z-Octenyl acetate; Methyl (xi)-3-nonenoate; ()-6-Methyl-5-hepten-2-yl acetate; 3-Decenoic acid; 9-Decenoic acid; 2-Octenyl acetate; Cyclohexylethyl acetate; Linalool oxide (trans-pyranoid); Linalool oxide III; 4-Hydroxy-26-dimethyl-7-octen-3-one; Ethyl 3-octenoate; 2-Methylbutyl 3-methyl-2-butenoate; 1-Octen-3-yl acetate; 6-Decanolide; cis-3-Hexenyl butyrate; Cyclohexyl butanoate; 2-Hexenyl butanoate; 3-Methyl-24-nonanedione; (SE)-26-Dimethyl-57-octadiene-23-diol; Cnidiol C; Citronellic acid; (E)-3-(Tetrahydro-55-dimethyl-2-furanyl)-2-buten-1-ol; Linalyl oxide; Lilac alcohol; Hexyl crotonate; 26-Dimethyl-37-octadiene-26-diol; 1-Cyclopropyl-4-methyl-13-cyclohexanediol; trans-p-Menth-2-ene-14-diol; delta-Decalactone; xi-5-Hexyldihydro-2(3H)-furanone; Ethyl (E)-2-octenoate; xi-Tetrahydro-3-pentyl-2H-pyran-2-one; 9-Hydroxygeraniol; ()-6-Hydroxy-26-dimethyl-7-octen-4-one; 5-Decenoic acid; Ethyl 4Z-octenoate; Methyl 2-nonenoate; 2-Propenyl heptanoate; cis-3-Hexenyl isobutyrate; Menthone lactone; Isopentyl 3-methyl-2-butenoate; ()-cis-Linalyl oxide; ()-trans-Linalyl oxide; 6-Decenoic acid; ()-cis-Linalool 37-oxide; 2-Exo-hydroxy-18-cineole; 2-Hydroxycineol | HMDB04980; HMDB10726; HMDB12183; HMDB29358; HMDB29859; HMDB30031; HMDB31002; HMDB31003; HMDB31297; HMDB31362; HMDB31440; HMDB31441; HMDB32119; HMDB32275; HMDB32393; HMDB32452; HMDB33203; HMDB33377; HMDB34427; HMDB34581; HMDB34880; HMDB35122; HMDB35365; HMDB35837; HMDB35839; HMDB35907; HMDB36099; HMDB36211; HMDB36990; HMDB36992; HMDB37021; HMDB37116; HMDB37217; HMDB37497; HMDB37632; HMDB38744; HMDB39149; HMDB39768; HMDB39795; HMDB40194; HMDB40208; HMDB40213; HMDB40330; HMDB40530; HMDB40726; HMDB40727; HMDB41012; HMDB41630; HMDB59610; HMDB59852 | M+H | - | - |
| C18 negative | 209.0301 | 19.8 | 0.28 (0.57) | 0.11 (0.88) | 0.17 (0, 0.34) | 0.0484 | 0.6181 | Galactaric acid; Glucaric acid; D-Glucaric acid | HMDB00639; HMDB00663; HMDB29881 | M-H | Tetrahydroxyhexanedioic acid (saccharate/galactarate) | M-H |
| HILIC positive | 864.4521 | 54 | 0.19 (0.26) | 0.09 (0.56) | 0.1 (0, 0.2) | 0.0484 | 0.8113 | Casomorphin | HMDB59787 | M+H | - | - |
| C18 negative | 377.2695 | 250.9 | 0.11 (0.95) | -0.08 (0.89) | 0.18 (0, 0.37) | 0.0486 | 0.6181 | 2-Arachidonylglycerol; MG(0:020:4(8Z11Z14Z17Z)0:0); MG(20:4(5Z8Z11Z14Z)0:00:0); MG(20:4(8Z11Z14Z17Z)0:00:0); 1-Acetoxy-2-hydroxy-51215-heneicosatrien-4-one; 12-Gingerol; Persenone A | HMDB04666; HMDB11549; HMDB11578; HMDB11579; HMDB35273; HMDB36356; HMDB36568 | M-H | - | - |
| C18 negative | 827.7476 | 41.4 | 0.24 (0.58) | 0.12 (0.53) | 0.12 (0, 0.24) | 0.0488 | 0.6181 | TG(15:018:3(6Z9Z12Z)o-18:0); TG(15:018:3(9Z12Z15Z)o-18:0); TG(15:0o-18:018:3(6Z9Z12Z)); TG(15:0o-18:018:3(9Z12Z15Z)); TG(14:1(9Z)18:2(9Z12Z)o-18:0); TG(14:1(9Z)20:2n6o-18:0); TG(14:1(9Z)o-18:018:2(9Z12Z)); TG(14:1(9Z)o-18:020:2n6); TG(18:2(9Z12Z)14:1(9Z)o-18:0); TG(18:3(6Z9Z12Z)15:0o-18:0); TG(20:2n614:1(9Z)o-18:0); TG(18:3(9Z12Z15Z)15:0o-18:0) | HMDB43452; HMDB43655; HMDB43816; HMDB43823; HMDB48091; HMDB48137; HMDB48399; HMDB48401; HMDB52515; HMDB52872; HMDB53367; HMDB55161 | M-H | - | - |
| C18 negative | 171.0662 | 218.3 | -0.04 (0.92) | 0.12 (0.66) | -0.17 (-0.33, 0) | 0.0491 | 0.6181 | 2-Octenedioic acid; cis-4-Octenedioic acid; trans-3-Octenedioic acid; 3-Isopropenylpentanedioic acid; Ethyl 24-dioxohexanoate; Diethyl fumarate | HMDB00341; HMDB04982; HMDB13312; HMDB32352; HMDB34851; HMDB38601 | M-H | - | - |

| | | | | | | | | | | | | |
|---|---|---|---|---|---|---|---|---|---|---|---|---|
| C18 negative | 277.0114 | 217.8 | -0.11 (1) | 0.12 (1.02) | -0.23 (-0.45, 0) | 0.0491 | 0.6181 | Fenthion | HMDB33209 | M-H | - | - |
| C18 negative | 197.0818 | 156.5 | -0.11 (1.14) | 0.11 (0.86) | -0.22 (-0.45, 0) | 0.0492 | 0.6181 | Guaifenesin; 5-(3E-Pentenyl)tetrahydro-2-oxo-3-furancarboxylic acid; 3-(4-Hydroxy-3-methoxyphenyl)-12-propanediol; (2E4E)-27-Dimethyl-24-octadienedioic acid; 34-Methylene suberic acid; 23-Methylene suberic acid | HMDB04998; HMDB30991; HMDB33093; HMDB34099; HMDB59768; HMDB59779 | M-H | - | - |
| C18 negative | 445.3319 | 235.9 | -0.11 (0.88) | 0.07 (0.72) | -0.17 (-0.35, 0) | 0.0495 | 0.6181 | 13-Carboxy-gamma-tocopherol; (3beta5alpha6beta7alpha22E24R)-Ergosta-822-diene-3567-tetrol; (3beta5alpha6alpha9alpha22E24R)-Ergosta-722-diene-3569-tetrol; 359-Trihydroxyergost-7-en-6-one; Secasterone; 3-Dehydroteasterone | HMDB12557; HMDB32107; HMDB32123; HMDB32875; HMDB40999; HMDB41527 | M-H | - | - |
| C18 negative | 484.2838 | 114.9 | -0.21 (0.98) | 0 (0.96) | -0.22 (-0.43, 0) | 0.0495 | 0.6181 | Deoxycholic acid glycine conjugate; Chenodeoxycholic acid glycine conjugate; Glycoursodeoxycholic acid; Chenodeoxyglycocholic acid | HMDB00631; HMDB00637; HMDB00708; HMDB06898 | M+Cl | - | - |
| C18 negative | 340.2856 | 183.1 | 0.07 (0.96) | -0.21 (1.34) | 0.27 (0, 0.55) | 0.0495 | 0.6181 | Stearoylglycine | HMDB13308 | M-H | - | - |
| C18 negative | 229.2074 | 223.3 | 0.08 (0.99) | -0.14 (1.07) | 0.22 (0, 0.43) | 0.0498 | 0.6181 | Myristic acid; 2610-Trimethylundecanoic acid; 12-Methyltridecanoic acid; Hexanal octane-13-diol acetal; Undecanal propyleneglycol acetal; Ethyl dodecanoate; Heptyl heptanoate; Hexyl octanoate; Decyl butanoate; 3-Methylbutyl nonanoate; Nonyl isovalerate; Dodecyl acetate; Isobutyl decanoate | HMDB00806; HMDB02221; HMDB31072; HMDB32318; HMDB32548; HMDB33788; HMDB34461; HMDB36216; HMDB37312; HMDB37495; HMDB38034; HMDB38969; HMDB59865 | M-H_[+2] | - | - |
| HILIC positive | 419.315 | 26.9 | 0.14 (0.72) | -0.05 (0.96) | 0.18 (0, 0.36) | 0.0500 | 0.8113 | 11-Carboxy-alpha-chromanol; Hexadecyl ferulate | HMDB12515; HMDB39317 | M+H | - | - |

SD: Standard Deviation; CI: Confidence Interval; FDR: False Discovery Rate.

**Supplemental Table 5.** Results of the Untargeted Metabolome-wide Association Study Comparing TB Resisters and Controls in the Indian Cohort (103 Pairs). Metabolites with P Value < 0.05 and Level 1 or 2 Confirmation are Shown.

| Platform | m/z | time | Resister Mean (SD) | Control Mean (SD) | Difference Resister vs. Control (95% CI) | P | FDR | Level 2 Confirmation | | | Level 1 Confirmation | |
|---|---|---|---|---|---|---|---|---|---|---|---|---|
| | | | | | | | | Name | HMDB ID | Adduct | Name | Adduct |
| C18 negative | 562.383 | 190.8 | -0.08 (1.02) | 0.27 (0.51) | -0.36 (-0.54, -0.18) | 0.0002 | 0.8915 | Cholesterol glucuronide | HMDB10330 | M-H_[-1] | - | - |
| HILIC positive | 221.1536 | 297.2 | 0.2 (0.68) | -0.19 (0.84) | 0.39 (0.19, 0.59) | 0.0002 | 0.8047 | 26-Di-tert-butylbenzoquinone; 3-Phenylpropyl isovalerate; 13-Nor-6-eremophilene-811-dione; Hexyl phenylacetate; 2-Phenylethyl hexanoate; (Z)-6-Tetradecene-13-diyne-58-diol; 13-Tetradecene-13-diyne-67-diol; 34-Dihydro-22578-pentamethyl-2H-1-benzopyran-6-ol; 2-Methyl-1-phenyl-2-propanyl butyrate | HMDB13817; HMDB36389; HMDB37606; HMDB37714; HMDB37718; HMDB38996; HMDB39539; HMDB40217; HMDB40226 | M+H | - | - |
| C18 negative | 346.2466 | 187.9 | 0.28 (0.79) | -0.09 (0.94) | 0.38 (0.17, 0.58) | 0.0005 | 0.8915 | Methyl-10-shogaol; Ginkgoic acid; Siderol; ent-17-Acetoxy-16b-kauran-19-al; Volemolide; (R)-34-Dihydro-8-hydroxy-3-tridecyl-1H-2-benzopyran-1-one | HMDB31465; HMDB33897; HMDB34944; HMDB36722; HMDB41001; HMDB41302 | M-H_[-1] | - | - |
| C18 negative | 613.1796 | 146.1 | -0.43 (0.95) | -0.08 (1.01) | -0.35 (-0.54, -0.16) | 0.0005 | 0.8915 | Catechin 34-diglucoside; Catechin 35-diglucoside; Catechin 37-diglucoside | HMDB37950; HMDB37951; HMDB37952 | M-H | - | - |
| C18 negative | 210.07 | 25.6 | -0.25 (1.37) | 0.21 (0.68) | -0.45 (-0.71, -0.2) | 0.0006 | 0.8915 | 1-Hydroxylorcaserin; 5-Hydroxylorcaserin; 7-Hydroxylorcaserin; N-Hydroxylorcaserin; 4-(4-chlorophenyl)-4-hydroxypiperidine | HMDB60704; HMDB60778; HMDB60800; HMDB60844; HMDB60902 | M-H | - | - |
| C18 negative | 415.3217 | 227.9 | -0.24 (1.16) | 0.15 (0.84) | -0.39 (-0.63, -0.16) | 0.0011 | 0.8915 | 2425-Dihydroxyvitamin D; 2526-dihydroxyvitamin D; Calcitriol; 7a12a-Dihydroxy-cholestene-3-one; 24R25-Dihydroxyvitamin D3; 23S25-dihydroxyvitamin D3; 3 beta-Hydroxy-5-cholestenoate; 7 alpha24-Dihydroxy-4-cholesten-3-one; 7 alpha26-Dihydroxy-4-cholesten-3-one; Paricalcitol; Sarsasapogenin; Neotigogenin; Smilagenin; Octadecyl cis-p-coumarate; (24R)-2425-Dihydroxycalciol; 7alpha25-Dihydroxy-4-cholesten-3-one; Secalciferol | HMDB00430; HMDB01420; HMDB01903; HMDB02197; HMDB06226; HMDB06720; HMDB12453; HMDB12457; HMDB12459; HMDB15046; HMDB30024; HMDB30047; HMDB30048; HMDB35300; HMDB60133; HMDB60425; HMDB60508 | M-H | - | - |
| HILIC positive | 259.1904 | 196.4 | -0.35 (1.27) | 0.1 (0.9) | -0.45 (-0.72, -0.18) | 0.0011 | 0.8047 | Tetradecanedioic acid; Diethyl decanedioate; Diisobutyl adipate; 2-Hydroxypropyl 2-isopropyl-5-methylcyclohexyl carbonate; 1-Hydroxypropan-2-yl 2-isopropyl-5-methylcyclohexyl carbonate | HMDB00872; HMDB40429; HMDB41618; HMDB59946; HMDB59947 | M+H | - | - |
| C18 negative | 491.193 | 169 | 0.12 (1.18) | -0.16 (1.41) | 0.28 (0.11, 0.45) | 0.0016 | 0.8915 | Isolariciresinol 9-O-alpha-L-arabinofuranoside | HMDB32906 | M-H | - | - |
| C18 negative | 252.1089 | 293.8 | -0.2 (1.25) | 0.21 (0.88) | -0.41 (-0.67, -0.15) | 0.0025 | 0.8915 | Choline bitartrate | HMDB40269 | M-H | - | - |
| HILIC positive | 778.5741 | 53.6 | 0.05 (0.38) | 0.17 (0.32) | -0.13 (-0.21, -0.04) | 0.0033 | 0.8047 | PE(22:4(7Z10Z13Z16Z)P-18:1(11Z)); PE(22:4(7Z10Z13Z16Z)P-18:1(9Z)); PE(22:5(4Z7Z10Z13Z16Z)dm18:0); PE(22:5(7Z10Z13Z16Z19Z)P-18:0); PE(P-18:022:5(4Z7Z10Z13Z16Z)); PE(P-18:022:5(7Z10Z13Z16Z19Z)); PE(P-18:1(11Z)22:4(7Z10Z13Z16Z)); PE(P-18:1(9Z)22:4(7Z10Z13Z16Z)) | HMDB09611; HMDB09612; HMDB09643; HMDB09676; HMDB11392; HMDB11393; HMDB11424; HMDB11457 | M+H | - | - |
| HILIC positive | 173.0298 | 64.1 | -0.07 (1.08) | 0.21 (0.79) | -0.28 (-0.47, -0.09) | 0.0039 | 0.8047 | Dihydrothymine; Squamolone; L-Cyclo(alanylglycyl) | HMDB00079; HMDB29874; HMDB31547 | M+2Na-H | - | - |
| C18 negative | 101.0608 | 292.1 | 0.18 (0.49) | 0.01 (0.42) | 0.17 (0.06, 0.29) | 0.0039 | 0.8915 | Isovaleric acid; Valeric acid; Ethylmethylacetic acid; Ethyl propionate; Tetrahydro-2-furanmethanol; Isopropyl acetate; 2-Methylpropyl formate; Methyl isobutyrate; 3-Hydroxy-2-pentanone; 24-Dimethyl-13-dioxolane; Ethylene oxidepropylene oxide copolymer; (S)-2-Methylbutanoic acid; Methyl butyrate; Propyl acetate; Tetrahydro-2-methyl-3-furanol; Butyl formate; Pivalic acid; 1-Hydroxy-2-pentanone | HMDB00718; HMDB00892; HMDB02176; HMDB30058; HMDB31175; HMDB31241; HMDB31247; HMDB31249; HMDB31516; HMDB32230; HMDB32263; HMDB33742; HMDB33890; HMDB34237; HMDB39799; HMDB40575; HMDB41992; HMDB59678 | M-H | - | - |
| HILIC positive | 876.6842 | 34.1 | -0.1 (1.09) | 0.26 (0.68) | -0.36 (-0.59, -0.12) | 0.0040 | 0.8047 | PC(o-22:022:6(4Z7Z10Z13Z16Z19Z)); PC(o-22:3(10Z13Z16Z)22:3(10Z13Z16Z)) | HMDB13450; HMDB13457 | M+H | - | - |
| C18 negative | 199.0378 | 22.8 | -0.19 (1.14) | 0.18 (0.81) | -0.37 (-0.62, -0.12) | 0.0041 | 0.8915 | Potassium 2-(1-ethoxy) ethoxypropanoate | HMDB37192 | M-H | - | - |
| C18 negative | 291.6261 | 187.9 | -0.24 (1.23) | 0.14 (0.77) | -0.39 (-0.65, -0.12) | 0.0042 | 0.8915 | 1112-Dimethoxydihydrokawain; Coriandrone A; Coriandrone B | HMDB30767; HMDB37774; HMDB37775 | M-H_[-1] | - | - |
| C18 negative | 179.0713 | 25 | -0.01 (0.82) | 0.28 (0.5) | -0.29 (-0.48, -0.09) | 0.0042 | 0.8915 | 3-Methoxybenzenepropanoic acid; Coniferyl alcohol; 4-Ethoxy-3-methoxybenzaldehyde; 2-Phenyl-13-dioxolane-4- | HMDB11751; HMDB12915; HMDB29685; HMDB31240; | M-H | - | - |

| | | | | | | | | | | | | |
|---|---|---|---|---|---|---|---|---|---|---|---|---|
| | | | | | | | | methanol; 1-(24-Dihydroxyphenyl)-1-butanone; Benzaldehyde glyceryl acetal; Propylparaben; Ethyl p-anisate; 4-Methoxybenzyl acetate; 4-Hydroxy-266-trimethyl-3-oxo-14-cyclohexadiene-1-carboxaldehyde; Propyl 2-furanacrylate; 3-(3-Hydroxyphenyl)-2-methylpropionic acid | HMDB32128; HMDB32174; HMDB32574; HMDB32640; HMDB33761; HMDB37602; HMDB37733; HMDB60734 | | | |
| C18 negative | 431.2438 | 69.2 | 0.25 (0.58) | -0.07 (1.09) | 0.32 (0.1, 0.54) | 0.0046 | 0.8915 | Erinacine E; Colupdox a; Erinacine A; Erinacine B | HMDB29656; HMDB39597; HMDB40291; HMDB40390 | M-H | - | - |
| C18 negative | 338.3145 | 300.7 | 0.06 (0.95) | 0.34 (0.64) | -0.28 (-0.47, -0.09) | 0.0053 | 0.8915 | Erucic acid; Cetoleic acid; Catelaidic acid; Phytyl acetate; 79-Docosanedione; 68-Docosanedione; 57-Docosanedione; 46-Docosanedione | HMDB02068; HMDB02884; HMDB31013; HMDB32470; HMDB35566; HMDB35567; HMDB35568; HMDB35569 | M-H_[-1] | - | - |
| C18 negative | 113.989 | 34.7 | -0.19 (1.24) | 0.2 (0.88) | -0.39 (-0.67, -0.11) | 0.0061 | 0.8915 | Trifluoroacetic acid | HMDB14118 | M-H_[-1] | - | - |
| C18 negative | 149.0607 | 25.1 | -0.16 (1.04) | 0.18 (0.79) | -0.35 (-0.59, -0.1) | 0.0065 | 0.8915 | Hydrocinnamic acid; 4-Ethylbenzoic acid; 3-Methylphenylacetic acid; 34-Dimethylbenzoic acid; 4-Coumaryl alcohol; 2-Phenylpropionate; 2-Methoxy-4-vinylphenol; Benzyl acetate; o-Tolyl acetate; p-Tolyl acetate; 2-Methoxyacetophenone; 4-Methoxyacetophenone; 2-Hydroxy-5-methylacetophenone; Methyl phenylacetate; Ethyl benzoate; 4-Ethoxybenzaldehyde; 1-Phenylethyl formate; 2-Ethyl-4-(2-furanyl)-2-propenal; 2-Phenylethyl formate; 2-(2-Furanyl)-3-methyl-2-butenal; 4-Hydroxy-3-methylacetophenone | HMDB00764; HMDB02097; HMDB02222; HMDB02237; HMDB03654; HMDB11743; HMDB13744; HMDB31310; HMDB32073; HMDB32075; HMDB32569; HMDB32570; HMDB32592; HMDB32617; HMDB33967; HMDB33970; HMDB37715; HMDB38355; HMDB40428; HMDB40925; HMDB59824 | M-H | - | - |
| HILIC positive | 999.4746 | 28.1 | -0.15 (1.16) | 0.22 (0.82) | -0.37 (-0.63, -0.1) | 0.0066 | 0.8047 | (S)-Nerolidol 3-O-a-L-Rhamnopyranosyl-(1-4)-a-L-rhamnopyranosyl-(1-2)-4-(4-hydroxy-3-methoxycinnamoyl)-(E)-a-L-rhamnopyranosyl-(1-6)-b-D-glucopyranoside | HMDB40848 | M+H | - | - |
| C18 negative | 219.0342 | 22.2 | 0.28 (1.1) | -0.09 (0.94) | 0.37 (0.1, 0.63) | 0.0067 | 0.8915 | 1-Propenyl 1-(1-propenylthio)propyl disulfide | HMDB38967 | M-H | - | - |
| C18 negative | 155.0713 | 217.2 | 0.2 (0.83) | -0.16 (1.17) | 0.37 (0.1, 0.63) | 0.0069 | 0.8915 | 8-Hydroxy-56-octadienoic acid; 25-Dimethyl-4-ethoxy-3(2H)-furanone | HMDB31101; HMDB32232 | M-H | - | - |
| C18 negative | 291.0831 | 20 | -0.65 (1.69) | -0.12 (0.93) | -0.53 (-0.91, -0.15) | 0.0070 | 0.8915 | Edetic Acid | HMDB15109 | M-H | - | - |
| HILIC positive | 431.3153 | 28 | -0.19 (1.27) | 0.19 (0.78) | -0.37 (-0.64, -0.1) | 0.0073 | 0.8047 | MG(0:024:6(6Z9Z12Z15Z18Z21Z)0:0); MG(24:6(6Z9Z12Z15Z18Z21Z)0:00:0); 7 alpha-Hydroxy-3-oxo-4-cholestenoate; Australigenin; Barogenin; Schidigeragenin C; 25-Hydroxyvitamin D3-2623-lactol; 24-Oxo-1alpha25-dihydroxyvitamin D3 | HMDB11560; HMDB11590; HMDB12458; HMDB30066; HMDB34403; HMDB36249; HMDB60127; HMDB60128 | M+H | - | - |
| C18 negative | 167.0342 | 21.3 | -0.24 (1.5) | 0.23 (0.82) | -0.46 (-0.8, -0.13) | 0.0078 | 0.8915 | Homogentisic acid; Vanillic acid; 3-Hydroxymandelic acid; p-Hydroxymandelic acid; 34-Dihydroxybenzeneacetic acid; 5-Methoxysalicylic acid; 34-Dihydroxymandelaldehyde; 246-Trihydroxyacetophenone; 26-Dimethoxy-14-benzoquinone; 3-Acetyl-4-hydroxy-6-methyl-2H-pyran-2-one; Thermophillin; 3-Methoxysalicylic acid; Isovanillic acid | HMDB00130; HMDB00484; HMDB00750; HMDB00822; HMDB01336; HMDB01868; HMDB06242; HMDB29644; HMDB29673; HMDB33129; HMDB34335; HMDB59713; HMDB60003 | M-H | Homogent isate | M-H |
| C18 negative | 491.3378 | 179.4 | -0.21 (1.22) | 0.16 (0.89) | -0.37 (-0.64, -0.1) | 0.0078 | 0.8915 | Homodolicholide | HMDB33907 | M-H | - | - |
| HILIC positive | 361.2008 | 30.2 | -0.18 (1.12) | 0.18 (0.75) | -0.36 (-0.62, -0.09) | 0.0081 | 0.8047 | Aldosterone; Cortisone; 19-Oic-deoxycorticosterone; Prednisolone; Tricyclodehydroisohumulone; 7-Methylrosmanol | HMDB00037; HMDB02802; HMDB12613; HMDB14998; HMDB30165; HMDB35813 | M+H | - | - |
| HILIC positive | 154.983 | 121 | -0.68 (1.45) | -0.26 (1.2) | -0.42 (-0.73, -0.11) | 0.0086 | 0.8047 | (E)-S-1-Propenyl thiosulfate | HMDB41079 | M+H | - | - |
| HILIC positive | 114.0662 | 36.1 | -0.08 (0.86) | 0.11 (0.75) | -0.19 (-0.33, -0.05) | 0.0087 | 0.8047 | Creatinine | HMDB00562 | M+H | Creatinine | M+H |
| C18 negative | 431.2819 | 289.5 | -0.22 (1.11) | 0.12 (0.85) | -0.33 (-0.58, -0.09) | 0.0087 | 0.8915 | Latanoprost | HMDB14792 | M-H | - | - |
| HILIC positive | 371.2984 | 27.6 | -0.08 (0.96) | 0.2 (0.58) | -0.28 (-0.49, -0.07) | 0.0096 | 0.8047 | cis-5-Tetradecenoylcarnitine; trans-2-Tetradecenoylcarnitine | HMDB02014; HMDB13329 | M+H_[+1] | - | - |
| C18 negative | 566.1816 | 142.8 | -0.16 (0.99) | 0.18 (1.02) | -0.33 (-0.58, -0.08) | 0.0096 | 0.8915 | Neoacrimarine H | HMDB33031 | M-H | - | - |
| C18 negative | 169.0253 | 21.3 | -0.17 (1.12) | 0.15 (0.91) | -0.33 (-0.57, -0.08) | 0.0097 | 0.8915 | Homogentisic acid; Uric acid; Vanillic acid; 3-Hydroxymandelic acid; p-Hydroxymandelic acid; 34-Dihydroxybenzeneacetic acid; 5-Methoxysalicylic acid; 34-Dihydroxymandelaldehyde; 246-Trihydroxyacetophenone; 26-Dimethoxy-14-benzoquinone; 3-Acetyl-4-hydroxy-6-methyl-2H-pyran-2-one; Thermophillin; 3-Methoxysalicylic acid; Isovanillic acid | HMDB00130; HMDB00289; HMDB00484; HMDB00750; HMDB00822; HMDB01336; HMDB01868; HMDB06242; HMDB29644; HMDB29673; HMDB33129; HMDB34335; HMDB59713; HMDB60003 | M-H_[+2] | - | - |
| HILIC positive | 784.6559 | 53.4 | 0.43 (0.31) | 0.29 (0.54) | 0.14 (0.03, 0.24) | 0.0101 | 0.8047 | CerP(d18:126:1(17Z)) | HMDB10707 | M+H | - | - |
| HILIC positive | 850.6676 | 34.1 | -0.15 (1.2) | 0.14 (0.62) | -0.3 (-0.52, -0.07) | 0.0102 | 0.8047 | PC(o-22:1(13Z)20:4(8Z11Z14Z17Z)) | HMDB13451 | M+H | - | - |
| C18 negative | 124.9914 | 21.4 | -0.2 (1.07) | 0.14 (0.9) | -0.34 (-0.6, -0.08) | 0.0106 | 0.8915 | 2-Hydroxyethanesulfonate; Ethyl hydrogen sulfate | HMDB03903; HMDB31233 | M-H | - | - |
| HILIC positive | 209.0916 | 53.1 | -0.08 (1.02) | 0.21 (0.89) | -0.29 (-0.51, -0.07) | 0.0106 | 0.8047 | L-Kynurenine; Formyl-5-hydroxykynurenamine; 4-Aminobenzoyl-(beta)-alanine | HMDB00684; HMDB12948; HMDB60757 | M+H | Kynurenin e | M+H |
| HILIC positive | 238.0388 | 79 | -0.68 (1.23) | -0.35 (1.18) | -0.32 (-0.57, -0.08) | 0.0107 | 0.8047 | S-Cysteinosuccinic acid | HMDB29418 | M+H | - | - |

| | | | | | | | | | | | | |
|---|---|---|---|---|---|---|---|---|---|---|---|---|
| C18 negative | 433.3323 | 293.1 | -0.19 (1.6) | 0.23 (0.61) | -0.43 (-0.75, -0.1) | 0.0108 | 0.8915 | 3a7a-Dihydroxycoprostanic acid; 3a7a12a-Trihydroxy-5b-cholestan-26-al | HMDB00359; HMDB03533 | M-H | - | - |
| C18 negative | 157.0366 | 295.9 | -0.12 (0.92) | 0.17 (0.93) | -0.29 (-0.51, -0.07) | 0.0108 | 0.8915 | Allantoin | HMDB00462 | M-H | - | - |
| C18 negative | 478.3253 | 178.6 | -0.24 (1.43) | 0.16 (0.79) | -0.39 (-0.69, -0.09) | 0.0110 | 0.8915 | Dolicholide; Polyporusterone A | HMDB34086; HMDB38495 | M-H_[-1] | - | - |
| HILIC positive | 379.1498 | 100.5 | 0.18 (0.82) | -0.11 (1.06) | 0.29 (0.07, 0.52) | 0.0120 | 0.8047 | Riboflavin; (1RS2RS)-Guaiacylglycerol 1-glucoside; (1x2x)-Guaiacylglycerol 2-glucoside; (1x2x)-Guaiacylglycerol 3-glucoside | HMDB00244; HMDB33300; HMDB33301; HMDB40600 | M+H_[+2] | - | - |
| HILIC positive | 317.2515 | 28.9 | -0.15 (1.14) | 0.17 (0.65) | -0.32 (-0.57, -0.07) | 0.0122 | 0.8047 | Decanoylcarnitine | HMDB00651 | M+H_[+1] | - | - |
| HILIC positive | 177.0951 | 287.6 | 0.2 (0.5) | 0.34 (0.25) | -0.14 (-0.25, -0.03) | 0.0122 | 0.8047 | Methionyl butyrate; Methylthiomethyl hexanoate; (S)-3-Mercaptohexyl acetate | HMDB32378; HMDB38300; HMDB38920 | M+H | - | - |
| HILIC positive | 344.2792 | 27.5 | -0.08 (1.04) | 0.2 (0.51) | -0.28 (-0.5, -0.06) | 0.0124 | 0.8047 | Dodecanoylcarnitine | HMDB02250 | M+H | Lauroylcar nitine | M+H |
| C18 negative | 314.0684 | 20.5 | -1.18 (0.73) | -1.04 (0.71) | -0.14 (-0.25, -0.03) | 0.0128 | 0.8915 | Avenanthramide C | HMDB38576 | M-H | - | - |
| HILIC positive | 658.5416 | 26.8 | -0.11 (1.14) | 0.22 (0.83) | -0.33 (-0.58, -0.07) | 0.0128 | 0.8047 | DG(16:022:6(4Z7Z10Z13Z16Z19Z)0:0); DG(16:1(9Z)22:5(4Z7Z10Z13Z16Z)0:0); DG(16:1(9Z)22:5(7Z10Z13Z16Z19Z)0:0); DG(18:1(11Z)20:5(5Z8Z11Z14Z17Z)0:0); DG(18:1(9Z)20:5(5Z8Z11Z14Z17Z)0:0); DG(18:2(9Z12Z)20:4(5Z8Z11Z14Z)0:0); DG(18:2(9Z12Z)20:4(8Z11Z14Z17Z)0:0); DG(18:3(6Z9Z12Z)20:3(5Z8Z11Z)0:0); DG(18:3(6Z9Z12Z)20:3(8Z11Z14Z)0:0); DG(18:3(9Z12Z15Z)20:3(5Z8Z11Z)0:0); DG(18:3(9Z12Z15Z)20:3(8Z11Z14Z)0:0); DG(18:4(6Z9Z12Z15Z)20:2(11Z14Z)0:0); DG(20:2(11Z14Z)18:4(6Z9Z12Z15Z)0:0); DG(20:3(5Z8Z11Z)18:3(6Z9Z12Z)0:0); DG(20:3(5Z8Z11Z)18:3(9Z12Z15Z)0:0); DG(20:3(8Z11Z14Z)18:3(6Z9Z12Z)0:0); DG(20:3(8Z11Z14Z)18:3(9Z12Z15Z)0:0); DG(20:4(5Z8Z11Z14Z)18:2(9Z12Z)0:0); DG(20:4(8Z11Z14Z17Z)18:2(9Z12Z)0:0); DG(20:5(5Z8Z11Z14Z17Z)18:1(11Z)0:0); DG(20:5(5Z8Z11Z14Z17Z)18:1(9Z)0:0); DG(22:5(4Z7Z10Z13Z16Z)16:1(9Z)0:0); DG(22:5(7Z10Z13Z16Z19Z)16:1(9Z)0:0); DG(22:6(4Z7Z10Z13Z16Z19Z)16:00:0); DG(16:00:022:6n3); DG(16:1n70:022:5n6); DG(16:1n70:022:5n3); DG(18:1n70:020:5n3); DG(18:1n90:020:5n3); DG(20:3n90:018:3n6); DG(20:3n90:018:3n3); DG(18:3n60:020:3n6); DG(20:3n60:018:3n3) | HMDB07121; HMDB07148; HMDB07149; HMDB07201; HMDB07230; HMDB07257; HMDB07258; HMDB07284; HMDB07285; HMDB07313; HMDB07314; HMDB07341; HMDB07425; HMDB07452; HMDB07453; HMDB07481; HMDB07482; HMDB07509; HMDB07538; HMDB07565; HMDB07566; HMDB07708; HMDB07737; HMDB07765; HMDB56035; HMDB56170; HMDB56175; HMDB56194; HMDB56213; HMDB56238; HMDB56245; HMDB56298; HMDB56326 | M+NH4 | - | - |
| C18 negative | 154.9989 | 293.5 | -0.07 (0.85) | 0.18 (0.76) | -0.25 (-0.45, -0.05) | 0.0129 | 0.8915 | 25-Furandicarboxylic acid; Methyl phenyl disulfide | HMDB04812; HMDB40939 | M-H | - | - |
| C18 negative | 155.0461 | 20.8 | 0.2 (0.11) | 0.25 (0.23) | -0.05 (-0.1, -0.01) | 0.0130 | 0.8915 | 5-Hydroxymethyl-4-methyluracil; 4-Imidazolone-5-propionic acid; Imidazolelactic acid | HMDB00544; HMDB01014; HMDB02320 | M-H | - | - |
| HILIC positive | 156.0421 | 58.3 | -0.17 (0.56) | -0.06 (0.62) | -0.11 (-0.2, -0.02) | 0.0138 | 0.8047 | N-Methylethanolaminium phosphate | HMDB60173 | M+H | - | - |
| HILIC positive | 253.0809 | 69 | 0.21 (0.92) | 0.48 (0.76) | -0.27 (-0.49, -0.06) | 0.0140 | 0.8047 | 5-Hydroxy-N-formylkynurenine; N5-(34-Dioxo-15-cyclohexadien-1-yl)-L-glutamine | HMDB04086; HMDB35993 | M+H | - | - |
| HILIC positive | 314.1037 | 282.4 | -0.14 (1.08) | 0.15 (0.91) | -0.29 (-0.51, -0.06) | 0.0140 | 0.8047 | Avenanthramide 1f; Avenanthramide E | HMDB29283; HMDB38944 | M+H | - | - |
| HILIC positive | 279.1802 | 287.4 | -0.11 (1.09) | 0.22 (0.83) | -0.33 (-0.59, -0.07) | 0.0141 | 0.8047 | (x)-2-Heptanol glucoside | HMDB35028 | M+H | - | - |
| C18 negative | 500.2094 | 192.5 | -0.14 (1.23) | 0.22 (0.71) | -0.36 (-0.65, -0.07) | 0.0141 | 0.8915 | D-Glucosaminide; Chitotriose | HMDB04270; HMDB06578 | M-H | - | - |
| HILIC positive | 151.024 | 90.6 | -0.29 (1.18) | 0.04 (0.82) | -0.33 (-0.6, -0.07) | 0.0144 | 0.8047 | Tartaric acid; D-Tartaric acid; 2-Methyl-2-(methyldithio)propanal; 23-Dihydroxybutanedioic acid | HMDB00956; HMDB29878; HMDB38889; HMDB59916 | M+H | - | - |
| HILIC positive | 95.0685 | 44.1 | 0.47 (0.19) | 0.42 (0.12) | 0.05 (0.01, 0.1) | 0.0148 | 0.8047 | Aniline | HMDB03012 | M+H_[+1] | - | - |
| HILIC positive | 282.1195 | 54.6 | -0.21 (1.54) | 0.19 (0.63) | -0.4 (-0.72, -0.08) | 0.0150 | 0.8047 | 1-Methyladenosine; N6-Methyladenosine; 2-O-Methyladenosine; 3-O-Methyladenosine; N-(1-Deoxy-1-fructosyl)threonine | HMDB03331; HMDB04044; HMDB04326; HMDB06023; HMDB37843 | M+H | - | - |
| C18 negative | 497.2882 | 182.2 | -0.15 (1.49) | 0.24 (0.65) | -0.39 (-0.7, -0.08) | 0.0150 | 0.8915 | Polyporusterone B; Polyporusterone C | HMDB38496; HMDB38497 | M+Na-2H | - | - |
| HILIC positive | 121.1011 | 28.6 | 0.18 (0.98) | -0.11 (0.92) | 0.29 (0.06, 0.52) | 0.0151 | 0.8047 | 124-Trimethylbenzene; Isopropylbenzene; 124-Tris(methylene)cyclohexane; Mesitylene; o-Ethyltoluene; m-Ethyltoluene; Propylbenzene; Hemimellitene | HMDB13733; HMDB34029; HMDB40458; HMDB41924; HMDB59819; HMDB59848; HMDB59877; HMDB59901 | M+H | - | - |
| C18 negative | 383.1899 | 128.8 | 0.03 (0.91) | 0.23 (0.6) | -0.2 (-0.36, -0.04) | 0.0153 | 0.8915 | Bortezomib | HMDB14334 | M-H | - | - |
| C18 negative | 157.0619 | 293.3 | 0.16 (0.98) | -0.21 (1.29) | 0.37 (0.07, 0.67) | 0.0154 | 0.8915 | 1-(Hydroxymethyl)-55-dimethyl-24-imidazolidinedione | HMDB31670 | M-H | - | - |
| HILIC positive | 437.1012 | 98.8 | 0.2 (0.74) | -0.02 (1.21) | 0.23 (0.04, 0.41) | 0.0159 | 0.8047 | 7-Chloro-334568-hexamethoxyflavone | HMDB32658 | M+H | - | - |
| HILIC positive | 972.5066 | 53.3 | 0.15 (0.64) | -0.06 (0.73) | 0.21 (0.04, 0.38) | 0.0160 | 0.8047 | Rose bengal | HMDB36613 | M+H | - | - |

| | | | | | | | | | | | | |
|---|---|---|---|---|---|---|---|---|---|---|---|---|
| HILIC positive | 288.2167 | 32.6 | -0.08 (1.02) | 0.19 (0.59) | -0.27 (-0.49, -0.05) | 0.0160 | 0.8047 | L-Octanoylcarnitine | HMDB00791 | M+H | Octanoylc arnitine | M+H |
| HILIC positive | 320.0345 | 110.6 | -0.38 (1.69) | 0.05 (0.6) | -0.43 (-0.77, -0.08) | 0.0162 | 0.8047 | L-Cysteinylglycine disulfide | HMDB00709 | M+Na | - | - |
| HILIC positive | 316.2481 | 28.5 | -0.07 (0.92) | 0.19 (0.61) | -0.25 (-0.46, -0.05) | 0.0162 | 0.8047 | Decanoylcarnitine | HMDB00651 | M+H | Decanoylc arnitine | M+H |
| HILIC positive | 203.1277 | 240.7 | 0.12 (0.82) | -0.2 (0.99) | 0.32 (0.06, 0.57) | 0.0163 | 0.8047 | Sebacic acid; R-2-Hydroxy-3-methylbutanoic acid 3-Methylbutanoyl; Oxalic acid dibutyl ester; 2-Ethylsuberic acid; Heptylmalonic acid; 3-Methylazelaic acid | HMDB00792; HMDB31510; HMDB40196; HMDB59708; HMDB59719; HMDB59754 | M+H | - | - |
| HILIC positive | 338.1434 | 74.9 | -0.91 (1.09) | -0.54 (1.13) | -0.37 (-0.68, -0.07) | 0.0164 | 0.8047 | 4-O-alpha-D-Galactopyranosylcalystegine B2 | HMDB38228 | M+H | - | - |
| C18 negative | 475.3427 | 228.1 | -0.12 (0.79) | 0.12 (0.75) | -0.24 (-0.43, -0.04) | 0.0165 | 0.8915 | Homodolichosterone; (2alpha3alpha5alpha22R23R)-232223-Tetrahydroxy-25-methylergost-24(28)en-6-one | HMDB34079; HMDB39443 | M-H | - | - |
| C18 negative | 209.0666 | 26 | -0.16 (1.2) | 0.17 (0.86) | -0.33 (-0.6, -0.06) | 0.0166 | 0.8915 | Sedoheptulose; L-Galacto-2-heptulose; D-Manno-2-heptulose; D-altro-D-manno-Heptose | HMDB03219; HMDB29901; HMDB29935; HMDB29952 | M-H | - | - |
| C18 negative | 561.3796 | 190.5 | -0.1 (1.33) | 0.22 (0.61) | -0.32 (-0.59, -0.06) | 0.0167 | 0.8915 | Cholesterol glucuronide | HMDB10330 | M-H | - | - |
| HILIC positive | 552.4025 | 36.1 | -0.27 (1.6) | 0.14 (0.92) | -0.41 (-0.75, -0.07) | 0.0170 | 0.8047 | LysoPC(20:0); Aliskiren | HMDB10390; HMDB15387 | M+H | - | - |
| HILIC positive | 127.1117 | 295.5 | 0.15 (0.38) | -0.08 (0.86) | 0.23 (0.04, 0.42) | 0.0170 | 0.8047 | (E)-2-octenal; 2-Octenal; (3xi5Z)-15-Octadien-3-ol; 244-Trimethylcyclopentanone; 2-Octen-4-one; 1-Octen-3-one; 3-Methyl-3-hepten-2-one; 5-Methyl-5-hepten-2-one; 6-Methyl-3-hepten-2-one trans-; 4-Octen-3-one; 3-Octen-2-one; (SE)-Filbertone; 5-Octen-2-one; Sulcatone; 2-Ethylidenehexanal; 6-Octenal; (Z)-5-Octenal; 25-Octadien-1-ol; (3E5Z)-35-Octadien-1-ol; (2E4E)-24-Octadien-1-ol | HMDB13809; HMDB30961; HMDB30966; HMDB31197; HMDB31301; HMDB31309; HMDB31590; HMDB31591; HMDB32403; HMDB32451; HMDB33547; HMDB35242; HMDB35390; HMDB35915; HMDB37152; HMDB39769; HMDB39770; HMDB40146; HMDB40149; HMDB40151 | M+H | - | - |
| HILIC positive | 241.1546 | 56.1 | -0.23 (1.58) | 0.18 (0.71) | -0.4 (-0.74, -0.07) | 0.0171 | 0.8047 | Pirbuterol | HMDB15407 | M+H | - | - |
| C18 negative | 329.2331 | 72 | 0.09 (0.98) | -0.15 (0.96) | 0.24 (0.04, 0.44) | 0.0172 | 0.8915 | 91213-TriHOME; 91013-TriHOME; 5812-Trihydroxy-9-octadecenoic acid; (9S10E12S13S)-91213-Trihydroxy-10-octadecenoic acid | HMDB04708; HMDB04710; HMDB30936; HMDB38555 | M-H | - | - |
| C18 negative | 213.0557 | 61 | 0.1 (1.22) | -0.23 (0.98) | 0.33 (0.06, 0.61) | 0.0172 | 0.8915 | Phenyl salicylate; 4-Hydroxy-2-biphenylcarboxylic acid; 3-Phenoxybenzoic acid | HMDB32018; HMDB32583; HMDB41807 | M-H | - | - |
| HILIC positive | 370.2951 | 27.4 | -0.09 (0.98) | 0.19 (0.66) | -0.28 (-0.5, -0.05) | 0.0172 | 0.8047 | cis-5-Tetradecenoylcarnitine; trans-2-Tetradecenoylcarnitine | HMDB02014; HMDB13329 | M+H | - | - |
| HILIC positive | 1023.743 | 63.2 | -0.43 (1.52) | -0.01 (1.09) | -0.42 (-0.76, -0.08) | 0.0172 | 0.8047 | TG(22:6(4Z7Z10Z13Z16Z19Z)22:6(4Z7Z10Z13Z16Z19Z)22:6(4Z7Z10Z13Z16Z19Z)) | HMDB10562 | M+H | - | - |
| C18 negative | 291.1244 | 186.6 | -0.11 (0.86) | 0.12 (0.76) | -0.24 (-0.43, -0.04) | 0.0174 | 0.8915 | 1112-Dimethoxydihydrokawain; Coriandrone A; Coriandrone B | HMDB30767; HMDB37774; HMDB37775 | M-H | - | - |
| HILIC positive | 203.1067 | 295.2 | 0.16 (0.66) | -0.17 (1.32) | 0.34 (0.06, 0.61) | 0.0174 | 0.8047 | 6-Acetyl-22-dimethyl-2H-1-benzopyran; 3-(3-Methylbutylidene)-1(3H)-isobenzofuranone | HMDB30816; HMDB31663 | M+H | - | - |
| HILIC positive | 221.1134 | 95.1 | 0.08 (1.04) | -0.3 (1.34) | 0.37 (0.07, 0.68) | 0.0179 | 0.8047 | N-Acetyl-b-glucosaminylamine; Threoninyl-Threonine | HMDB01104; HMDB29071 | M+H | - | - |
| HILIC positive | 157.0456 | 57.4 | -0.16 (0.55) | -0.06 (0.6) | -0.11 (-0.2, -0.02) | 0.0182 | 0.8047 | N-Methylethanolaminium phosphate | HMDB60173 | M+H_[+1] | - | - |
| C18 negative | 239.0772 | 24.8 | 0.22 (1.06) | -0.11 (0.97) | 0.34 (0.06, 0.62) | 0.0184 | 0.8915 | D-glycero-L-galacto-Octulose | HMDB29954 | M-H | - | - |
| C18 negative | 411.202 | 133.6 | -0.04 (0.89) | 0.22 (0.96) | -0.26 (-0.47, -0.04) | 0.0184 | 0.8915 | 5-(23-Dihydroxy-3-methylbutyl)-4-(34-epoxy-4-methylpentanoyl)-34-dihydroxy-2-isopentanoyl-2-cyclopenten-1-one; Abscisic alcohol 11-glucoside | HMDB30082; HMDB39636 | M-H | - | - |
| HILIC positive | 309.2271 | 295.2 | 0.13 (0.66) | -0.18 (1.23) | 0.31 (0.05, 0.57) | 0.0184 | 0.8047 | 123-Tris(1-ethoxyethoxy)propane | HMDB37162 | M+H | - | - |
| HILIC positive | 567.4191 | 26.9 | -0.05 (1.09) | 0.24 (0.83) | -0.29 (-0.52, -0.05) | 0.0187 | 0.8047 | 3-Hydroxy-ee-caroten-3-one; 3-Hydroxy-be-caroten-3-one; Nigroxanthin; Cryptocapsone; 2-Dehydroplectaniaxanthin | HMDB02020; HMDB03448; HMDB37122; HMDB40657; HMDB40743 | M+H | - | - |
| HILIC positive | 260.1854 | 35.9 | -0.18 (1.2) | 0.14 (0.74) | -0.32 (-0.58, -0.05) | 0.0189 | 0.8047 | Hexanoylcarnitine; L-Hexanoylcarnitine | HMDB00705; HMDB00756 | M+H | Hexanoylc arnitine | M+H |
| C18 negative | 311.2227 | 223.7 | -0.26 (1.62) | 0.15 (0.77) | -0.41 (-0.76, -0.07) | 0.0190 | 0.8915 | 13-L-Hydroperoxylinoleic acid; 8(R)-Hydroperoxylinoleic acid; 9(S)-HPODE; 1213-DiHODE; 1516-DiHODE; 910-DiHODE; ()-(E)-13-Hydroxy-10-oxo-11-octadecenoic acid | HMDB03871; HMDB04706; HMDB06940; HMDB10201; HMDB10208; HMDB10221; HMDB40900 | M-H | - | - |
| HILIC positive | 274.0595 | 68.6 | -0.11 (1.15) | 0.18 (0.76) | -0.28 (-0.52, -0.05) | 0.0196 | 0.8047 | Ergothioneine | HMDB03045 | M+2Na-H | - | - |
| C18 negative | 297.1343 | 45.5 | 0.31 (0.79) | 0.07 (0.77) | 0.23 (0.04, 0.43) | 0.0198 | 0.8915 | Toxin T2 tetrol; 37815-Scirpenetetrol | HMDB36159; HMDB37560 | M-H | - | - |
| C18 negative | 154.9987 | 17 | -0.02 (1.16) | -0.42 (1.43) | 0.4 (0.06, 0.73) | 0.0198 | 0.8915 | 25-Furandicarboxylic acid; Methyl phenyl disulfide | HMDB04812; HMDB40939 | M-H | - | - |
| HILIC positive | 794.607 | 32.7 | -0.22 (1.17) | 0.11 (0.89) | -0.33 (-0.6, -0.05) | 0.0199 | 0.8047 | PC(20:3(5Z8Z11Z)P-18:1(11Z)); PC(20:3(5Z8Z11Z)P-18:1(9Z)); PC(20:3(8Z11Z14Z)P-18:1(11Z)); PC(20:3(8Z11Z14Z)P-18:1(9Z)); PC(20:4(5Z8Z11Z14Z)P-18:0); PC(20:4(8Z11Z14Z17Z)P-18:0); PC(22:4(7Z10Z13Z16Z)P-16:0); PC(P-16:022:4(7Z10Z13Z16Z)); PC(P-18:020:4(5Z8Z11Z14Z)); PC(P-18:020:4(8Z11Z14Z17Z)); PC(P-18:1(11Z)20:3(5Z8Z11Z)); PC(P-18:1(11Z)20:3(8Z11Z14Z)); | HMDB08392; HMDB08393; HMDB08424; HMDB08425; HMDB08456; HMDB08489; HMDB08652; HMDB11226; HMDB11253; HMDB11254; HMDB11284; HMDB11285; HMDB11317; HMDB11318; HMDB11450; HMDB13432 | M+H | - | - |

| | | | | | | | | | | | | |
|---|---|---|---|---|---|---|---|---|---|---|---|---|
| | | | | | | | | PC(P-18:1(9Z)20:3(5Z8Z11Z)); PC(P-18:1(9Z)20:3(8Z11Z14Z)); PE(P-18:1(9Z)20:3(8Z11Z14Z)); PC(o-18:1(9Z)20:4(8Z11Z14Z17Z)) | | | | |
| HILIC positive | 193.1224 | 107.2 | 0.22 (0.35) | -0.13 (1.54) | 0.36 (0.06, 0.66) | 0.0201 | 0.8047 | 2-Methyl-1-phenyl-2-propanyl acetate; 4-Phenyl-2-butyl acetate; Ethyl 4-phenylbutanoate; Benzyl 3-methylbutanoate; 3-Methylbutyl benzoate; 2-Methylpropyl phenylacetate; 2-Phenylethyl butanoate; 2-Phenylethyl 2-methylpropanoate; 5-Isopropyl-2-methylphenol acetate; 3-Phenylpropyl propanoate; 111213-Trinor-1(10)-spirovetivene-27-dione; 4-Methyl-2-(1-phenylethyl)-13-dioxolane 9CI; 4-Methylphenyl 3-methylbutanoate; 1-Phenylethyl isobutyrate; 1-Phenylethyl butyrate; 1-Ethoxy-2-methoxy-4-(1-propenyl)benzene; 2-(2-Hydroxy-4-methylphenyl)-3-pentanone; 2-Ethoxy-1-methoxy-4-(1-propenyl)benzene; 111213-Trinor-135-bisabolatrien-10-oic acid; 2-Benzyl-45-dimethyl-13-dioxolane; Butyl phenylacetate | HMDB31571; HMDB31614; HMDB31618; HMDB32043; HMDB33380; HMDB35010; HMDB35014; HMDB35015; HMDB35447; HMDB36388; HMDB36734; HMDB37165; HMDB37709; HMDB37716; HMDB37717; HMDB37792; HMDB38180; HMDB39496; HMDB39643; HMDB39841; HMDB40427 | M+H | - | - |
| HILIC positive | 204.123 | 44.8 | -0.2 (1.4) | 0.14 (0.56) | -0.34 (-0.62, -0.05) | 0.0202 | 0.8047 | L-Acetylcarnitine | HMDB00201 | M+H | Acetylcarnitine | M+H |
| C18 negative | 178.051 | 22.6 | -0.13 (1) | 0.16 (0.69) | -0.29 (-0.53, -0.05) | 0.0202 | 0.8915 | Hippuric acid; 3-Succinoylpyridine; Adrenochrome; Methyl n-formylanthranilate; 1-(4-Methoxyphenyl)-2-nitroethylene | HMDB00714; HMDB00992; HMDB12884; HMDB32398; HMDB32595 | M-H | - | - |
| HILIC positive | 257.1217 | 60 | 0.12 (1.02) | 0.4 (0.79) | -0.28 (-0.52, -0.04) | 0.0202 | 0.8047 | Echothiophate | HMDB15190 | M+H | - | - |
| HILIC positive | 122.0271 | 183 | 0.17 (0.71) | -0.08 (0.85) | 0.25 (0.04, 0.46) | 0.0203 | 0.8047 | L-Cysteine; D-Cysteine | HMDB00574; HMDB03417 | M+H | - | - |
| HILIC positive | 795.6088 | 30.1 | -0.21 (1.19) | 0.12 (0.86) | -0.33 (-0.61, -0.05) | 0.0205 | 0.8047 | PC(20:3(5Z8Z11Z)P-18:1(11Z)); PC(20:3(5Z8Z11Z)P-18:1(9Z)); PC(20:3(8Z11Z14Z)P-18:1(11Z)); PC(20:3(8Z11Z14Z)P-18:1(9Z)); PC(20:4(5Z8Z11Z14Z)P-18:0); PC(20:4(8Z11Z14Z17Z)P-18:0); PC(22:4(7Z10Z13Z16Z)P-16:0); PC(P-16:022:4(7Z10Z13Z16Z)); PC(P-18:020:4(5Z8Z11Z14Z)); PC(P-18:020:4(8Z11Z14Z17Z)); PC(P-18:1(11Z)20:3(5Z8Z11Z)); PC(P-18:1(11Z)20:3(8Z11Z14Z)); PC(P-18:1(9Z)20:3(5Z8Z11Z)); PC(P-18:1(9Z)20:3(8Z11Z14Z)); PE(P-18:1(9Z)20:3(8Z11Z14Z)); PC(o-18:1(9Z)20:4(8Z11Z14Z17Z)) | HMDB08392; HMDB08393; HMDB08424; HMDB08425; HMDB08456; HMDB08489; HMDB08652; HMDB11226; HMDB11253; HMDB11254; HMDB11284; HMDB11285; HMDB11317; HMDB11318; HMDB11450; HMDB13432 | M+H_[+1] | - | - |
| HILIC positive | 205.1264 | 44.5 | -0.21 (1.47) | 0.14 (0.5) | -0.35 (-0.64, -0.05) | 0.0205 | 0.8047 | L-Acetylcarnitine; Hexyl 3-mercaptobutanoate; (S)-3-Mercaptohexyl butyrate; 3-Mercaptohexyl butyrate | HMDB00201; HMDB32327; HMDB37765; HMDB38948 | M+H_[+1]; M+H | - | - |
| HILIC positive | 140.0817 | 60.1 | 0.05 (0.81) | 0.25 (0.66) | -0.2 (-0.37, -0.03) | 0.0206 | 0.8047 | Histidinal | HMDB12234 | M+H | - | - |
| HILIC positive | 552.4273 | 27.1 | -0.16 (1.5) | 0.22 (0.63) | -0.37 (-0.69, -0.06) | 0.0207 | 0.8047 | 3-cis-Hydroxy-be-Caroten-3-one | HMDB02890 | M+H_[+1] | - | - |
| C18 negative | 313.1655 | 292.7 | -0.15 (1.18) | 0.16 (0.84) | -0.31 (-0.57, -0.05) | 0.0208 | 0.8915 | (-)-trans-Carveol glucoside; ()-trans-Carveol glucoside; Perilloside A | HMDB29848; HMDB29849; HMDB38706 | M-H | - | - |
| C18 negative | 420.9085 | 30.7 | -1.09 (0.75) | -0.93 (0.76) | -0.16 (-0.3, -0.03) | 0.0209 | 0.8915 | Benzbromarone | HMDB41834 | M-H | - | - |
| HILIC positive | 146.0922 | 115.9 | -0.13 (1.53) | 0.23 (0.53) | -0.36 (-0.66, -0.05) | 0.0213 | 0.8047 | 4-Guanidinobutanoic acid | HMDB03464 | M+H | - | - |
| C18 negative | 834.5752 | 221.6 | -0.07 (1.01) | -0.33 (1.04) | 0.26 (0.04, 0.48) | 0.0216 | 0.8915 | 3-O-Sulfogalactosylceramide (d18:120:0) | HMDB12315 | M-H | - | - |
| HILIC positive | 322.186 | 236 | 0.15 (0.85) | -0.15 (0.92) | 0.3 (0.04, 0.55) | 0.0216 | 0.8047 | Arginyl-Phenylalanine; Phenylalanyl-Arginine | HMDB28716; HMDB28989 | M+H | - | - |
| HILIC positive | 792.5895 | 29.6 | -0.29 (1.24) | 0.03 (0.75) | -0.32 (-0.58, -0.05) | 0.0218 | 0.8047 | PC(20:4(5Z8Z11Z14Z)P-18:1(11Z)); PC(20:4(5Z8Z11Z14Z)P-18:1(9Z)); PC(20:4(8Z11Z14Z17Z)P-18:1(11Z)); PC(20:4(8Z11Z14Z17Z)P-18:1(9Z)); PC(20:5(5Z8Z11Z14Z17Z)P-18:0); PC(22:5(4Z7Z10Z13Z16Z)P-16:0); PC(22:5(7Z10Z13Z16Z19Z)P-16:0); PC(P-16:022:5(4Z7Z10Z13Z16Z)); PC(P-16:022:5(7Z10Z13Z16Z19Z)); PC(P-18:020:5(5Z8Z11Z14Z17Z)); PC(P-18:1(11Z)20:4(5Z8Z11Z14Z)); PC(dm18:1(11Z)20:4(8Z11Z14Z17Z)); PC(P-18:1(9Z)20:4(5Z8Z11Z14Z)); PC(P-18:1(9Z)20:4(8Z11Z14Z17Z)); PC(o-16:022:6(4Z7Z10Z13Z16Z19Z)); 2-O-(4710131619-Docosahexaenoyl)-1-O-hexadecylglycero-3-phosphocholine | HMDB08457; HMDB08458; HMDB08490; HMDB08491; HMDB08522; HMDB08685; HMDB08718; HMDB11227; HMDB11228; HMDB11255; HMDB11286; HMDB11287; HMDB11319; HMDB11320; HMDB13409; HMDB39529 | M+H | - | - |
| HILIC positive | 158.0403 | 57.4 | -0.18 (0.57) | -0.07 (0.64) | -0.11 (-0.2, -0.02) | 0.0219 | 0.8047 | N-Methylethanolaminium phosphate | HMDB60173 | M+H_[+2] | - | - |
| C18 negative | 856.7194 | 46.9 | 0.07 (1.14) | -0.39 (1.71) | 0.45 (0.07, 0.84) | 0.0221 | 0.8915 | PC(24:0P-18:0); PC(P-18:024:0); PC(o-18:1(9Z)24:0); PC(o-22:020:1(11Z)) | HMDB08785; HMDB11263; HMDB13434; HMDB13447 | M-H | - | - |
| C18 negative | 140.011 | 15.8 | -0.3 (1.33) | 0 (1.18) | -0.3 (-0.56, -0.04) | 0.0222 | 0.8915 | 5-Hydroxymethyl-2-furanoate | HMDB29188 | M-H | - | - |
| HILIC positive | 280.1754 | 250.7 | 0.08 (0.74) | -0.28 (1.38) | 0.36 (0.05, 0.66) | 0.0224 | 0.8047 | Etamiphylline | HMDB41889 | M+H | - | - |
| C18 negative | 429.301 | 182.1 | -0.19 (1.42) | 0.16 (0.75) | -0.35 (-0.66, -0.05) | 0.0226 | 0.8915 | MG(0:024:6(6Z9Z12Z15Z18Z21Z)0:0); MG(24:6(6Z9Z12Z15Z18Z21Z)0:00:0); 7 alpha-Hydroxy-3- | HMDB11560; HMDB11590; HMDB12458; HMDB30066; | M-H | - | - |

| | | | | | | | | | | | | |
|---|---|---|---|---|---|---|---|---|---|---|---|---|
| | | | | | | | | oxo-4-cholestenoate; Australigenin; Barogenin; Schidigeragenin C; 25-Hydroxyvitamin D3-2623-lactol; 24-Oxo-1alpha25-dihydroxyvitamin D3 | HMDB34403; HMDB36249; HMDB60127; HMDB60128 | | | |
| HILIC positive | 183.138 | 25 | 0.2 (1.24) | -0.18 (0.93) | 0.38 (0.05, 0.7) | 0.0230 | 0.8047 | 2277-Tetramethyl-16-dioxaspiro44non-3-ene; Linalyl formate; alpha-Terpineol formate; 5-Pentyl-3h-furan-2-one; Methyl 48-decadienoate; Methyl (2E4Z)-decadienoate; Neryl formate; 4-Hydroxy-4-methyl-7-decenoic acid gamma-lactone; Ethyl octynecarboxylate; Tsibulin 2; 6-(3-Hexenyl)tetrahydro-2H-pyran-2-one; xi-2-Hexyl-5-methyl-3(2H)-furanone; (3E6Z)-Nonadien-1-yl acetate; (2E6Z)-26-Nonadien-1-Yl acetate; Bornyl formate; cis-3-Hexenyl tiglate; Allyl cyclohexylacetate; Methyl geranate | HMDB30006; HMDB30424; HMDB32050; HMDB32463; HMDB34708; HMDB34730; HMDB35156; HMDB36191; HMDB36215; HMDB37388; HMDB37829; HMDB38005; HMDB38068; HMDB38079; HMDB38245; HMDB38279; HMDB40595; HMDB59816 | M+H | - | - |
| HILIC positive | 193.0351 | 90.3 | -0.35 (1.31) | -0.07 (0.81) | -0.28 (-0.52, -0.04) | 0.0231 | 0.8047 | Citric acid; Isocitric acid; D-threo-Isocitric acid; Diketogulonic acid; 23-Diketo-L-gulonate; (1R2R)-Isocitric acid; D-Glucaro-14-lactone | HMDB00094; HMDB00193; HMDB01874; HMDB05971; HMDB06511; HMDB33717; HMDB41862 | M+H | - | - |
| HILIC positive | 218.0854 | 67.9 | -1.18 (0.67) | -1.06 (0.74) | -0.12 (-0.21, -0.02) | 0.0231 | 0.8047 | Captopril | HMDB15328 | M+H | - | - |
| HILIC positive | 178.0845 | 294.7 | -1.07 (0.49) | -0.88 (0.69) | -0.18 (-0.34, -0.03) | 0.0232 | 0.8047 | Alanyl-Serine; Glycyl-Threonine; Serinyl-Alanine; Threoninyl-Glycine | HMDB28696; HMDB28851; HMDB29032; HMDB29061 | M+H_[+1] | - | - |
| HILIC positive | 85.0648 | 163.5 | -0.04 (0.85) | 0.21 (0.85) | -0.25 (-0.47, -0.03) | 0.0234 | 0.8047 | Methyl propenyl ketone; 3-Methyl-2-butenal; Cyclopentanone; Tiglic aldehyde; 3-Methyl-3-buten-2-one; 2-Pentenal; 4-Penten-2-one; 1-Penten-3-one; 4-Pentenal; xi-23-Dihydro-3-methylfuran; 23-Dihydro-4-methylfuran | HMDB01184; HMDB12157; HMDB31407; HMDB31512; HMDB31530; HMDB31601; HMDB31606; HMDB31607; HMDB32458; HMDB37788; HMDB40584 | M+H | - | - |
| C18 negative | 477.3219 | 178.9 | -0.13 (1.18) | 0.16 (0.75) | -0.29 (-0.54, -0.04) | 0.0235 | 0.8915 | Dolicholide; Polyporusterone A | HMDB34086; HMDB38495 | M-H | - | - |
| C18 negative | 424.1976 | 106.5 | -0.16 (1.17) | 0.14 (0.78) | -0.3 (-0.56, -0.04) | 0.0235 | 0.8915 | O-Desmethyl-tramado glucuronide; NO-Didesmethylvenlafaxine glucuronide | HMDB60856; HMDB61167 | M-H | - | - |
| C18 negative | 1090.704 | 209.2 | -0.09 (1.15) | 0.22 (0.75) | -0.31 (-0.58, -0.04) | 0.0236 | 0.8915 | Ganglioside GA2 (d18:19Z-18:1) | HMDB04889 | M-H_[-1] | - | - |
| C18 negative | 181.0508 | 22.2 | -0.15 (1.25) | 0.17 (0.89) | -0.32 (-0.6, -0.04) | 0.0237 | 0.8915 | Homovanillic acid; Isohomovanillic acid; 34-Dihydroxyhydrocinnamic acid; Hydroxyphenyllactic acid; 3-(3-Hydroxyphenyl)-3-hydroxypropanoic acid; 3-Methoxy-4-hydroxyphenylglycolaldehyde; 3-Hydroxyphenyllactate; 26-Dimethoxybenzoic acid; 26-Dihydroxy-4-methoxyacetophenone; ()-2-Hydroxy-3-(2-hydroxyphenyl)propanoic acid; Maltol propionate; 24-Dihydroxy-6-methoxyacetophenone; 34-Dimethoxybenzoic acid | HMDB00118; HMDB00333; HMDB00423; HMDB00755; HMDB02643; HMDB04061; HMDB29232; HMDB29273; HMDB29646; HMDB33624; HMDB37274; HMDB41270; HMDB59763 | M-H | Hydroxyphenyllactate_homovanillate | M-H |
| C18 negative | 112.9857 | 35.1 | -0.16 (1.27) | 0.14 (0.8) | -0.3 (-0.57, -0.04) | 0.0237 | 0.8915 | Trifluoroacetic acid | HMDB14118 | M-H | - | - |
| C18 negative | 197.0818 | 156.5 | 0.16 (0.89) | -0.14 (1.04) | 0.3 (0.04, 0.56) | 0.0238 | 0.8915 | Guaifenesin; 5-(3E-Pentenyl)tetrahydro-2-oxo-3-furancarboxylic acid; 3-(4-Hydroxy-3-methoxyphenyl)-12-propanediol; (2E4E)-27-Dimethyl-24-octadienedioic acid; 34-Methylene suberic acid; 23-Methylene suberic acid | HMDB04998; HMDB30991; HMDB33093; HMDB34099; HMDB59768; HMDB59779 | M-H | - | - |
| HILIC positive | 636.5548 | 27.2 | -0.08 (1.2) | 0.24 (0.85) | -0.31 (-0.59, -0.04) | 0.0239 | 0.8047 | DG(14:1(9Z)22:2(13Z16Z)0:0); DG(16:020:3(5Z8Z11Z)0:0); DG(16:020:3(8Z11Z14Z)0:0); DG(16:1(9Z)20:2(11Z14Z)0:0); DG(18:018:3(6Z9Z12Z)0:0); DG(18:018:3(9Z12Z15Z)0:0); DG(18:1(11Z)18:2(9Z12Z)0:0); DG(18:1(9Z)18:2(9Z12Z)0:0); DG(18:2(9Z12Z)18:1(11Z)0:0); DG(18:2(9Z12Z)18:1(9Z)0:0); DG(18:3(6Z9Z12Z)18:00:0); DG(18:3(9Z12Z15Z)18:00:0); DG(20:2(11Z14Z)16:1(9Z)0:0); DG(20:3(5Z8Z11Z)16:00:0); DG(20:3(8Z11Z14Z)16:00:0); DG(22:2(13Z16Z)14:1(9Z)0:0); DG(16:00:020:3n9); DG(16:00:020:3n6); DG(18:00:018:3n6); DG(18:00:018:3n3); DG(14:1n50:022:2n6) | HMDB07059; HMDB07110; HMDB07111; HMDB07138; HMDB07162; HMDB07163; HMDB07190; HMDB07219; HMDB07246; HMDB07247; HMDB07274; HMDB07303; HMDB07418; HMDB07446; HMDB07475; HMDB07647; HMDB56019; HMDB56025; HMDB56049; HMDB56056; HMDB56147 | M+NH4 | - | - |
| HILIC positive | 147.1209 | 47.2 | -0.18 (1.38) | 0.14 (0.65) | -0.32 (-0.6, -0.04) | 0.0240 | 0.8047 | 3-Dehydroxycarnitine; Dibutyl sulfide; 2-Ethyl-1-hexanethiol | HMDB06831; HMDB31660; HMDB41477 | M+H_[+1]; M+H | - | - |
| HILIC positive | 188.1758 | 65.2 | -0.16 (1.35) | 0.19 (0.76) | -0.35 (-0.66, -0.05) | 0.0241 | 0.8047 | N1-Acetylspermidine; N8-Acetylspermidine; N(8)-Acetylspermidinium | HMDB01276; HMDB02189; HMDB60288 | M+H | - | - |
| C18 negative | 383.1532 | 24.5 | -0.09 (1.14) | 0.15 (0.64) | -0.25 (-0.46, -0.03) | 0.0241 | 0.8915 | 3b16a-Dihydroxyandrostenone sulfate | HMDB00386 | M-H | - | - |
| HILIC positive | 209.0129 | 175.8 | -0.55 (0.96) | -0.28 (0.96) | -0.27 (-0.51, -0.04) | 0.0242 | 0.8047 | (Z)-3-(Methylsulfinyl)-1-propenyl 2-propenyl disulfide | HMDB32653 | M+H | - | - |
| C18 negative | 292.1276 | 186.7 | -0.27 (1.44) | 0.08 (0.7) | -0.35 (-0.65, -0.05) | 0.0243 | 0.8915 | 1112-Dimethoxydihydrokawain; Coriandrone A; Coriandrone B | HMDB30767; HMDB37774; HMDB37775 | M-H_[-1] | - | - |
| C18 negative | 157.0366 | 166.2 | -0.11 (1.13) | 0.19 (0.75) | -0.3 (-0.56, -0.04) | 0.0248 | 0.8915 | Allantoin | HMDB00462 | M-H | - | - |
| HILIC positive | 171.0895 | 84.3 | 0.01 (0.75) | 0.19 (0.47) | -0.18 (-0.34, -0.02) | 0.0251 | 0.8047 | 1-Methylhistidine; 3-Methylhistidine | HMDB00001; HMDB00479 | M+H_[+1] | - | - |

| | | | | | | | | | | | | |
|---|---|---|---|---|---|---|---|---|---|---|---|---|
| HILIC positive | 749.5302 | 28.2 | -0.21 (1.24) | 0.1 (0.81) | -0.31 (-0.58, -0.04) | 0.0252 | 0.8047 | PG(16:018:1(11Z)); PG(16:018:1(9Z)); PG(16:1(9Z)18:0); PG(18:016:1(9Z)); PG(18:1(11Z)16:0); PG(18:1(9Z)16:0) | HMDB10573; HMDB10574; HMDB10587; HMDB10601; HMDB10615; HMDB10630 | M+H | - | - |
| HILIC positive | 232.1543 | 38.3 | -0.24 (1.54) | 0.13 (0.66) | -0.37 (-0.69, -0.05) | 0.0253 | 0.8047 | Isobutyryl-L-carnitine; Butyrylcarnitine | HMDB00736; HMDB02013 | M+H | Butyrylcar nitine | M+H |
| C18 negative | 423.275 | 90.1 | 0.3 (0.48) | 0.11 (0.66) | 0.19 (0.02, 0.36) | 0.0254 | 0.8915 | 1b-Hydroxycholic acid; 3b4b7a12a-Tetrahydroxy-5b-cholanoic acid; 2b3a7a12a-Tetrahydroxy-5b-cholanoic acid; 3a7a12a19-Tetrahydroxy-5b-cholanoic acid; 3a6b7b12a-Tetrahydroxy-5b-cholanoic acid; 3a6b7b12b-Tetrahydroxy-5b-cholanoic acid; 3a6b7a12a-Tetrahydroxy-5b-cholanoic acid; 13712-Tetrahydroxycholan-24-oic acid; 3a4b7a12a-Tetrahydroxy-5b-cholanoic acid; 1b3a7b-Trihydroxy-5b-cholanoic acid; 3a7b21-Trihydroxy-5b-cholanoic acid | HMDB00307; HMDB00311; HMDB00316; HMDB00340; HMDB00367; HMDB00377; HMDB00399; HMDB00433; HMDB00437; HMDB13158; HMDB13192 | M-H | - | - |
| C18 negative | 405.1917 | 107.5 | -0.22 (1.36) | 0.11 (0.71) | -0.33 (-0.62, -0.04) | 0.0257 | 0.8915 | (1R3R4R5S6S8x)-1-Acetoxy-8-angeloyloxy-34-epoxy-5-hydroxy-7(14)10-bisaboladien-2-one; 1-(4-Hydroxy-35-dimethoxyphenyl)-7-(4-hydroxy-3-methoxyphenyl)-35-heptanediol | HMDB39971; HMDB41091 | M-H | - | - |
| C18 negative | 127.1128 | 295 | -0.29 (1.95) | 0.17 (0.56) | -0.46 (-0.86, -0.06) | 0.0259 | 0.8915 | Octanal; 25-Diethyltetrahydrofuran; (R)-Sulcatol; 2-Octanone; Ethyl pentyl ketone; (E)-2-Octen-1-ol; (Z)-5-Octen-1-ol; (R)-1-Octen-3-ol; ()-(E)-2-Octen-4-ol; 2-Methyl-3-heptanone; trans-3-Octen-2-ol; (Z)-3-Octen-1-ol; (E)-3-Octen-1-ol; xi-7-Octen-2-ol | HMDB01140; HMDB29574; HMDB30030; HMDB31294; HMDB31295; HMDB31296; HMDB31298; HMDB31299; HMDB31300; HMDB32402; HMDB32450; HMDB35231; HMDB35768; HMDB37181 | M-H | - | - |
| HILIC positive | 259.1905 | 287.9 | -0.47 (1.5) | -0.13 (1.16) | -0.34 (-0.64, -0.04) | 0.0260 | 0.8047 | Tetradecanedioic acid; Diethyl decanedioate; Diisobutyl adipate; 2-Hydroxypropyl 2-isopropyl-5-methylcyclohexyl carbonate; 1-Hydroxypropan-2-yl 2-isopropyl-5-methylcyclohexyl carbonate | HMDB00872; HMDB40429; HMDB41618; HMDB59946; HMDB59947 | M+H | - | - |
| C18 negative | 197.0431 | 25.1 | -0.28 (1.47) | 0.08 (0.73) | -0.36 (-0.67, -0.04) | 0.0264 | 0.8915 | 4-Methyldibenzothiophene | HMDB40164 | M-H | - | - |
| HILIC positive | 844.7368 | 105.3 | -0.03 (0.92) | 0.23 (0.68) | -0.26 (-0.48, -0.03) | 0.0266 | 0.8047 | SM(d18:126:0) | HMDB11698 | M+H | - | - |
| HILIC positive | 243.1079 | 182.6 | 0.04 (0.92) | 0.27 (0.83) | -0.22 (-0.42, -0.03) | 0.0268 | 0.8047 | Histidinyl-Serine; Serinyl-Histidine; D-erythro-D-galacto-octitol; (SS)-Nt-Histidinylalanine; (SS)-Np-Histidinylalanine | HMDB28894; HMDB29041; HMDB29953; HMDB40248; HMDB40249 | M+H | - | - |
| HILIC positive | 286.2376 | 244.4 | 0.12 (0.79) | -0.17 (1.17) | 0.29 (0.03, 0.54) | 0.0270 | 0.8047 | Myristoylglycine | HMDB13250 | M+H | - | - |
| HILIC positive | 463.3411 | 27.2 | 0.34 (0.85) | 0.56 (0.65) | -0.22 (-0.42, -0.03) | 0.0271 | 0.8047 | Dolichosterone; Polyporusterone F | HMDB34336; HMDB38499 | M+H | - | - |
| C18 negative | 114.9899 | 35.1 | -0.16 (1.23) | 0.16 (0.89) | -0.33 (-0.62, -0.04) | 0.0273 | 0.8915 | Trifluoroacetic acid | HMDB14118 | M-H_[+2] | - | - |
| C18 negative | 384.2391 | 149.2 | -0.03 (0.96) | 0.2 (0.47) | -0.23 (-0.43, -0.03) | 0.0275 | 0.8915 | Buspirone | HMDB14633 | M-H | - | - |
| HILIC positive | 287.2366 | 27.1 | -0.23 (1.41) | 0.09 (0.93) | -0.32 (-0.6, -0.04) | 0.0276 | 0.8047 | Vitamin A; 11-cis-Retinol; 9-cis-Retinol; 13-cis Retinol; (x)-p-Menth-1-en-4-yl 5-isopropyl-2-methylphenyl ether; Abietinal; ent-16-Kauren-19-al; ent-19-Trachylobanal; 813-Abietadien-18-al | HMDB00305; HMDB06216; HMDB06217; HMDB06221; HMDB30403; HMDB34735; HMDB36728; HMDB36841; HMDB38709 | M+H | - | - |
| HILIC positive | 229.1546 | 51.3 | -0.08 (1.01) | 0.14 (0.71) | -0.22 (-0.41, -0.02) | 0.0278 | 0.8047 | L-isoleucyl-L-proline ; L-leucyl-L-proline ; Isoleucyl-Proline; Leucyl-Proline | HMDB11174; HMDB11175; HMDB28915; HMDB28937 | M+H | - | - |
| HILIC positive | 247.1732 | 37.5 | -0.18 (1.41) | 0.16 (0.8) | -0.34 (-0.65, -0.04) | 0.0278 | 0.8047 | 2-Methylbutyroylcarnitine; Isovalerylcarnitine; Valerylcarnitine; Pivaloylcarnitine; (S)-3-Methylthiohexyl hexanoate | HMDB00378; HMDB00688; HMDB13128; HMDB41993; HMDB38947 | M+H_[+1]; M+H | - | - |
| HILIC positive | 368.1549 | 244.3 | 0.18 (0.51) | -0.01 (0.91) | 0.19 (0.02, 0.37) | 0.0280 | 0.8047 | N-Acetyl-6-O-L-fucosyl-D-glucosamine; 2-Acetamido-2-deoxy-6-O-a-L-fucopyranosyl-D-glucose; 3-O-fucopyranosyl-2-acetamido-2-deoxyglucopyranose; Casuarine 6-alpha-D-glucoside | HMDB02220; HMDB05817; HMDB06700; HMDB31999 | M+H | - | - |
| HILIC positive | 835.6022 | 30.2 | -0.24 (1.23) | 0.04 (0.94) | -0.28 (-0.53, -0.03) | 0.0282 | 0.8047 | PC(18:022:6(4Z7Z10Z13Z16Z19Z)); PC(18:1(11Z)22:5(4Z7Z10Z13Z16Z)); PC(18:1(11Z)22:5(7Z10Z13Z16Z19Z)); PC(18:1(9Z)22:5(4Z7Z10Z13Z16Z)); PC(18:1(9Z)22:5(7Z10Z13Z16Z19Z)); PC(18:2(9Z12Z)22:4(7Z10Z13Z16Z)); PC(18:4(6Z9Z12Z15Z)22:2(13Z16Z)); PC(20:1(11Z)20:5(5Z8Z11Z14Z17Z)); PC(20:2(11Z14Z)20:4(5Z8Z11Z14Z)); PC(20:2(11Z14Z)20:4(8Z11Z14Z17Z)); PC(20:3(5Z8Z11Z)20:3(5Z8Z11Z)); PC(20:3(5Z8Z11Z)20:3(8Z11Z14Z)); PC(20:3(8Z11Z14Z)20:3(5Z8Z11Z)); PC(20:3(8Z11Z14Z)20:3(8Z11Z14Z)); PC(20:4(5Z8Z11Z14Z)20:2(11Z14Z)); | HMDB08057; HMDB08088; HMDB08089; HMDB08121; HMDB08122; HMDB08153; HMDB08251; HMDB08314; HMDB08345; HMDB08346; HMDB08376; HMDB08377; HMDB08408; HMDB08409; HMDB08440; HMDB08473; HMDB08505; HMDB08601; HMDB08631; HMDB08662; HMDB08663; HMDB08695; HMDB08696; HMDB08727 | M+H_[+1] | - | - |

| | | | | | | | | | | | | |
|---|---|---|---|---|---|---|---|---|---|---|---|---|
| | | | | | | | | PC(20:4(8Z11Z14Z17Z)20:2(11Z14Z)); PC(20:5(5Z8Z11Z14Z17Z)20:1(11Z)); PC(22:2(13Z16Z)18:4(6Z9Z12Z15Z)); PC(22:4(7Z10Z13Z16Z)18:2(9Z12Z)); PC(22:5(4Z7Z10Z13Z16Z)18:1(11Z)); PC(22:5(4Z7Z10Z13Z16Z)18:1(9Z)); PC(22:5(7Z10Z13Z16Z19Z)18:1(11Z)); PC(22:5(7Z10Z13Z16Z19Z)18:1(9Z)); PC(22:6(4Z7Z10Z13Z16Z19Z)18:0) | | | | |
| HILIC positive | 146.1176 | 45.6 | -0.14 (1.25) | 0.15 (0.74) | -0.29 (-0.55, -0.03) | 0.0282 | 0.8047 | 3-Dehydroxycarnitine | HMDB06831 | M+H | Acetylcholine | M+H |
| C18 negative | 200.041 | 22.8 | -0.2 (1.23) | 0.08 (0.86) | -0.28 (-0.53, -0.03) | 0.0286 | 0.8915 | Potassium 2-(1-ethoxy) ethoxypropanoate | HMDB37192 | M-H_[-1] | - | - |
| HILIC positive | 399.33 | 27.1 | -0.16 (1.16) | 0.1 (0.94) | -0.26 (-0.5, -0.03) | 0.0289 | 0.8047 | trans-Hexadec-2-enoyl carnitine; 9-Hexadecenoylcarnitine | HMDB06317; HMDB13207 | M+H_[+1] | - | - |
| HILIC positive | 161.1285 | 73.2 | 0.01 (0.63) | 0.17 (0.63) | -0.16 (-0.3, -0.02) | 0.0291 | 0.8047 | N(6)-Methyllysine; Isoputreanine | HMDB02038; HMDB06009 | M+H | - | - |
| HILIC positive | 160.0605 | 297.7 | 0.06 (0.33) | 0.17 (0.47) | -0.11 (-0.2, -0.01) | 0.0292 | 0.8047 | L-2-Amino-4-methylenepentanedioic acid; L-trans-alpha-Amino-2-carboxycyclopropaneacetic acid | HMDB29433; HMDB34442 | M+H | - | - |
| C18 negative | 168.0244 | 20.4 | -0.13 (1.04) | 0.14 (0.92) | -0.28 (-0.53, -0.03) | 0.0292 | 0.8915 | Uric acid | HMDB00289 | M-H_[-1] | - | - |
| HILIC positive | 132.0565 | 74.5 | -0.18 (1.07) | 0.06 (0.91) | -0.24 (-0.45, -0.02) | 0.0293 | 0.8047 | Pyroglutamic acid; Pyrrolidonecarboxylic acid; Pyrroline hydroxycarboxylic acid; N-Acryloylglycine; 1-Pyrroline-4-hydroxy-2-carboxylate; 5-Oxoprolinate; dimethadione | HMDB00267; HMDB00805; HMDB01369; HMDB01843; HMDB02234; HMDB60262; HMDB61093 | M+H_[+2] | - | - |
| HILIC positive | 263.0661 | 92.7 | 0.48 (0.61) | 0.32 (0.7) | 0.17 (0.02, 0.32) | 0.0295 | 0.8047 | Cinoxacin; 5-Fluorouridine | HMDB14965; HMDB60396 | M+H | - | - |
| HILIC positive | 222.1124 | 179.3 | -0.18 (1.2) | 0.12 (0.68) | -0.3 (-0.56, -0.03) | 0.0296 | 0.8047 | Metaxalone; Carbofuran; Hydrocotarnine | HMDB14798; HMDB31770; HMDB33701 | M+H | - | - |
| HILIC positive | 189.1121 | 235.7 | 0.13 (0.82) | -0.22 (1.26) | 0.35 (0.03, 0.67) | 0.0303 | 0.8047 | Azelaic acid; Nonate; cis- and trans-Ethyl 24-dimethyl-13-dioxolane-2-acetate; (-)-Ethyl 3-acetoxy-2-methylbutyrate; (-)-Methyl 5-acetoxyhexanoate; Butyl ethyl malonate; 24-Dimethylpimelic acid; 3-Methylsuberic acid; Diethyl methylsuccinate; Diethyl glutarate | HMDB00784; HMDB11717; HMDB32200; HMDB32258; HMDB32387; HMDB36233; HMDB59760; HMDB59783; HMDB59814; HMDB59879 | M+H | - | - |
| C18 negative | 129.0557 | 24.8 | -0.28 (1.39) | 0.05 (0.76) | -0.33 (-0.62, -0.03) | 0.0305 | 0.8915 | 2-Methyl-3-ketovaleric acid; 3-Methyl-2-oxovaleric acid; Ketoleucine; 2-Ketohexanoic acid; Mevalonolactone; 3-Oxohexanoic acid; Adipate semialdehyde; 5-Ethoxy-45-dihydro-2(3H)furanone; Ethyl acetoacetate; Ethyl 3-oxobutanoate; Sherry lactone; (4S6S)-3456-Tetrahydro-4-hydroxy-6-methyl-2H-pyran-2-one; Acetoin acetate; Methyl levulinate; Pantolactone | HMDB00408; HMDB00491; HMDB00695; HMDB01864; HMDB06024; HMDB10717; HMDB12882; HMDB31214; HMDB31216; HMDB31218; HMDB33286; HMDB33302; HMDB33698; HMDB41602; HMDB59876 | M-H | - | - |
| C18 negative | 861.5478 | 226.6 | -0.06 (1.11) | 0.21 (0.98) | -0.27 (-0.52, -0.03) | 0.0306 | 0.8915 | PI(16:020:2(11Z14Z)); PI(18:018:2(9Z12Z)); PI(18:1(11Z)18:1(11Z)); PI(18:1(11Z)18:1(9Z)); PI(18:1(9Z)18:1(11Z)); PI(18:1(9Z)18:1(9Z)); PI(18:2(9Z12Z)18:0); PI(20:2(11Z14Z)16:0) | HMDB09786; HMDB09809; HMDB09824; HMDB09825; HMDB09836; HMDB09837; HMDB09847; HMDB09875 | M-H | - | - |
| HILIC positive | 501.3932 | 26.8 | 0.03 (1.08) | 0.29 (0.79) | -0.26 (-0.5, -0.03) | 0.0306 | 0.8047 | Norambreinolide; 3-Methyl-alpha-ionyl acetate; 22-Bis(2-methylpropoxy)ethylbenzene; Ganoderiol C | HMDB35293; HMDB37631; HMDB37712; HMDB37781 | 2M+H; M+H-H2O | - | - |
| C18 negative | 221.0667 | 294.9 | -0.16 (1.21) | 0.12 (0.81) | -0.28 (-0.53, -0.03) | 0.0306 | 0.8915 | Ethyl glucuronide | HMDB10325 | M-H | - | - |
| HILIC positive | 482.3236 | 37.2 | 0.13 (0.91) | -0.15 (1.22) | 0.28 (0.03, 0.54) | 0.0309 | 0.8047 | LysoPC(15:0); LysoPE(0:018:0); LysoPE(18:00:0) | HMDB10381; HMDB11129; HMDB11130 | M+H | - | - |
| C18 negative | 524.2781 | 191.5 | -0.14 (0.89) | 0.08 (0.85) | -0.22 (-0.42, -0.02) | 0.0310 | 0.8915 | LysoPE(0:022:6(4Z7Z10Z13Z16Z19Z)); LysoPE(22:6(4Z7Z10Z13Z16Z19Z)0:0) | HMDB11496; HMDB11526 | M-H | - | - |
| C18 negative | 201.1497 | 166.7 | 0.12 (1.26) | -0.21 (0.98) | 0.33 (0.03, 0.64) | 0.0312 | 0.8915 | 3-Hydroxynonyl acetate | HMDB32443 | M-H | - | - |
| C18 negative | 138.031 | 294.5 | -0.18 (1.19) | 0.14 (0.83) | -0.32 (-0.61, -0.03) | 0.0314 | 0.8915 | 5-Hydroxypyrazinamide | HMDB01978 | M-H | - | - |
| HILIC positive | 89.0418 | 99.4 | 0.57 (0.49) | 0.39 (0.75) | 0.18 (0.02, 0.34) | 0.0323 | 0.8047 | 3-(Methylthio)-1-propene; cis-23-Dimethylthiirane; (1E)-1-(methylthio)prop-1-ene | HMDB31653; HMDB39807; HMDB59843 | M+H | - | - |
| C18 negative | 131.0462 | 21.7 | -0.1 (1.19) | 0.22 (0.84) | -0.33 (-0.63, -0.03) | 0.0324 | 0.8915 | Ureidopropionic acid; L-Asparagine; Glycyl-glycine; N-Carbamoylsarcosine; D-Asparagine | HMDB00026; HMDB00168; HMDB11733; HMDB12265; HMDB33780 | M-H | Ureidopropionate | M-H |
| C18 negative | 229.1443 | 295.8 | 0.16 (0.66) | -0.1 (1.09) | 0.26 (0.02, 0.49) | 0.0328 | 0.8915 | Dodecanedioic acid; Talaromycin A; Bis(1-methylethyl) hexanedioate; Dipropyl hexanedioate | HMDB00623; HMDB30143; HMDB41613; HMDB41614 | M-H | - | - |
| C18 negative | 191.1077 | 294.3 | 0.12 (0.95) | -0.16 (1.01) | 0.28 (0.02, 0.54) | 0.0331 | 0.8915 | 2-Methyl-1-phenyl-2-propanyl acetate; 4-Phenyl-2-butyl acetate; Ethyl 4-phenylbutanoate; Benzyl 3-methylbutanoate; 3-Methylbutyl benzoate; 2-Methylpropyl phenylacetate; 2-Phenylethyl butanoate; 2-Phenylethyl 2-methylpropanoate; 5-Isopropyl-2-methylphenol acetate; 3-Phenylpropyl propanoate; 111213-Trinor-1(10)-spirovetivene-27-dione; 4-Methyl-2-(1-phenylethyl)-13-dioxolane 9CI; 4-Methylphenyl 3-methylbutanoate; 1-Phenylethyl isobutyrate; 1-Phenylethyl butyrate; 1-Ethoxy-2-methoxy-4-(1-propenyl)benzene; 2-(2- | HMDB31571; HMDB31614; HMDB31618; HMDB32043; HMDB33380; HMDB35010; HMDB35014; HMDB35015; HMDB35447; HMDB36388; HMDB36734; HMDB37165; HMDB37709; HMDB37716; HMDB37717; HMDB37792; HMDB38180; HMDB39496; | M-H | - | - |

| | | | | | | | | | | | | |
|---|---|---|---|---|---|---|---|---|---|---|---|---|
| | | | | | | | | Hydroxy-4-methylphenyl)-3-pentanone; 2-Ethoxy-1-methoxy-4-(1-propenyl)benzene; 111213-Trinor-135-bisabolatrien-10-oic acid; 2-Benzyl-45-dimethyl-13-dioxolane; Butyl phenylacetate | HMDB39643; HMDB39841; HMDB40427 | | | |
| HILIC positive | 290.1961 | 274.1 | 0.12 (0.9) | -0.15 (1.03) | 0.27 (0.02, 0.52) | 0.0332 | 0.8047 | Dihydro-246-tris(2-methylpropyl)-4h-135-dithiazine | HMDB32221 | M+H | - | - |
| HILIC positive | 378.7914 | 26.1 | -0.13 (1.04) | 0.16 (0.72) | -0.29 (-0.56, -0.02) | 0.0336 | 0.8047 | ()-Naled | HMDB31806 | M+H | - | - |
| HILIC positive | 180.1593 | 30.1 | 0.16 (1.09) | -0.06 (0.87) | 0.22 (0.02, 0.43) | 0.0340 | 0.8047 | (3R7R)-137-Octanetriol | HMDB33625 | M+NH4 | - | - |
| C18 negative | 167.021 | 20.5 | -0.13 (1.04) | 0.14 (0.92) | -0.27 (-0.52, -0.02) | 0.0340 | 0.8915 | Uric acid | HMDB00289 | M-H | Urate | M-H |
| C18 negative | 183.1025 | 290.5 | -0.16 (1.08) | 0.13 (1.01) | -0.3 (-0.57, -0.02) | 0.0346 | 0.8915 | (1R)-Nepetalic acid; (S)-Oleuropeic acid; (4S8R)-89-Dihydroxy-p-menth-1(6)-en-2-one; 1-Acetylcyclohexyl acetate; (E)-10-Oxo-8-decenoic acid | HMDB36117; HMDB36998; HMDB39052; HMDB39710; HMDB40883 | M-H | - | - |
| HILIC positive | 597.4484 | 25.7 | -0.09 (1.23) | 0.17 (0.78) | -0.27 (-0.51, -0.02) | 0.0346 | 0.8047 | DG(15:018:4(6Z9Z12Z15Z)0:0); DG(18:4(6Z9Z12Z15Z)15:00:0); DG(15:00:018:4n3) | HMDB07077; HMDB07329; HMDB56004 | M+Na | - | - |
| HILIC positive | 235.1321 | 26.9 | 0.17 (0.96) | -0.15 (1.26) | 0.33 (0.02, 0.63) | 0.0349 | 0.8047 | (10S11S)-Pterosin C; Dehydrooreadone; (S)-Pterosin P; Pterosin N | HMDB30763; HMDB36046; HMDB36607; HMDB38218 | M+H | - | - |
| HILIC positive | 834.5977 | 31.6 | -0.23 (1.21) | 0.04 (0.95) | -0.27 (-0.51, -0.02) | 0.0349 | 0.8047 | PC(18:022:6(4Z7Z10Z13Z16Z19Z)); PC(18:1(11Z)22:5(4Z7Z10Z13Z16Z)); PC(18:1(11Z)22:5(7Z10Z13Z16Z19Z)); PC(18:1(9Z)22:5(4Z7Z10Z13Z16Z)); PC(18:1(9Z)22:5(7Z10Z13Z16Z19Z)); PC(18:2(9Z12Z)22:4(7Z10Z13Z16Z)); PC(18:4(6Z9Z12Z15Z)22:2(13Z16Z)); PC(20:1(11Z)20:5(5Z8Z11Z14Z17Z)); PC(20:2(11Z14Z)20:4(5Z8Z11Z14Z)); PC(20:2(11Z14Z)20:4(8Z11Z14Z17Z)); PC(20:3(5Z8Z11Z)20:3(5Z8Z11Z)); PC(20:3(5Z8Z11Z)20:3(8Z11Z14Z)); PC(20:3(8Z11Z14Z)20:3(5Z8Z11Z)); PC(20:3(8Z11Z14Z)20:3(8Z11Z14Z)); PC(20:4(5Z8Z11Z14Z)20:2(11Z14Z)); PC(20:4(8Z11Z14Z17Z)20:2(11Z14Z)); PC(20:5(5Z8Z11Z14Z17Z)20:1(11Z)); PC(22:2(13Z16Z)18:4(6Z9Z12Z15Z)); PC(22:4(7Z10Z13Z16Z)18:2(9Z12Z)); PC(22:5(4Z7Z10Z13Z16Z)18:1(11Z)); PC(22:5(4Z7Z10Z13Z16Z)18:1(9Z)); PC(22:5(7Z10Z13Z16Z19Z)18:1(11Z)); PC(22:5(7Z10Z13Z16Z19Z)18:1(9Z)); PC(22:6(4Z7Z10Z13Z16Z19Z)18:0) | HMDB08057; HMDB08088; HMDB08089; HMDB08121; HMDB08122; HMDB08153; HMDB08251; HMDB08314; HMDB08345; HMDB08346; HMDB08376; HMDB08377; HMDB08408; HMDB08409; HMDB08440; HMDB08473; HMDB08505; HMDB08601; HMDB08631; HMDB08662; HMDB08663; HMDB08695; HMDB08696; HMDB08727 | M+H | - | - |
| C18 negative | 315.2327 | 229.9 | -0.05 (0.99) | -0.28 (1.03) | 0.23 (0.02, 0.45) | 0.0351 | 0.8915 | Pregnenolone; 20a-Dihydroprogesterone; 5a-Pregnane-320-dione; Calusterone ; Ethyltestosterone; Norbolethone; Bolasterone; (3beta5alpha)-3-Hydroxypregn-16-en-20-one; 1-Phenyl-13-pentadecanedione; Kaurenoic acid methyl ester; 5-(811-Pentadecadienyl)-13-benzenediol; 5-alpha-Pregnan-320-dione | HMDB00253; HMDB03069; HMDB03759; HMDB04627; HMDB06002; HMDB06026; HMDB06048; HMDB34369; HMDB35580; HMDB36729; HMDB38534; HMDB59652 | M-H | - | - |
| C18 negative | 408.2107 | 106.5 | -0.25 (1.47) | 0.09 (0.77) | -0.34 (-0.66, -0.02) | 0.0352 | 0.8915 | Cascarillin; Anhydrocinnzeylanine | HMDB36836; HMDB36864 | M-H_[-1] | - | - |
| C18 negative | 375.1812 | 141.4 | -0.25 (1.2) | 0.06 (1.02) | -0.3 (-0.58, -0.02) | 0.0352 | 0.8915 | 18-Oxocortisol; 111214-Trihydroxy-7-methoxy-81113-abietatrien-206-olide; 12alpha-Hydroxy-1318-dehydroparain; 16alpha;-hydroxyprednisolone | HMDB00332; HMDB31932; HMDB39557; HMDB61016 | M-H | - | - |
| C18 negative | 387.2023 | 132.9 | -0.07 (1.17) | 0.2 (0.56) | -0.27 (-0.52, -0.02) | 0.0352 | 0.8915 | 5a6a-Epoxy-7E-megastigmene-3a9e-diol 3-glucoside; 913-Dihydroxy-4-megastigmen-3-one 9-glucoside; Icariside B8; 5a6a-Epoxy-7E-megastigmene-3b9e-diol 9-glucoside; Dihydroroseoside | HMDB31676; HMDB36318; HMDB36846; HMDB38306; HMDB40614 | M-H | - | - |
| HILIC positive | 427.361 | 27.1 | -0.11 (1.01) | 0.15 (0.86) | -0.26 (-0.5, -0.02) | 0.0352 | 0.8047 | Oleoylcarnitine; Vaccenyl carnitine; Elaidic carnitine; 11Z-Octadecenylcarnitine | HMDB05065; HMDB06351; HMDB06464; HMDB13338 | M+H_[+1] | - | - |
| HILIC positive | 203.1391 | 52.2 | -0.13 (1.27) | 0.15 (0.64) | -0.29 (-0.55, -0.02) | 0.0352 | 0.8047 | Alanyl-Isoleucine; Alanyl-Leucine; Isoleucyl-Alanine; Leucyl-Alanine | HMDB28690; HMDB28691; HMDB28900; HMDB28922 | M+H | - | - |
| HILIC positive | 251.2005 | 26.9 | 0.04 (1.3) | 0.29 (0.75) | -0.25 (-0.49, -0.02) | 0.0352 | 0.8047 | Norambreinolide; 3-Methyl-alpha-ionyl acetate; 22-Bis(2-methylpropoxy)ethylbenzene | HMDB35293; HMDB37631; HMDB37712 | M+H | - | - |
| HILIC positive | 283.1904 | 232.2 | -0.13 (1.07) | 0.16 (0.95) | -0.28 (-0.54, -0.02) | 0.0359 | 0.8047 | Lactapiperanol C | HMDB33630 | M+H | - | - |
| HILIC positive | 205.1587 | 285.5 | -0.13 (1.23) | 0.17 (0.82) | -0.3 (-0.58, -0.02) | 0.0359 | 0.8047 | 24-Di-tert-butylphenol; (6R7S)-67-Epoxy-13-tetradecadiyne; 2-Benzylidene-1-heptanol; 3-Benzyl-4-heptanone | HMDB13816; HMDB31776; HMDB36170; HMDB41482 | M+H | - | - |
| C18 negative | 153.0921 | 295.7 | 0.16 (0.59) | -0.08 (1.03) | 0.24 (0.02, 0.47) | 0.0360 | 0.8915 | Methyl 2-octynoate; 24-Hexadienyl propionate; 2-Nonenoic acid gamma-lactone; (E)-26-Dimethyl-25-heptadienoic acid; 355-Trimethyl-12-cyclohexanedione; Enol-355-Trimethyl-12-cyclohexanedione; Octahydro-2H-1-benzopyran-2-one; Allyl | HMDB31302; HMDB32314; HMDB32442; HMDB35135; HMDB36177; HMDB36239; HMDB37276; HMDB38268; | M-H | - | - |

| | | | | | | | | | | | | |
|---|---|---|---|---|---|---|---|---|---|---|---|---|
| | | | | | | | | hexenoate; xi-226-Trimethyl-14-cyclohexanedione; Anapear; 4-Oxo-2-nonenal | HMDB38697; HMDB40285; HMDB60285 | | | |
| HILIC positive | 110.0601 | 183.5 | -0.21 (1.39) | 0.11 (0.69) | -0.32 (-0.61, -0.02) | 0.0366 | 0.8047 | 4-Aminophenol; 2-Acetylpyrrole; 1-Methyl-2-pyrrolecarboxaldehyde | HMDB01169; HMDB35882; HMDB40582 | M+H | - | - |
| HILIC positive | 380.2572 | 34.7 | -0.21 (1.27) | 0.06 (0.86) | -0.27 (-0.53, -0.02) | 0.0367 | 0.8047 | Sphingosine 1-phosphate; N-palmitoyl-phosphoethanolamine | HMDB00277; HMDB59630 | M+H | - | - |
| C18 negative | 202.9976 | 20.9 | -0.11 (1.07) | 0.16 (0.91) | -0.26 (-0.51, -0.02) | 0.0368 | 0.8915 | Uric acid | HMDB00289 | M+Cl | - | - |
| C18 negative | 525.2817 | 191.4 | -0.15 (0.93) | 0.08 (0.89) | -0.23 (-0.44, -0.01) | 0.0369 | 0.8915 | LysoPE(0:022:6(4Z7Z10Z13Z16Z19Z)); LysoPE(22:6(4Z7Z10Z13Z16Z19Z)0:0); Desmosine; Isodesmosine | HMDB11496; HMDB11526; HMDB00572; HMDB00739 | M-H_[-1]; M-H | - | - |
| C18 negative | 475.3796 | 266.7 | -0.03 (1.08) | 0.22 (0.77) | -0.25 (-0.48, -0.01) | 0.0372 | 0.8915 | Panaxatriol | HMDB38244 | M-H | - | - |
| C18 negative | 213.1131 | 292.7 | 0.11 (0.92) | -0.12 (0.99) | 0.23 (0.01, 0.45) | 0.0373 | 0.8915 | 5-Hexyltetrahydro-2-oxo-3-furancarboxylic acid; alpha-Carboxy-delta-decalactone; 2-Carboxy-57-dimethyl-4-octanolide | HMDB30984; HMDB30985; HMDB30986 | M-H | - | - |
| HILIC positive | 269.2474 | 19.7 | -0.14 (0.89) | 0.13 (0.86) | -0.28 (-0.54, -0.02) | 0.0375 | 0.8047 | Cyclohexaneundecanoic acid; (Z)-9-Heptadecenoic acid; 24-Heptadecanedione; 10Z-Heptadecenoic acid | HMDB30997; HMDB31046; HMDB40888; HMDB60038 | M+H | - | - |
| HILIC positive | 277.1796 | 27.9 | -0.14 (1.09) | 0.12 (0.76) | -0.26 (-0.5, -0.01) | 0.0377 | 0.8047 | Cyclandelate; 8-Shogaol; Panaquinquecol 2; Ginsenoyne C; Ginsenoyne K; Panaquinquecol 7 | HMDB15586; HMDB31463; HMDB38938; HMDB38994; HMDB40375; HMDB41201 | M+H | - | - |
| C18 negative | 189.0114 | 23.8 | -0.31 (1.44) | 0.02 (0.79) | -0.33 (-0.64, -0.02) | 0.0379 | 0.8915 | p-Cresol sulfate; Benzyl sulfate | HMDB11635; HMDB34782 | M-H_[+2] | - | - |
| HILIC positive | 813.6829 | 36 | -0.15 (1.09) | 0.11 (1.1) | -0.26 (-0.51, -0.01) | 0.0380 | 0.8047 | SM(d18:124:1(15Z)) | HMDB12107 | M+H | - | - |
| C18 negative | 485.3121 | 142.4 | 0.18 (0.62) | 0.35 (0.55) | -0.16 (-0.32, -0.01) | 0.0381 | 0.8915 | Hovenine A | HMDB30200 | M-H | - | - |
| C18 negative | 212.002 | 24.1 | -0.1 (0.79) | 0.14 (0.75) | -0.24 (-0.48, -0.01) | 0.0385 | 0.8915 | Indoxyl sulfate | HMDB00682 | M-H | - | - |
| C18 negative | 315.05 | 19.3 | -1.16 (0.88) | -0.99 (0.76) | -0.17 (-0.33, -0.01) | 0.0387 | 0.8915 | Isorhamnetin; 3457-Tetrahydroxy-6-methoxyflavone; 2-(34-Dihydroxyphenyl)-56-dihydroxy-7-methoxy-4H-1-benzopyran-4-one; Pollenitin; 1358-Tetrahydroxy-6-methoxy-2-methylanthraquinone; Malaoxon | HMDB02655; HMDB30547; HMDB30804; HMDB38572; HMDB39210; HMDB60627 | M-H; M-H_[+2] | - | - |
| HILIC positive | 559.4711 | 26.8 | 0.01 (1.28) | 0.28 (0.79) | -0.27 (-0.52, -0.01) | 0.0387 | 0.8047 | DG(15:018:3(6Z9Z12Z)0:0); DG(15:018:3(9Z12Z15Z)0:0); DG(18:3(6Z9Z12Z)15:00:0); DG(18:3(9Z12Z15Z)15:00:0); DG(15:00:018:3n6); DG(15:00:018:3n3); DG(14:1n50:018:2n6); DG(14:1n50:020:2n6) | HMDB07075; HMDB07076; HMDB07271; HMDB07300; HMDB55996; HMDB56003; HMDB56142; HMDB56144 | M+H-H2O | - | - |
| C18 negative | 257.2121 | 156.3 | 0.17 (0.88) | -0.1 (1.03) | 0.27 (0.01, 0.53) | 0.0388 | 0.8915 | 10-Hydroxymyristic acid methyl ester; Ethyl 3-hydroxytridecanoate | HMDB31061; HMDB59866 | M-H | - | - |
| HILIC positive | 793.5929 | 29.6 | -0.26 (1.17) | 0.02 (0.79) | -0.29 (-0.56, -0.01) | 0.0388 | 0.8047 | PC(20:4(5Z8Z11Z14Z)P-18:1(11Z)); PC(20:4(5Z8Z11Z14Z)P-18:1(9Z)); PC(20:4(8Z11Z14Z17Z)P-18:1(11Z)); PC(20:4(8Z11Z14Z17Z)P-18:1(9Z)); PC(20:5(5Z8Z11Z14Z17Z)P-18:0); PC(22:5(4Z7Z10Z13Z16Z)P-16:0); PC(22:5(7Z10Z13Z16Z19Z)P-16:0); PC(P-16:022:5(4Z7Z10Z13Z16Z)); PC(P-16:022:5(7Z10Z13Z16Z19Z)); PC(P-18:020:5(5Z8Z11Z14Z17Z)); PC(P-18:1(11Z)20:4(5Z8Z11Z14Z)); PC(dm18:1(11Z)20:4(8Z11Z14Z17Z)); PC(P-18:1(9Z)20:4(5Z8Z11Z14Z)); PC(P-18:1(9Z)20:4(8Z11Z14Z17Z)); PC(o-16:022:6(4Z7Z10Z13Z16Z19Z)); 2-O-(4710131619-Docosahexaenoyl)-1-O-hexadecylglycero-3-phosphocholine | HMDB08457; HMDB08458; HMDB08490; HMDB08491; HMDB08522; HMDB08685; HMDB08718; HMDB11227; HMDB11228; HMDB11255; HMDB11286; HMDB11287; HMDB11319; HMDB11320; HMDB13409; HMDB39529 | M+H_[+1] | - | - |
| HILIC positive | 190.1186 | 72.2 | -0.03 (1) | 0.18 (0.45) | -0.21 (-0.41, -0.01) | 0.0391 | 0.8047 | Homocitrulline | HMDB00679 | M+H | - | - |
| C18 negative | 112.0517 | 26.1 | 0 (0.65) | 0.11 (0.62) | -0.11 (-0.21, -0.01) | 0.0392 | 0.8915 | Creatinine | HMDB00562 | M-H | - | - |
| C18 negative | 289.2019 | 153.3 | 0.16 (0.74) | -0.14 (1.28) | 0.29 (0.01, 0.58) | 0.0395 | 0.8915 | Glycerol 1-(5-hydroxydodecanoate) | HMDB40168 | M-H | - | - |
| HILIC positive | 661.4802 | 24.9 | -0.05 (0.99) | 0.18 (0.86) | -0.24 (-0.47, -0.01) | 0.0395 | 0.8047 | DG(18:4(6Z9Z12Z15Z)22:6(4Z7Z10Z13Z16Z19Z)0:0); DG(20:5(5Z8Z11Z14Z17Z)20:5(5Z8Z11Z14Z17Z)0:0); DG(22:6(4Z7Z10Z13Z16Z19Z)18:4(6Z9Z12Z15Z)0:0); DG(18:4n30:022:6n3); DG(20:5n30:020:5n3) | HMDB07353; HMDB07578; HMDB07773; HMDB56376; HMDB56381 | M+H | - | - |
| HILIC positive | 214.0174 | 98.6 | -0.15 (1.11) | 0.1 (0.99) | -0.26 (-0.5, -0.01) | 0.0397 | 0.8047 | Indoxyl sulfate | HMDB00682 | M+H | - | - |
| C18 negative | 475.3062 | 182.4 | -0.17 (1.54) | 0.17 (0.71) | -0.34 (-0.67, -0.02) | 0.0402 | 0.8915 | Polyporusterone B; Polyporusterone C | HMDB38496; HMDB38497 | M-H | - | - |
| HILIC positive | 369.2828 | 27.8 | -0.05 (0.96) | 0.17 (0.65) | -0.23 (-0.45, -0.01) | 0.0409 | 0.8047 | 3 5-Tetradecadiencarnitine | HMDB13331 | M+H_[+1] | - | - |
| C18 negative | 481.1642 | 171.4 | 0.47 (0.91) | 0.23 (1) | 0.23 (0.01, 0.45) | 0.0410 | 0.8915 | Melleolide D | HMDB37042 | M-H | - | - |
| C18 negative | 591.39 | 200.8 | -0.09 (1.19) | 0.17 (0.76) | -0.26 (-0.51, -0.01) | 0.0416 | 0.8915 | Tuberoside | HMDB39054 | M-H | - | - |
| HILIC positive | 426.3575 | 27.1 | -0.1 (0.96) | 0.15 (0.89) | -0.25 (-0.49, -0.01) | 0.0416 | 0.8047 | Oleoylcarnitine; Vaccenyl carnitine; Elaidic carnitine; 11Z-Octadecenylcarnitine | HMDB05065; HMDB06351; HMDB06464; HMDB13338 | M+H | - | - |
| C18 negative | 335.0345 | 20.4 | -1.15 (0.89) | -0.98 (0.78) | -0.17 (-0.34, -0.01) | 0.0416 | 0.8915 | Malaoxon | HMDB60627 | M+Na-2H | - | - |
| C18 negative | 423.074 | 214.2 | 0.09 (1) | 0.37 (0.93) | -0.28 (-0.54, -0.01) | 0.0417 | 0.8915 | BL V | HMDB39567 | M-H | - | - |
| C18 negative | 585.2624 | 183.7 | -0.09 (0.93) | 0.11 (0.62) | -0.2 (-0.39, -0.01) | 0.0421 | 0.8915 | Bilirubin; 4E15Z-Bilirubin IXa | HMDB00054; HMDB00488 | M-H_[+2] | - | - |
| C18 negative | 227.0924 | 294.2 | -0.04 (0.79) | 0.17 (0.8) | -0.22 (-0.43, -0.01) | 0.0423 | 0.8915 | Propyl propane thiosulfonate | HMDB32496 | M-H | - | - |

| | | | | | | | | | | | | |
|---|---|---|---|---|---|---|---|---|---|---|---|---|
| C18 negative | 204.9954 | 21.9 | -0.21 (1.36) | 0.13 (0.92) | -0.34 (-0.66, -0.01) | 0.0424 | 0.8915 | O-methoxycatechol-O-sulphate | HMDB60013 | M-H_[+2] | - | - |
| HILIC positive | 398.2323 | 27.6 | 0.11 (0.79) | -0.15 (1.39) | 0.26 (0.01, 0.51) | 0.0424 | 0.8047 | Drotaverine | HMDB15669 | M+H | - | - |
| C18 negative | 862.5536 | 227.6 | -0.05 (1.09) | 0.2 (0.99) | -0.25 (-0.5, -0.01) | 0.0424 | 0.8915 | PI(16:020:2(11Z14Z)); PI(18:018:2(9Z12Z)); PI(18:1(11Z)18:1(11Z)); PI(18:1(11Z)18:1(9Z)); PI(18:1(9Z)18:1(11Z)); PI(18:1(9Z)18:1(9Z)); PI(18:2(9Z12Z)18:0); PI(20:2(11Z14Z)16:0) | HMDB09786; HMDB09809; HMDB09824; HMDB09825; HMDB09836; HMDB09837; HMDB09847; HMDB09875 | M-H_[-1] | - | - |
| C18 negative | 277.1646 | 156.5 | 0.13 (0.69) | -0.18 (1.34) | 0.31 (0.01, 0.61) | 0.0425 | 0.8915 | (x)-2-Heptanol glucoside | HMDB35028 | M-H | - | - |
| HILIC positive | 315.2358 | 30 | -0.09 (1.08) | 0.14 (0.59) | -0.23 (-0.46, -0.01) | 0.0430 | 0.8047 | 9-Decenoylcarnitine | HMDB13205 | M+H_[+1] | - | - |
| C18 negative | 143.035 | 25.2 | 0.19 (1.23) | -0.11 (0.92) | 0.3 (0.01, 0.59) | 0.0433 | 0.8915 | 2-Hydroxyadipic acid; 3-Hydroxyadipic acid; 3-Hydroxymethylglutaric acid; 2(R)-Hydroxyadipic acid; Levoglucosan; D-1-Deoxy-erythro-hexo-23-diulose; Diethyl dicarbonate; D-15-Anhydrofructose; 2-Hydroxy-2-ethylsuccinic acid; 3-Hexenedioic acid; 3-Methylglutaconic acid; (E)-2-Methylglutaconic acid; trans-2-Hexenedioic acid; 3-Hydroxyadipic acid 36-lactone; Dimethyl fumarate; Maleic acid homopolymer; Ethyl hydrogen fumarate | HMDB00321; HMDB00345; HMDB00355; HMDB00368; HMDB00640; HMDB29934; HMDB32873; HMDB41561; HMDB59758; HMDB00393; HMDB00522; HMDB02266; HMDB13311; HMDB29171; HMDB31257; HMDB36232; HMDB40207 | M-H2O-H; M-H | - | - |
| C18 negative | 149.1335 | 102.3 | -0.21 (1.11) | 0.07 (0.95) | -0.29 (-0.56, -0.01) | 0.0435 | 0.8915 | Cystophorene; (E)-48-Dimethyl-137-nonatriene | HMDB30944; HMDB35792 | M-H | - | - |
| HILIC positive | 231.0207 | 94.4 | 0.52 (0.36) | 0.42 (0.5) | 0.1 (0, 0.19) | 0.0441 | 0.8047 | Guanabenz | HMDB14767 | M+H | - | - |
| C18 negative | 168.018 | 20.7 | -0.13 (1.08) | 0.14 (0.91) | -0.27 (-0.53, -0.01) | 0.0444 | 0.8915 | Uric acid | HMDB00289 | M-H_[-1] | - | - |
| HILIC positive | 728.5586 | 28.8 | -0.45 (1.59) | -0.07 (1.23) | -0.38 (-0.75, -0.01) | 0.0448 | 0.8047 | PE(18:1(11Z)P-18:1(11Z)); PE(18:1(11Z)P-18:1(9Z)); PE(18:1(9Z)P-18:1(11Z)); PE(18:1(9Z)P-18:1(9Z)); PE(18:2(9Z12Z)P-18:0); PE(20:2(11Z14Z)P-16:0); PE(P-16:020:2(11Z14Z)); PE(P-18:018:2(9Z12Z)); PE(P-18:1(11Z)18:1(11Z)); PE(P-18:1(11Z)18:1(9Z)); PE(P-18:1(9Z)18:1(11Z)); PE(P-18:1(9Z)18:1(9Z)) | HMDB09050; HMDB09051; HMDB09083; HMDB09084; HMDB09115; HMDB09312; HMDB11349; HMDB11376; HMDB11407; HMDB11408; HMDB11440; HMDB11441 | M+H | - | - |
| C18 negative | 476.3096 | 183 | -0.17 (1.51) | 0.16 (0.74) | -0.33 (-0.66, -0.01) | 0.0449 | 0.8915 | Polyporusterone B; Polyporusterone C; Gentamicin | HMDB38496; HMDB38497; HMDB14936 | M-H_[-1]; M-H | - | - |
| C18 negative | 329.066 | 18.9 | -1.16 (0.86) | -1 (0.75) | -0.16 (-0.32, 0) | 0.0454 | 0.8915 | 37-Dimethylquercetin; Aflatoxin G2; Aflatoxin M2; Demethoxysudachitin; Prudomestin; 34-Dihydroxyphenacyl caffeate; 135-Trihydroxy-67-dimethoxy-2-methylanthraquinone; Trifolian; Aflatoxin B2a; Pilosin; Caryatin; 34-Di-O-methylquercetin; Cicerin; 57-Dihydroxy-84-dimethoxyisoflavone | HMDB29263; HMDB30475; HMDB30480; HMDB30696; HMDB30811; HMDB31706; HMDB33268; HMDB34009; HMDB35598; HMDB37096; HMDB37351; HMDB37354; HMDB38659; HMDB41690 | M-H | - | - |
| HILIC positive | 601.4808 | 25.4 | -0.39 (1.43) | -0.05 (1.27) | -0.34 (-0.67, -0.01) | 0.0455 | 0.8047 | DG(15:020:5(5Z8Z11Z14Z17Z)0:0); DG(20:5(5Z8Z11Z14Z17Z)15:00:0); DG(15:00:020:5n3) | HMDB07085; HMDB07561; HMDB56006 | M+H | - | - |
| HILIC positive | 224.0668 | 22.6 | -0.03 (0.9) | 0.17 (0.45) | -0.2 (-0.4, 0) | 0.0456 | 0.8047 | 1-(1-Propenylthio)propyl propyl disulfide; (E)-1-Propenyl 1-(propylthio)propyl disulfide; 224466-Hexamethyl-135-trithiane; 246-Triethyl-135-trithiane | HMDB33041; HMDB33072; HMDB40243; HMDB40262 | M+H_[+1] | - | - |
| HILIC positive | 225.1485 | 292.3 | -0.18 (1.58) | 0.14 (0.69) | -0.32 (-0.64, -0.01) | 0.0456 | 0.8047 | Epoxyeremopetasinorol; Annuionone A; Annuionone C; 13-Oxo-911-tridecadienoic acid; Methyl jasmonate; (8alpha10beta11beta)-3-Hydroxy-415-dinor-1(5)-xanthen-128-olide; Octyl 2-furoate; Furfuryl octanoate; 3-Methylbutyl 2-furanbutanoate | HMDB29670; HMDB32679; HMDB32689; HMDB34564; HMDB36583; HMDB37244; HMDB37726; HMDB37730; HMDB37736 | M+H | - | - |
| HILIC positive | 227.1642 | 290.4 | -0.13 (1.31) | 0.16 (0.77) | -0.29 (-0.58, -0.01) | 0.0460 | 0.8047 | Methyl dihydrojasmonate; Riesling acetal; 45-Dihydrovomifoliol | HMDB31740; HMDB37562; HMDB40615 | M+H | - | - |
| HILIC positive | 161.0961 | 296.4 | 0.1 (0.91) | -0.27 (1.65) | 0.37 (0.01, 0.74) | 0.0466 | 0.8047 | 3-Methyl-4-phenyl-3-buten-2-one; 2-Phenyl-4-pentenal; 3-Phenyl-4-pentenal | HMDB31567; HMDB35207; HMDB38790 | M+H | - | - |
| HILIC positive | 103.548 | 93.4 | -0.11 (0.69) | 0.06 (0.59) | -0.17 (-0.34, 0) | 0.0467 | 0.8047 | Cycloserine; 3-Amino-2-oxazolidone | HMDB14405; HMDB60739 | M+H_[+1] | - | - |
| C18 negative | 314.1225 | 146.9 | -0.11 (1.18) | 0.14 (0.81) | -0.25 (-0.5, 0) | 0.0468 | 0.8915 | Neosaxitoxin; N-Hydroxysaxitoxin | HMDB29369; HMDB33664 | M-H | - | - |
| C18 negative | 240.099 | 294 | 0.35 (0.44) | 0.19 (0.72) | 0.16 (0, 0.33) | 0.0469 | 0.8915 | 5-Methyldeoxycytidine; 3-Amino-3-deoxythimidine | HMDB02224; HMDB60750 | M-H | - | - |
| HILIC positive | 286.2376 | 27.8 | -0.21 (0.97) | 0.09 (1.2) | -0.3 (-0.59, 0) | 0.0469 | 0.8047 | Myristoylglycine | HMDB13250 | M+H | - | - |
| HILIC positive | 351.1091 | 291.4 | -1.01 (0.99) | -0.8 (0.99) | -0.2 (-0.4, 0) | 0.0474 | 0.8047 | 3-Feruloyl-15-quinolactone; 4-Feruloyl-15-quinolactone | HMDB29289; HMDB29290 | M+H | - | - |
| HILIC positive | 101.0597 | 297.4 | 0.15 (0.49) | -0.18 (1.57) | 0.33 (0, 0.65) | 0.0474 | 0.8047 | Senecioic acid; Tiglic acid; 2-Ethylacrylic acid; 3-Methylbutyrolactone; Glutaral; Angelic acid; Dihydro-2-methyl-3(2H)-furanone; 23-Pentanedione; 4-Pentenoic acid; 24-Pentanedione; Isopropenyl acetate; Methyl methacrylate; 2-Pentenoic acid; Dihydro-5-methyl-2(3H)-furanone; Ethyl acrylate; Dihydro-3-methyl-2(3H)-furanone | HMDB00509; HMDB01470; HMDB01862; HMDB02167; HMDB29599; HMDB29608; HMDB31178; HMDB31598; HMDB31602; HMDB31648; HMDB32351; HMDB32385; HMDB32459; HMDB33840; HMDB33978; HMDB35143 | M+H | - | - |
| HILIC positive | 230.0959 | 69 | -0.03 (0.92) | 0.17 (0.76) | -0.2 (-0.39, 0) | 0.0475 | 0.8047 | Ergothioneine | HMDB03045 | M+H | - | - |
| HILIC positive | 202.1437 | 30 | -0.28 (1.16) | 0 (0.89) | -0.28 (-0.56, 0) | 0.0475 | 0.8047 | Capryloylglycine; Valproylglycine; N-(5-Methyl-3-oxohexyl)alanine; N-Acetylaminooctanoic acid | HMDB00832; HMDB13116; HMDB41540; HMDB59745 | M+H | - | - |

| | | | | | | | | | | | | |
|---|---|---|---|---|---|---|---|---|---|---|---|---|
| HILIC positive | 125.1325 | 294 | 0.16 (0.56) | -0.19 (1.66) | 0.35 (0, 0.69) | 0.0479 | 0.8047 | 26-Dimethyl-24-heptadiene; Isobutylene-isoprene copolymer; 2-Isopropyl-14-hexadiene | HMDB31415; HMDB32344; HMDB37777 | M+H | - | - |
| HILIC positive | 162.0761 | 94.9 | 0.12 (0.45) | 0.22 (0.28) | -0.11 (-0.22, 0) | 0.0482 | 0.8047 | Aminoadipic acid; Acetylhomoserine; ()-22-Iminobispropanoic acid | HMDB00510; HMDB29423; HMDB33747 | M+H | Alpha-aminoadipate | M+H |
| C18 negative | 493.2802 | 127 | 0.18 (0.88) | 0.41 (0.74) | -0.23 (-0.46, 0) | 0.0485 | 0.8915 | (3b4b11b14b)-11-Ethoxy-34-epoxy-14-hydroxy-12-cyathen-15-al 14-xyloside | HMDB34617 | M-H | - | - |
| HILIC positive | 315.2528 | 25.9 | -0.27 (1.5) | 0.09 (1.16) | -0.36 (-0.72, 0) | 0.0486 | 0.8047 | Octadecanedioic acid; 910-DHOME; 1213-DHOME; (9xi10xi12xi)-910-Dihydroxy-12-octadecenoic acid; Dibutyl decanedioate | HMDB00782; HMDB04704; HMDB04705; HMDB31679; HMDB41220 | M+H | - | - |
| C18 negative | 357.2645 | 215.7 | 0.13 (0.89) | -0.08 (0.71) | 0.22 (0, 0.43) | 0.0488 | 0.8915 | 1314-Dihydro PGF-1a | HMDB05076 | M-H | - | - |
| HILIC positive | 517.3881 | 26.8 | 0.08 (1.08) | 0.32 (0.82) | -0.25 (-0.49, 0) | 0.0490 | 0.8047 | 22-Acetylpriverogenin B | HMDB34645 | M+H | - | - |
| HILIC positive | 149.0558 | 94.5 | -0.28 (1.58) | -0.02 (1.02) | -0.26 (-0.51, 0) | 0.0490 | 0.8047 | Hydroxylated lecithin | HMDB32332 | M+H | - | - |
| HILIC positive | 146.06 | 42.6 | -0.24 (1.41) | 0.06 (0.79) | -0.3 (-0.59, 0) | 0.0490 | 0.8047 | 1H-Indole-3-carboxaldehyde | HMDB29737 | M+H | - | - |
| HILIC positive | 314.2325 | 30.1 | -0.06 (0.98) | 0.15 (0.6) | -0.21 (-0.41, 0) | 0.0490 | 0.8047 | 9-Decenoylcarnitine | HMDB13205 | M+H | - | - |
| HILIC positive | 85.0284 | 75.8 | -0.08 (1.18) | 0.19 (0.75) | -0.27 (-0.55, 0) | 0.0491 | 0.8047 | 4-Hydroxy-2-butenoic acid gamma-lactone | HMDB32330 | M+H | - | - |
| HILIC positive | 697.2156 | 46.1 | -0.74 (1.15) | -0.5 (1.11) | -0.24 (-0.47, 0) | 0.0494 | 0.8047 | Licorice glycoside B; Licorice glycoside D1 | HMDB31988; HMDB31994 | M+H | - | - |
| C18 negative | 283.2279 | 210.1 | -0.02 (0.5) | 0.12 (0.53) | -0.14 (-0.28, 0) | 0.0494 | 0.8915 | Avocadyne; Muricatacin | HMDB35473; HMDB38685 | M-H | - | - |
| C18 negative | 174.056 | 25 | -0.28 (1.5) | 0.09 (0.97) | -0.37 (-0.73, 0) | 0.0495 | 0.8915 | Indoleacetic acid; 5-Hydroxyindoleacetaldehyde; Hydroxymethyl indol-3-yl ketone; 1-Methoxy-1H-indole-3-carboxaldehyde | HMDB00197; HMDB04073; HMDB38628; HMDB40972 | M-H | Indoleacetate | M-H |
| C18 negative | 257.1757 | 178.2 | -0.21 (1.6) | 0.14 (0.73) | -0.35 (-0.7, 0) | 0.0496 | 0.8915 | Tetradecanedioic acid; Diethyl decanedioate; Diisobutyl adipate; 2-Hydroxypropyl 2-isopropyl-5-methylcyclohexyl carbonate; 1-Hydroxypropan-2-yl 2-isopropyl-5-methylcyclohexyl carbonate | HMDB00872; HMDB40429; HMDB41618; HMDB59946; HMDB59947 | M-H | - | - |
| C18 negative | 570.2769 | 155.3 | 0.18 (0.72) | -0.07 (1.03) | 0.25 (0, 0.51) | 0.0498 | 0.8915 | Ganoderic acid F; Kinetensin 4-7 | HMDB35988; HMDB12986 | M-H_[-1]; M-H | - | - |

SD: Standard Deviation; CI: Confidence Interval; FDR: False Discovery Rate.

**Supplemental Table 6.** Results of the Untargeted Metabolome-wide Association Study Comparing TB Resisters and Controls in the South African Cohort (11 Pairs). Metabolites with P Value < 0.05 and Level 1 or 2 Confirmation are Shown.

| Platform | m/z | time | Resister Mean (SD) | Control Mean (SD) | Difference Resister vs. Control (95% CI) | P | FDR | Level 2 Confirmation | | | Level 1 Confirmation | |
|---|---|---|---|---|---|---|---|---|---|---|---|---|
| | | | | | | | | Name | HMDB ID | Adduct | Name | Adduct |
| HILIC positive | 132.0565 | 74.5 | -0.21 (0.93) | 0.88 (0.83) | -1.09 (-1.51, -0.67) | 0.0002 | 0.5612 | Pyroglutamic acid; Pyrrolidonecarboxylic acid; Pyrroline hydroxycarboxylic acid; N-Acryloylglycine; 1-Pyrroline-4-hydroxy-2-carboxylate; 5-Oxoprolinate; dimethadione | HMDB00267; HMDB00805; HMDB01369; HMDB01843; HMDB02234; HMDB60262; HMDB61093 | M+H_[+2] | - | - |
| HILIC positive | 424.342 | 27.2 | 0.65 (0.84) | -0.22 (0.37) | 0.87 (0.41, 1.33) | 0.0018 | 0.8637 | Linoelaidyl carnitine; Linoleyl carnitine | HMDB06461; HMDB06469 | M+H | - | - |
| HILIC positive | 425.3453 | 27.3 | 0.62 (0.76) | -0.18 (0.36) | 0.8 (0.38, 1.23) | 0.0018 | 0.8637 | Linoelaidyl carnitine; Linoleyl carnitine | HMDB06461; HMDB06469 | M+H_[+1] | - | - |
| C18 negative | 495.1851 | 134 | -0.55 (0.45) | 0.48 (0.73) | -1.03 (-1.57, -0.48) | 0.0019 | 0.8508 | (7R8R)-3-Methoxy-34799-pentahydroxy-84-oxyneolignan 4-xyloside | HMDB40325 | M-H | - | - |
| C18 negative | 411.202 | 133.6 | -0.52 (0.87) | 0.43 (0.96) | -0.95 (-1.47, -0.43) | 0.0022 | 0.8508 | 5-(23-Dihydroxy-3-methylbutyl)-4-(34-epoxy-4-methylpentanoyl)-34-dihydroxy-2-isopentanoyl-2-cyclopenten-1-one; Abscisic alcohol 11-glucoside | HMDB30082; HMDB39636 | M-H | - | - |
| HILIC positive | 233.0632 | 250.2 | 0.17 (0.84) | -0.53 (1.08) | 0.71 (0.31, 1.1) | 0.0026 | 0.8637 | (2E11Z)-5-5-(Methylthio)-4-penten-2-ynyl-2-furanacrolein | HMDB31070 | M+H | - | - |
| C18 negative | 469.3346 | 196.2 | 0.54 (0.44) | 0.02 (0.22) | 0.52 (0.22, 0.83) | 0.0035 | 0.8508 | Glycyrrhetinic acid ; Murrayenol; (3alpha20R24Z)-3-Hydroxy-21-oxoeupha-824-dien-26-oic acid; 6beta-Hydroxy-3-oxo-12-oleanen-28-oic acid; beta-Glycyrrhetinic acid; 23-Hydroxy-3-oxocycloart-24-en-26-oic acid; 3beta-324-Dihydroxy-9(11)12-oleanadien-30-oic acid; Ganoderiol B; Ganodermic acid Jb; Liquiritic acid; Pomonic acid; Rubinic acid; 11-Keto-beta-boswellic acid; Lansic acid; Colubrinic acid; Koetjapic acid; 28-Hydroxymangiferonic acid; 2-Hydroxy-3-oxo-12-oleanen-28-oic acid; 16-Hydroxy-3-oxo-12-oleanen-28-oic acid; Secoisobryononic acid; Secobryononic acid | HMDB11628; HMDB29345; HMDB33213; HMDB34508; HMDB34517; HMDB34568; HMDB35252; HMDB35327; HMDB35334; HMDB35788; HMDB35912; HMDB36655; HMDB36671; HMDB36786; HMDB36847; HMDB38653; HMDB40415; HMDB40494; HMDB40618; HMDB40995; HMDB40996 | M-H | - | - |
| HILIC positive | 742.5741 | 30.8 | -0.42 (0.94) | 0.4 (0.55) | -0.83 (-1.32, -0.33) | 0.0039 | 0.8637 | PC(16:1(9Z)P-18:1(11Z)); PC(16:1(9Z)P-18:1(9Z)); PC(18:2(9Z12Z)P-16:0); PC(P-16:018:2(9Z12Z)); PC(P-18:1(11Z)16:1(9Z)); PC(P-18:1(9Z)16:1(9Z)); PC(o-16:1(9Z)18:2(9Z12Z)) | HMDB08029; HMDB08030; HMDB08159; HMDB11211; HMDB11273; HMDB11306; HMDB13413 | M+H | - | - |
| HILIC positive | 132.0542 | 75 | -0.25 (0.81) | 0.79 (0.67) | -1.04 (-1.68, -0.4) | 0.0048 | 0.8637 | Pipecolic acid; Pyroglutamic acid; L-Pipecolic acid; Pyrrolidonecarboxylic acid; Pyrroline hydroxycarboxylic acid; N-Acryloylglycine; 1-Pyrroline-4-hydroxy-2-carboxylate; N4-Acetylaminobutanal; D-Pipecolic acid; Vigabatrin; L-trans-4-Methyl-2-pyrrolidinecarboxylic acid; 2-Pyrrolidineacetic acid; 3-Acetamidobutanal; 5-Oxoprolinate; dimethadione | HMDB00070; HMDB00267; HMDB00716; HMDB00805; HMDB01369; HMDB01843; HMDB02234; HMDB04226; HMDB05960; HMDB15212; HMDB29435; HMDB29444; HMDB59649; HMDB60262; HMDB61093 | M+H_[+2] | - | - |
| C18 negative | 412.2056 | 133.8 | -0.38 (0.71) | 0.36 (0.83) | -0.74 (-1.21, -0.28) | 0.0050 | 0.8508 | 5-(23-Dihydroxy-3-methylbutyl)-4-(34-epoxy-4-methylpentanoyl)-34-dihydroxy-2-isopentanoyl-2-cyclopenten-1-one; Abscisic alcohol 11-glucoside | HMDB30082; HMDB39636 | M-H_[-1] | - | - |
| HILIC positive | 288.1665 | 86.1 | 0.17 (0.38) | 0.52 (0.41) | -0.35 (-0.56, -0.13) | 0.0050 | 0.8637 | Arginyl-Hydroxyproline; Hydroxyprolyl-Arginine | HMDB28710; HMDB28857 | M+H | - | - |
| C18 negative | 193.0869 | 155.7 | -0.2 (0.34) | 0.24 (0.37) | -0.43 (-0.7, -0.16) | 0.0051 | 0.8508 | (R)-3-Hydroxy-5-phenylpentanoic acid; 25-Dimethoxy-4-(2-propenyl)phenol; Butylparaben; Zingerone; Ethyl 4-methylphenoxyacetate; 4-Methoxybenzyl propanoate; Isobutyl salicylate; 26-Dimethoxy-4-(1-propenyl)phenol; Butyl salicylate; Methoxyeugenol; 5-Hydroxy-2-benzyl-13-dioxane; 2-Benzyl-5-hydroxymethyl-13-dioxolane; 3-Hydroxyphenyl-valeric acid; Ethyl 4-ethoxybenzoate; 2-(2-Methylphenyl)-13-dioxolan-4-ylmethanol; 2-(3-Methylphenyl)-13-dioxolan-4-ylmethanol; 2-(4-Methylphenyl)-13-dioxolan-4-ylmethanol; 2-(2-Methylphenyl)-13-dioxan-5-ol; 2-(3-Methylphenyl)-13-dioxan-5-ol; 2-(4-Methylphenyl)-13-dioxan-5-ol | HMDB31517; HMDB31984; HMDB32575; HMDB32590; HMDB34206; HMDB34990; HMDB36432; HMDB37271; HMDB40730; HMDB41194; HMDB41505; HMDB41506; HMDB41666; HMDB59898; HMDB59940; HMDB59941; HMDB59942; HMDB59943; HMDB59944; HMDB59945 | M-H | - | - |
| C18 negative | 411.2393 | 166.9 | -1.02 (1.26) | 0.55 (0.71) | -1.57 (-2.55, -0.59) | 0.0052 | 0.8508 | LysoPC(10:0) | HMDB03752 | M-H | - | - |
| HILIC positive | 343.1234 | 88.6 | 0.45 (0.91) | -0.63 (0.78) | 1.07 (0.4, 1.74) | 0.0052 | 0.8637 | Melibiose; Cellobiose; D-Maltose; Alpha-Lactose; Sucrose; Lactulose; Trehalose; Isomaltose; Galactinol; 3-b-Galactopyranosyl glucose; Epimelibiose; Turanose; Kojibiose; Neotrehalose; Sakebiose; Inulobiose; Galabiose; Maltulose; Mannobiose; DEAE-cellulose; Allolactose; Glucinol; | HMDB00048; HMDB00055; HMDB00163; HMDB00186; HMDB00258; HMDB00740; HMDB00975; HMDB02923; HMDB05826; HMDB06603; | M+H | - | - |

| | | | | | | | | | | | | |
|---|---|---|---|---|---|---|---|---|---|---|---|---|
| | | | | | | | | Trehalulose; Fagopyritol A1; beta-Lactose; Glucose-13-mannose oligosaccharide | HMDB06792; HMDB11740; HMDB11742; HMDB29880; HMDB29882; HMDB29898; HMDB29902; HMDB29919; HMDB29933; HMDB33368; HMDB38489; HMDB39237; HMDB39727; HMDB39876; HMDB41627; HMDB60068 | | | |
| C18 negative | 173.0567 | 19.8 | -0.19 (0.88) | 0.53 (0.55) | -0.72 (-1.18, -0.26) | 0.0057 | 0.8508 | Formiminoglutamic acid; N-Acetylasparagine | HMDB00854; HMDB06028 | M-H | - | - |
| C18 negative | 169.087 | 288.3 | -0.56 (0.72) | 0.32 (0.87) | -0.88 (-1.45, -0.3) | 0.0067 | 0.8508 | 14-Ipomeadiol; cis-3-Hexenyl pyruvate | HMDB30471; HMDB38276 | M-H | - | - |
| HILIC positive | 426.3575 | 27.1 | 0.62 (0.75) | -0.29 (0.55) | 0.91 (0.31, 1.51) | 0.0070 | 0.8637 | Oleoylcarnitine; Vaccenyl carnitine; Elaidic carnitine; 11Z-Octadecenylcarnitine | HMDB05065; HMDB06351; HMDB06464; HMDB13338 | M+H | - | - |
| C18 negative | 363.2549 | 166.1 | -0.28 (1.16) | 0.89 (0.14) | -1.17 (-1.94, -0.4) | 0.0071 | 0.8508 | 6-O-Acetylaustroinulin; 7-O-Acetylaustroinulin | HMDB36803; HMDB36804 | M-H | - | - |
| HILIC positive | 508.3396 | 37 | -0.62 (1.11) | 0.51 (0.62) | -1.14 (-1.89, -0.39) | 0.0071 | 0.8637 | LysoPE(0:020:1(11Z)); LysoPE(20:1(11Z)0:0); Gymnodimine | HMDB11482; HMDB11512; HMDB41430 | M+H | - | - |
| HILIC positive | 373.3141 | 27.7 | 0.42 (0.26) | -0.02 (0.28) | 0.44 (0.15, 0.74) | 0.0075 | 0.8637 | Tetradecanoylcarnitine | HMDB05066 | M+H_[+1] | - | - |
| HILIC positive | 313.2372 | 28.1 | -0.31 (0.86) | 0.4 (0.63) | -0.72 (-1.2, -0.23) | 0.0082 | 0.8637 | 13-L-Hydroperoxylinoleic acid; 8(R)-Hydroperoxylinoleic acid; 9(S)-HPODE; 1213-DiHODE; 1516-DiHODE; 910-DiHODE; ()-(E)-13-Hydroxy-10-oxo-11-octadecenoic acid | HMDB03871; HMDB04706; HMDB06940; HMDB10201; HMDB10208; HMDB10221; HMDB40900 | M+H | - | - |
| HILIC positive | 427.361 | 27.1 | 0.61 (0.75) | -0.28 (0.57) | 0.89 (0.29, 1.5) | 0.0083 | 0.8637 | Oleoylcarnitine; Vaccenyl carnitine; Elaidic carnitine; 11Z-Octadecenylcarnitine | HMDB05065; HMDB06351; HMDB06464; HMDB13338 | M+H_[+1] | - | - |
| HILIC positive | 151.0478 | 74.7 | -0.1 (0.85) | 0.7 (0.49) | -0.8 (-1.34, -0.25) | 0.0085 | 0.8637 | L-Glutamine; Ureidoisobutyric acid; D-Glutamine; Alanylglycine; Alanyl-Glycine | HMDB00641; HMDB02031; HMDB03423; HMDB06899; HMDB28687 | M+H_[+4] | - | - |
| C18 negative | 591.39 | 200.8 | -0.14 (0.24) | 0.13 (0.26) | -0.27 (-0.45, -0.08) | 0.0085 | 0.8508 | Tuberoside | HMDB39054 | M-H | - | - |
| C18 negative | 205.0174 | 22.8 | -0.4 (0.85) | 0.26 (0.68) | -0.66 (-1.12, -0.21) | 0.0086 | 0.8508 | Daucic acid | HMDB31665 | M-H_[+2] | - | - |
| HILIC positive | 219.0975 | 72 | 0.05 (0.75) | 0.63 (0.84) | -0.58 (-0.97, -0.18) | 0.0088 | 0.8637 | Glutamylalanine; 5-L-Glutamyl-L-alanine; Hydroxyprolyl-Serine; Serinyl-Hydroxyproline; N-gamma-L-Glutamyl-D-alanine | HMDB03764; HMDB06248; HMDB28872; HMDB29040; HMDB36301 | M+H | - | - |
| C18 negative | 256.0939 | 16.2 | 0.34 (0.18) | 0.18 (0.08) | 0.16 (0.05, 0.28) | 0.0096 | 0.8508 | 5-Methylcytidine; Pyro-L-glutaminyl-L-glutamine | HMDB00982; HMDB39229 | M-H | - | - |
| HILIC positive | 121.0509 | 272.4 | 0.34 (0.79) | -0.61 (0.89) | 0.95 (0.29, 1.61) | 0.0096 | 0.8637 | Purine | HMDB01366 | M+H | - | - |
| C18 negative | 886.7305 | 41.3 | -0.99 (1.33) | 0.52 (0.62) | -1.51 (-2.57, -0.45) | 0.0099 | 0.8508 | PE(22:024:0); PE(24:022:0) | HMDB09508; HMDB09732 | M-H | - | - |
| HILIC positive | 178.0862 | 30.6 | 0.24 (0.92) | -0.74 (0.88) | 0.98 (0.29, 1.67) | 0.0102 | 0.8637 | 5-Hydroxytryptophol; 12-Dehydrosalsolinol; Plantagonine; 2-Propenyl 2-aminobenzoate; 5-2H-Pyrrol-4-(3H)-ylidenemethyl-2-furanmethanol; NN-dimethylindoliumolate | HMDB01855; HMDB12490; HMDB33482; HMDB37693; HMDB40045; HMDB60063 | M+H | - | - |
| HILIC positive | 259.071 | 85.5 | -1.03 (0.15) | -0.05 (1.05) | -0.98 (-1.67, -0.29) | 0.0103 | 0.8637 | Thalidomide | HMDB15175 | M+H | - | - |
| HILIC positive | 901.7241 | 73 | -1.36 (1.14) | -0.14 (1.15) | -1.22 (-2.08, -0.36) | 0.0103 | 0.8637 | TG(16:1(9Z)20:4(5Z8Z11Z14Z)20:4(5Z8Z11Z14Z))iso3; TG(18:2(9Z12Z)18:2(9Z12Z)20:5(5Z8Z11Z14Z17Z))iso3; TG(18:3(9Z12Z15Z)16:022:6(4Z7Z10Z13Z16Z19Z))iso6; TG(18:3(9Z12Z15Z)18:1(9Z)20:5(5Z8Z11Z14Z17Z))iso6; TG(20:4(5Z8Z11Z14Z)18:2(9Z12Z)18:3(9Z12Z15Z))iso6; TG(14:020:3(5Z8Z11Z)22:6(4Z7Z10Z13Z16Z19Z)); TG(14:020:3n622:6(4Z7Z10Z13Z16Z19Z)); TG(14:020:4(5Z8Z11Z14Z)22:5(4Z7Z10Z13Z16Z)); TG(14:020:4(5Z8Z11Z14Z)22:5(7Z10Z13Z16Z19Z)); TG(14:022:4(7Z10Z13Z16Z)20:5(5Z8Z11Z14Z17Z)); TG(14:022:5(4Z7Z10Z13Z16Z)20:4(5Z8Z11Z14Z)); TG(14:022:5(4Z7Z10Z13Z16Z)20:4(8Z11Z14Z17Z)); TG(14:020:4(8Z11Z14Z17Z)22:5(4Z7Z10Z13Z16Z)); TG(14:020:4(8Z11Z14Z17Z)22:5(7Z10Z13Z16Z19Z)); TG(14:020:5(5Z8Z11Z14Z17Z)22:4(7Z10Z13Z16Z)); TG(14:022:5(7Z10Z13Z16Z19Z)20:4(5Z8Z11Z14Z)); TG(14:022:5(7Z10Z13Z16Z19Z)20:4(8Z11Z14Z17Z)); TG(14:022:6(4Z7Z10Z13Z16Z19Z)20:3(5Z8Z11Z)); TG(14:022:6(4Z7Z10Z13Z16Z19Z)20:3n6); TG(16:018:3(6Z9Z12Z)22:6(4Z7Z10Z13Z16Z19Z)); TG(16:020:4(5Z8Z11Z14Z)20:5(5Z8Z11Z14Z17Z)); TG(16:022:5(4Z7Z10Z13Z16Z)18:4(6Z9Z12Z15Z)); TG(16:018:3(9Z12Z15Z)22:6(4Z7Z10Z13Z16Z19Z)); TG(16:018:4(6Z9Z12Z15Z)22:5(4Z7Z10Z13Z16Z)); TG(16:018:4(6Z9Z12Z15Z)22:5(7Z10Z13Z16Z19Z)); TG(16:020:4(8Z11Z14Z17Z)20:5(5Z8Z11Z14Z17Z)); TG(16:020:5(5Z8Z11Z14Z17Z)20:4(5Z8Z11Z14Z)); TG(16:020:5(5Z8Z11Z14Z17Z)20:4(8Z11Z14Z17Z)); TG(16:022:5(7Z10Z13Z16Z19Z)18:4(6Z9Z12Z15Z)); | HMDB05448; HMDB10493; HMDB10500; HMDB10504; HMDB10527; HMDB42449; HMDB42629; HMDB42653; HMDB42658; HMDB42717; HMDB42740; HMDB42746; HMDB42833; HMDB42838; HMDB42862; HMDB42890; HMDB42896; HMDB42913; HMDB42919; HMDB44303; HMDB44385; HMDB44467; HMDB44499; HMDB44521; HMDB44526; HMDB44553; HMDB44574; HMDB44580; HMDB44607; HMDB44627; HMDB44634; HMDB45339; HMDB45391; HMDB48015; HMDB48020; HMDB48153; HMDB48158; HMDB48175; HMDB48219; HMDB48225; HMDB48235; HMDB48241; HMDB48313; HMDB48350; HMDB48356; HMDB48695; HMDB48779; HMDB48784; HMDB48827; HMDB48848; HMDB48891; HMDB48905; HMDB48912; HMDB48933; HMDB48938; HMDB48954; HMDB48974; HMDB48980; | M+H | - | - |

| | | | | | | | | | |
|---|---|---|---|---|---|---|---|---|---|
| | | | | | | | | TG(16:022:6(4Z7Z10Z13Z16Z19Z)18:3(6Z9Z12Z));<br>TG(16:022:6(4Z7Z10Z13Z16Z19Z)18:3(9Z12Z15Z));<br>TG(18:018:4(6Z9Z12Z15Z)20:5(5Z8Z11Z14Z17Z));<br>TG(18:020:5(5Z8Z11Z14Z17Z)18:4(6Z9Z12Z15Z));<br>TG(14:1(9Z)20:3(5Z8Z11Z)22:5(4Z7Z10Z13Z16Z));<br>TG(14:1(9Z)20:3(5Z8Z11Z)22:5(7Z10Z13Z16Z19Z));<br>TG(14:1(9Z)20:3n622:5(4Z7Z10Z13Z16Z));<br>TG(14:1(9Z)20:3n622:5(7Z10Z13Z16Z19Z));<br>TG(14:1(9Z)20:4(5Z8Z11Z14Z)22:4(7Z10Z13Z16Z));<br>TG(14:1(9Z)22:4(7Z10Z13Z16Z)20:4(5Z8Z11Z14Z));<br>TG(14:1(9Z)22:4(7Z10Z13Z16Z)20:4(8Z11Z14Z17Z));<br>TG(14:1(9Z)22:5(4Z7Z10Z13Z16Z)20:3(5Z8Z11Z));<br>TG(14:1(9Z)22:5(4Z7Z10Z13Z16Z)20:3n6);<br>TG(14:1(9Z)20:4(8Z11Z14Z17Z)22:4(7Z10Z13Z16Z));<br>TG(14:1(9Z)22:5(7Z10Z13Z16Z19Z)20:3(5Z8Z11Z));<br>TG(14:1(9Z)22:5(7Z10Z13Z16Z19Z)20:3n6);<br>TG(16:1(9Z)20:3(5Z8Z11Z)20:5(5Z8Z11Z14Z17Z));<br>TG(16:1(9Z)18:3(6Z9Z12Z)22:5(4Z7Z10Z13Z16Z));<br>TG(16:1(9Z)18:3(6Z9Z12Z)22:5(7Z10Z13Z16Z19Z));<br>TG(16:1(9Z)20:3n620:5(5Z8Z11Z14Z17Z));<br>TG(16:1(9Z)20:4(5Z8Z11Z14Z)20:4(8Z11Z14Z17Z));<br>TG(16:1(9Z)22:4(7Z10Z13Z16Z)18:4(6Z9Z12Z15Z));<br>TG(16:1(9Z)22:5(4Z7Z10Z13Z16Z)18:3(6Z9Z12Z));<br>TG(16:1(9Z)22:5(4Z7Z10Z13Z16Z)18:3(9Z12Z15Z));<br>TG(16:1(9Z)18:3(9Z12Z15Z)22:5(4Z7Z10Z13Z16Z));<br>TG(16:1(9Z)18:3(9Z12Z15Z)22:5(7Z10Z13Z16Z19Z));<br>TG(16:1(9Z)18:4(6Z9Z12Z15Z)22:4(7Z10Z13Z16Z));<br>TG(16:1(9Z)20:4(8Z11Z14Z17Z)20:4(5Z8Z11Z14Z));<br>TG(16:1(9Z)20:4(8Z11Z14Z17Z)20:4(8Z11Z14Z17Z));<br>TG(16:1(9Z)20:5(5Z8Z11Z14Z17Z)20:3(5Z8Z11Z));<br>TG(16:1(9Z)20:5(5Z8Z11Z14Z17Z)20:3n6);<br>TG(16:1(9Z)22:5(7Z10Z13Z16Z19Z)18:3(6Z9Z12Z));<br>TG(16:1(9Z)22:5(7Z10Z13Z16Z19Z)18:3(9Z12Z15Z));<br>TG(18:1(11Z)18:3(6Z9Z12Z)20:5(5Z8Z11Z14Z17Z));<br>TG(18:1(11Z)20:4(5Z8Z11Z14Z)18:4(6Z9Z12Z15Z));<br>TG(18:1(11Z)18:3(9Z12Z15Z)20:5(5Z8Z11Z14Z17Z));<br>TG(18:1(11Z)18:4(6Z9Z12Z15Z)20:4(5Z8Z11Z14Z));<br>TG(18:1(11Z)18:4(6Z9Z12Z15Z)20:4(8Z11Z14Z17Z));<br>TG(18:1(11Z)20:4(8Z11Z14Z17Z)18:4(6Z9Z12Z15Z));<br>TG(18:1(11Z)20:5(5Z8Z11Z14Z17Z)18:3(6Z9Z12Z));<br>TG(18:1(11Z)20:5(5Z8Z11Z14Z17Z)18:3(9Z12Z15Z));<br>TG(18:1(9Z)18:3(6Z9Z12Z)20:5(5Z8Z11Z14Z17Z));<br>TG(18:1(9Z)20:4(5Z8Z11Z14Z)18:4(6Z9Z12Z15Z));<br>TG(18:1(9Z)18:3(9Z12Z15Z)20:5(5Z8Z11Z14Z17Z));<br>TG(18:1(9Z)18:4(6Z9Z12Z15Z)20:4(5Z8Z11Z14Z));<br>TG(18:1(9Z)18:4(6Z9Z12Z15Z)20:4(8Z11Z14Z17Z));<br>TG(18:1(9Z)20:4(8Z11Z14Z17Z)18:4(6Z9Z12Z15Z));<br>TG(18:1(9Z)20:5(5Z8Z11Z14Z17Z)18:3(6Z9Z12Z));<br>TG(18:1(9Z)20:5(5Z8Z11Z14Z17Z)18:3(9Z12Z15Z));<br>TG(20:1(11Z)18:4(6Z9Z12Z15Z)18:4(6Z9Z12Z15Z));<br>TG(20:3(5Z8Z11Z)14:022:6(4Z7Z10Z13Z16Z19Z));<br>TG(20:3(5Z8Z11Z)14:1(9Z)22:5(4Z7Z10Z13Z16Z));<br>TG(20:3(5Z8Z11Z)14:1(9Z)22:5(7Z10Z13Z16Z19Z));<br>TG(20:3(5Z8Z11Z)16:1(9Z)20:5(5Z8Z11Z14Z17Z));<br>TG(20:3(5Z8Z11Z)18:3(6Z9Z12Z)18:3(6Z9Z12Z));<br>TG(20:3(5Z8Z11Z)18:3(6Z9Z12Z)18:3(9Z12Z15Z));<br>TG(20:3(5Z8Z11Z)18:3(9Z12Z15Z)18:3(6Z9Z12Z));<br>TG(20:3(5Z8Z11Z)18:3(9Z12Z15Z)18:3(9Z12Z15Z));<br>TG(18:3(6Z9Z12Z)16:022:6(4Z7Z10Z13Z16Z19Z));<br>TG(18:3(6Z9Z12Z)16:1(9Z)22:5(4Z7Z10Z13Z16Z));<br>TG(18:3(6Z9Z12Z)16:1(9Z)22:5(7Z10Z13Z16Z19Z));<br>TG(18:3(6Z9Z12Z)18:1(11Z)20:5(5Z8Z11Z14Z17Z));<br>TG(18:3(6Z9Z12Z)18:1(9Z)20:5(5Z8Z11Z14Z17Z));<br>TG(18:3(6Z9Z12Z)20:3(5Z8Z11Z)18:3(6Z9Z12Z));<br>TG(18:3(6Z9Z12Z)20:3(5Z8Z11Z)18:3(9Z12Z15Z));<br>TG(18:3(6Z9Z12Z)18:3(6Z9Z12Z)20:3n6);<br>TG(18:3(6Z9Z12Z)20:3n618:3(6Z9Z12Z)); | HMDB48989; HMDB48995;<br>HMDB49015; HMDB49022;<br>HMDB49425; HMDB49486;<br>HMDB49572; HMDB49586;<br>HMDB49592; HMDB49612;<br>HMDB49625; HMDB49632;<br>HMDB50037; HMDB50095;<br>HMDB50177; HMDB50190;<br>HMDB50196; HMDB50215;<br>HMDB50227; HMDB50234;<br>HMDB50769; HMDB50885;<br>HMDB51005; HMDB51010;<br>HMDB51027; HMDB51161;<br>HMDB51168; HMDB51287;<br>HMDB51294; HMDB52885;<br>HMDB52963; HMDB52968;<br>HMDB52981; HMDB52995;<br>HMDB53013; HMDB53020;<br>HMDB53071; HMDB53097;<br>HMDB53104; HMDB53169;<br>HMDB53663; HMDB53741;<br>HMDB53746; HMDB53757;<br>HMDB53850; HMDB53934;<br>HMDB54015; HMDB54020;<br>HMDB54041; HMDB54091;<br>HMDB54100; HMDB54106;<br>HMDB54116; HMDB54127;<br>HMDB54645; HMDB54707;<br>HMDB54715; HMDB54912;<br>HMDB54927; HMDB54974;<br>HMDB55208; HMDB55214;<br>HMDB55232; HMDB55274;<br>HMDB55372; HMDB55377;<br>HMDB55412; HMDB55418;<br>HMDB55423; HMDB55538;<br>HMDB55547; HMDB55576 |

| | | | | | | | | | | | | |
|---|---|---|---|---|---|---|---|---|---|---|---|---|
| | | | | | | | | TG(18:3(6Z9Z12Z)20:3n618:3(9Z12Z15Z)); TG(18:3(6Z9Z12Z)18:3(9Z12Z15Z)20:3n6); TG(20:3n614:022:6(4Z7Z10Z13Z16Z19Z)); TG(20:3n614:1(9Z)22:5(4Z7Z10Z13Z16Z)); TG(20:3n614:1(9Z)22:5(7Z10Z13Z16Z19Z)); TG(20:3n616:1(9Z)20:5(5Z8Z11Z14Z17Z)); TG(20:3n618:3(6Z9Z12Z)18:3(9Z12Z15Z)); TG(20:3n618:3(9Z12Z15Z)18:3(9Z12Z15Z)); TG(20:4(5Z8Z11Z14Z)14:022:5(4Z7Z10Z13Z16Z)); TG(20:4(5Z8Z11Z14Z)14:022:5(7Z10Z13Z16Z19Z)); TG(20:4(5Z8Z11Z14Z)16:020:5(5Z8Z11Z14Z17Z)); TG(20:4(5Z8Z11Z14Z)14:1(9Z)22:4(7Z10Z13Z16Z)); TG(20:4(5Z8Z11Z14Z)16:1(9Z)20:4(5Z8Z11Z14Z)); TG(20:4(5Z8Z11Z14Z)16:1(9Z)20:4(8Z11Z14Z17Z)); TG(20:4(5Z8Z11Z14Z)18:1(11Z)18:4(6Z9Z12Z15Z)); TG(20:4(5Z8Z11Z14Z)18:1(9Z)18:4(6Z9Z12Z15Z)); TG(22:4(7Z10Z13Z16Z)14:020:5(5Z8Z11Z14Z17Z)); TG(22:4(7Z10Z13Z16Z)14:1(9Z)20:4(8Z11Z14Z17Z)); TG(22:4(7Z10Z13Z16Z)16:1(9Z)18:4(6Z9Z12Z15Z)); TG(22:5(4Z7Z10Z13Z16Z)14:020:4(8Z11Z14Z17Z)); TG(22:5(4Z7Z10Z13Z16Z)16:018:4(6Z9Z12Z15Z)); TG(22:5(4Z7Z10Z13Z16Z)16:1(9Z)18:3(9Z12Z15Z)); TG(18:3(9Z12Z15Z)16:1(9Z)22:5(7Z10Z13Z16Z19Z)); TG(18:3(9Z12Z15Z)18:1(11Z)20:5(5Z8Z11Z14Z17Z)); TG(18:3(9Z12Z15Z)20:3(5Z8Z11Z)18:3(9Z12Z15Z)); TG(18:3(9Z12Z15Z)20:3n618:3(9Z12Z15Z)); TG(18:4(6Z9Z12Z15Z)16:022:5(7Z10Z13Z16Z19Z)); TG(18:4(6Z9Z12Z15Z)18:020:5(5Z8Z11Z14Z17Z)); TG(18:4(6Z9Z12Z15Z)18:1(11Z)20:4(8Z11Z14Z17Z)); TG(18:4(6Z9Z12Z15Z)18:1(9Z)20:4(8Z11Z14Z17Z)); TG(18:4(6Z9Z12Z15Z)20:1(11Z)18:4(6Z9Z12Z15Z)); TG(20:4(8Z11Z14Z17Z)14:022:5(7Z10Z13Z16Z19Z)); TG(20:4(8Z11Z14Z17Z)16:020:5(5Z8Z11Z14Z17Z)); TG(20:4(8Z11Z14Z17Z)16:1(9Z)20:4(8Z11Z14Z17Z)) | | | | |
| C18 negative | 475.2668 | 184.2 | -0.27 (0.35) | 0.38 (0.58) | -0.65 (-1.11, -0.19) | 0.0105 | 0.8508 | LysoPE(0:018:3(6Z9Z12Z)); LysoPE(0:018:3(9Z12Z15Z)); LysoPE(18:3(6Z9Z12Z)0:0); LysoPE(18:3(9Z12Z15Z)0:0) | HMDB11478; HMDB11479; HMDB11508; HMDB11509 | M-H_[-1] | - | - |
| C18 negative | 170.9443 | 25.1 | 0.37 (0.73) | -0.56 (1.03) | 0.93 (0.27, 1.59) | 0.0107 | 0.8508 | 2-Bromophenol; Bromobenzene-23-oxide; Bromobenzene-34-oxide | HMDB32059; HMDB60446; HMDB60448 | M-H | - | - |
| HILIC positive | 299.1042 | 91 | 0.51 (0.27) | 0.7 (0.17) | -0.19 (-0.32, -0.05) | 0.0108 | 0.8637 | 8-Deoxy-11-hydroxy-13-chlorogrosheimin | HMDB41037 | M+H | - | - |
| HILIC positive | 169.0586 | 76.6 | -0.03 (0.75) | 0.67 (0.52) | -0.7 (-1.21, -0.2) | 0.0109 | 0.8637 | L-Glutamine; Ureidoisobutyric acid; D-Glutamine; Alanylglycine; Alanyl-Glycine | HMDB00641; HMDB02031; HMDB03423; HMDB06899; HMDB28687 | M+Na | - | - |
| HILIC positive | 387.3615 | 26.8 | -1.1 (1.14) | -0.15 (0.99) | -0.95 (-1.63, -0.27) | 0.0109 | 0.8637 | Cholesterol; 5alpha-Cholestanone; Lathosterol; 5a-Cholest-8-en-3b-ol; 5beta-Cholestanone; Doristerol; 5-beta-Cholestan-3-one | HMDB00067; HMDB00871; HMDB01170; HMDB06841; HMDB11182; HMDB29815; HMDB59604 | M+H | - | - |
| HILIC positive | 387.3468 | 27.2 | 0.36 (1.25) | -0.38 (0.99) | 0.74 (0.21, 1.26) | 0.0110 | 0.8637 | DG(18:0e2:00:0); MG(0:020:00:0); MG(20:00:00:0) | HMDB11147; HMDB11542; HMDB11572 | M+H | - | - |
| C18 negative | 127.1128 | 161.7 | 0.52 (0.41) | -0.4 (0.91) | 0.92 (0.25, 1.59) | 0.0120 | 0.8508 | Octanal; 25-Diethyltetrahydrofuran; (R)-Sulcatol; 2-Octanone; Ethyl pentyl ketone; (E)-2-Octen-1-ol; (Z)-5-Octen-1-ol; (R)-1-Octen-3-ol; ()-(E)-2-Octen-4-ol; 2-Methyl-3-heptanone; trans-3-Octen-2-ol; (Z)-3-Octen-1-ol; (E)-3-Octen-1-ol; xi-7-Octen-2-ol | HMDB01140; HMDB29574; HMDB30030; HMDB31294; HMDB31295; HMDB31296; HMDB31298; HMDB31299; HMDB31300; HMDB32402; HMDB32450; HMDB35231; HMDB35768; HMDB37181 | M-H | - | - |
| C18 negative | 201.0879 | 288 | -0.45 (0.49) | 0.33 (0.89) | -0.77 (-1.34, -0.21) | 0.0125 | 0.8508 | Alanyl-Hydroxyproline; Hydroxyprolyl-Alanine; Prolyl-Serine; Serinyl-Proline; L-Coprine | HMDB28688; HMDB28856; HMDB29026; HMDB29047; HMDB34266 | M-H | - | - |
| C18 negative | 239.0928 | 294 | -0.31 (0.95) | 0.33 (0.88) | -0.64 (-1.11, -0.17) | 0.0126 | 0.8508 | 3-(345-Trimethoxyphenyl)propanoic acid; Ethyl 345-trimethoxybenzoate; Isopropyl 3-(34-dihydroxyphenyl)-2-hydroxypropanoate; 3-Carboxy-4-methyl-5-propyl-2-furanpropionic acid | HMDB30254; HMDB38627; HMDB41756; HMDB61112 | M-H | - | - |
| C18 negative | 151.0401 | 23.8 | 0.34 (0.49) | -0.08 (0.49) | 0.42 (0.11, 0.74) | 0.0130 | 0.8508 | p-Hydroxyphenylacetic acid; 3-Hydroxyphenylacetic acid; Ortho-Hydroxyphenylacetic acid; Mandelic acid; p-Anisic acid; 3-Cresotinic acid; 34-Dihydroxyphenylacetaldehyde; 4-Hydroxy-3-methylbenzoic acid; Vanillin; 24- | HMDB00020; HMDB00440; HMDB00669; HMDB00703; HMDB01101; HMDB02390; HMDB03791; HMDB04815; | M-H | Hydroxyphenylacetate | M-H |

| | | | | | | | | | | | | |
|---|---|---|---|---|---|---|---|---|---|---|---|---|
| | | | | | | | | Dihydroxyacetophenone; 23-Dihydroxyacetophenone; 26-Dihydroxyacetophenone; 34-Dihydroxyacetophenone; 35-Dihydroxyacetophenone; Phenoxyacetic acid; Methyl furfuracrylate; Methylparaben; 2-(Hydroxymethyl)benzoic acid; 2-Methoxybenzoic acid; 3-Methoxybenzoic acid; 25-Dihydroxyacetophenone; Ethyl 2-furanyl diketone; 1-(5-Methyl-2-furanyl)-12-propanedione; Methyl 2-hydroxybenzoate; 2-Propenyl 2-furancarboxylate; 1-(2-Furanyl)-13-butanedione; 4-Hydroxyphenyl acetate | HMDB12308; HMDB29657; HMDB29658; HMDB29659; HMDB29660; HMDB29661; HMDB29662; HMDB31609; HMDB32399; HMDB32572; HMDB32594; HMDB32604; HMDB32606; HMDB32629; HMDB32919; HMDB33003; HMDB34172; HMDB36907; HMDB41283; HMDB60390 | | | |
| HILIC positive | 772.5841 | 31 | -0.56 (0.72) | 0.37 (0.91) | -0.93 (-1.61, -0.24) | 0.0132 | 0.8637 | PC(15:020:2(11Z14Z)); PC(20:2(11Z14Z)15:0); PE(14:1(9Z)24:1(15Z)); PE(16:022:2(13Z16Z)); PE(16:1(9Z)22:1(13Z)); PE(18:020:2(11Z14Z)); PE(18:1(11Z)20:1(11Z)); PE(18:1(9Z)20:1(11Z)); PE(18:2(9Z12Z)20:0); PE(20:018:2(9Z12Z)); PE(20:1(11Z)18:1(11Z)); PE(20:1(11Z)18:1(9Z)); PE(20:2(11Z14Z)18:0); PE(22:1(13Z)16:1(9Z)); PE(22:2(13Z16Z)16:0); PE(24:1(15Z)14:1(9Z)); PE-NMe2(18:1(9Z)18:1(9Z)) | HMDB07946; HMDB08330; HMDB08882; HMDB08942; HMDB08974; HMDB09000; HMDB09032; HMDB09065; HMDB09097; HMDB09225; HMDB09256; HMDB09257; HMDB09288; HMDB09518; HMDB09550; HMDB09746; HMDB10564 | M+H | - | - |
| HILIC positive | 152.0512 | 74.7 | -0.08 (0.58) | 0.61 (0.48) | -0.69 (-1.2, -0.18) | 0.0132 | 0.8637 | Mevalonic acid; (R) 23-Dihydroxy-3-methylvalerate; Glycerol 1-propanoate; (R)-mevalonate; (2-Methoxyethoxy)propanoic acid | HMDB00227; HMDB12140; HMDB31639; HMDB59629; HMDB59705 | M+H_[+3] | - | - |
| C18 negative | 289.0565 | 135.7 | -0.29 (0.58) | 0.3 (0.64) | -0.59 (-1.02, -0.15) | 0.0135 | 0.8508 | Isoprothiolane; Dedimethylchlorpromazine; N-Dedimethylchlorpromazine | HMDB31779; HMDB61134; HMDB61160 | M-H | - | - |
| C18 negative | 505.354 | 296.7 | 0.46 (0.33) | 0.05 (0.48) | 0.41 (0.11, 0.72) | 0.0135 | 0.8508 | Theasapogenol A | HMDB34519 | M-H | - | - |
| HILIC positive | 131.0469 | 74.9 | -0.16 (0.94) | 0.73 (0.63) | -0.89 (-1.56, -0.22) | 0.0144 | 0.8637 | Pipecolic acid; Pyroglutamic acid; L-Pipecolic acid; Pyrrolidonecarboxylic acid; Pyrroline hydroxycarboxylic acid; N-Acryloylglycine; 1-Pyrroline-4-hydroxy-2-carboxylate; N4-Acetylaminobutanal; D-Pipecolic acid; Vigabatrin; Flucytosine; L-trans-4-Methyl-2-pyrrolidinecarboxylic acid; 2-Pyrrolidineacetic acid; 3-Acetamidobutanal; 5-Oxoprolinate; dimethadione | HMDB00070; HMDB00267; HMDB00716; HMDB00805; HMDB01369; HMDB01843; HMDB02234; HMDB04226; HMDB05960; HMDB15212; HMDB15231; HMDB29435; HMDB29444; HMDB59649; HMDB60262; HMDB61093 | M+H_[+1] | - | - |
| HILIC positive | 399.33 | 27.1 | 0.41 (0.21) | 0.15 (0.13) | 0.26 (0.06, 0.46) | 0.0148 | 0.8637 | trans-Hexadec-2-enoyl carnitine; 9-Hexadecenoylcarnitine | HMDB06317; HMDB13207 | M+H_[+1] | - | - |
| C18 negative | 130.0509 | 21.9 | -0.36 (0.79) | 0.32 (0.91) | -0.67 (-1.19, -0.16) | 0.0152 | 0.8508 | 4-Hydroxyproline; N-Acetyl-L-alanine; Propionylglycine; 5-Aminolevulinic acid; L-Glutamic gamma-semialdehyde; 3-Hydroxy-L-proline; 4-Hydroxy-L-proline; 5-Amino-2-oxopentanoic acid; 4-Hydroxy-2-pyrrolidinecarboxylic acid; Trans-3-hydroxy-L-proline; cis-4-Hydroxy-D-proline | HMDB00725; HMDB00766; HMDB00783; HMDB01149; HMDB02104; HMDB02113; HMDB06055; HMDB06272; HMDB36576; HMDB59659; HMDB60460 | M-H | - | - |
| HILIC positive | 823.6396 | 31.9 | -0.2 (0.57) | 0.26 (0.63) | -0.46 (-0.81, -0.11) | 0.0153 | 0.8637 | PC(22:4(7Z10Z13Z16Z)P-18:0); PC(P-18:022:4(7Z10Z13Z16Z)); PC(o-20:1(11Z)20:4(8Z11Z14Z17Z)) | HMDB08653; HMDB11259; HMDB13444 | M+H_[+1] | - | - |
| C18 negative | 279.0386 | 23.3 | 0.53 (0.88) | -0.27 (0.82) | 0.8 (0.19, 1.41) | 0.0154 | 0.8508 | Uridine; Pseudouridine | HMDB00296; HMDB00767 | M+Cl | - | - |
| C18 negative | 370.2961 | 235.1 | 0.66 (0.47) | 0.12 (0.67) | 0.53 (0.13, 0.94) | 0.0154 | 0.8508 | Tetradecanoylcarnitine | HMDB05066 | M-H | - | - |
| HILIC positive | 282.2216 | 294 | 0.42 (0.8) | -0.56 (1.16) | 0.99 (0.23, 1.74) | 0.0157 | 0.8637 | Alverine | HMDB15554 | M+H | - | - |
| HILIC positive | 160.1162 | 285.2 | -0.5 (0.84) | 0.55 (0.6) | -1.04 (-1.84, -0.24) | 0.0158 | 0.8637 | ()-2-Pentylthiazolidine | HMDB40057 | M+H | - | - |
| C18 negative | 245.1395 | 295.6 | -0.43 (0.89) | 0.36 (0.85) | -0.79 (-1.39, -0.18) | 0.0159 | 0.8508 | 3-Hydroxydodecanedioic acid; Dibutyl malate | HMDB00413; HMDB31696 | M-H | - | - |
| HILIC positive | 153.0407 | 44.9 | 0.43 (0.29) | 0.12 (0.31) | 0.3 (0.07, 0.54) | 0.0159 | 0.8637 | Xanthine; Oxypurinol; 68-Dihydroxypurine | HMDB00292; HMDB00786; HMDB01182 | M+H | Xanthine | M+H |
| HILIC positive | 179.1067 | 107.7 | 0.24 (0.47) | -0.44 (0.82) | 0.68 (0.16, 1.21) | 0.0163 | 0.8637 | 5-Phenylvaleric acid; Phenylmethyl 2-methylpropanoate; Methyleugenol; Phenylmethyl butanoate; 3-Phenylpropyl acetate; 4-(4-Methoxyphenyl)-2-butanone; 2-Phenylethyl propanoate; alphaalpha-Dimethylphenethyl formate; Methyl 4-phenylbutanoate; 2-Methylphenyl 2-methylpropanoate; p-Tolyl isobutyrate; 3-(2-Hydroxy-4-methylphenyl)-2-butanone; 1-Phenylethyl propanoate; Osmorhizole; Ethyl 3-phenylpropanoate; Propyl phenylacetate; Isopropyl phenylacetate; 2-Methylpropyl benzoate; 2-Ethoxy-5-(1-propenyl)phenol; Methylisoeugenol | HMDB02043; HMDB31250; HMDB31864; HMDB33376; HMDB33386; HMDB33597; HMDB35013; HMDB36235; HMDB36385; HMDB37680; HMDB37708; HMDB38179; HMDB40287; HMDB40356; HMDB40411; HMDB40425; HMDB40426; HMDB40583; HMDB40800; HMDB41553 | M+H | - | - |
| C18 negative | 469.2842 | 282.4 | 0.48 (0.63) | -0.35 (1.14) | 0.84 (0.19, 1.48) | 0.0166 | 0.8508 | Ceanothine C | HMDB29340 | M-H | - | - |
| HILIC positive | 147.0471 | 75.2 | -0.13 (0.75) | 0.69 (0.66) | -0.82 (-1.47, -0.18) | 0.0171 | 0.8637 | Dihydro-2-methoxy-2-methyl-3(2H)-thiophenone | HMDB40237 | M+H | - | - |
| C18 negative | 161.0568 | 295.3 | -0.3 (0.71) | 0.4 (0.67) | -0.69 (-1.24, -0.15) | 0.0172 | 0.8508 | Glycyl-Serine; Serinyl-Glycine; L-4-Hydroxyglutamine | HMDB28850; HMDB29039; HMDB29424 | M-H | - | - |
| HILIC positive | 131.0533 | 75.4 | -0.16 (0.99) | 0.72 (0.69) | -0.88 (-1.57, -0.19) | 0.0176 | 0.8637 | Pipecolic acid; Pyroglutamic acid; L-Pipecolic acid; Pyrrolidonecarboxylic acid; Pyrroline hydroxycarboxylic acid; | HMDB00070; HMDB00267; HMDB00716; HMDB00805; | M+H_[+1] | - | - |

| | | | | | | | | | | | | |
|---|---|---|---|---|---|---|---|---|---|---|---|---|
| | | | | | | | | N-Acryloylglycine; 1-Pyrroline-4-hydroxy-2-carboxylate; N4-Acetylaminobutanal; D-Pipecolic acid; Vigabatrin; L-trans-4-Methyl-2-pyrrolidinecarboxylic acid; 2-Pyrrolidineacetic acid; 3-Acetamidobutanal; 5-Oxoprolinate; dimethadione | HMDB01369; HMDB01843; HMDB02234; HMDB04226; HMDB05960; HMDB15212; HMDB29435; HMDB29444; HMDB59649; HMDB60262; HMDB61093 | | | |
| HILIC positive | 294.1487 | 75.2 | 0.05 (0.47) | 0.5 (0.25) | -0.45 (-0.8, -0.1) | 0.0178 | 0.8637 | Koenimbine; Dehydronuciferine | HMDB30210; HMDB33737 | M+H | - | - |
| HILIC positive | 773.5876 | 31.3 | -0.65 (0.85) | 0.36 (0.85) | -1.01 (-1.81, -0.22) | 0.0178 | 0.8637 | PC(15:020:2(11Z14Z)); PC(20:2(11Z14Z)15:0); PE(14:1(9Z)24:1(15Z)); PE(16:022:2(13Z16Z)); PE(16:1(9Z)22:1(13Z)); PE(18:020:2(11Z14Z)); PE(18:1(11Z)20:1(11Z)); PE(18:1(9Z)20:1(11Z)); PE(18:2(9Z12Z)20:0); PE(20:018:2(9Z12Z)); PE(20:1(11Z)18:1(11Z)); PE(20:1(11Z)18:1(9Z)); PE(20:2(11Z14Z)18:0); PE(22:1(13Z)16:1(9Z)); PE(22:2(13Z16Z)16:0); PE(24:1(15Z)14:1(9Z)); PE-NMe2(18:1(9Z)18:1(9Z)) | HMDB07946; HMDB08330; HMDB08882; HMDB08942; HMDB08974; HMDB09000; HMDB09032; HMDB09065; HMDB09097; HMDB09225; HMDB09256; HMDB09257; HMDB09288; HMDB09518; HMDB09550; HMDB09746; HMDB10564 | M+H_[+1] | - | - |
| HILIC positive | 415.3205 | 28.3 | 0.46 (0.53) | -0.74 (1.5) | 1.2 (0.25, 2.14) | 0.0179 | 0.8637 | Smilagenone; Setariol | HMDB36941; HMDB41583 | M+H | - | - |
| HILIC positive | 160.0369 | 62.1 | -1.34 (1.34) | -0.49 (1.32) | -0.84 (-1.51, -0.18) | 0.0180 | 0.8637 | Trigonelline; 2-Aminobenzoic acid; p-Aminobenzoic acid; 3-Pyridylacetic acid; m-Aminobenzoic acid; Salicylamide; Methyl nicotinate; 2-Pyridylacetic acid | HMDB00875; HMDB01123; HMDB01392; HMDB01538; HMDB01891; HMDB15687; HMDB29806; HMDB60722 | M+Na | - | - |
| C18 negative | 363.2161 | 178.9 | 0.78 (0.21) | 0.61 (0.12) | 0.17 (0.04, 0.31) | 0.0182 | 0.8508 | 13-Hydroxy-9-methoxy-10-oxo-11-octadecenoic acid; Tetrahydrocortisone; Dihydrocortisol; 3a11b21-Trihydroxy-20-oxo-5b-pregnan-18-al; 11b21-Dihydroxy-320-oxo-5b-pregnan-18-al; 11b17a21-Trihydroxypreg-nenolone; 18-Hydroxy-11-dehydrotetrahydrocorticosterone; Vanillin 3-(L-menthoxy)propane-12-diol acetal | HMDB40901; HMDB00903; HMDB03259; HMDB06753; HMDB06754; HMDB06760; HMDB13157; HMDB32553 | M+Na-2H; M-H | - | - |
| HILIC positive | 124.0757 | 34.5 | -0.07 (0.44) | 0.43 (0.29) | -0.5 (-0.89, -0.1) | 0.0183 | 0.8637 | p-Anisidine; 2-Propionylpyrrole; 4-Hydroxybenzylamine; 1-Ethyl-1H-pyrrole-2-carboxaldehyde; 2-Acetyl-1-methylpyrrole | HMDB29300; HMDB32492; HMDB32578; HMDB36058; HMDB40360 | M+H | - | - |
| HILIC positive | 151.0754 | 23 | -0.59 (0.25) | 0.4 (1.2) | -0.99 (-1.78, -0.21) | 0.0183 | 0.8637 | Hydrocinnamic acid; 4-Ethylbenzoic acid; 3-Methylphenylacetic acid; 34-Dimethylbenzoic acid; 4-Coumaryl alcohol; 2-Phenylpropionate; 2-Methoxy-4-vinylphenol; Benzyl acetate; o-Tolyl acetate; p-Tolyl acetate; 2-Methoxyacetophenone; 4-Methoxyacetophenone; 2-Hydroxy-5-methylacetophenone; Methyl phenylacetate; Ethyl benzoate; 4-Ethoxybenzaldehyde; 1-Phenylethyl formate; 2-Ethyl-4-(2-furanyl)-2-propenal; 2-Phenylethyl formate; 2-(2-Furanyl)-3-methyl-2-butenal; 4-Hydroxy-3-methylacetophenone | HMDB00764; HMDB02097; HMDB02222; HMDB02237; HMDB03654; HMDB11743; HMDB13744; HMDB31310; HMDB32073; HMDB32075; HMDB32569; HMDB32570; HMDB32592; HMDB32617; HMDB33967; HMDB33970; HMDB37715; HMDB38355; HMDB40428; HMDB40925; HMDB59824 | M+H | - | - |
| C18 negative | 500.9256 | 156.2 | 0.18 (1) | -0.77 (1.19) | 0.95 (0.19, 1.71) | 0.0188 | 0.8508 | Perfluorooctanesulfonic acid | HMDB59586 | M-H_[+2] | - | - |
| HILIC positive | 315.1272 | 74.7 | 0.02 (0.61) | 0.55 (0.46) | -0.53 (-0.96, -0.1) | 0.0196 | 0.8637 | L-Glutamine; Ureidoisobutyric acid; D-Glutamine; Alanylglycine; Alanyl-Glycine | HMDB00641; HMDB02031; HMDB03423; HMDB06899; HMDB28687 | 2M+Na | - | - |
| C18 negative | 213.0493 | 25.6 | -0.31 (0.75) | 0.33 (0.8) | -0.64 (-1.16, -0.12) | 0.0200 | 0.8508 | 1-Methyl-3-(2-thiazolyl)-1H-indole | HMDB34965 | M-H | - | - |
| C18 negative | 133.051 | 21.3 | -1.11 (2.22) | 0.36 (0.66) | -1.47 (-2.65, -0.28) | 0.0201 | 0.8508 | 23-Dihydroxyvaleric acid; 1-Deoxy-D-xylulose; Deoxyribose; (R)-23-Dihydroxy-isovalerate; (R)-glycerol 1-acetate; 23-Dihydroxy-2-methylbutanoic acid | HMDB00421; HMDB01292; HMDB03224; HMDB12141; HMDB12485; HMDB29576 | M-H | - | - |
| HILIC positive | 239.1278 | 203.6 | 0.35 (0.36) | -0.03 (0.65) | 0.38 (0.07, 0.68) | 0.0201 | 0.8637 | 2244-Tetramethyl-6-(1-oxopropyl)-135-cyclohexanetrione; 1234-Tetramethoxy-5-(2-propenyl)benzene | HMDB33191; HMDB34046 | M+H | - | - |
| HILIC positive | 784.6559 | 53.4 | -0.26 (1.32) | -1.43 (1.45) | 1.17 (0.22, 2.12) | 0.0202 | 0.8637 | CerP(d18:126:1(17Z)) | HMDB10707 | M+H | - | - |
| C18 negative | 225.1652 | 203.5 | 0.63 (0.75) | -0.14 (1.04) | 0.77 (0.15, 1.4) | 0.0202 | 0.8508 | (8E15E)-1815-Heptadecatriene-1113-diyne | HMDB31041 | M-H | - | - |
| HILIC positive | 136.1049 | 30.5 | 0.4 (0.36) | -0.85 (1.58) | 1.25 (0.24, 2.26) | 0.0203 | 0.8637 | Polypropylene glycol (m w 1200-3000) | HMDB32478 | M+H_[+1] | - | - |
| HILIC positive | 341.305 | 27.3 | 0.39 (1.31) | -0.36 (1.05) | 0.75 (0.14, 1.36) | 0.0207 | 0.8637 | MG(18:00:00:0); MG(0:018:00:0); Glycerol 1-octadecanoate; Polyoxyethylene (600) monoricinoleate | HMDB11131; HMDB11535; HMDB31075; HMDB32476 | M+H-H2O; M+H | - | - |
| C18 negative | 201.088 | 218.5 | -0.64 (1.17) | 0.26 (0.84) | -0.9 (-1.64, -0.17) | 0.0209 | 0.8508 | Alanyl-Hydroxyproline; Hydroxyprolyl-Alanine; Prolyl-Serine; Serinyl-Proline; L-Coprine | HMDB28688; HMDB28856; HMDB29026; HMDB29047; HMDB34266 | M-H | - | - |
| HILIC positive | 290.1596 | 113.8 | 0.42 (0.47) | -0.04 (0.54) | 0.47 (0.08, 0.85) | 0.0214 | 0.8637 | 3-Methylglutarylcarnitine | HMDB00552 | M+H | - | - |
| C18 negative | 161.0275 | 295.7 | 0.04 (0.41) | 0.39 (0.29) | -0.35 (-0.63, -0.06) | 0.0218 | 0.8508 | 12-Dihydroxy-3-keto-5-methylthiopentene; 1-(Methylsulfanyl)-1-oxopropan-2-yl acetate | HMDB12134; HMDB36204 | M-H | - | - |
| HILIC positive | 154.0479 | 71.7 | -0.64 (1.42) | 0.41 (0.8) | -1.06 (-1.93, -0.19) | 0.0219 | 0.8637 | 4-Hydroxyproline; N-Acetyl-L-alanine; Propionylglycine; 5-Aminolevulinic acid; L-Glutamic gamma-semialdehyde; 3-Hydroxy-L-proline; 4-Hydroxy-L-proline; 5-Amino-2- | HMDB00725; HMDB00766; HMDB00783; HMDB01149; HMDB02104; HMDB02113; HMDB06055; HMDB06272; | M+Na | - | - |

| | | | | | | | | | | | | |
|---|---|---|---|---|---|---|---|---|---|---|---|---|
| | | | | | | | | oxopentanoic acid; 4-Hydroxy-2-pyrrolidinecarboxylic acid; Trans-3-hydroxy-L-proline; cis-4-Hydroxy-D-proline | HMDB36576; HMDB59659; HMDB60460 | | | |
| C18 negative | 372.239 | 140.2 | 0.42 (0.57) | -0.15 (0.33) | 0.57 (0.1, 1.04) | 0.0219 | 0.8508 | Dodecanedioylcarnitine | HMDB13327 | M-H | - | - |
| HILIC positive | 153.0659 | 260.7 | 0.44 (0.75) | -0.29 (0.78) | 0.74 (0.13, 1.35) | 0.0223 | 0.8637 | N1-Methyl-2-pyridone-5-carboxamide; N1-Methyl-4-pyridone-3-carboxamide | HMDB04193; HMDB04194 | M+H | - | - |
| C18 negative | 253.0831 | 59.5 | 0.55 (0.02) | 0.58 (0.04) | -0.03 (-0.05, 0) | 0.0225 | 0.8508 | L-Agaridoxin; p-Hydroxyfelbamate; 2-Hydroxyfelbamate | HMDB29445; HMDB60669; HMDB60670 | M-H | - | - |
| C18 negative | 167.0714 | 295.5 | -0.43 (1.02) | 0.38 (0.58) | -0.82 (-1.5, -0.14) | 0.0234 | 0.8508 | 26-Dimethoxy-4-methylphenol; 4-Ipomeanol; 1-Ipomeanol; Epoxyoxophorone; Ethyl 2-furanpropionate; 2-Furanylmethyl butanoate; (4-Hydroxy-3-methoxyphenyl)ethanol; 135-Trimethoxybenzene | HMDB29680; HMDB30472; HMDB30473; HMDB32252; HMDB32921; HMDB37729; HMDB38925; HMDB59963 | M-H | - | - |
| HILIC positive | 86.06 | 70.8 | -0.1 (0.72) | 0.55 (1.03) | -0.66 (-1.2, -0.11) | 0.0234 | 0.8637 | 2-Pyrrolidinone; Acetone cyanohydrin | HMDB02039; HMDB60427 | M+H | - | - |
| HILIC positive | 276.1554 | 147.8 | 0.1 (0.44) | 0.39 (0.31) | -0.3 (-0.54, -0.05) | 0.0235 | 0.8637 | Epsilon-(gamma-Glutamyl)-lysine; L-a-glutamyl-L-Lysine | HMDB03869; HMDB04207 | M+H | - | - |
| C18 negative | 159.0662 | 290.1 | 0.59 (0.73) | -0.59 (1.16) | 1.18 (0.19, 2.16) | 0.0235 | 0.8508 | 3-Methyladipic acid; Pimelic acid; 33-Dimethylglutaric acid; 2-Methyladipic acid; Diethyl malonate; Propyleneglycol diacetate; Mono-methyl-adipate; 2-Ethylglutaric acid; Ethyl methylsuccinate | HMDB00555; HMDB00857; HMDB02441; HMDB29167; HMDB29573; HMDB41605; HMDB59722; HMDB59738; HMDB59893 | M-H | - | - |
| HILIC positive | 219.138 | 234.3 | -0.46 (1.08) | 0.44 (0.72) | -0.9 (-1.65, -0.15) | 0.0235 | 0.8637 | (R)-Pterosin B; Dimethylbenzyl carbinyl crotonate; Isoamyl cinnamate; 2-(3-Hydroxy-4-methylphenyl)-5-methyl-4-hexen-3-one; cis-3-Hexenyl phenylacetate; Cinnamyl isovalerate | HMDB30759; HMDB32227; HMDB37704; HMDB37738; HMDB38281; HMDB41318 | M+H | - | - |
| HILIC positive | 493.1357 | 291.9 | 0.26 (0.93) | 0.92 (0.22) | -0.66 (-1.22, -0.11) | 0.0240 | 0.8637 | Tricin 7-glucoside; Licoagroside A; Limocitrin 3-rhamnoside; 38-Dimethoxyapigenin 7-glucoside; 345-Trihydroxy-37-dimethoxyflavone 5-glucoside; 37-Dimethoxy-458-trihydroxyflavone 8-glucoside | HMDB30553; HMDB35448; HMDB37578; HMDB38825; HMDB39336; HMDB40479 | M+H | - | - |
| C18 negative | 378.241 | 170.3 | 0.4 (0.67) | -0.34 (0.91) | 0.74 (0.12, 1.36) | 0.0241 | 0.8508 | Sphingosine 1-phosphate; N-palmitoyl-phosphoethanolamine | HMDB00277; HMDB59630 | M-H | - | - |
| HILIC positive | 637.4801 | 25.2 | 0.04 (0.15) | 0.39 (0.42) | -0.34 (-0.63, -0.05) | 0.0242 | 0.8637 | DG(14:022:5(4Z7Z10Z13Z16Z)0:0); DG(14:022:5(7Z10Z13Z16Z19Z)0:0); DG(14:1(9Z)22:4(7Z10Z13Z16Z)0:0); DG(16:020:5(5Z8Z11Z14Z17Z)0:0); DG(16:1(9Z)20:4(5Z8Z11Z14Z)0:0); DG(16:1(9Z)20:4(8Z11Z14Z17Z)0:0); DG(18:1(11Z)18:4(6Z9Z12Z15Z)0:0); DG(18:1(9Z)18:4(6Z9Z12Z15Z)0:0); DG(18:2(9Z12Z)18:3(6Z9Z12Z)0:0); DG(18:2(9Z12Z)18:3(9Z12Z15Z)0:0); DG(18:3(6Z9Z12Z)18:2(9Z12Z)0:0); DG(18:3(9Z12Z15Z)18:2(9Z12Z)0:0); DG(18:4(6Z9Z12Z15Z)18:1(11Z)0:0); DG(18:4(6Z9Z12Z15Z)18:1(9Z)0:0); DG(20:4(5Z8Z11Z14Z)16:1(9Z)0:0); DG(20:4(8Z11Z14Z17Z)16:1(9Z)0:0); DG(20:5(5Z8Z11Z14Z17Z)16:00:0); DG(22:4(7Z10Z13Z16Z)14:1(9Z)0:0); DG(22:5(4Z7Z10Z13Z16Z)14:00:0); DG(22:5(7Z10Z13Z16Z19Z)14:00:0); DG(14:00:022:5n6); DG(14:00:022:5n3); DG(16:00:020:5n3); DG(14:1n50:022:4n6); DG(16:1n70:020:4n6); DG(16:1n70:020:4n3); DG(18:1n70:018:4n3); DG(18:1n90:018:4n3); DG(18:3(6Z9Z12Z)20:5(5Z8Z11Z14Z17Z)0:0); DG(18:3(9Z12Z15Z)20:5(5Z8Z11Z14Z17Z)0:0); DG(18:4(6Z9Z12Z15Z)20:4(5Z8Z11Z14Z)0:0); DG(18:4(6Z9Z12Z15Z)20:4(8Z11Z14Z17Z)0:0); DG(20:4(5Z8Z11Z14Z)18:4(6Z9Z12Z15Z)0:0); DG(20:4(8Z11Z14Z17Z)18:4(6Z9Z12Z15Z)0:0); DG(20:5(5Z8Z11Z14Z17Z)18:3(6Z9Z12Z)0:0); DG(20:5(5Z8Z11Z14Z17Z)18:3(9Z12Z15Z)0:0); DG(18:3n60:020:5n3); DG(20:4n60:018:4n3); DG(18:3n30:020:5n3); DG(18:4n30:020:4n3) | HMDB07032; HMDB07033; HMDB07060; HMDB07114; HMDB07141; HMDB07142; HMDB07193; HMDB07222; HMDB07249; HMDB07250; HMDB07277; HMDB07306; HMDB07333; HMDB07334; HMDB07505; HMDB07534; HMDB07562; HMDB07676; HMDB07704; HMDB07733; HMDB55974; HMDB55979; HMDB56033; HMDB56148; HMDB56167; HMDB56173; HMDB56192; HMDB56211; HMDB07288; HMDB07317; HMDB07344; HMDB07345; HMDB07512; HMDB07541; HMDB07568; HMDB07569; HMDB56306; HMDB56337; HMDB56369; HMDB56373 | M+Na; M+H | - | - |
| HILIC positive | 125.0961 | 295.6 | 0.2 (0.17) | -0.03 (0.2) | 0.23 (0.04, 0.42) | 0.0251 | 0.8637 | 23-Dimethyl-2-cyclohexen-1-one; 6-Methyl-35-heptadien-2-one; (EE)-24-Octadienal; trans trans-35-Octadien-2-one; 15-Octadien-3-one; 2-Isopropyl-5-methylfuran; (EE)-26-Octadienal; 355-Trimethyl-2-cyclopenten-1-one; 2-Butylfuran | HMDB31414; HMDB31582; HMDB31686; HMDB32446; HMDB32447; HMDB33238; HMDB39844; HMDB40246; HMDB40272 | M+H | - | - |

| | | | | | | | | | | | | |
|---|---|---|---|---|---|---|---|---|---|---|---|---|
| HILIC positive | 147.091 | 75.2 | -0.4 (1.38) | 0.77 (0.49) | -1.17 (-2.18, -0.17) | 0.0259 | 0.8637 | Dimethylbenzimidazole; 1H-Indole-3-methanamine; Myosmine; 14-Dimethylpyrrolo12-apyrazine; 34-Dimethylpyrrolo12-apyrazine; 13-Dimethylpyrrolo12-apyrazine | HMDB03701; HMDB29740; HMDB30386; HMDB30906; HMDB30907; HMDB30908 | M+H | - | - |
| HILIC positive | 160.0605 | 297.7 | -0.48 (1.12) | 0.27 (0.95) | -0.75 (-1.39, -0.11) | 0.0259 | 0.8637 | L-2-Amino-4-methylenepentanedioic acid; L-trans-alpha-Amino-2-carboxycyclopropaneacetic acid | HMDB29433; HMDB34442 | M+H | - | - |
| HILIC positive | 285.0407 | 296.6 | 0.43 (0.61) | 0.91 (0.16) | -0.48 (-0.89, -0.07) | 0.0259 | 0.8637 | Rhein; Lucernol; Repensol; Aureol | HMDB32876; HMDB34042; HMDB34138; HMDB37921 | M+H | - | - |
| HILIC positive | 695.1919 | 74.4 | 0.01 (0.53) | 0.4 (0.26) | -0.39 (-0.72, -0.06) | 0.0261 | 0.8637 | 5-Hydroxy-734-trimethoxy-8-methylisoflavone 5-neohesperidoside; Egonol gentiobioside | HMDB30627; HMDB34281 | M+2Na-H | - | - |
| C18 negative | 150.0561 | 146.7 | 0.27 (0.54) | -0.36 (0.57) | 0.63 (0.09, 1.17) | 0.0264 | 0.8508 | Acetaminophen; 2-Phenylglycine; Dopamine quinone; Leukoaminochrome; Methyl 2-aminobenzoate; 2-(Methylamino)benzoic acid; 3alpha477alpha-Tetrahydro-1H-isoindole-13(2H)-dione; Ethyl nicotinate; 2-Amino-3-methylbenzoate | HMDB01859; HMDB02210; HMDB12219; HMDB12992; HMDB29703; HMDB32609; HMDB33903; HMDB59846; HMDB60680 | M-H | - | - |
| C18 negative | 859.5313 | 220.8 | -0.55 (1.07) | 0.53 (0.8) | -1.08 (-2.01, -0.15) | 0.0265 | 0.8508 | PI(16:020:3(5Z8Z11Z)); PI(16:020:3(8Z11Z14Z)); PI(18:018:3(6Z9Z12Z)); PI(18:018:3(9Z12Z15Z)); PI(18:1(11Z)18:2(9Z12Z)); PI(18:1(9Z)18:2(9Z12Z)); PI(18:2(9Z12Z)18:1(11Z)); PI(18:2(9Z12Z)18:1(9Z)); PI(18:3(6Z9Z12Z)18:0); PI(18:3(9Z12Z15Z)18:0); PI(20:3(5Z8Z11Z)16:0); PI(20:3(8Z11Z14Z)16:0) | HMDB09787; HMDB09788; HMDB09810; HMDB09811; HMDB09826; HMDB09838; HMDB09848; HMDB09849; HMDB09858; HMDB09862; HMDB09879; HMDB09886 | M-H | - | - |
| HILIC positive | 722.5114 | 28.1 | -0.37 (1.22) | 0.58 (0.18) | -0.96 (-1.78, -0.13) | 0.0270 | 0.8637 | PE(18:4(6Z9Z12Z15Z)P-18:1(11Z)); PE(18:4(6Z9Z12Z15Z)P-18:1(9Z)); PE(20:5(5Z8Z11Z14Z17Z)P-16:0); PE(P-16:020:5(5Z8Z11Z14Z17Z)); PE(P-18:1(11Z)18:4(6Z9Z12Z15Z)); PE(P-18:1(9Z)18:4(6Z9Z12Z15Z)) | HMDB09215; HMDB09216; HMDB09477; HMDB11354; HMDB11412; HMDB11445 | M+H | - | - |
| HILIC positive | 398.3264 | 27.4 | 0.55 (0.67) | -0.09 (0.28) | 0.65 (0.09, 1.2) | 0.0271 | 0.8637 | trans-Hexadec-2-enoyl carnitine; 9-Hexadecenoylcarnitine | HMDB06317; HMDB13207 | M+H | - | - |
| HILIC positive | 139.039 | 109.5 | -0.32 (0.85) | 0.22 (0.82) | -0.54 (-1.01, -0.07) | 0.0272 | 0.8637 | 4-Hydroxybenzoic acid; Salicylic acid; 3-Hydroxybenzoic acid; Gentisate aldehyde; 2-Methoxy-14-benzoquinone; alpha-Furyl methyl diketone; Sesamol; 34-Dihydroxybenzaldehyde | HMDB00500; HMDB01895; HMDB02466; HMDB04062; HMDB32576; HMDB32920; HMDB33812; HMDB59965 | M+H | - | - |
| HILIC positive | 149.0769 | 75.2 | -0.12 (0.59) | 0.5 (0.51) | -0.61 (-1.14, -0.08) | 0.0275 | 0.8637 | L-Glutamine; Ureidoisobutyric acid; D-Glutamine; Alanylglycine; Alanyl-Glycine | HMDB00641; HMDB02031; HMDB03423; HMDB06899; HMDB28687 | M+H_[+2] | - | - |
| C18 negative | 203.0205 | 22.8 | -0.55 (1.11) | 0.36 (0.87) | -0.91 (-1.7, -0.12) | 0.0275 | 0.8508 | Daucic acid | HMDB31665 | M-H | - | - |
| C18 negative | 155.0349 | 293.2 | -0.35 (0.74) | 0.25 (0.84) | -0.6 (-1.12, -0.08) | 0.0286 | 0.8508 | Ascladiol; 23-Methyleneglutaric acid | HMDB29610; HMDB59731 | M-H | - | - |
| C18 negative | 817.3859 | 22.5 | 0.77 (0.53) | 0.22 (0.72) | 0.55 (0.07, 1.03) | 0.0286 | 0.8508 | (3x5x10x)-910-Didehydroisohumbertiol O-rhamnosyl-(1-4)-rhamnosyl-(1-2)-rhamnosyl-(1-6)-glucoside | HMDB40687 | M-H | - | - |
| HILIC positive | 792.5895 | 29.6 | -0.52 (1.03) | 0.23 (0.94) | -0.75 (-1.4, -0.09) | 0.0287 | 0.8637 | PC(20:4(5Z8Z11Z14Z)P-18:1(11Z)); PC(20:4(5Z8Z11Z14Z)P-18:1(9Z)); PC(20:4(8Z11Z14Z17Z)P-18:1(11Z)); PC(20:4(8Z11Z14Z17Z)P-18:1(9Z)); PC(20:5(5Z8Z11Z14Z17Z)P-18:0); PC(22:5(4Z7Z10Z13Z16Z)P-16:0); PC(22:5(7Z10Z13Z16Z19Z)P-16:0); PC(P-16:022:5(4Z7Z10Z13Z16Z)); PC(P-16:022:5(7Z10Z13Z16Z19Z)); PC(P-18:020:5(5Z8Z11Z14Z17Z)); PC(P-18:1(11Z)20:4(5Z8Z11Z14Z)); PC(dm18:1(11Z)20:4(8Z11Z14Z17Z)); PC(P-18:1(9Z)20:4(5Z8Z11Z14Z)); PC(P-18:1(9Z)20:4(8Z11Z14Z17Z)); PC(o-16:022:6(4Z7Z10Z13Z16Z19Z)); 2-O-(4710131619-Docosahexaenoyl)-1-O-hexadecylglycero-3-phosphocholine | HMDB08457; HMDB08458; HMDB08490; HMDB08491; HMDB08522; HMDB08685; HMDB08718; HMDB11227; HMDB11228; HMDB11255; HMDB11286; HMDB11287; HMDB11319; HMDB11320; HMDB13409; HMDB39529 | M+H | - | - |
| C18 negative | 111.02 | 220.5 | 0.36 (0.25) | 0.1 (0.23) | 0.27 (0.03, 0.5) | 0.0288 | 0.8508 | Uracil; 4-Carboxypyrazole | HMDB00300; HMDB60760 | M-H | - | - |
| HILIC positive | 127.0727 | 292.8 | 0.49 (0.22) | 0.34 (0.34) | 0.15 (0.02, 0.28) | 0.0289 | 0.8637 | Melamine | HMDB41922 | M+H | - | - |
| HILIC positive | 817.7663 | 143.1 | 0.45 (0.69) | 0.94 (0.18) | -0.48 (-0.9, -0.06) | 0.0291 | 0.8637 | TG(14:1(9Z)18:1(11Z)o-18:0); TG(14:1(9Z)18:1(9Z)o-18:0); TG(14:1(9Z)o-18:018:1(11Z)); TG(14:1(9Z)o-18:018:1(9Z)); TG(16:1(9Z)16:1(9Z)o-18:0); TG(16:1(9Z)o-18:016:1(9Z)); TG(18:1(11Z)14:1(9Z)o-18:0); TG(18:1(9Z)14:1(9Z)o-18:0) | HMDB47953; HMDB47976; HMDB48393; HMDB48394; HMDB48610; HMDB49051; HMDB49239; HMDB49860 | M+H | - | - |
| HILIC positive | 112.0393 | 75.2 | -0.12 (0.97) | 0.59 (0.59) | -0.71 (-1.34, -0.09) | 0.0295 | 0.8637 | Pyrrole-2-carboxylic acid; 2-Acetyloxazole | HMDB04230; HMDB38175 | M+H | - | - |
| HILIC positive | 298.1006 | 95.6 | -0.03 (0.8) | 0.38 (0.62) | -0.4 (-0.76, -0.05) | 0.0298 | 0.8637 | L-Menthone 12-glyceryl ketal | HMDB32367 | M+H | - | - |
| C18 negative | 393.1565 | 161.8 | -0.08 (0.81) | 0.49 (0.73) | -0.57 (-1.07, -0.07) | 0.0304 | 0.8508 | Gibberellin A39; Gibberellin A43 | HMDB35045; HMDB35046 | M-H | - | - |
| C18 negative | 166.9938 | 31.2 | -0.69 (0.85) | 0.01 (0.86) | -0.69 (-1.31, -0.08) | 0.0305 | 0.8508 | Desflurane; Hexafluoroisopropanol | HMDB15320; HMDB60648 | M-H | - | - |
| HILIC positive | 88.0393 | 111.4 | -0.21 (1.13) | 0.42 (0.6) | -0.63 (-1.2, -0.07) | 0.0306 | 0.8637 | 2-Aminoacrylic acid; 3-Isoxazolidinone | HMDB03609; HMDB39426 | M+H | - | - |
| HILIC positive | 197.1285 | 32.6 | -0.53 (0.33) | 0.16 (0.95) | -0.68 (-1.29, -0.08) | 0.0306 | 0.8637 | 2-Isopropyl-35-dimethoxy-6-methylpyrazine; L-alpha-Amino-1H-pyrrole-1-hexanoic acid | HMDB29741; HMDB40551 | M+H | - | - |

| | | | | | | | | | | | | |
|---|---|---|---|---|---|---|---|---|---|---|---|---|
| HILIC positive | 551.503 | 27.4 | 0.23 (0.75) | -0.33 (0.93) | 0.57 (0.06, 1.07) | 0.0312 | 0.8637 | Artemoin A; Artemoin B; Artemoin C; Artemoin D | HMDB33604; HMDB33605; HMDB33606; HMDB33607 | M+H | - | - |
| HILIC positive | 195.1377 | 29.4 | -0.33 (0.6) | 0.31 (0.65) | -0.65 (-1.22, -0.07) | 0.0314 | 0.8637 | 4-Hydroxypropofol; (SZ)-Lyratol acetate; Hexylresorcinol; Neocnidilide; ()-Myrtenyl acetate; Ethyl (2E4E7Z)-Decatrienoate; Ethyl (2E4Z7Z)-Decatrienoate; (2S4R)-p-Mentha-1(7)5-dien-2-ol acetate; (S)-p-Mentha-18-dien-10-yl acetate; Perillyl acetate; (R)-2-Acetoxy-p-mentha-18-diene; Carvyl acetate; Menthadienyl acetate; 8-Ocimenyl acetate; 1-(2-Furanyl)-1-octanone; (S)-Neolyratyl acetate; (22-Diethoxyethyl)benzene | HMDB14018; HMDB31840; HMDB32567; HMDB34450; HMDB35126; HMDB35375; HMDB35376; HMDB35801; HMDB37018; HMDB37231; HMDB37389; HMDB38046; HMDB38292; HMDB39685; HMDB40439; HMDB41497; HMDB41610 | M+H | - | - |
| C18 negative | 421.155 | 180.9 | -0.15 (0.76) | 0.47 (0.53) | -0.62 (-1.17, -0.07) | 0.0314 | 0.8508 | Losartan | HMDB14816 | M-H | - | - |
| HILIC positive | 180.0997 | 57.6 | 0.55 (0.1) | 0.49 (0.02) | 0.06 (0.01, 0.12) | 0.0318 | 0.8637 | Tranexamic Acid; (S)-Homostachydrine | HMDB14447; HMDB33433 | M+Na | - | - |
| HILIC positive | 130.0408 | 75 | -0.15 (0.98) | 0.65 (0.75) | -0.8 (-1.52, -0.08) | 0.0322 | 0.8637 | Flucytosine | HMDB15231 | M+H | - | - |
| HILIC positive | 101.0961 | 297.8 | 0.23 (0.32) | -0.11 (0.35) | 0.34 (0.03, 0.64) | 0.0323 | 0.8637 | 3-Hexanone; 4-Methylpentanal; Methyl isobutyl ketone; 2-Oxohexane; Ethyl isopropyl ketone; Hexanal; (E)-3-Hexen-1-ol; 2-Hexen-1-ol; 2-Ethylbutanal; (E)-4-Hexen-1-ol; 1-Hexen-3-ol; 2-Methylpentanal | HMDB00753; HMDB01318; HMDB02939; HMDB05842; HMDB05846; HMDB05994; HMDB30003; HMDB30952; HMDB31220; HMDB31502; HMDB31503; HMDB31578 | M+H | - | - |
| C18 negative | 138.9802 | 122.6 | 0.45 (0.3) | -0.19 (0.78) | 0.65 (0.06, 1.23) | 0.0328 | 0.8508 | Acetylphosphate; Phosphonoacetate | HMDB01494; HMDB04110 | M-H | - | - |
| HILIC positive | 220.0816 | 203.7 | 0.3 (0.7) | -0.35 (1.01) | 0.65 (0.06, 1.23) | 0.0330 | 0.8637 | 1-Isothiocyanato-7-(methylsulfinyl)heptane | HMDB38441 | M+H | - | - |
| HILIC positive | 342.3077 | 27.3 | 0.34 (1.25) | -0.28 (1) | 0.62 (0.06, 1.18) | 0.0330 | 0.8637 | Polyoxyethylene (600) monoricinoleate | HMDB32476 | M+H_[+1] | - | - |
| C18 negative | 190.072 | 294.7 | -0.24 (0.82) | 0.46 (0.76) | -0.69 (-1.32, -0.07) | 0.0331 | 0.8508 | Calystegine C1 | HMDB31346 | M-H | - | - |
| HILIC positive | 265.1798 | 295.5 | -0.26 (1.02) | 0.33 (0.67) | -0.58 (-1.11, -0.05) | 0.0337 | 0.8637 | 12-Oxo-23-dinor-1015-phytodienoic acid | HMDB32090 | M+H | - | - |
| HILIC positive | 213.1597 | 30.4 | -0.11 (0.23) | 0.53 (0.84) | -0.64 (-1.23, -0.06) | 0.0337 | 0.8637 | 14-Bipiperidine-1-carboxylic acid | HMDB60336 | M+H | - | - |
| HILIC positive | 793.5929 | 29.6 | -0.47 (1.03) | 0.2 (0.87) | -0.67 (-1.28, -0.06) | 0.0339 | 0.8637 | PC(20:4(5Z8Z11Z14Z)P-18:1(11Z)); PC(20:4(5Z8Z11Z14Z)P-18:1(9Z)); PC(20:4(8Z11Z14Z17Z)P-18:1(11Z)); PC(20:4(8Z11Z14Z17Z)P-18:1(9Z)); PC(20:5(5Z8Z11Z14Z17Z)P-18:0); PC(22:5(4Z7Z10Z13Z16Z)P-16:0); PC(22:5(7Z10Z13Z16Z19Z)P-16:0); PC(P-16:022:5(4Z7Z10Z13Z16Z)); PC(P-16:022:5(7Z10Z13Z16Z19Z)); PC(P-18:020:5(5Z8Z11Z14Z17Z)); PC(P-18:1(11Z)20:4(5Z8Z11Z14Z)); PC(dm18:1(11Z)20:4(8Z11Z14Z17Z)); PC(P-18:1(9Z)20:4(5Z8Z11Z14Z)); PC(P-18:1(9Z)20:4(8Z11Z14Z17Z)); PC(o-16:022:6(4Z7Z10Z13Z16Z19Z)); 2-O-(4710131619-Docosahexaenoyl)-1-O-hexadecylglycero-3-phosphocholine | HMDB08457; HMDB08458; HMDB08490; HMDB08491; HMDB08522; HMDB08685; HMDB08718; HMDB11227; HMDB11228; HMDB11255; HMDB11286; HMDB11287; HMDB11319; HMDB11320; HMDB13409; HMDB39529 | M+H_[+1] | - | - |
| HILIC positive | 189.1234 | 257.9 | -0.17 (0.57) | 0.35 (0.59) | -0.52 (-0.99, -0.05) | 0.0341 | 0.8637 | N6-Acetyl-L-lysine; N-Alpha-acetyllysine; Glycyl-L-leucine; Alanyl-Valine; Glycyl-Isoleucine; Isoleucyl-Glycine; Leucyl-Glycine; Valyl-Alanine | HMDB00206; HMDB00446; HMDB00759; HMDB28700; HMDB28844; HMDB28907; HMDB28929; HMDB29120 | M+H | - | - |
| C18 negative | 453.1355 | 64 | -0.17 (1.09) | 0.49 (0.67) | -0.66 (-1.26, -0.06) | 0.0341 | 0.8508 | epsilon-Viniferin; trans-delta-Viniferin; Ampelopsin D | HMDB30604; HMDB32657; HMDB40894 | M-H | - | - |
| C18 negative | 243.0621 | 23.5 | 0.51 (0.87) | -0.47 (1.06) | 0.98 (0.09, 1.87) | 0.0345 | 0.8508 | Uridine; Pseudouridine | HMDB00296; HMDB00767 | M-H | Uridine | M-H |
| HILIC positive | 147.0764 | 77.1 | -0.11 (0.93) | 0.69 (0.66) | -0.8 (-1.54, -0.07) | 0.0350 | 0.8637 | Pyroglutamic acid; L-Glutamine; Pyrrolidonecarboxylic acid; Pyrroline hydroxycarboxylic acid; N-Acryloylglycine; Ureidoisobutyric acid; 1-Pyrroline-4-hydroxy-2-carboxylate; D-Glutamine; Alanylglycine; Alanyl-Glycine; 5-Oxoprolinate; dimethadione | HMDB00267; HMDB00641; HMDB00805; HMDB01369; HMDB01843; HMDB02031; HMDB02234; HMDB03423; HMDB06899; HMDB28687; HMDB60262; HMDB61093 | M+NH4; M+H | Glutamine | M+H |
| C18 negative | 227.1652 | 292.4 | 0.32 (0.84) | -0.26 (0.46) | 0.58 (0.05, 1.11) | 0.0350 | 0.8508 | 11-Hydroxy-9-tridecenoic acid; Menthone 12-glyceryl ketal; Menthyl lactate; Menthone 13-glyceryl ketal | HMDB35881; HMDB36139; HMDB37212; HMDB40004 | M-H | - | - |
| HILIC positive | 148.0798 | 75.8 | -0.12 (0.97) | 0.68 (0.69) | -0.79 (-1.52, -0.07) | 0.0354 | 0.8637 | L-Lysine; L-Glutamine; Ureidoisobutyric acid; D-Lysine; D-Glutamine; Alanylglycine; (3S)-36-Diaminohexanoate; (3S5S)-35-Diaminohexanoate; Alanyl-Glycine | HMDB00182; HMDB00641; HMDB02031; HMDB03405; HMDB03423; HMDB06899; HMDB12114; HMDB12115; HMDB28687 | M+H_[+1] | - | - |
| HILIC positive | 132.0656 | 70.7 | -0.04 (0.65) | 0.51 (0.96) | -0.55 (-1.06, -0.04) | 0.0358 | 0.8637 | 4-Hydroxyproline; N-Acetyl-L-alanine; Propionylglycine; 5-Aminolevulinic acid; L-Glutamic gamma-semialdehyde; 3-Hydroxy-L-proline; 4-Hydroxy-L-proline; 5-Amino-2-oxopentanoic acid; 4-Hydroxy-2-pyrrolidinecarboxylic acid; Trans-3-hydroxy-L-proline; cis-4-Hydroxy-D-proline | HMDB00725; HMDB00766; HMDB00783; HMDB01149; HMDB02104; HMDB02113; HMDB06055; HMDB06272; | M+H | Hydroxypr oline | M+H |

| | | | | | | | | | | | | |
|---|---|---|---|---|---|---|---|---|---|---|---|---|
| | | | | | | | | | HMDB36576; HMDB59659; HMDB60460 | | | |
| HILIC positive | 130.0499 | 76.6 | -0.14 (1.05) | 0.66 (0.65) | -0.81 (-1.55, -0.06) | 0.0361 | 0.8637 | Pyroglutamic acid; Pyrrolidonecarboxylic acid; Pyrroline hydroxycarboxylic acid; N-Acryloylglycine; 1-Pyrroline-4-hydroxy-2-carboxylate; 5-Oxoprolinate; dimethadione | HMDB00267; HMDB00805; HMDB01369; HMDB01843; HMDB02234; HMDB60262; HMDB61093 | M+H | - | - |
| HILIC positive | 505.1562 | 74.9 | -0.06 (0.6) | 0.38 (0.37) | -0.44 (-0.84, -0.03) | 0.0363 | 0.8637 | 6-Caffeoylsucrose | HMDB35486 | M+H | - | - |
| C18 negative | 166.0622 | 294.2 | -1.08 (1.27) | 0.06 (0.89) | -1.14 (-2.2, -0.09) | 0.0364 | 0.8508 | (S)-Spinacine; 24-Diamino-6-nitrotoluene | HMDB29873; HMDB60362 | M-H | - | - |
| HILIC positive | 124.1121 | 284.1 | -0.57 (1.69) | 0.45 (0.82) | -1.02 (-1.96, -0.08) | 0.0364 | 0.8637 | 2-(1-Propenyl)-delta1-piperideine | HMDB33365 | M+H | - | - |
| HILIC positive | 476.3793 | 27.2 | 0.45 (0.82) | -0.22 (0.8) | 0.67 (0.05, 1.29) | 0.0367 | 0.8637 | Soyasapogenol A; Camelliagenin A; Priverogenin B; (3alphaOH20S24S)-319:2024-Diepoxydammarane-325-diol; Ganoderiol A; 2024-Epoxy-2526-dihydroxydammaran-3-one | HMDB34505; HMDB34528; HMDB34644; HMDB34683; HMDB35326; HMDB39692 | M+H_[+1] | - | - |
| C18 negative | 347.1501 | 139.8 | -0.05 (1.07) | -0.87 (0.87) | 0.82 (0.06, 1.59) | 0.0368 | 0.8508 | Gibberellin A60; Gibberellin A58; Gibberellin A63; Gibberellin A54; Gibberellin A67; Gibberellin A77; 8-Angeloylegelolide; Gibberellin A81; Triphasiol; Gibberellin A90; 3-Epigibberellin A1 | HMDB34988; HMDB35043; HMDB35044; HMDB35048; HMDB35049; HMDB37004; HMDB37773; HMDB38080; HMDB39062; HMDB39622; HMDB40944 | M-H | - | - |
| HILIC positive | 142.9837 | 265.6 | 0.12 (0.74) | -0.39 (0.41) | 0.51 (0.04, 0.97) | 0.0370 | 0.8637 | Mesna | HMDB03745 | M+H | - | - |
| HILIC positive | 148.0736 | 75 | -0.11 (0.99) | 0.69 (0.69) | -0.8 (-1.54, -0.06) | 0.0372 | 0.8637 | L-Lysine; 2-Methylglutaric acid; Adipic acid; L-Glutamine; Methylglutaric acid; Monomethyl glutaric acid; Ureidoisobutyric acid; 22-Dimethylsuccinic acid; Solerol; D-Lysine; D-Glutamine; Alanylglycine; (S)-2-Aceto-2-hydroxybutanoic acid; (3S)-36-Diaminohexanoate; (3S5S)-35-Diaminohexanoate; Alanyl-Glycine; Dimethyl succinate | HMDB00182; HMDB00422; HMDB00448; HMDB00641; HMDB00752; HMDB00858; HMDB02031; HMDB02074; HMDB02173; HMDB03405; HMDB03423; HMDB06899; HMDB06900; HMDB12114; HMDB12115; HMDB28687; HMDB33837 | M+H_[+1] | - | - |
| HILIC positive | 163.1328 | 29.8 | -0.9 (1.85) | 0.49 (0.32) | -1.39 (-2.69, -0.09) | 0.0389 | 0.8637 | (3R7R)-137-Octanetriol | HMDB33625 | M+H | - | - |
| HILIC positive | 325.3096 | 27.2 | 0.38 (0.91) | -0.16 (0.82) | 0.54 (0.03, 1.05) | 0.0391 | 0.8637 | 68-Heneicosanedione; 46-Heneicosanedione | HMDB35570; HMDB35571 | M+H | - | - |
| HILIC positive | 211.1441 | 31.1 | -0.32 (0.36) | 0.16 (0.77) | -0.48 (-0.92, -0.03) | 0.0391 | 0.8637 | LL-Cyclo(leucylprolyl) | HMDB34276 | M+H | - | - |
| C18 negative | 857.5185 | 219 | -0.49 (0.88) | 0.52 (0.89) | -1 (-1.94, -0.06) | 0.0394 | 0.8508 | PI(16:020:4(5Z8Z11Z14Z)); PI(16:020:4(8Z11Z14Z17Z)); PI(18:1(11Z)18:3(6Z9Z12Z)); PI(18:1(11Z)18:3(9Z12Z15Z)); PI(18:1(9Z)18:3(6Z9Z12Z)); PI(18:1(9Z)18:3(9Z12Z15Z)); PI(18:2(9Z12Z)18:2(9Z12Z)); PI(18:3(6Z9Z12Z)18:1(11Z)); PI(18:3(6Z9Z12Z)18:1(9Z)); PI(18:3(9Z12Z15Z)18:1(11Z)); PI(18:3(9Z12Z15Z)18:1(9Z)); PI(20:4(5Z8Z11Z14Z)16:0); PI(20:4(8Z11Z14Z17Z)16:0) | HMDB09789; HMDB09790; HMDB09827; HMDB09828; HMDB09839; HMDB09840; HMDB09850; HMDB09859; HMDB09860; HMDB09863; HMDB09864; HMDB09893; HMDB09899 | M-H | - | - |
| HILIC positive | 314.0748 | 96.1 | 0.27 (0.58) | 0.58 (0.3) | -0.31 (-0.61, -0.02) | 0.0396 | 0.8637 | Phosphoribosylformylglycineamidine; 5-phosphoribosyl-a-N-formylglycineamidine | HMDB00999; HMDB02305 | M+H | - | - |
| C18 negative | 257.2122 | 294.6 | -0.44 (0.87) | 0.59 (0.78) | -1.03 (-2.01, -0.06) | 0.0399 | 0.8508 | 10-Hydroxymyristic acid methyl ester; Ethyl 3-hydroxytridecanoate | HMDB31061; HMDB59866 | M-H | - | - |
| HILIC positive | 293.1454 | 75.1 | 0.01 (0.73) | 0.67 (0.51) | -0.66 (-1.28, -0.04) | 0.0401 | 0.8637 | L-Glutamine; Ureidoisobutyric acid; D-Glutamine; Alanylglycine; Alanyl-Glycine | HMDB00641; HMDB02031; HMDB03423; HMDB06899; HMDB28687 | 2M+H | - | - |
| HILIC positive | 330.2638 | 28.1 | 0.41 (0.41) | -0.01 (0.39) | 0.42 (0.02, 0.82) | 0.0404 | 0.8637 | 48 Dimethylnonanoyl carnitine; Undecanoylcarnitine | HMDB06202; HMDB13321 | M+H | - | - |
| HILIC positive | 372.3112 | 27.4 | 0.41 (0.69) | -0.21 (0.28) | 0.61 (0.03, 1.2) | 0.0406 | 0.8637 | Tetradecanoylcarnitine | HMDB05066 | M+H | - | - |
| C18 negative | 161.0819 | 290 | 0.33 (0.97) | -0.48 (0.95) | 0.81 (0.04, 1.58) | 0.0406 | 0.8508 | Dimethicone; 2-(1-Ethoxyethoxy)propanoic acid | HMDB33532; HMDB59918 | M-H | - | - |
| C18 negative | 367.1472 | 25.4 | -0.14 (0.92) | 0.45 (0.29) | -0.59 (-1.15, -0.03) | 0.0408 | 0.8508 | N-Desmethyleletriptan | HMDB13919 | M-H | - | - |
| C18 negative | 566.1816 | 142.8 | 0.69 (0.81) | -0.3 (0.97) | 0.99 (0.05, 1.94) | 0.0412 | 0.8508 | Neoacrimarine H | HMDB33031 | M-H | - | - |
| HILIC positive | 651.2291 | 75 | -0.04 (0.74) | 0.53 (0.22) | -0.58 (-1.13, -0.03) | 0.0415 | 0.8637 | 5-Hydroxy-734-trimethoxy-8-methylisoflavone 5-neohesperidoside; Egonol gentiobioside | HMDB30627; HMDB34281 | M+H | - | - |
| HILIC positive | 517.3881 | 26.8 | -0.52 (1.36) | 0.56 (0.34) | -1.08 (-2.11, -0.05) | 0.0416 | 0.8637 | 22-Acetylpriverogenin B | HMDB34645 | M+H | - | - |
| HILIC positive | 179.0024 | 181.7 | 0.29 (0.5) | 0.57 (0.23) | -0.28 (-0.55, -0.01) | 0.0417 | 0.8637 | Allitridin | HMDB31154 | M+H | - | - |
| HILIC positive | 101.0709 | 75.7 | -0.07 (0.94) | 0.69 (0.66) | -0.77 (-1.5, -0.03) | 0.0418 | 0.8637 | N-Nitroso-pyrrolidine; Gyromitrin | HMDB31642; HMDB33952 | M+H | - | - |
| C18 negative | 471.3485 | 237 | -0.56 (1.27) | 0.45 (0.49) | -1.01 (-1.97, -0.04) | 0.0419 | 0.8508 | Maslinic acid; Lucidumol A; Queretaroic acid; Priverogenin A; (3beta23xi)-323-Dihydroxycycloart-24-en-26-oic acid; Pomolic acid; 20beta-Hydroxyursolic acid; Azukisapogenol; delta-Maslinic acid; Sebiferenic acid; Momordicin I; Ganoderic acid U; Ganodermanontriol; Epoxyganoderiol A; (3b22S24E)-322-Dihydroxycycloart-24-en-26-oic acid; 27-Hydroxyisomangiferolic acid; Albigenic acid; Rubitic acid; Ganoderiol E; 3alpha-Corosolic acid | HMDB02392; HMDB33233; HMDB34523; HMDB34530; HMDB34567; HMDB35106; HMDB35107; HMDB35258; HMDB35260; HMDB35748; HMDB35758; HMDB35962; HMDB35977; HMDB36001; HMDB36063; HMDB36064; HMDB36638; HMDB36654; HMDB37779; HMDB41043 | M-H | - | - |

| | | | | | | | | | | | | |
|---|---|---|---|---|---|---|---|---|---|---|---|---|
| C18 negative | 533.2556 | 147.1 | 0.64 (0.4) | 0.3 (0.33) | 0.33 (0.01, 0.65) | 0.0419 | 0.8508 | Pyrophaeophorbide a | HMDB31150 | M-H | - | - |
| C18 negative | 292.0557 | 19.1 | 0.58 (0.26) | 0.71 (0.2) | -0.14 (-0.26, -0.01) | 0.0420 | 0.8508 | Canavaninosuccinate; Lumiracoxib | HMDB12197; HMDB15403 | M-H_[-1]; M-H | - | - |
| HILIC positive | 768.5898 | 30.9 | -0.49 (0.93) | 0.21 (0.77) | -0.7 (-1.37, -0.03) | 0.0423 | 0.8637 | PC(18:2(9Z12Z)P-18:1(11Z)); PC(18:2(9Z12Z)P-18:1(9Z)); PC(18:3(6Z9Z12Z)P-18:0); PC(18:3(9Z12Z15Z)P-18:0); PC(20:3(5Z8Z11Z)P-16:0); PC(20:3(8Z11Z14Z)P-16:0); PC(P-16:020:3(5Z8Z11Z)); PC(P-16:020:3(8Z11Z14Z)); PC(P-18:018:3(6Z9Z12Z)); PC(P-18:018:3(9Z12Z15Z)); PC(P-18:1(11Z)18:2(9Z12Z)); PC(P-18:1(9Z)18:2(9Z12Z)); PC(o-16:020:4(8Z11Z14Z17Z)); PC(o-18:2(9Z12Z)18:2(9Z12Z)) | HMDB08161; HMDB08162; HMDB08193; HMDB08226; HMDB08390; HMDB08422; HMDB11218; HMDB11219; HMDB11245; HMDB11246; HMDB11277; HMDB11310; HMDB13407; HMDB13435 | M+H | - | - |
| C18 negative | 145.0619 | 20.8 | -0.51 (1.2) | 0.41 (0.75) | -0.92 (-1.8, -0.04) | 0.0429 | 0.8508 | L-Glutamine; Ureidoisobutyric acid; D-Glutamine; Alanylglycine; Alanyl-Glycine | HMDB00641; HMDB02031; HMDB03423; HMDB06899; HMDB28687 | M-H | - | - |
| HILIC positive | 131.0562 | 75.3 | -0.06 (0.86) | 0.58 (0.57) | -0.64 (-1.25, -0.02) | 0.0437 | 0.8637 | Pipecolic acid; Pyroglutamic acid; L-Pipecolic acid; Pyrrolidonecarboxylic acid; Pyrroline hydroxycarboxylic acid; N-Acryloylglycine; 1-Pyrroline-4-hydroxy-2-carboxylate; N4-Acetylaminobutanal; D-Pipecolic acid; Vigabatrin; L-trans-4-Methyl-2-pyrrolidinecarboxylic acid; 2-Pyrrolidineacetic acid; 3-Acetamidobutanal; 5-Oxoprolinate; dimethadione | HMDB00070; HMDB00267; HMDB00716; HMDB00805; HMDB01369; HMDB01843; HMDB02234; HMDB04226; HMDB05960; HMDB15212; HMDB29435; HMDB29444; HMDB59649; HMDB60262; HMDB61093 | M+H_[+1] | - | - |
| C18 negative | 910.5527 | 225.3 | 0.02 (0.34) | 0.49 (0.67) | -0.47 (-0.93, -0.02) | 0.0437 | 0.8508 | PI(18:022:6(4Z7Z10Z13Z16Z19Z)); PI(18:3(6Z9Z12Z)22:3(10Z13Z16Z)); PI(18:3(9Z12Z15Z)22:3(10Z13Z16Z)); PI(20:3(5Z8Z11Z)20:3(5Z8Z11Z)); PI(20:3(5Z8Z11Z)20:3(8Z11Z14Z)); PI(20:3(8Z11Z14Z)20:3(5Z8Z11Z)); PI(20:3(8Z11Z14Z)20:3(8Z11Z14Z)); PI(22:3(10Z13Z16Z)18:3(6Z9Z12Z)); PI(22:3(10Z13Z16Z)18:3(9Z12Z15Z)); PI(22:6(4Z7Z10Z13Z16Z19Z)18:0) | HMDB09821; HMDB09861; HMDB09865; HMDB09884; HMDB09885; HMDB09891; HMDB09892; HMDB09910; HMDB09911; HMDB09920 | M-H_[-1] | - | - |
| HILIC positive | 223.2057 | 290.2 | 0.44 (0.61) | -0.57 (1.26) | 1 (0.03, 1.97) | 0.0437 | 0.8637 | Farnesol; beta-Triticene; alpha-Triticene; Pogostol; Valerianol; 10-Epijunenol; (-)-Pacifigorgiol; Sesquisabinene hydrate; 4beta-8-Daucen-4-ol; 6-Epi-beta-bisabolol; Panasinsanol B; Nerolidol; (S)-Nerolidol; Panasinsanol A; Shyobunol; Rosifoliol; beta-Bisabolol; (-)-alpha-Bisabolol; ()-alpha-Bisabolol; (-)-Valeranone; Bulnesol; Guaioxide; alpha-Cadinol; Carotol; 58-Epoxydaucane; Isointermedeol; Globulol; Cedrelanol; Faurinone; alpha-Caryophyllene alcohol; Sesquicineol; Epicubenol; beta-Caryophyllene alcohol; Zingiberenol; Koraiol; 3xi-4-Copanol; Maalialcohol; Humulol; (6Z9Z)-69-Pentadecadien-2-one; Ginsenol; Chamomillol; Isokessane; Cubenol; (Z)-Farnesol | HMDB04305; HMDB30938; HMDB30939; HMDB33239; HMDB34951; HMDB34982; HMDB35021; HMDB35199; HMDB35351; HMDB35364; HMDB35383; HMDB35662; HMDB35663; HMDB35711; HMDB35811; HMDB35868; HMDB36111; HMDB36197; HMDB36198; HMDB36421; HMDB36445; HMDB36447; HMDB36452; HMDB36579; HMDB36580; HMDB36642; HMDB36645; HMDB36646; HMDB36679; HMDB36732; HMDB36908; HMDB37031; HMDB37066; HMDB37067; HMDB37234; HMDB37394; HMDB38167; HMDB38213; HMDB39538; HMDB39744; HMDB40422; HMDB41575; HMDB59841; HMDB59849 | M+H | - | - |
| HILIC positive | 325.2006 | 225.3 | -0.98 (1.93) | 0.28 (0.39) | -1.26 (-2.48, -0.04) | 0.0438 | 0.8637 | Lactapiperanol D; Cibaric acid | HMDB33631; HMDB38580 | M+H | - | - |
| HILIC positive | 276.1806 | 236 | -0.24 (1.03) | 0.47 (0.53) | -0.71 (-1.4, -0.02) | 0.0439 | 0.8637 | Hydroxyhexanoycarnitine | HMDB13131 | M+H | - | - |
| HILIC positive | 359.3154 | 27.5 | 0.35 (1.33) | -0.36 (1.04) | 0.7 (0.02, 1.38) | 0.0441 | 0.8637 | MG(18:00:00:0); MG(0:018:00:0); Glycerol 1-octadecanoate | HMDB11131; HMDB11535; HMDB31075 | M+H | - | - |
| HILIC positive | 819.6081 | 29.8 | -0.18 (0.52) | 0.31 (0.49) | -0.49 (-0.97, -0.02) | 0.0441 | 0.8637 | PC(22:5(4Z7Z10Z13Z16Z)P-18:1(11Z)); PC(22:5(4Z7Z10Z13Z16Z)P-18:1(9Z)); PC(22:5(7Z10Z13Z16Z19Z)P-18:1(11Z)); PC(22:5(7Z10Z13Z16Z19Z)P-18:1(9Z)); PC(22:6(4Z7Z10Z13Z16Z19Z)P-18:0); PC(P-18:022:6(4Z7Z10Z13Z16Z19Z)); PC(P-18:1(11Z)22:5(4Z7Z10Z13Z16Z)); PC(P-18:1(11Z)22:5(7Z10Z13Z16Z19Z)); PC(P- | HMDB08687; HMDB08688; HMDB08720; HMDB08721; HMDB08752; HMDB11262; HMDB11293; HMDB11294; HMDB11326; HMDB11327 | M+H_[+1] | - | - |

| | | | | | | | | | | | | |
|---|---|---|---|---|---|---|---|---|---|---|---|---|
| | | | | | | | | 18:1(9Z)22:5(4Z7Z10Z13Z16Z)); PC(P-18:1(9Z)22:5(7Z10Z13Z16Z19Z)) | | | | |
| C18 negative | 858.522 | 219 | -0.47 (0.89) | 0.5 (0.87) | -0.97 (-1.91, -0.03) | 0.0442 | 0.8508 | PI(16:020:4(5Z8Z11Z14Z)); PI(16:020:4(8Z11Z14Z17Z)); PI(18:1(11Z)18:3(6Z9Z12Z)); PI(18:1(11Z)18:3(9Z12Z15Z)); PI(18:1(9Z)18:3(6Z9Z12Z)); PI(18:1(9Z)18:3(9Z12Z15Z)); PI(18:2(9Z12Z)18:2(9Z12Z)); PI(18:3(6Z9Z12Z)18:1(11Z)); PI(18:3(6Z9Z12Z)18:1(9Z)); PI(18:3(9Z12Z15Z)18:1(11Z)); PI(18:3(9Z12Z15Z)18:1(9Z)); PI(20:4(5Z8Z11Z14Z)16:0); PI(20:4(8Z11Z14Z17Z)16:0) | HMDB09789; HMDB09790; HMDB09827; HMDB09828; HMDB09839; HMDB09840; HMDB09850; HMDB09859; HMDB09860; HMDB09863; HMDB09864; HMDB09893; HMDB09899 | M-H_[-1] | - | - |
| C18 negative | 449.3286 | 216.8 | -0.71 (1.45) | 0.53 (0.75) | -1.23 (-2.42, -0.04) | 0.0443 | 0.8508 | Coprocholic acid; 3a7a12a-Trihydroxy-5b-cholestanoic acid; 3alpha7alpha12alpha25-Tetrahydroxy-5beta-cholestane-24-one | HMDB00601; HMDB03873; HMDB60137 | M-H | - | - |
| C18 negative | 378.2404 | 254 | 0.44 (0.4) | -0.79 (1.83) | 1.23 (0.04, 2.42) | 0.0443 | 0.8508 | Sphingosine 1-phosphate; N-palmitoyl-phosphoethanolamine | HMDB00277; HMDB59630 | M-H | - | - |
| HILIC positive | 216.1595 | 110.2 | -0.26 (0.76) | 0.39 (0.54) | -0.65 (-1.28, -0.02) | 0.0449 | 0.8637 | N-Nonanoylglycine | HMDB13279 | M+H | - | - |
| HILIC positive | 149.0832 | 75.2 | 0.22 (0.29) | 0.42 (0.2) | -0.21 (-0.41, -0.01) | 0.0451 | 0.8637 | L-Glutamine; Ureidoisobutyric acid; D-Glutamine; Alanylglycine; Alanyl-Glycine | HMDB00641; HMDB02031; HMDB03423; HMDB06899; HMDB28687 | M+H_[+2] | - | - |
| C18 negative | 140.011 | 15.8 | 0.12 (0.99) | -0.84 (1.49) | 0.96 (0.02, 1.89) | 0.0454 | 0.8508 | 5-Hydroxymethyl-2-furanoate | HMDB29188 | M-H | - | - |
| HILIC positive | 201.9838 | 160.9 | 0.39 (0.34) | -0.03 (0.75) | 0.42 (0.01, 0.82) | 0.0454 | 0.8637 | Cysteine-S-sulfate | HMDB00731 | M+H | - | - |
| C18 negative | 275.1503 | 294.6 | 0.28 (0.88) | -0.51 (0.81) | 0.79 (0.02, 1.56) | 0.0455 | 0.8508 | 4-Hydroxycinnamoylagmatine | HMDB33460 | M-H | - | - |
| HILIC positive | 360.3188 | 27.4 | 0.37 (1.31) | -0.36 (1.04) | 0.73 (0.02, 1.45) | 0.0455 | 0.8637 | MG(18:00:00:0); MG(0:018:00:0); Glycerol 1-octadecanoate | HMDB11131; HMDB11535; HMDB31075 | M+H_[+1] | - | - |
| HILIC positive | 275.0663 | 279.8 | 0.38 (0.88) | 0.94 (0.24) | -0.56 (-1.1, -0.01) | 0.0458 | 0.8637 | cistrans-5-Hydroxythalidomide; 5-Hydroxythalidomide; Thalidomide arene oxide | HMDB13870; HMDB13871; HMDB13872 | M+H | - | - |
| HILIC positive | 237.1122 | 294 | -0.24 (1.02) | 0.25 (0.63) | -0.49 (-0.97, -0.01) | 0.0463 | 0.8637 | Ethyl vanillin isobutyrate; Carboxy-ibuprofen | HMDB37683; HMDB60564 | M+H | - | - |
| HILIC positive | 235.1692 | 295.9 | 0.4 (0.35) | -0.12 (0.64) | 0.52 (0.01, 1.03) | 0.0475 | 0.8637 | Valerenic acid; Epiacoronene; 410-Longipinanedione; Curcumanolide A; Curcumenone; Curcumenol; 9-Pentadecene-1214-diyne-111-diol; Saussurea lactone; Oxysolavetivone; 7-Hydroxycostal; Dehydrocurdione; 110-Epoxygermacrone; (4alpha5alpha)-11-Eremophilene-29-dione; Petasalbin; beta-Costic acid; alpha-Rotunol; Germacrone 45-epoxide; Dihydroisoalantolactone; Marasmene; 3-Phenylpropyl hexanoate; (Z)-alpha-Bergamotenoic acid; Procurcumenol; (1alpha4beta5beta)-4-Hydroxy-7(11)10(14)-guaiadien-8-one; Zerumbone oxide; Isocurcumenol; Cyclodehydroisolubimin; p-Hydroxynonanophenone; 2-Methyl-4-phenyl-2-butyl 2-methylpropanoate; 4-Methylphenyl octanoate; 7-(13-Cyclohexadienyl)-5-hydroxy-26-dimethyl-2-hepten-4-one; Rishitinol; 37-Bisaboladiene-28-dione; xi-2-Hydroxy-135-bisabolatrien-9-one; 79-Illudadiene-314-diol; 27(14)-Illudadiene-1015-diol; 46-Decadiyn-1-ol isovalerate; beta-Santalic acid; Sugeonol | HMDB30016; HMDB31349; HMDB32081; HMDB33252; HMDB33253; HMDB33960; HMDB34562; HMDB34742; HMDB34956; HMDB35202; HMDB35407; HMDB35408; HMDB35413; HMDB35639; HMDB35794; HMDB35869; HMDB35889; HMDB35935; HMDB36035; HMDB36390; HMDB36392; HMDB36427; HMDB36449; HMDB36466; HMDB36647; HMDB37038; HMDB37391; HMDB37640; HMDB37710; HMDB37739; HMDB38190; HMDB38198; HMDB38698; HMDB38795; HMDB38796; HMDB38914; HMDB39621; HMDB41036 | M+H | - | - |
| HILIC positive | 178.0863 | 284.9 | -0.08 (0.96) | -1 (0.75) | 0.93 (0.01, 1.85) | 0.0484 | 0.8637 | 5-Hydroxytryptophol; 12-Dehydrosalsolinol; Plantagonine; 2-Propenyl 2-aminobenzoate; 5-2H-Pyrrol-4-(3H)-ylidenemethyl-2-furanmethanol; NN-dimethylindoliumolate | HMDB01855; HMDB12490; HMDB33482; HMDB37693; HMDB40045; HMDB60063 | M+H | - | - |
| C18 negative | 498.3201 | 188.6 | -1.21 (2.24) | 0.24 (0.37) | -1.45 (-2.89, -0.01) | 0.0485 | 0.8508 | Hydroxybuprenorphine | HMDB60547 | M-H | - | - |
| C18 negative | 353.2333 | 152 | 0.6 (0.27) | 0.17 (0.75) | 0.43 (0, 0.85) | 0.0485 | 0.8508 | Prostaglandin F2a; Prostaglandin E1; Prostaglandin F2b; Troxilin B3; Trioxilin A3; 8-isoprostaglandin PGF2b; 111215-THETA; 1314-Dihydro-15-keto PGF2a; 8-Isoprostaglandin E1; 111415-THETA; 8-Isoprostaglandin F2a; Prostaglandin D1; 8-iso-1314-dihydro-15-keto-PGF2a; 111215-TriHETRE; 11b-PGF2a; 11-Epi-PGF2a; 1314-Dihydro- lipoxin A4; Prostaglandin H1; Sterebin G; 15-keto-PGF1alpha; (5Z9E12S14Z)-81112-Trihydroxyicosa-5914-trienoate | HMDB01139; HMDB01442; HMDB01483; HMDB01965; HMDB01977; HMDB02115; HMDB04684; HMDB04685; HMDB04686; HMDB04694; HMDB05083; HMDB05102; HMDB06562; HMDB10170; HMDB10199; HMDB11137; HMDB12563; HMDB13041; HMDB35380; HMDB60045; HMDB60107 | M-H | - | - |
| HILIC positive | 782.6096 | 31.7 | -0.35 (0.97) | 0.34 (1.01) | -0.69 (-1.37, -0.01) | 0.0486 | 0.8637 | PE(22:2(13Z16Z)P-18:1(11Z)); PE(22:2(13Z16Z)P-18:1(9Z)); PE(P-18:1(11Z)22:2(13Z16Z)); PE(P-18:1(9Z)22:2(13Z16Z)) | HMDB09578; HMDB09579; HMDB11423; HMDB11456 | M+H | - | - |
| HILIC positive | 386.29 | 29.4 | 0.5 (0.38) | 0.15 (0.3) | 0.35 (0, 0.71) | 0.0492 | 0.8637 | 3-Hydroxy-cis-5-tetradecenoylcarnitine | HMDB13330 | M+H | - | - |
| HILIC positive | 326.3051 | 27.8 | 0.65 (0.62) | -0.13 (0.99) | 0.78 (0, 1.56) | 0.0496 | 0.8637 | N-Oleoylethanolamine | HMDB02088 | M+H | - | - |
| C18 negative | 147.0452 | 25.3 | -0.43 (1.2) | 0.3 (0.83) | -0.73 (-1.46, 0) | 0.0500 | 0.8508 | Cinnamic acid; trans-Cinnamic acid; (E)-3-(2-Hydroxyphenyl)-2-propenal; Di-2-furanylmethane; 1-Phenyl-12-propanedione; 34-Dihydro-2H-1-benzopyran-2-one; (E)-3-(4-Hydroxyphenyl)-2-propenal | HMDB00567; HMDB00930; HMDB31725; HMDB32947; HMDB35243; HMDB36626; HMDB40986 | M-H | Cinnamic acid | M-H |

SD: Standard Deviation; CI: Confidence Interval; FDR: False Discovery Rate.

**Supplemental Figure 1.** GWAS Resistance & Infection Hierarchy.

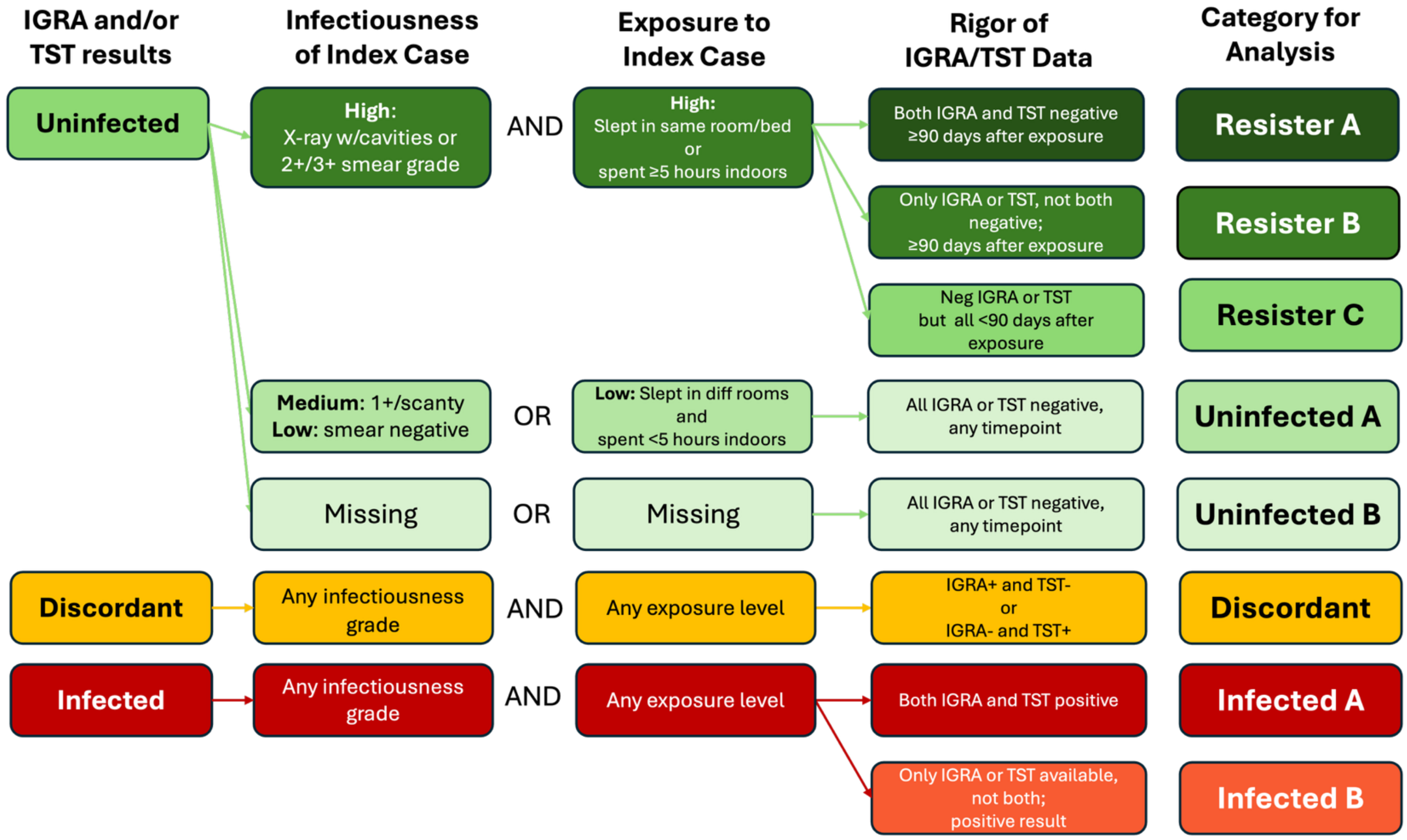

**Supplemental Figure 2.** Volcano Plot of the Metabolome-wide Association Study Comparing TB Resisters and Controls in the Overall Cohort (269 Pairs). Horizontal Line Represents P Value of 0.05. Confirmed Metabolites are Labeled.

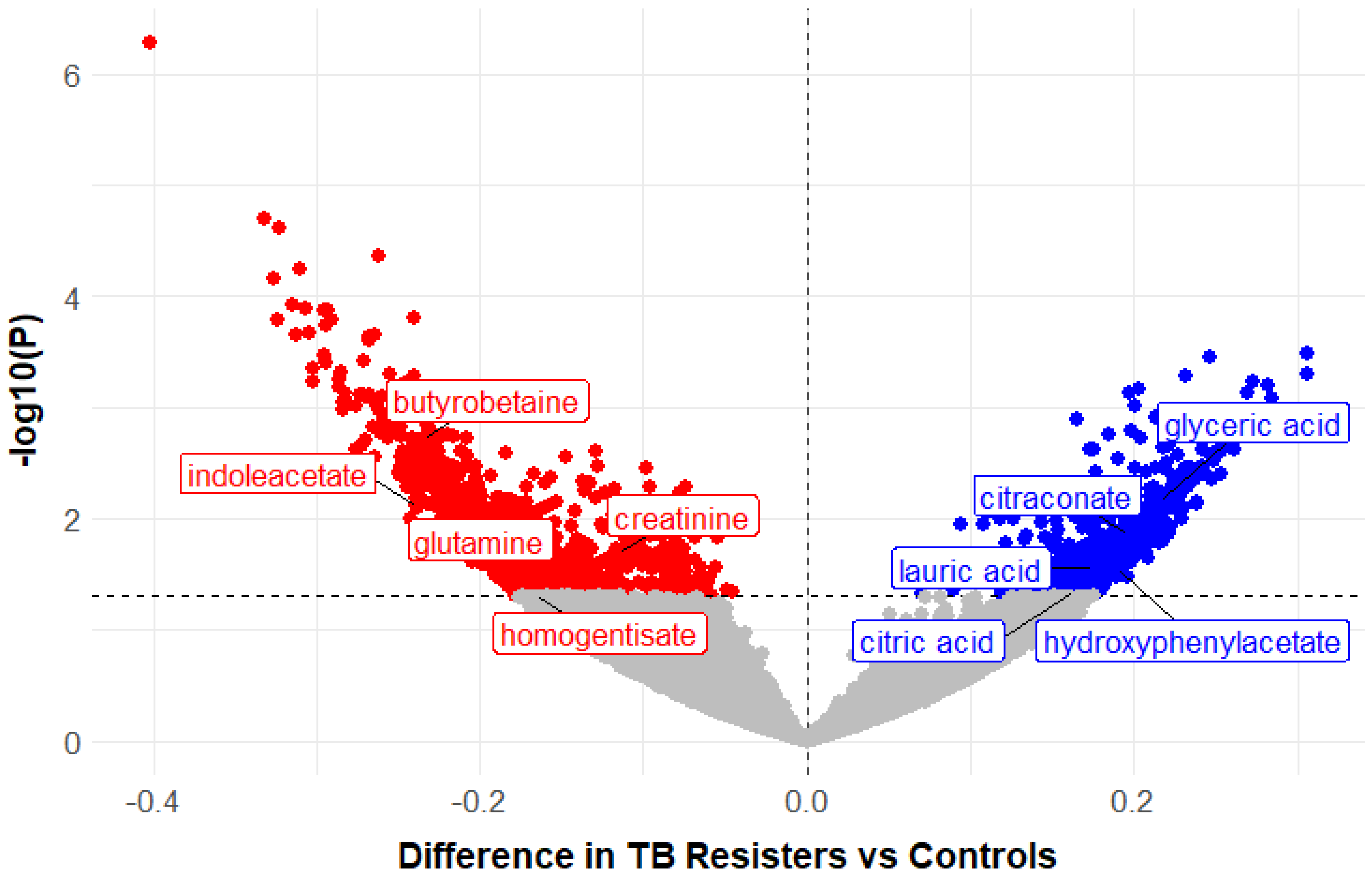

**Supplemental Figure 3.** Volcano Plot of the Metabolome-wide Association Study Comparing TB Resisters and Controls in the Brazilian Cohort (155 Pairs). Horizontal Line Represents P Value of 0.05. Confirmed Metabolites are Labeled.

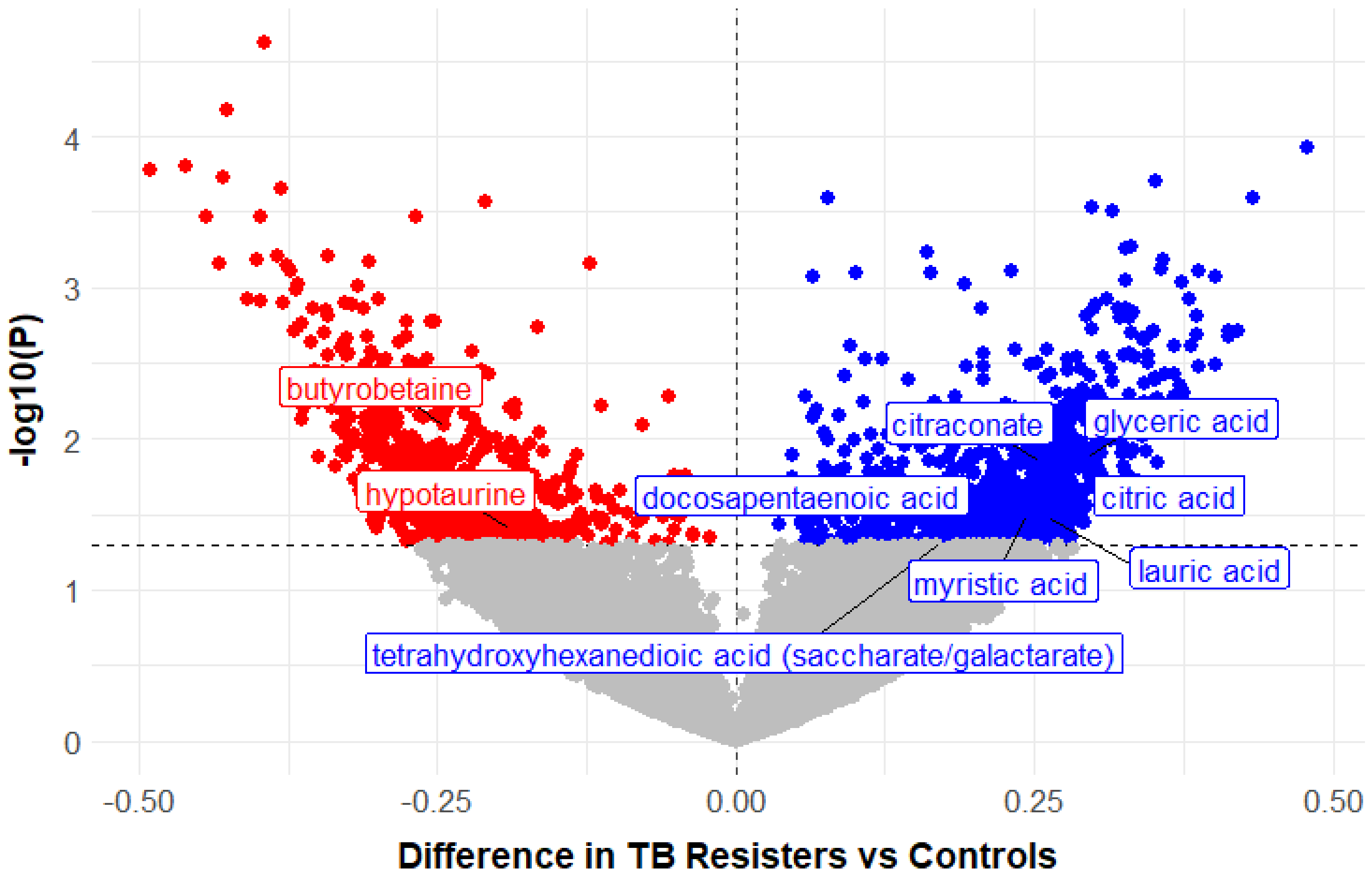

**Supplemental Figure 4.** Volcano Plot of the Metabolome-wide Association Study Comparing TB Resisters and Controls in the Indian Cohort (103 Pairs). Horizontal Line Represents P Value of 0.05. Confirmed Metabolites are Labeled.

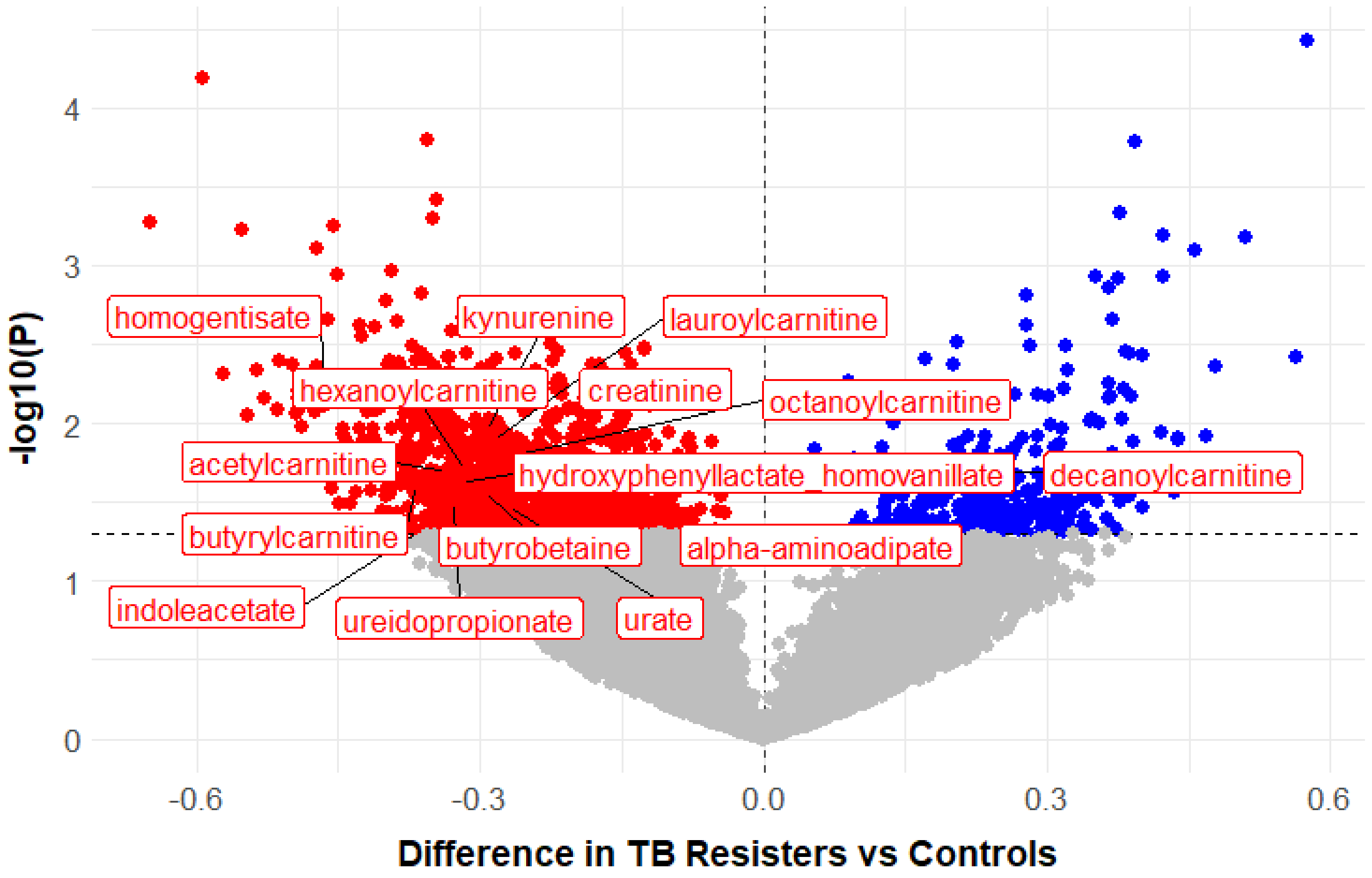

**Supplemental Figure 5.** Volcano Plot of the Metabolome-wide Association Study Comparing TB Resisters and Controls in the South African Cohort (11 Pairs). Horizontal Line Represents P Value of 0.05. Confirmed Metabolites are Labeled.

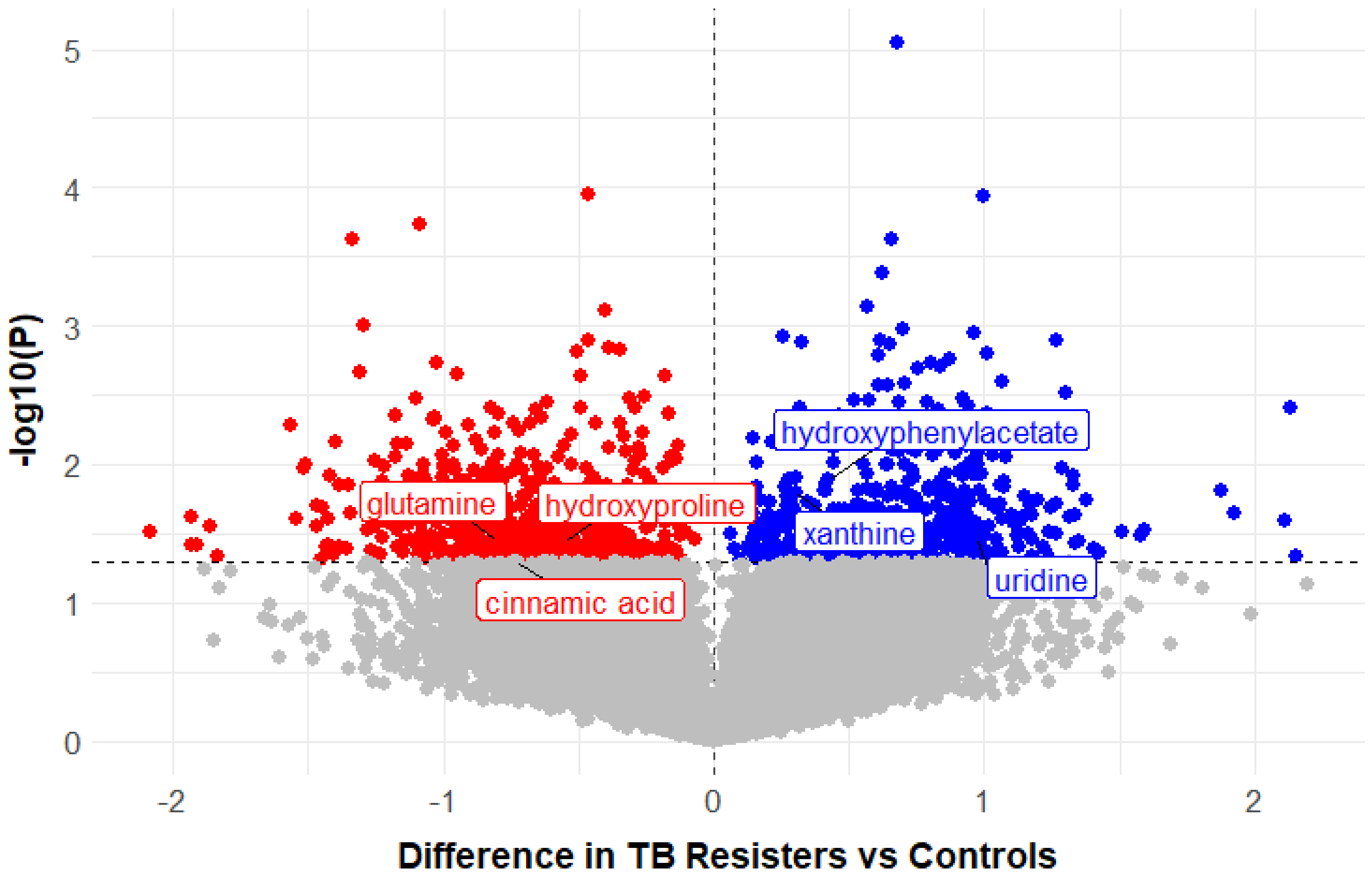
